# Digital Registrar: A Schema-First Framework for Multi-Cancer Privacy-Preserving Pathology Abstraction via Local LLMs

**DOI:** 10.1101/2025.10.21.25338475

**Authors:** Nan-Haw Chow, Han Chang, Hung-Kai Chen, Chen-Yuan Lin, Ying-Lung Liu, Po-Yen Tseng, Li-Ju Shiu, Yen-Wei Chu, Pau-Choo Chung, Kai-Po Chang

**Affiliations:** Center for Precision Medicine, China Medical University Hospital, Taichung, Taiwan; School of Medicine, China Medical University, Taichung, Taiwan; Department of Pathology, China Medical University Hospital, Taichung, Taiwan; Signal1, Toronto, ON, Canada; Department of Computer Science and Information Engineering, National Taiwan University of Science and Technology, Taipei, Taiwan; Department of Foreign Languages and Literature, National Chi Nan University, Nantou, Taiwan; Graduate Institute of Genomics and Bioinformatics, National Chung Hsing University, Taichung, Taiwan; Department of Electrical Engineering, National Cheng Kung University, Tainan, Taiwan

**Keywords:** clinical ontology, cancer registry, CAP protocol, large language model, structured data extraction, DSPy, interoperability, privacy-preserving AI

## Abstract

**Background/Objectives:** Free-text surgical pathology reports hinder automated cancer registry entry and secondary analytics. This study introduces a clinically governed schema layer for interoperability, testing whether a locally-deployable Large Language Model (LLM) pipeline can deliver robust, registry-grade extraction across institutions.

**Methods:** We developed a College of American Pathologists (CAP)-aligned clinical ontology encompassing 10 cancer types, 192 per-organ scalar fields, key biomarkers, and nested structures for lymph nodes and margins. Encoded via Declarative Self-improving Python (DSPy) signatures with grammar-constrained decoding, this model-agnostic pipeline was benchmarked on 893 internal reports against a pathologist-adjudicated gold standard. External validation utilized 242 The Cancer Genome Atlas (TCGA) reports. Hardware feasibility was confirmed on a single 48-gigabyte (GB) Graphics Processing Unit (GPU), ensuring suitability for privacy-preserving, on-premise deployment.

**Results:** Using the gpt-oss-20b model, the framework achieved 92.0% macro-mean exact-match accuracy on internal data, demonstrating near-perfect run-to-run reliability. Critical prognostic indicators, including breast estrogen receptor/progesterone receptor (ER/PR) (98.7%) and margin positivity (>93%), maintained high fidelity. On the external TCGA cohort, accuracy was 77.5%, rising to 88.0% after excluding structurally silent fields absent in older narratives. Operationally, the model processed reports in 40-70 seconds, optimally balancing speed and accuracy.

**Conclusions:** This schema-first abstraction layer successfully decouples clinical logic from specific Artificial Intelligence (Al) models. By reliably transforming narrative reports into machine-readable structures, it establishes a portable, privacy-preserving foundation for automated cancer surveillance, institutional data reuse, and future multimodal clinical systems.

## 1. Introduction

Surgical pathology reports contain the most granular descriptions of cancer diagnosis, staging, margin status, lymph node involvement, and biomarker findings, making them indispensable for cancer surveillance, quality assessment, and secondary clinical research [1–13]. Yet, these data are still documented predominantly as narrative free text. This creates a translational gap between what pathologists report and what registries, analytics pipelines, and downstream clinical systems can computationally reuse. As a result, high-value pathological detail often must be manually re-entered, simplified, or discarded before it becomes available for structured surveillance.

The bottleneck is therefore not only extraction accuracy, but also representation. A durable abstraction system must preserve organ-specific semantics, nested relationships, and variable-length structures such as lymph node groups, specimen margins, and biomarker panels. Flat field lists or ad hoc prompt outputs are insufficient because they frequently collapse clinically meaningful context, making results harder to validate, compare across institutions, and integrate into longitudinal data infrastructures.

Recent large language model (LLM) studies have shown encouraging performance for oncology information extraction, but most have focused on narrow tasks, limited variable sets, or model-specific demonstrations [14–28]. These advances are important, yet model capabilities evolve rapidly and implementation details can age quickly. In contrast, the clinical ontology that governs what should be abstracted, how values are typed, and how repeating structures are organized is the more durable scientific contribution. A schema-first design can therefore outlast any single generation of models while improving reproducibility and interoperability.

The CAP protocols provide an appropriate foundation for such a design because they define stable, clinically governed data elements for cancer reporting. Translating these standards into strictly typed hierarchical schemas makes it possible to explicitly encode registry logic, programmatically constrain outputs, and separate clinical representation from the inference engine itself. This separation is especially important for privacy-preserving deployment, because institutions can retain on-premise control of protected health information while updating local models over time without rewriting the clinical abstraction layer.

Accordingly, we present Digital Registrar, a schema-first framework for privacy-preserving pathology abstraction using local LLMs. Rather than treating structured extraction solely as a model-performance problem, the framework implements CAP-aligned clinical ontologies across ten major cancer types and 192 per-organ scalar field cells (60 unique field names when same-name fields shared across organs are de-duplicated) - including eight clinically acted-on biomarker sub-fields (breast ER/PR/HER2/Ki-67 and colorectal MLH1/MSH2/MSH6/PMS2) and two variable-length nested structures (regional lymph-node groups and surgical margin enumerations) - within a model-agnostic DSPy pipeline executed on a single 48 GB GPU; the full schema composition is summarised in Section 2.2.1 with the per-organ enumeration in Supplementary Tables S1-S10. We evaluate the framework on 893 internal pathology reports under a thirty-run multi-seed protocol and on 242 external TCGA reports under a twenty-two-run protocol, demonstrating registry-grade accuracy while establishing a portable foundation for automated cancer surveillance, institutional data reuse, and future multimodal clinical systems [29-33].

## 2. Materials and Methods

### 2.1. Data Source and Annotation

#### 2.1.1. Data Source

This study was conducted under Research Ethics Committee approval (REC #CMUH114-REC2-037) from China Medical University Hospital and in accordance with all relevant guidelines and regulations. The dataset comprised a total of 893 de-identified surgical pathology reports from the years 2023-2024. The final validation cohort consisted of 694 eligible cancer excision reports. Of these, 664 reports described resected, invasive primary malignancies across ten major organ systems and were used for organ-specific extraction analyses, while 30 reports were categorized as “Others” (dual primaries or out-of-scope cancers) and retained for triage and organ-classification evaluation only; the detailed composition is summarized in Table 1. An additional 199 reports describing biopsies, non-invasive tumors, or benign conditions served as negative controls for the initial classification stages of the pipeline.

**Table 1.**
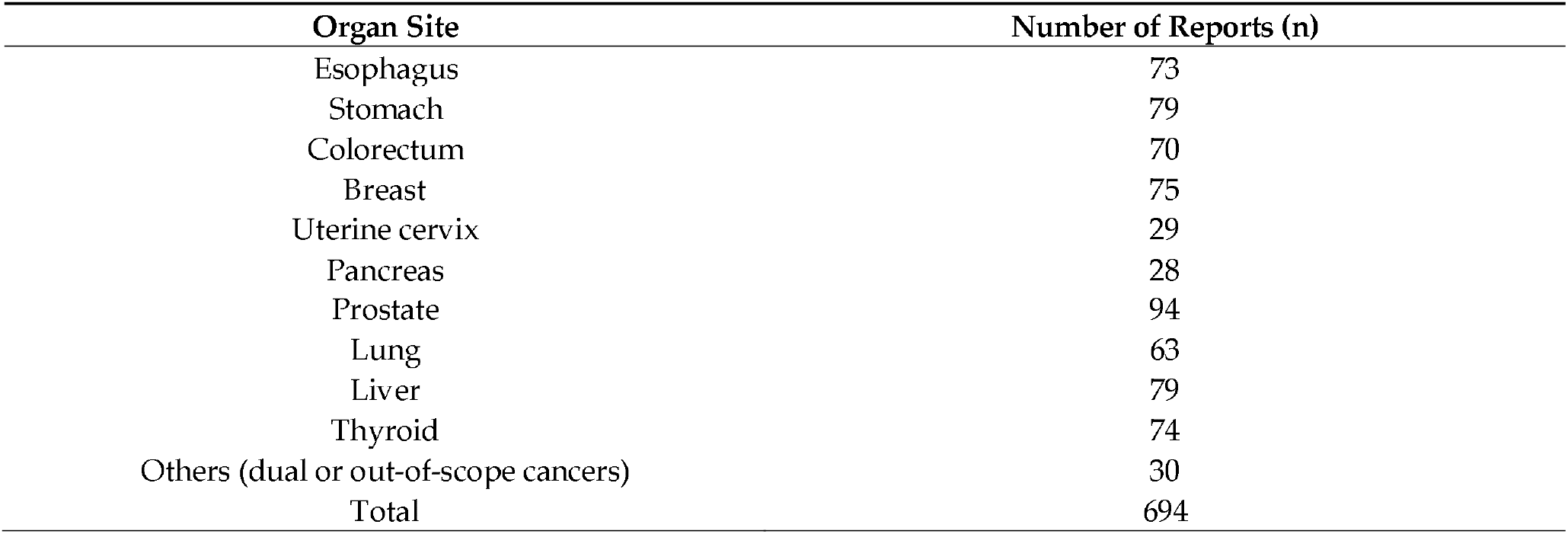
Composition of the cancer excision cohort (n = 694).

| Organ Site | Number of Reports (n) |
| --- | --- |
| Esophagus | 73 |
| Stomach | 79 |
| Colorectum | 70 |
| Breast | 75 |
| Uterine cervix | 29 |
| Pancreas | 28 |
| Prostate | 94 |
| Lung | 63 |
| Liver | 79 |
| Thyroid | 74 |
| Others (dual or out-of-scope cancers) | 30 |
| Total | 694 |

#### 2.1.3 Reference Standard Construction

The gold standard used throughout Section 3 was produced by a two-pathologist annotation committee - Kai-Po Chang (KPC) and Nan-Haw Chow (NHC), both co-authors - with adjudication by a senior third pathologist, Han Chang, also a co-author. The committee re-annotated all 893 reports from scratch under a dedicated GUI annotation tool that persists each annotator’s labels in canonical form and records adjudication decisions as a structured audit trail (Supplementary Table S58). Two arms were run on the same case set: a with-LLM-seed primary arm on all 893 reports (the operational workflow the manuscript proposes), and a without-LLM-seed bias-control arm on a stratified 196-case subset (∼22 %, balanced across organs) that directly tests the same-model-bias concern raised in R1.b. The committee-resolved labels from the primary arm form the gold reference for every Section 3 result. The Δκ between arms (mean +0.015, median 0.0; reported in Section 3.7) confirms the pre-annotation seed does not systematically anchor annotators on the LLM’s output. Full bias-control arm allocation, adjudication χ^2^ protocol (uniformity test against the equal-probability null on (matches_KPC, matches_NHC) outcomes; Holm-corrected across 230 (organ, field) cells), and the annotation tooling reference are in Supplementary Section S1.1 (extended_methods/S1_1_annotation_protocol.md).

#### 2.1.3. Rationale for the Ten-Organ Scope

The ten cancer types covered by the schema (lung, female breast, colorectum, prostate, stomach, liver, thyroid, uterine cervix, esophagus, and pancreas) were selected to align with the world’s most impactful malignancies as ranked by GLOBOCAN 2022 global new-case incidence [39]. We retained the eight cancers in GLOBOCAN ranks 1-8 and added ranks 11 and 12 (esophagus and pancreas), excluding the two intervening entries that do not generate registry-grade surgical-pathology reports under standard clinical workflow: bladder cancer, because most newly diagnosed cases are managed by transurethral resection of bladder tumor rather than CAP-aligned resection reporting; and non-Hodgkin lymphoma, because lymphoma staging is clinical and imaging-based rather than derived from a resected primary specimen. The retained ten cancer types collectively account for approximately 63% of all new cancer cases worldwide (12.5 of 20.0 million in 2022) and each has a published CAP cancer protocol that anchors the schema in stable, clinically governed data elements.

#### 2.1.4. Forward References

Cohen’s kappa is defined as the gold-standard validity metric in Section 2.4. Headline inter-annotator agreement on the committee, the Δκ analysis on the without-LLM-seed arm, and the per-(organ, field) committee-adjudication audit are reported in Section 3.7. All per-field kappa values, the 8-pair kappa matrix, the Krippendorff alpha cross-check, and the per-(annotator, organ, field) Δκ tables are in Supplementary Tables S55-S63.

### 2.2. Schema Composition and Evaluation Scopes

We evaluated the extraction results under several field-count scopes, because different analyses addressed different parts of the schema. The main text performance analysis in Section 3.3 used a 192-cell scalar-field scope across the ten organ systems. These 192 cells correspond to 60 unique scalar field names after fields shared across organs, such as pt_category, grade, procedure, lymphovascular_invasion, and histology, are counted only once. The distinction between 192 per-organ field cells and 60 unique field names is intentional: the same field name may carry different permitted values or clinical meaning across organs, and therefore must be evaluated within its organ-specific schema.

A broader cascade-evaluation scope was used for the component-ablation and multi-run reliability analyses in Sections 3.6 and 3.8. This scope included the 192 scalar field cells, eight clinically acted-on biomarker sub-fields–breast ER, PR, HER2, and Ki-67, and colorectal MLH1, MSH2, MSH6, and PMS2–and three prostate margin-related scalar fields. Together, these form 71 unique field names, corresponding to 203 per-organ field cells. The inter-annotator agreement analysis in Section 3.7 was restricted to 41 categorical fields that were independently re-annotated by the two pathologists under the bias-control protocol. The external TCGA evaluation in Section 3.9 used the subset applicable to the six TCGA organ systems, comprising approximately 124 per-organ scalar field cells and 51 unique scalar field names; this TCGA scope is therefore not directly comparable to the full ten-organ internal scope.

In addition to scalar fields, the schema represents two variable-length nested structures: surgical margins and regional lymph-node groups. Surgical margins are evaluated across the nine organs with explicit margin categories, with prostate handled separately through prostate-specific scalar margin fields. Regional lymph-node groups are evaluated across the ten organs using organ-specific station maps where applicable, while organs without standard nodal-station schemas fall back on counted lymph-node scoring. The complete per-organ schema, including data types, allowed values, nullability, and per-organ field enumeration, is provided in Supplementary Tables S1-S10.

### 2.3. Ontology-Guided Extraction Workflow

The deterministic extraction workflow was implemented in DSPy 2.1 using dspy.Predict() modules with predefined dspy.Signature specifications that encoded essential clinical instructions and ontology constraints. The pipeline was executed on-premises using the gpt-oss-20b model served via Ollama. No model training, fine-tuning, or few-shot prompting was applied; each report was processed in a single forward pass.

#### 2.3.1. Eligibility Classification

Eligibilitysignature predicts whether a report represents a cancer surgery (cancer_excision_report: bool), the organ category (cancer_category), and a free-text descriptor, labeled as “others”.

#### 2.3.2. Organ Detection

If cancer_excision_report = True, a secondary signature (CancerTypeSignature) verifies or refines the organ label prior to extraction.

#### 2.3.3. Organ-Specific Extraction

Ten sets of organ-specific DSPy modules were used for organ-specific extraction. For example, the breast cancer pipeline included BreastCancerNonnested, BreastCancerMargins, BreastCancerLN, and BreastCancerBiomarkers. These modules receive both the original pathology report and the JSON(JavaScript Object Notation) summary previously generated by gpt-oss-20b as input, and validated JSON schemas matching their respective schemas are output. The pipeline is illustrated in Figure 1. The modules extract tumor characteristics, American Joint Committee on Cancer/Tumor-Node-Metastasis (AJCC/TNM) staging, surgical margins, and lymph node details for each cancer type. For breast and colorectal cancers, biomarker status is also extracted. Supplementary Appendix S25 provides examples of model output.

**Figure 1.**
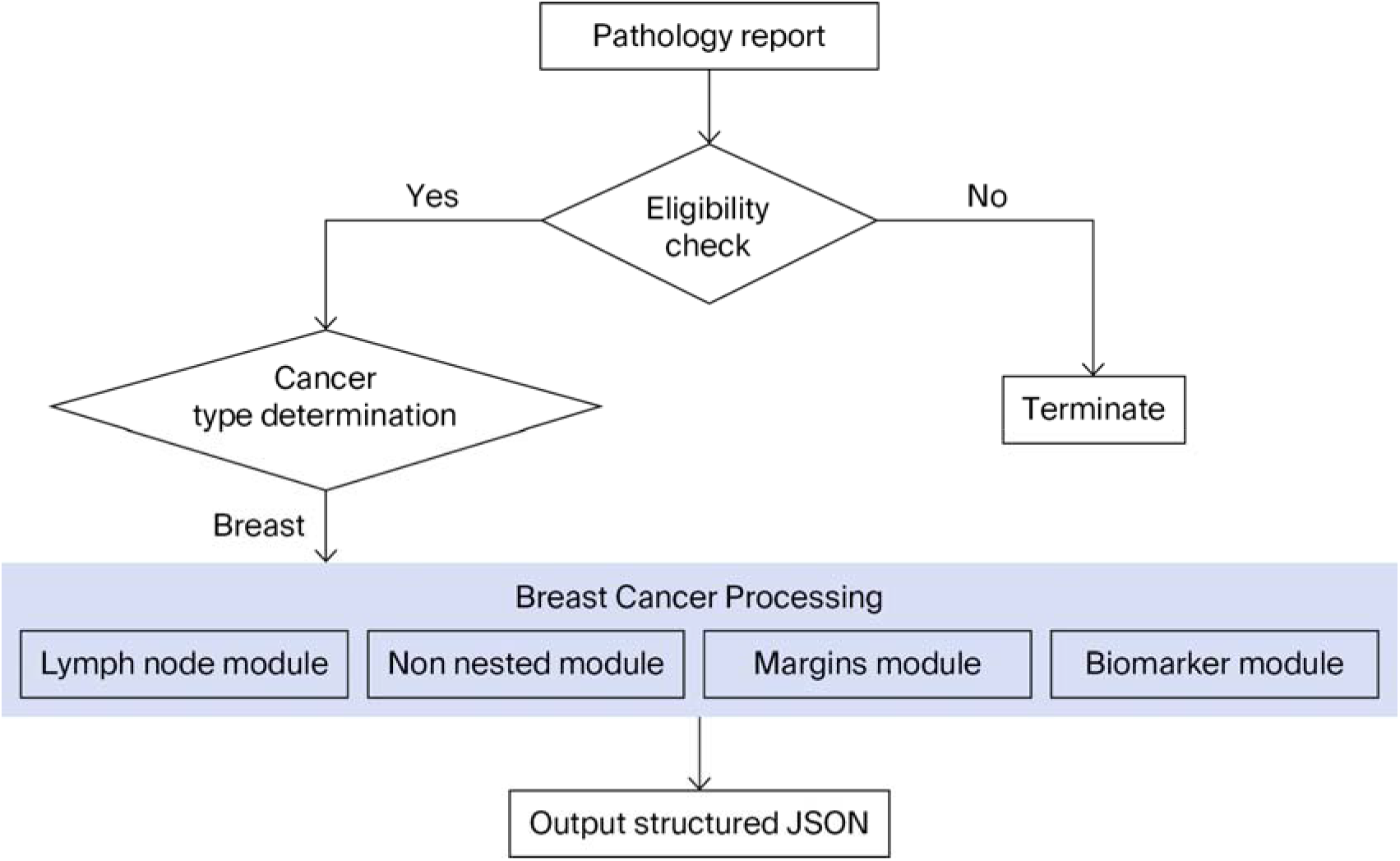
Ontology-guided extraction pipeline for structured cancer data from surgical pathology 205 reports. The eligibility classifier (Stage A) feeds the organ classifier (Stage B); per-organ DSPy 206 modules then emit JSON-schema-validated structured fields (Stage C).

#### 2.3.4. Validation of Model Agnosticism and Hardware Feasibility

To validate the architectural independence of the extraction framework and assess the feasibility of deploying the framework on standard workstation hardware, an identical pipeline structure was instantiated and executed using two alternative open-weight transformer architectures: Qwen3-30B-A3B (sparse mixture of experts (MoE)) and Gemma 3 27B (dense). The foundational design of the AI pipeline explicitly embraces principles of model agnosticism, utilizing the DSPy 2.1 framework to decouple the extraction logic from specific model weights.

All comparative models were executed on-premises using the same dedicated NVIDIA RTX A6000 Ada Generation GPU (48 GB video RAM (VRAM)). This hardware constraint was strictly enforced to evaluate whether models could operate efficiently within the memory envelope of a standard high-end medical workstation without requiring multi-GPU datacenter configurations. The overall ontology-guided extraction workflow is summarized in Figure 1.

### 2.4. Evaluation Protocol and Statistical Methods

#### 2.4.1. Reference Comparison and Metric Definitions

Model performance was evaluated by comparing each generated JSON output against a committee-resolved gold standard constructed by two annotating pathologists with adjudication by a senior third pathologist (see Section 2.1). Because all schema variables are strictly typed (Literal, bool, int, or None), evaluation was performed using deterministic exact-string matching rather than similarity-based scoring. For list-typed variables - surgical margins and lymph-node groups - corresponding elements were aligned by canonical category before comparison; biomarker scope was restricted to the eight clinically-acted-on markers (er, pr, her2, ki67 for breast; mlh1, msh2, msh6, pms2 for colorectal); and four meta-fields (ajcc_version, treatment_effect, margins [*].description, regional_lymph_node[*].station_name) were excluded from per-field statistics in favor of category-aggregated scoring (full exclusion list in Supplementary Table S57).

The primary metrics were per-field exact-match accuracy, Cohen’s kappa relative to gold, F1 (with macro and micro averages where applicable), and balanced accuracy for multiclass classification. Composite metrics for the margin and lymph-node modules - margin status correctness, margin distance correctness, total lymph-node accuracy (with ±1 tolerance on integer counts), and lymph-node station accuracy - are defined in Table 2

**Table 2.** Evaluation metrics and definitions.

| Metric | Definition |
| --- | --- |
| Margin status correctness | Exact match of both margin_category and margin_involved for every reported margin element. |
| Margin distance correctness | Exact match of margin_category, margin_involved, and distance (integer, mm) for each margin element. |
| Total lymph node accuracy | Exact match of examined and involved integer counts within each organ's regional_lymph_node list ($\pm 1$ tolerance). |
| Lymph node station accuracy | Exact match of station_name, examined, and involved values for each lymph node group. |
| Accuracy of other fields | Proportion of all remaining scalar fields that exactly match the expert reference. |

Throughout Section 3, per-attempt effective accuracy = n_correct / n_eligible (a missing emission counts as wrong against any non-null gold cell); coverage = n_attempted / n_eligible (the emission rate). For the two upstream classification end-points (eligibility and organ routing), thirty repeated inferences are first collapsed to a report-level majority result, and Wilson intervals use the number of independent reports as the denominator. For scalar extraction summaries, repeated inferences are first collapsed to a case-field-level majority correctness label, and Wilson intervals use eligible case-fields rather than case-field-runs as the denominator; the per-organ macro mean averages per-organ majority accuracies with equal weight per organ. Pooled case-run and case-run-field accuracies are retained only as descriptive repeated-inference summaries. Patient-level cohort size remains the number of unique reports. Formal definitions can be found in supplementary table S1-10.

#### 2.4.2. Multi-Run Protocol

To quantify run-to-run stability under fixed inputs, the full extraction was repeated across thirty independent inference runs of the gpt-oss-20b model with identical prompts and decoding parameters but distinct random seeds. The external TCGA evaluation in Section 3.9 uses a separate k = 22 multi-seed pass for compute-budget reasons. Reliability is analysed separately from the accuracy intervals by treating the thirty seed outputs as a case-by-run result matrix: ICC(2,1) summarises single-run reproducibility, ICC(3,k) summarises the thirty-run mean (Spearman-Brown extrapolation), Cronbach’s alpha summarises internal consistency across seeds, and flip rate reports how often a case-level correctness indicator changes across runs. Per-case run SD complements the ICC for list-typed fields. Per-stage and per-organ reliability is in Section 3.8 and Supplementary Table S35.

#### 2.4.3. Component Ablation Protocol

To quantify the contribution of the pipeline’s engineering choices, we performed a five-cell component-ablation study on the same internal CMUH cohort against the same committee-resolved gold standard. The evaluated cells were the proposed modular DSPy pipeline and four progressively stripped-down variants that removed, in sequence, per-organ decomposition, the Reportjsonize pre-pass, the DSPy structured-decoding framework, and prompt-level schema discipline. The resulting variants were dspy_modular, dspy_monolithic, dspy_monolithic_no_jsonize, raw_json, and free_text_regex.

This design was intended as a component-level lesion study rather than a fully orthogonal factorial experiment, because each stripped-down variant removes one additional upstream engineering layer relative to the variant above it. The ablation therefore tests whether the removal of each structural constraint produces a measurable degradation in extraction performance. Run-to-run reliability of the proposed pipeline is reported separately in Section 3.8.

Headline ablation results are presented in Section 3.6. Detailed cell definitions, implementation paths, field-type stratification, and the full variant-component matrix are provided in Supplementary Section S1.3.

#### 2.4.4. Confidence Intervals, Hypothesis Tests, and Effect Sizes (Summary)

For proportion-based outcomes, 95% Wilson confidence intervals were computed on the appropriate majority-aggregated scoring unit. For report-level classification end-points, repeated inference runs were first collapsed to one majority result per report. For scalar extraction endpoints, repeated runs were first collapsed to one majority correctness label per eligible case-field. Thus, Wilson intervals used independent reports or eligible case-fields as denominators, rather than pooled case-runs or case-run-fields. Pooled repeated-inference accuracies were retained only as descriptive summaries of run-level performance.

Run-to-run reliability was evaluated separately using intraclass correlation coefficients, Cronbach’s *α*, flip rates, and per-case run standard deviations where applicable. Paired binary comparisons used McNemar’s test, and multiclass comparisons used Stuart-Maxwell tests. When multiple comparisons were performed within the same comparison family, p-values were adjusted using the Holm-Bonferroni method. Full derivations, sensitivity analyses, and software version information are provided in Supplementary Section SI .3.

### 2.5. Independent External Validation (TCGA Cohort)

The external validation cohort was the publicly released TCGA Pathology Reports dataset (Kefeli and Tatonetti, 2024 [33]). For this revision, the external evaluation was broadened to 242 reports spanning six per-organ schemas: breast, colorectal, esophagus, liver, stomach, and thyroid. These reports were processed end-to-end under a 22-seed multi-run protocol with identical prompts and decoding parameters; only the random seed varied across runs. The same TCGA cohort was also used for the four-system cross-architecture benchmark in Section 3.10.

TCGA is external to CMUH along three clinically relevant axes: jurisdictional setting, institutional structure, and reporting style. The internal cohort was drawn from a single Taiwanese hospital with relatively consistent reporting conventions, whereas the TCGA cohort reflects a federated multi-institutional corpus with heterogeneous templates and historical reporting practices. In addition, the TCGA-Reports release is derived from OCR-processed pathology-report PDFs and is described by the corpus authors as moderately curated, with residual text-quality limitations. These corpus-side factors are expected to contribute to the residual extraction error observed in Sections 3.9 and 3.10.

The TCGA-applicable schema covered approximately 124 per-organ scalar field cells, corresponding to roughly 51 unique scalar field names across the six organs. This external scope is therefore not directly comparable to the full ten-organ internal scope. The TCGA schema included the relevant scalar staging and diagnostic fields, breast ER/PR/HER2/Ki-67 biomarkers, colorectal mismatch-repair biomarkers, and the same variable-length list structures for surgical margins and regional lymph-node groups where applicable. Scalar field extraction was evaluated using the same majority-aggregation policy described in Section 2.4: pooled case-run and case-run-field outputs were retained only as repeated-inference summaries, whereas headline Wilson intervals were computed after collapsing repeated runs to report-level or case-field-level majority units.

The full-schema TCGA evaluation intentionally retained 13 fields with known source-documentation limitations, including older breast biomarker panels, colorectal mismatch-repair immunohistochemistry, AJCC-edition-dependent stage-group fields, and pm_category coding conventions. These fields were retained in the full-schema headline to provide an honest external validation result, and a sensitivity analysis excluding them is reported in Section 3.9 to estimate performance on fields consistently carried by the TCGA source narratives.

### 2.6. Baseline Systems for Cross-Architecture Comparison

To contextualize the proposed local-LLM pipeline against established methods, four systems were benchmarked head-to-head on the TCGA external cohort (Section 2.5): (i) a deterministic rule-based extractor (regex + lexicon per ten organ schemas; coverage policy: omit-on-no-signal)–the engineering floor; (ii) a BERT-merged encoder reference (Bio_ClinicalBERT shared encoder with per-field classification heads and a SQuAD-style extractive question-answering head for numeric spans; fine-tuned on the full CMUH annotation pool, predicted on held-out TCGA)–the encoder-only structural ceiling, with nested variable-length list fields unreachable by construction; (iii) the proposed local LLM (gpt-oss-20b served by Ollama with llama.cpp decoding through DSPy Signature declarations)–which runs on-premises with no protected health information(PHI) leaving the institutional perimeter; and (iv) an API LLM (gpt-5.4-mini via OpenAI’s native response_format = {“type”:”json_schema”) mode)–the closed-frontier reference. TCGA is the only corpus that admits a fair four-way comparison: the API model cannot be sent CMUH data for privacy reasons, and the BERT baseline was fine-tuned on CMUH, such that predicting on CMUH would leak training cases. All four systems consumed the same plain-text reports, emitted the same per-organ schema, and were scored under the identical three-stage cascade defined in Section 2.4. Full implementation specifications are provided in Supplementary Section S1 .2 (Extended Methods).

## 3. Results

### 3.1. Assessment of Model Agnosticism and Computational Feasibility

The proposed framework is architecturally model-agnostic: the DSPy 2.1 layer compiles each dspy.Signature declaration into a JSON schema, which the inference backend converts to a grammar-constrained sampling mask. The extraction logic–per-organ module enumeration, cascade gating, and schema typing–is independent of the model weights; an LLM enters the pipeline as an interchangeable inference engine. To validate this architectural property in practice, we screened three open-weight LLMs end to end on the 893-report CMUH cohort in a single-run pilot model-selection study against the v1 working gold standard (i.e., prior to the committee-resolved gold standard adjudication step described in Sections 2.1.2 and 3.7). All three models executed an identical pipeline structure on the same dedicated NVIDIA RTX A6000 Ada Generation GPU (48 GB VRAM). Table 3 shows that gpt-oss-20b was selected as the production inference engine on the basis of the best speed-accuracy balance: it achieved the highest pilot exact-match accuracy and 2-3× faster per-report latency than the alternatives. All headline accuracies reported in Sections 3.3−3.10 of this paper use gpt-oss-20b under the 30-run multi-seed committee-resolved gold standard protocol.

**Table 3.** Pilot model-selection study (single-run, v1 working gold standard, 193-field v10 evaluation scope). Three open-weight LLMs were screened on the 893-report CMUH cohort to identify the production inference engine; gpt-oss-20b was selected based on the best speed/accuracy balance. All headline accuracies in Sections 3.3–3.10 use gpt-oss-20b under the 30-run committee-resolved gold standard protocol and are not directly comparable to these pilot values (see the closing note in Section 3.3).

| Model | Architecture | Total/Active Params | Pilot Mean Exact-Match Accuracy | Per-Report Latency |
| --- | --- | --- | --- | --- |
| gpt-oss-20b | Sparse MoE | 20.9 B/3.6 B | 94.30% | 40–70 s |
| Qwen3-30B-A3B | Sparse MoE | 30.5 B/3.3 B | 92.90% | 140–>200 s |
| Gemma 3 27B | Dense | 27 B | 89.80% | 50–130 s |

Based on these pilot accuracies, gpt-oss-20b was selected as the production engine. Under the 30-run multi-seed protocol against the committee-resolved gold standard, the corresponding gpt-oss-20b per-organ macro-mean accuracy on the 192-field scope is 90.9% (Section 3.3). The lower headline value reflects four protocol differences from this pilot: (i) 30-run macro average vs. single run; (ii) 192-field paper scope vs. 193-field v10 scope; (iii) committee-resolved gold standard vs. v1 working gold standard; and (iv) per-organ macro mean vs. dataset-pooled mean. The architectural claim–that LLMs serve as interchangeable inference engines under DSPy–stands on this pilot; the headline performance is what the selected production pipeline delivers. Detailed architectural rationale (per-organ DSPy module enumeration, sparse-MoE routing overhead and bandwidth notes, decoding parameters) is provided in Supplementary Section S1.4.

### 3.2. Automated Report Triage and Organ Classification

Across the fixed 893-report validation cohort, repeated over 30 independent inference runs, the eligibility classifier achieved a report-level majority accuracy of 95.74% [95% Wilson CI: 94.21, 96.88], using independent reports as the interval denominator. Report-level sensitivity was 99.85% [99.18,99.97] (684/685 gold-eligible reports), specificity was 82.21% [76.44, 86.81] (171/208 gold-ineligible reports), and Cohen’s κ was 0.873 (Table 4). No report tied under the 30-run majority rule. The corresponding pooled case-run accuracy was 95.68% (25,632/26,790 case-runs), retained only as a repeated-inference performance summary rather than as an independent clinical sample size. Run-to-run reliability is reported separately in Section 3.8 and Supplementary Table S35. The false positives clustered in clinically ambiguous instances, such as large metastatic deposits or extensive biopsy specimens.

**Table 4.** Eligibility classifier confusion matrix aggregated across 30 independent runs on the 893-report CMUH validation cohort (n = 26,790 case-runs) against the committee-resolved gold standard. The cell counts from Section 3.2 show an accuracy of 95.68%, a sensitivity of 99.91%, and a specificity of 81.75%. The v10 single-run analog (run 1 against the v1 working gold standard) is shown in Supplementary Table S25.

|  | Predicted Eligible | Predicted Ineligible | Total |
| --- | --- | --- | --- |
| Actual Eligible | 684 TP | 1 FN | 685 |
| Actual Ineligible | 37 FP | 171 TN | 208 |
| Total | 721 | 172 | 893 |

For organ classification, the report-level endpoint was restricted to gold-eligible reports that also passed the majority eligibility gate (n = 684; one gold-eligible report failed the majority eligibility gate). Majority organ assignment reached 96.93% [95.35, 97.98] accuracy, with macro-F1 = 0.942 and Cohen’s κ = 0.966. The corresponding pooled Stage-B case-run summary was 97.11% (19,924/20,517 case-runs) within this report-level denominator; the chapter-wide pooled summary was 19,938/20,531 when all Stage-B case-runs were counted. These pooled summaries are retained only as repeated-inference descriptions. F1 was ≥0.95 across the ten in-scope organs in the pooled per-class table, and the dominant residual error pattern involved the gold-”others” class, quantified separately in Section 3.2.1.

#### 3.2.1. Multi-Primary and Out-of-Scope Triage

Across the 30 multi-seed runs, 1138 gold-standard “others” ledger entries were evaluated. At Stage B, the dispositions were as follows: 449 (39.5%) were correctly triaged to “others”; 631 (55.4%) were misrouted into a single in-scope organ; and 58 (5.1%) were over-flagged as “others”. Decomposing the 631 misroutes by the free-text annotation yielded 180 (28.5%) synchronous multi-primaries within an in-scope organ system, 163 (25.8%) out-of-scope sites approximated to a neighboring organ, and 288 (45.6%) phrasings that the regex could not confidently classify; lung was the dominant misroute attractor (n = 150), followed by pancreas (n = 108). A full disposition table is provided in Figure 2/Supplementary Table S30; per-organ misroute counts are shown in Supplementary Figure S8/Table S31.

**Figure 2.**
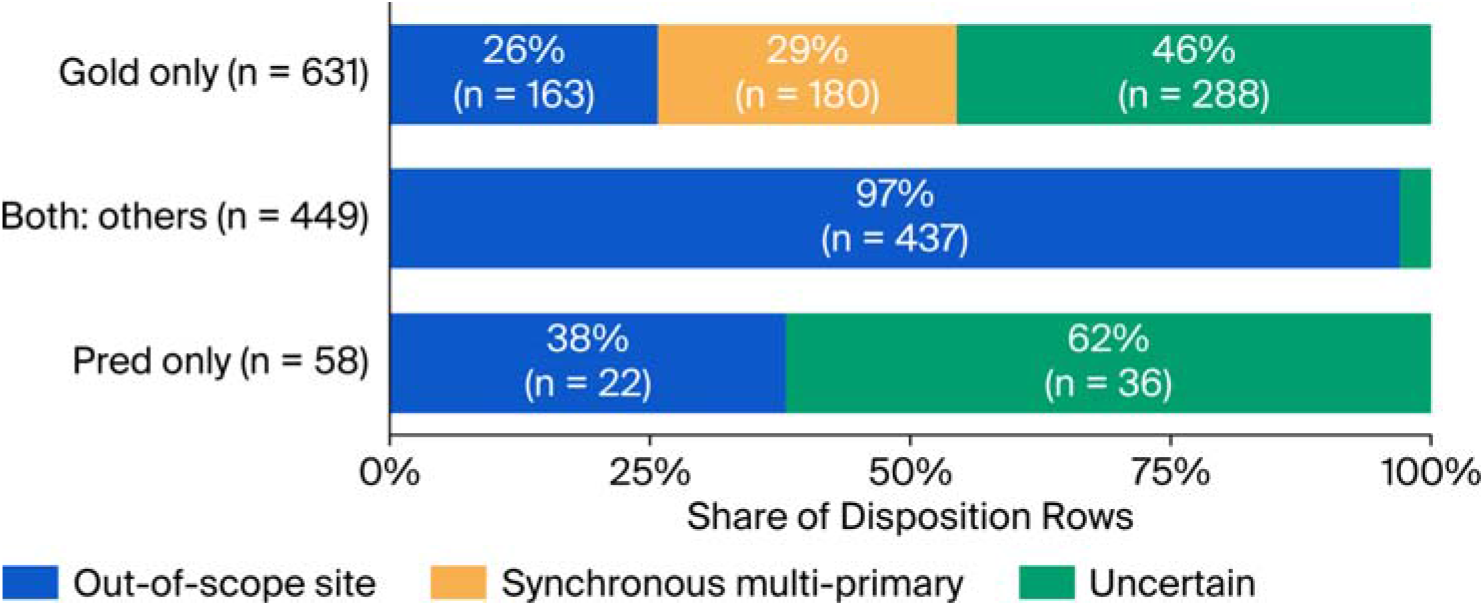
Multi-primary triage outcomes across the 30 multi-seed runs (1138 repeated-inference 427 ledger entries involving gold-standard “others” cases). The Stage B dispositions and three-way 428 decomposition of the misrouted entries are shown.

### 3.3. Per-Organ Extraction Accuracy Across Ontology Fields

After classification, each cancer-surgery report was processed through the corresponding organ-specific extraction modules. Per-organ schemas are listed in Supplementary Tables S1-S10, and per-organ per-field accuracy tables are provided in Supplementary Tables S14-S23. For the Section 3.3 main-text evaluation scope, 192 registry field cells across ten organ systems were evaluated after collapsing repeated inferences to case-field-level majority correctness labels before Wilson intervals were computed.

Per-organ majority accuracy ranged from 89.35% [95% Wilson CI: 86.09, 91.92] in cervix to 95.36% [94.43, 96.14] in prostate, with a per-organ macro mean of 92.03% (Table 5; Figure 3; Supplementary Table S27). The corresponding pooled case-run-field macro mean was 90.75% and is retained only as a descriptive repeated-inference summary. The bottom-decile field error attribution underlying this gradient is decomposed in Section 3.3.2 using a six-mechanism taxonomy: M1, source-bound qualitative phrasing; M2, het-erogeneous reporting / glossary-rule fixable; M3, anatomic ontology gap; M4, AJCC convention enforcement; M5, schema shape; and M6, unresolved model-side semantic conflation. The breast pathologic-versus-anatomic stage-group comparison is reported in Section 3.3.1. Run-to-run stability is reported separately in Section 3.8; no field exhibited a parse error or schema-validation failure across the thirty runs.

**Table 5.** Per-organ extraction-accuracy headline after case-field-level majority aggregation. Repeated inferences were collapsed to case-field-level majority correctness labels before Wilson intervals were computed. The denominator n case-fields is the number of eligible case-field units, not case-field-runs. Rows are sorted in descending order by majority accuracy. The pooled repeated-inference companion summary is retained in Supplementary Table S27.

| Organ | n_fields | Effective Accuracy [95% Wilson CI] |
| --- | --- | --- |
| Prostate | 25 | 95.36% [94.43, 96.14] |
| Colorectal | 20 | 93.96% [92.61, 95.08] |
| Esophagus | 16 | 93.41% [91.84, 94.69] |
| Thyroid | 19 | 93.06% [91.58, 94.29] |
| Breast | 26 | 93.03% [91.81, 94.07] |
| Lung | 20 | 91.45% [89.65, 92.97] |
| Liver | 17 | 91.20% [87.90, 93.67] |
| Pancreas | 15 | 90.00% [88.29, 91.48] |
| Stomach | 18 | 89.45% [87.75, 90.94] |
| Cervix | 16 | 89.35% [86.09, 91.92] |
| Macro mean |  | 92.03% |

**Figure 3.**
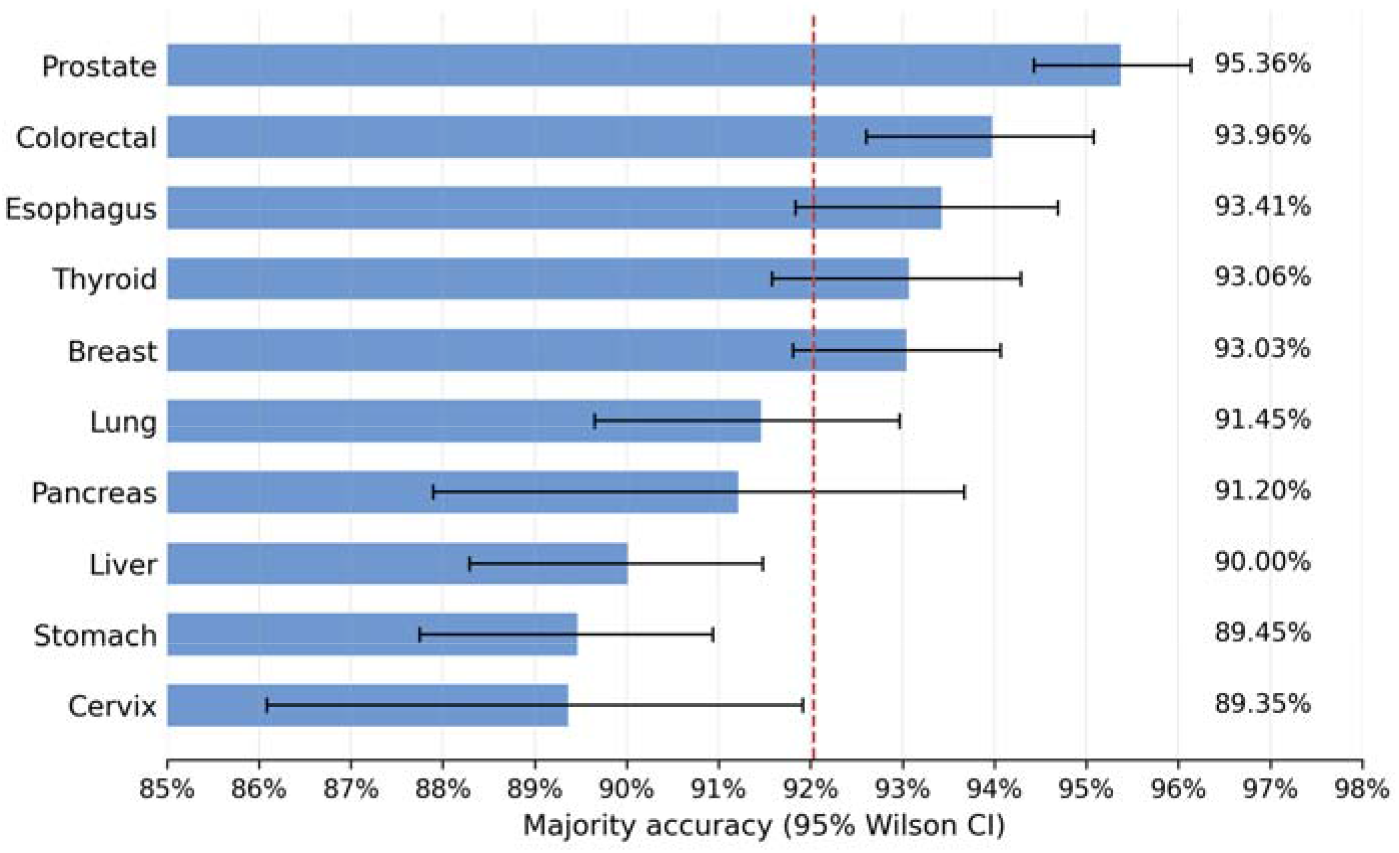
Per-organ extraction accuracy after case-field majority aggregation with 95% Wilson confidence intervals.

#### Note for readers comparing to Section 3.1 / Table 3

The Section 3.1 / Table 3 pilot value for gpt-oss-20b (94.30%, single run against the v1 working gold standard over the 193-field v10 evaluation scope) is not directly comparable to the Section 3.3 / Table 5 headline of 92.03%. The Section 3.3 protocol differs from the pilot on four axes: (i) case-field-level majority aggregation across thirty repeated inference runs versus a single run; (ii) 192-field paper scope versus 193-field v10 scope; (iii) committee-resolved gold standard versus v1 working gold standard prior to committee adjudication; and (iv) per-organ macro mean versus dataset pooled mean The values in Section 3.3 / Table 5 therefore represent the headline performance of the production pipeline under the revised committee-gold protocol.

#### 3.3.1. Anatomic vs. Pathologic Stage Group Disambiguation in Breast Reports

The breast schema separates two stage-group fields, anatomic_stage_group and pathologic_stage_group, evaluated on the same 75 independent breast reports. Repeated inferences were collapsed to case-field-level majority correctness before interval estimation. The two fields showed markedly different performance: anatomic_stage_group reached 98.67% [95% Wilson CI: 92.83, 99.76], whereas pathologic_stage_group reached 80.00% [69.59,87.49], an 18.67 percentage-point gap (Figure 4; Supplementary Table S28). The corresponding pooled repeated-inference accuracies were 97.95% and 74.87%, respectively, and are retained only as descriptive repeated-inference summaries.

**Figure 4.**
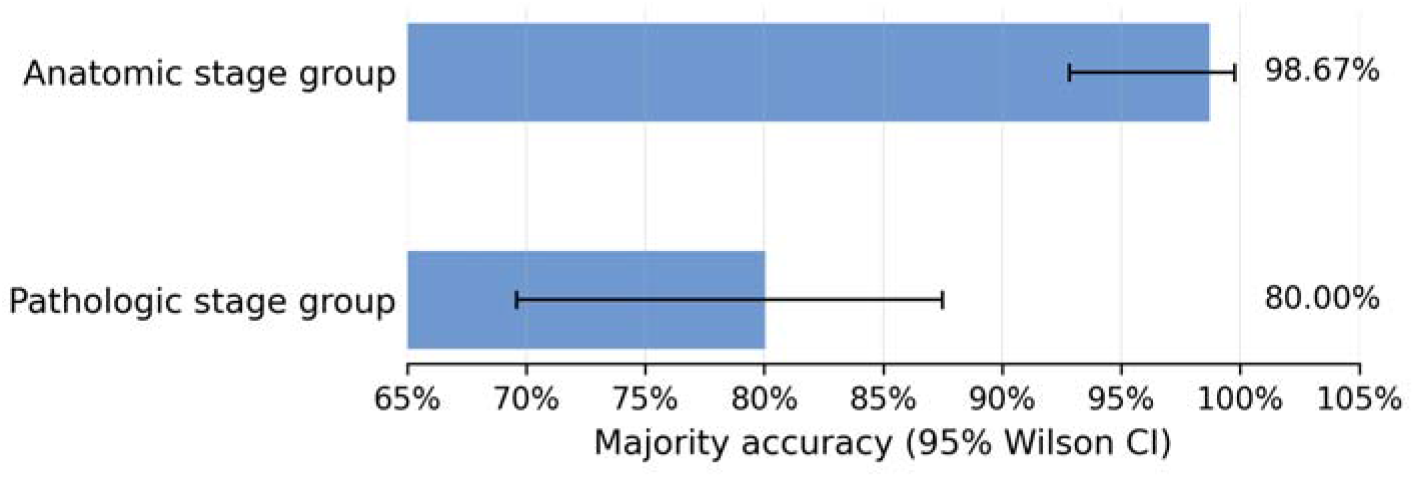
Breast anatomic versus pathologic stage-group extraction after majority aggregation

**Figure 5.**
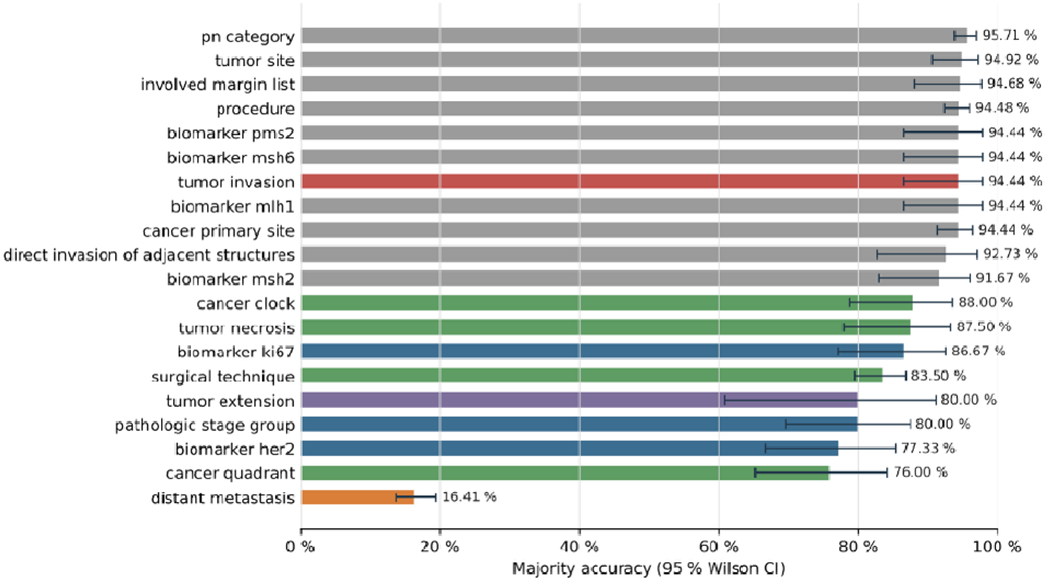
Bottom-decile field majority accuracy with six-mechanism error attribution.

Manual review indicated that the residual errors were structural rather than random noise. Three AJCC 8-related mechanisms accounted for most discrepancies: (i) post-treatment y-descriptor cases, for which AJCC 8 directs assignment to clinical prognostic stage, while the institutional source reports often provide only anatomic_stage_group; (ii) HER2 2+ equivocal cases pending fluorescence in situ hybridization (FISH), for which prognostic stage is genuinely indeterminate from the pathology report alone; and (iii) residual narrative ambiguity in non-y reports, where the model tended to default to anatomic staging. Two complementary structural mitigations–AJCC 8 stage-table injection and a multi-valued pathologic_stage_group representation for indeterminate cases–are proposed in Section 4.

#### 3.3.2. Bottom-Decile Field Error Attribution

Nine registry fields fell below 90% majority accuracy after repeated inferences were collapsed at the case-field level. We attributed these residual errors using a six-mechanism taxonomy that separates likely engineering remedies: M1, source-bound qualitative phrasing; M2, heterogeneous reporting or null-on-no-clue behavior; M3, anatomic ontology gap; M4, AJCC convention enforcement; M5, schema shape; and M6, unresolved model-side semantic conflation. M1 did not surface under the current majority-aggregated denominator.

Four fields fell under M2: surgical_technique (83.5%), tumor_necrosis (87.5%), cancer_clock (88.0%), and cancer_quadrant (76.0%). The surgical-technique residual was driven largely by heterogeneous prefix vocabulary, such as 3D or robotic descriptors, that the current schema does not fully disambiguate. One M4 case was distant_metastasis, with 16.4% [95% Wilson CI: 13.8,19.4] majority accuracy, reflecting an AJCC convention problem in which absent evidence of metastasis is inconsistently represented between the source narrative and registry gold.

M5 cases included pathologic_stage_group (80.0%; Section 3.3.1) and the two breast biomarker fields HER2 (77.3%) and Ki-67 (86.7%), where the schema over-specifies the biomarker sub-record. Specifically, HER2 should expose only clinically meaningful score/status information, whereas Ki-67 should expose only percentage; extraneous schema slots invite hallucinated or mismatched values. The pooled repeated-inference companion table still flagged colorectal tumor_invasion as a model-side semantic-confusion residual, but after case-field majority aggregation its accuracy rose above the bottom-decile threshold. Similarly, the global procedure field did not fall below 90% after majority aggregation, although liver procedure remains the canonical M3 example because mapping segmental liver resections onto CAP partial-hepatectomy categories requires anatomic segment-counting. This decomposition reframes the residual from a generic “low-performing field” problem into specific components that are closable by glossary/rule fixes, schema revision, ontology expansion, or further model-side semantic improvement.

### 3.4. Lymph Node and Surgical Margin Status

Across the nine organ systems for which the schema enumerates explicit surgical-margin categories, excluding prostate because its CAP protocol uses prostate-specific scalar margin fields, the pipeline identified margin involvement, defined as any-margin positivity, with high report-level majority accuracy. After repeated inferences were collapsed to report-level majority results, any-margin positivity ranged from 93.33% [95% Wilson CI: 85.32, 97.12] in breast to 100.00% in cervix, colorectum, lung, and thyroid. The residual list-level errors were dominated by hallucinated margin entries, defined as predicted margin elements with no gold counterpart, rather than missed margin elements. Hallucination rates were highest in lung, thyroid, liver, esophagus, and stomach, whereas miss rates remained uniformly low. These hallucination and miss rates are retained as pooled list-level descriptive summaries rather than report-level Wilson end-points. Because hallucinated entries generally defaulted to margin_involved = false in the gold-aligned scoring, this list-level hallucination pattern did not materially degrade the clinically critical any-margin-positive endpoint. Per-organ margin summaries are provided in Supplementary Table S32, with the prostate scalar-field margin callout reported in Supplementary Table S33.

Any-positive nodal status, the clinically critical endpoint asking whether any regional lymph node was involved, was near ceiling after report-level majority aggregation. Nine of ten organ systems reached 100.00% majority correctness, and thyroid reached 98.61% [92.54, 99.75]. More granular lymph-node endpoints remained more challenging. In the pooled repeated-inference summary, total-count concordance with ±1 tolerance ranged from 96.0% in colorectum to 45.5% in esophagus. The harder per-station group-recall endpoint showed a recall-precision asymmetry: prostate, cervix, and esophagus had low recall but comparatively high precision, consistent with station-name canonicalization gaps rather than node fabrication. Because any-positive nodal correctness, the endpoint most directly affecting downstream stage assignment, was near ceiling in nine of ten organs, the remaining station-level errors are best interpreted as recoverable engineering and ontology-mapping issues rather than immediate clinical-safety failures. Per-organ lymph-node summaries are provided in Supplementary Table S34 and Supplementary Figure S9.

### 3.5. Breast Biomarker Extraction Performance

Breast biomarker extraction was evaluated on 75 independent breast reports for the four clinically actionable receptor markers, with repeated inferences collapsed to report-biomarker majority correctness before Wilson intervals were computed. ER and PR each reached 98.67% [95% Wilson CI: 92.83, 99.76] majority accuracy (74/75), Ki-67 reached 86.67% [77.17, 92.59] (65/75), and HER2 was the lowest of the four at 77.33% [66.66, 85.34] (58/75). The corresponding pooled repeated-inference summaries were similar in point estimate—ER 98.67%, PR 98.31%, Ki-67 86.25%, and HER2 77.31%—and are retained only descriptively.

The bottom-decile field attribution in Section 3.3.2 identifies HER2 and Ki-67 as M5 schema-shape residuals. HER2 has a single clinically meaningful immunohistochemistry score/status dimension, whereas the current schema exposes score, percentage, and positivity slots; the extraneous slots have no clinical referent and invite hallucinated values. Ki-67 is the complementary case: the clinically meaningful value is the staining percentage, whereas the additional score and positivity slots create unnecessary error surfaces. A schema revision that keeps score/status only for HER2 and percentage only for Ki-67 was identified during re-audit and is queued for the next pipeline iteration. ER and PR sit near the upper performance limit because their schemas are already appropriately narrow.

Colorectal mismatch-repair biomarkers—MSH2, MSH6, MLH1, and PMS2—were evaluated on 72 independent colorectal reports after case-field majority aggregation. Majority accuracy was 91.67% [82.99, 96.12] for MSH2 (66/72) and 94.44% [86.57, 97.82] for MSH6, MLH1, and PMS2 (68/72 each). The MSH2 accuracy is partly bounded by source coverage, because a minority of CMUH colorectal reports do not document MMR immunohistochemistry; these gold-null cases occasionally elicit a default model output, the same “silent source to LLM hallucination” pattern catalogued for TCGA in Section 3.9, but at a lower rate in the internal cohort where MMR immunohistochemistry is usually part of the routine workup.

Run-to-run reliability across the thirty runs was at the noise floor for the four breast biomarkers: within-case accuracy SD was below 0.01 for ER, PR, and HER2 categorical positivity, and the accuracy flip rate was below 1% across all four. These observations indicate that the remaining biomarker errors are primarily source- or schema-bound rather than run-stochastic.

For the same breast cohort, BRCA1, BRCA2, and TP53 mutation status were not present at meaningful rates in the source surgical-pathology narratives. These markers are typically reported in separate molecular-pathology workups rather than in surgical-resection reports. Their integration is therefore deferred to the planned multimodal genomics extension, rather than added to the current schema without a consistent source-data layer.

### 3.6. Component Ablations

To quantify the contribution of the engineering choices that distinguish Digital Registrar from a naïve free-text or single-prompt approach, we performed a five-cell component-ablation study on the internal CMUH cohort against the same committee-resolved gold standard. The ablation tested progressively stripped-down variants of the proposed pipeline by removing per-organ decomposition, the Reportjsonize pre-pass, the DSPy structured-decoding framework, and prompt-level schema discipline, as defined in Section 2.4.3.

Using the majority/natural case-field sensitivity analysis for the chapter-3 scalar-field subset, the proposed dspy_modular pipeline achieved 88.26% [95% Wilson CI: 87.75, 88.75] accuracy. Performance decreased to 84.36% [83.78, 84.91] for dspy_monolithic_no_jsonize, 83.74% [83.16, 84.31] for dspy_monolithic, and 81.53% [80.92, 82.12] for rawjson. The schema-blind free_text_regex baseline fell to 18.17% [17.57, 18.79] (Figure 6; Supplementary Tables S45-S54). These ablation results are not directly comparable to the Section 3.3 per-organ macro mean of 92.03%, because the aggregation target and run protocol differ.

**Figure 6.**
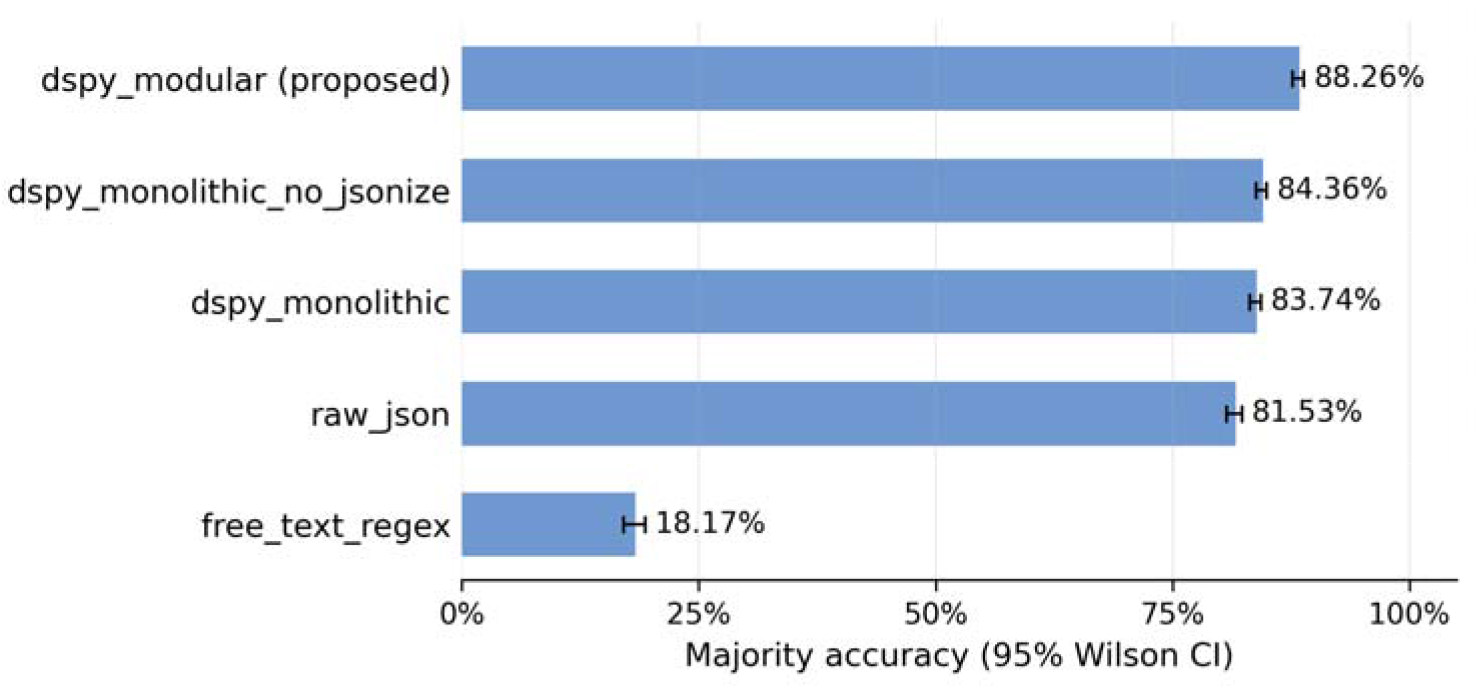
Component-ablation headline comparison across the five engineering cells.

The main qualitative conclusion was robust across the ablation grid: prompt-level schema discipline was load-bearing, as the schema-blind free-text baseline showed a large performance drop relative to the proposed pipeline. The DSPy framework was especially important for the chapter-5 lymph-node group extraction task, where removal of the framework in the rawjson variant markedly reduced micro-F1 and increased hallucination and miss rates. In contrast, several scalar field subsets showed smaller differences among the DSPy-or JSON-constrained variants. Detailed field-type stratification, nested-list error modes, matched-pair analyses, and the single-seed interpretation caveat for the stripped-down variants are provided in Supplementary Tables S45-S54 and Supplementary Section S1.3.1.

### 3.7. Inter-Annotator Agreement and Pre-Annotation Effect

The committee-resolved gold standard was constructed by two annotating pathologists, Kai-Po Chang and Nan-Haw Chow, with adjudication by a senior third pathologist, Han Chang. Under the proposed clinical workflow, in which both annotators weighted-mean Cohen’s κ of 0.844 across 41 categorical fields (n = 9,598 case-field pairings). At the highest-level report-triage organ-class decision across 12 categories, Cohen’s κ was 0.936 [0.919, 0.950], indicating high inter-annotator agreement (Figure 7; Supplementary Tables S56-S58). A secondary nominal Krippendorff s *a* cross-check was concordant at 0.968.

**Figure 7.**
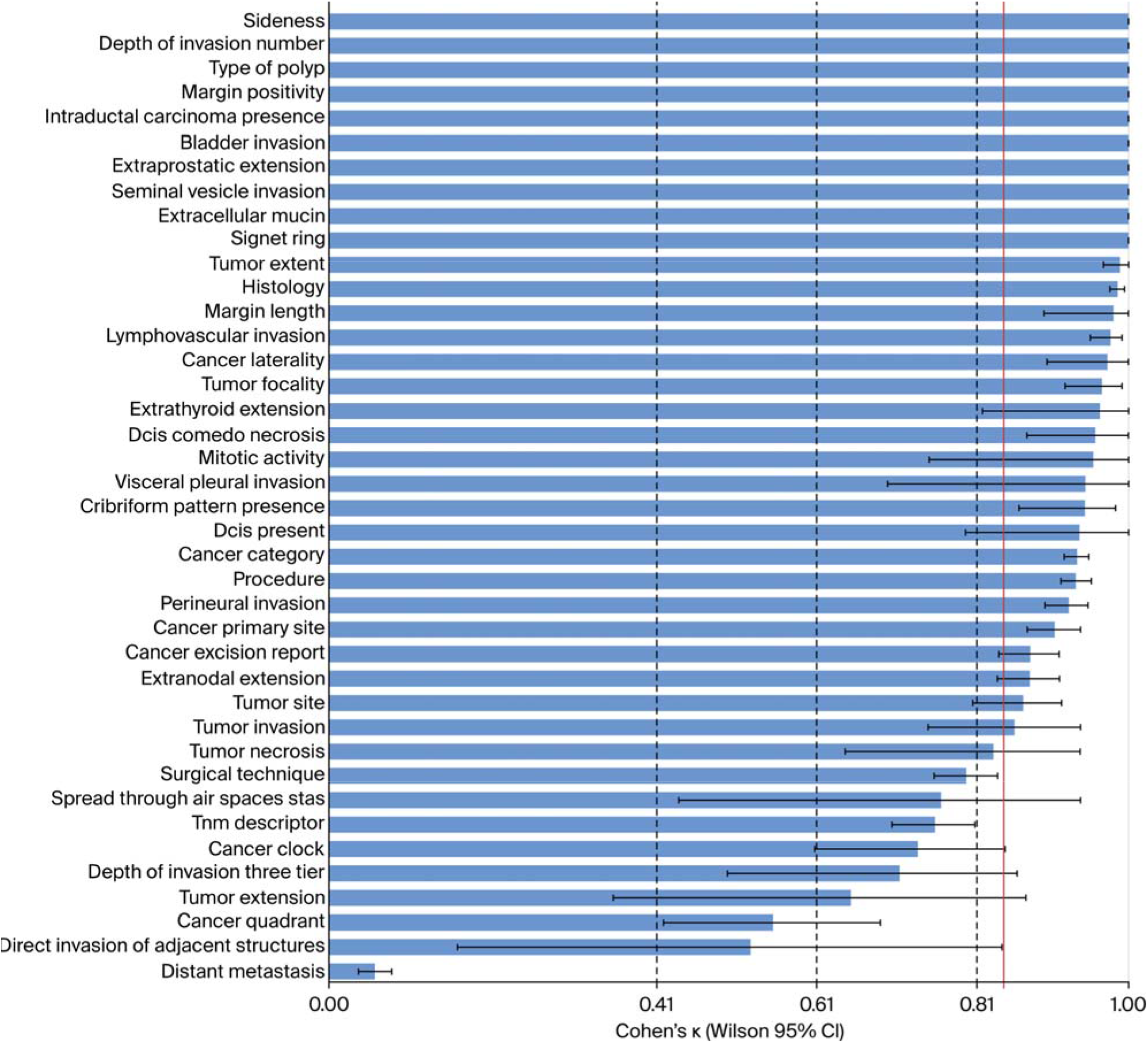
Per-field inter-annotator agreement (Cohen’s κ) under the operational workflow with 636 LLM seed, computed across the 41 categorical fields in scope.

To test whether LLM pre-annotation systematically anchored the annotators, both annotators independently re-annotated a stratified 196-case subset without the LLM seed. Across 93 annotator x organ x field cells with defined Δκ, the mean Δκ was +0.015 and the median was 0.0; 14 cells improved, 7 worsened, and 72 were unchanged (Supplementary Figure S16 and Supplementary Tables S59–S63). Thus, the headline κ would have been numerically similar without the LLM seed. Finally, adjudication fairness was assessed among cells with at least one KPC-NHC disagreement; no systematic bias toward either annotator was detected after Holm correction (Supplementary Table S57).

### 3.8. Multi-Run Reliability

Across the 30 independent inference runs of the gpt-oss-20b pipeline, run-to-run reliability of the upstream classification stages was very high. For eligibility classification, ICC(3, k = 30) was 0.9959 [0.9955, 0.9963], with single-run ICC(2,1) = 0.889 and a per-case flip rate of 3.6%. For organ classification, ICC(3,k) was 0.9963 [0.9959, 0.9967], with ICC(2,1) = 0.900 and a flip rate of 1.8%. These values indicate near-perfect reliability of the 30-run aggregate, while the single-run ICC and flip-rate values quantify the residual seed-to-seed variability.

Per-field reliability was computed on the same 71-field cascade-evaluation set used in the component-ablation analysis. This scope corresponds to the 192-cell Section 3.3 paper scope expanded by the separately reported biomarker and prostate-margin sub-fields described in Section 2.2.1. Thus, Sections 3.3, 3.6, and 3.8 use the same underlying inference outputs but differ in reporting denominator and aggregation target: Section 3.3 reports per-organ macro means after case-field majority aggregation, Section 3.6 reports component-ablation summaries, and Section 3.8 reports seed-to-seed reliability statistics.

For list-typed extractions, per-case F1 standard deviation was organ-dependent. The noisiest cells were prostate lymph-node groups (SD 0.40), cervix lymph-node groups (0.36), esophagus lymph-node groups (0.35), and lung margins (0.16), reflecting the same station-name canonicalization and anatomic-complexity gradients described in Section 3.4. Full per-organ per-field reliability results are provided in Supplementary Table S35 and Supplementary Figures S14–S15.

Accuracy intervals and multi-run reliability therefore answer distinct questions: Wilson intervals estimate uncertainty in majority-collapsed report-level or case-field-level accuracy, whereas ICC, Cronbach’s *α*, flip rates, and per-case run SD quantify seed-to-seed stability without treating the 30 runs as 30 independent clinical cohorts.

### 3.9. Multi-Run TCGA Cascade Evaluation (Full Schema, « =242 Reports)

We evaluated the proposed local-LLM pipeline on the full per-organ schema across 242 TCGA reports spanning breast, colorectal, esophagus, liver, stomach, and thyroid. The external cohort was processed under a 22-seed multi-run protocol using identical prompts and decoding parameters, with only the random seed varying across runs. The difference between the 30-run internal protocol and the 22-run external protocol was due to compute-budget constraints rather than a methodological difference. The TCGA evaluation scope included approximately 124 per-organ scalar field cells, corresponding to roughly 51 unique scalar field names across the six organs, and is therefore not directly comparable to the full ten-organ internal scope described in Section 2.2.1.

After report-level majority aggregation, eligibility triage reached 99.59% [95% Wilson CI: 97.70, 99.93], and organ classification reached 98.76% [96.40, 99.58], consistent with the internal cohort and indicating that the pipeline transported cleanly at the cascade-gating stages. At the per-field extraction stage, after case-field-level majority aggregation, the per-organ macro mean was 77.48% (Supplementary Table S36; Supplementary Figure S6), ranging from 84.77% [82.05, 87.13] in esophagus to 72.92% [70.33, 75.35] in colorectal. Pooled repeated-inference summaries are retained only as descriptive companions.

The gap between the internal per-organ macro mean of 92.03% and the external TCGA mean of 77.48% appears to be driven largely by TCGA source-documentation properties rather than by model capacity alone. Older TCGA breast reports often omit hormone-receptor workups; mismatch-repair immunohistochemistry is not consistently performed or reported across TCGA contributing institutions; AJCC stage cannot be reliably assigned without an edition statement, which most TCGA reports do not carry; and pathologists use different conventions for coding the absence of distant metastasis, such as pM0 versus Mx. Thirteen of the 51 evaluated fields were therefore tagged in Supplementary Table S38 with specific documentation caveats explaining their low effective accuracy in TCGA.

Excluding these 13 caveat-tagged fields raised case-field majority accuracy to 88.02% [86.99, 88.97] in a within-v2 sensitivity analysis. Both readings are reported because they answer different questions: 77.48% represents the full-schema external-validation headline, whereas 88.02% estimates performance after documentation-side caveats are set aside. The four-system comparison in Section 3.10 shows that the rule-based extractor, BERT encoder, local language model, and commercial API model all showed reduced performance on the same caveat-tagged fields, supporting the interpretation that the residual external-validation gap is largely a corpus-documentation property rather than a failure of any single extraction architecture.

### 3.10. Baseline Comparison on the External TCGA Cohort

Four systems were evaluated head-to-head on TCGA-Reports (n = 242 across breast, colorectal, esophagus, liver, stomach, and thyroid) under the same Stage A/B/C cascade defined in Section 2.4. Stage C scalar-field effective accuracies were local LLM, 77.02% [76.76, 77.27]; API LLM, 75.22% [74.74, 75.70]; rule-based, 65.16%; and BERT-merged, 40.06% (Figure 8; Supplementary Table S39). On variable-length list fields, the API LLM wins as expected (margins F1 0.831 vs. 0.783; lymph node F1 0.686 vs. 0.587). The local pipeline is operationally comparable to the closed-frontier API on scalar accuracy while keeping the cohort on-prem; the structural ceiling on rule-based and BERT-merged systems (which cannot emit nested variable-length structures by construction) explains why neither is a viable substitute for schema-rich extraction.

**Figure 8.**
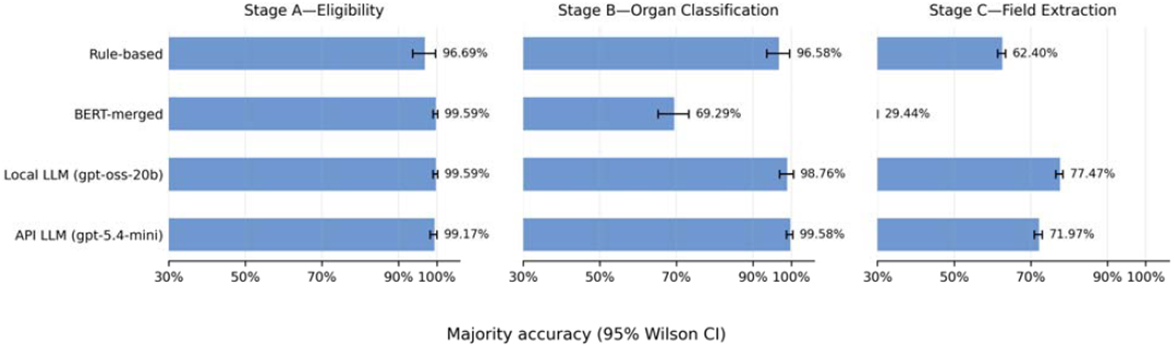
Baseline cascade comparison on the external TCGA cohort.

## 4. Discussion

This study demonstrates that a clinically governed ontology, paired with schema-bound local-LLM execution, can convert free-text surgical pathology reports into registry-grade structured data at scale. The pipeline achieved high case-field-level majority accuracy across ten malignancies and the 192-cell paper scope while remaining feasible on a single workstation-class GPU, supporting practical deployment for document triage, organ classification, and fine-grained abstraction.

Although LLMs now show strong performance on clinical extraction tasks, the field has faced both computational and representational barriers [24,34]. Our CAP-aligned ontology addresses the representational barrier by defining a reusable clinical target that can outlast rapid changes in model architecture, while the on-premises gpt-oss-20b deployment addresses the computational and privacy barriers. Separating schema logic from model weights also allowed the same clinical abstraction layer to be executed across different inference backends.

The four-system cascade comparison on the external TCGA cohort (Section 3.10) clarifies where the proposed local-LLM pipeline sits in the methodological landscape. On Stage-C scalar field extraction, the local pipeline reached 77.47%, compared with 71.97% for the commercial API reference, 62.40% for the rule-based extractor, and 29.44% for the BERT-merged encoder. These results support the value of schema-constrained language-model extraction over narrower rule-based or encoder-only approaches, while not establishing a categorical local-versus-commercial model ordering because the API and local configurations were not matched for context length or model family. On variable-length list fields, the commercial API retained an advantage, consistent with the need for further work on nested extraction. The BERT-merged system additionally illustrates an architectural limitation of encoder-only baselines: they do not natively emit nested variable-length structures required for schema-rich registry workflows.

A consistent and clinically meaningful failure mode involved the distinction between anatomic and pathologic stage groups under AJCC 8 prognostic staging. On the same 75 paired breast reports (Section 3.3.1), after case-field-level majority aggregation, anatomic_stage_group reached 98.67% [92.83, 99.76], whereas pathologic_stage_group reached 80.00% [69.59, 87.49], an 18.67 percentage-point gap. This gap is not primarily a stochastic model-disambiguation failure; rather, it decomposes into structural dimensions including post-treatment y-descriptor cases, HER2 2+ equivocal cases requiring FISH to finalize prognostic stage, and residual narrative ambiguity in non-y reports. The appropriate mitigations are therefore structural: AJCC 8 stage-table injection and a multi-valued pathologic_stage_group representation for indeterminate cases.

The current cancer-data schema models a single primary tumor per report, a deliberate choice aligned with the registry target of one extraction record per primary, but one that creates a structural blind spot for synchronous multi-primary disease. The Stage-B triage analysis in Section 3.2.1 quantified this directly: among 1,138 repeated-inference ledger entries involving gold-standard “others” dispositions, 449 (39.5%) were correctly triaged to “others”, 631 (55.4%) were misrouted into a single in-scope organ, and 58 (5.1%) were over-flagged. This motivates a flag-for-review workflow in the current revision and a future schema-cardinality extension to support multiple linked cancer-entity records when more than one primary tumor is present.

The bottom-decile audit after case-field majority aggregation supports a six-mechanism attribution rather than a single undifferentiated error category. Some residuals are likely closable by glossary or rule updates, such as heterogeneous reporting of surgical technique, tumor necrosis, cancer clock, and cancer quadrant. Others reflect schema-shape problems, including over-specified biomarker sub-records for HER2 and Ki-67, or AJCC convention issues such as pM0 versus Mx handling. Still others, such as liver procedure mapping and lymph-node station canonicalization, point to anatomic ontology gaps. Importantly, no bottom-decile pattern was driven by parse-error or schema-validation failure, supporting the reliability of the structured-output layer itself.

Hereditary and tumor-suppressor markers, including BRCA1, BRCA2, and TP53, are clinically important but were not present at meaningful rates in the surgical-resection narratives in either the CMUH cohort or the TCGA breast surgical-pathology subset used for external validation. These markers are typically documented in separate molecular-pathology workups under distinct reporting workflows. Adding them as schema fields without supporting source-document content would create structural false negatives. The appropriate next step is therefore to integrate molecular workups as a separately linked source layer with explicit provenance, rather than forcing these variables into the surgical-pathology extraction schema.

A key future direction is extending this framework into a multimodal system. The schema-first JSON design can accommodate multimodal extension without breaking existing fields: whole-slide image features can be added as a linked image-derived layer, and genomic data can be added as a sibling molecular layer with its own source identifiers, extractor versions, and timestamps. Pathology-tuned vision models, including recent vision-language systems [36], could therefore be incorporated as additional extractors without rewriting the clinical ontology. In this design, the CAP-aligned clinical core remains the durable anchor, while new modalities attach as provenance-preserving layers.

The limitations of this study include the single-institution internal development cohort, the current single-primary schema cardinality, source-documentation sensitivity in the TCGA cohort, and unresolved model-side semantic conflation for a small subset of fields. The next validation extension should therefore include a federated multi-institutional cohort, explicit multi-primary representation, and linked molecular-pathology and image-derived data sources.

## 5. Conclusions

By combining a College of American Pathologists (CAP)-aligned clinical ontology with schema-bound local large-language-model execution, this work provides a practical route from narrative surgical-pathology reports to interoperable, machine-readable cancer data. The durable contribution is the ontology and schema-first abstraction layer, rather than any single model family: it preserves clinically meaningful structure, enables privacy-preserving local deployment, and provides a portable foundation for future multimodal and real-time cancer data infrastructures.

## Supplementary Materials

Hie following supporting information can be downloaded at https://www.mdpi.com/article/doi/s1. Supplementary materials referenced in the manuscript include Supplementary Section S1 (Extended Methods Section S1.1. Annotation Protocol; Section S1.2. Baseline Implementations; Section S1.3. Statistical Framework; and Section S1.4 Decoding/Hardware), Supplementary Tables S1-S63, Supplementary Figures S1-S16, and Supplementary Appendices A1-A10 (case studies and in-depth information complementing the summaries in Section 4).

## Author Contributions

Conceptualization, K.-P.C.; methodology, K.-P.C., H.-K.C., and P.-C.C.; software, H.-K.C., C.-Y.L., and Y.-L.L.; validation, H.-K.C., P.-Y.T., N.-H.C., and H.C.; formal analysis, H.-K.C. and Y.-W.C.; investigation, N.-H.C., H.C., and K.-P.C.; resources, N.-H.C. and H.C.; data curation, P.-Y.T.; writing–original draft preparation, K.-P.C.; writing–review and editing, K.-P.C., P.-C.C., Y.-W.C., and L.-J.S.; supervision, N.-H.C., H.C., and K.-P.C.; funding acquisition, K.-P.C. and H.-K.C. All authors have read and agreed to the published version of the manuscript.

## Institutional Review Board Statement

This study was conducted in accordance with the Declaration of Helsinki and approved by the Research Ethics Committee of China Medical University & Hospital (protocol code CMUH114-REC2-037).

## Informed Consent Statement

Patient consent was waived by the Research Ethics Committee of China Medical University & Hospital due to the retrospective nature of the study and the use of de-identified datasets.

## Data Availability Statement

The internal development cohort consists of de-identified surgical pathology reports from China Medical University Hospital and is not publicly available because of patient privacy protections. The external validation cohort, including the 242 selected TCGA pathology reports, expert-annotated ground-truth labels, and model outputs, has been deposited in Zenodo (https://doi.org/10.5281/zenodo.17689361). The original TCGA source data remain accessible through the NCI Genomic Data Commons Data Portal. The source code for the Digital Registrar extraction pipeline is openly available at https://github.com/kblab2024/digitalregistrar.

## Acknowledgments

We are deeply grateful to Ting-Sung Hsieh of UT Southwestern Medical Center, and Ching-Hang Chien and Yi-Chung Dzeng of the Research Center for Applied Sciences, Academia Sinica. During the preparation of this manuscript, the authors used OpenAI GPT-5 for non-confidential language editing and limited drafting assistance on selected textual passages. The authors reviewed and edited the output and take full responsibility for the content of this publication.

## Disclaimer/Publisher’s Note

The statements, opinions and data contained in all publications are solely those of the individual author(s) and contributor(s) and not of MDPI and/or the editor(s). MDPI and/or the editor(s) disclaim responsibility for any injury to people or property resulting from any ideas, methods, instructions or products referred to in the content.

