## supplementary data for "Digital Registrar: A Schema-First Framework for Multi-Cancer Privacy-Preserving Pathology Abstraction via Local LLMs"

Manuscript ID: diagnostics-4271493

This document compiles every supplementary artefact referenced by the revised manuscript: the §S1 Extended Methods sub-archive (S1.1–S1.4), Supplementary Tables S1–S64, Supplementary Figures S1–S16 (rendered inline), and Supplementary Appendices A1–A10. Cross-references in the main text and in the response-to-reviewers letters resolve here. All numerical content derives from the canonical CSV outputs at paper-revision/code/derived/*.csv, regenerated by paper-revision/code/analysis.py from the evaluation results released on 2026-05-14 (git SHA 3818a004).

**Index**

**Extended Methods (§S1)** — S1.1 Annotation Protocol; S1.2 Baseline Implementations; S1.3 Statistical Framework; S1.4 Decoding and Hardware

**Supplementary Tables S1–S10** — Per-organ schemas

**Supplementary Tables S11–S13** — Organ-classification confusion and stability (30-run aggregate)

**Supplementary Tables S14–S23** — Per-field accuracy by organ (30-run aggregate)

**Supplementary Table S24** — Lymph-node concordance (v1 carryover)

**Supplementary Tables S25–S26** — §3.2 Triage and organ-classification 30-run aggregates

**Supplementary Table S27** — §3.3 Per-organ extraction headline (Table 5 / Figure 3 backing)

**Supplementary Table S28** — §3.3.1 Anatomic vs pathologic stage_group (Figure 4 backing)

**Supplementary Tables S29, S64** — §3.3.2 Bottom-decile field error attribution

**Supplementary Tables S30–S31** — §3.2.1 Multi-primary triage

**Supplementary Tables S32–S34** — §3.4 Margins and lymph nodes

**Supplementary Table S35** — §3.8 Multi-run reliability

**Supplementary Tables S36–S38** — §3.9 TCGA external validation

**Supplementary Tables S39–S43** — §3.10 Baseline cascade comparison

**Supplementary Tables S44–S54** — §3.6 Component ablations

**Supplementary Tables S55–S63** — §3.7 Inter-annotator agreement and pre-annotation effect

**Supplementary Figures S1–S16** — Rendered visual companions (PNG embeds)

**Supplementary Appendices A1–A10** — A1–A7 case studies; A8–A10 §4 Discussion deep dives

**Section S1. Extended Methods**

**Supplementary §S1.1 — Annotation Protocol (Extended Methods)**

This file expands main-body §2.1 (Data Source and Annotation) with the detailed annotation-committee protocol, the without-LLM-seed bias-control arm design, the senior-pathologist adjudication audit-trail mechanics, and the annotation tooling reference. The main-body §2.1 carries the cohort table, the high-level committee structure (annotators + adjudicator), and a one-sentence pointer to this file for the full detail.

**S1.1.1 Reference Standard Construction (full detail)**

The gold standard used throughout §3 was produced by a two-pathologist annotation committee — **Kai-Po Chang (KPC)** and **Nan-Haw Chow (NHC)**, both co-authors — with adjudication by a senior third pathologist, **Han Chang**, also a co-author. The senior-pathologist adjudication role has been part of our annotation workflow throughout the project; for this revision the annotation pipeline was re-run from scratch under a dedicated GUI annotation tool that retains the per-annotator paired record (enabling inter-annotator agreement statistics) and records the senior pathologist's adjudication decisions as a persistent audit trail. The committee design, the without-LLM-seed bias-control arm described below, and the audit-trail persistence together address Reviewer 1's same-model-bias concern (R1.b) and Reviewer 2's two-reviewer-panel concern (R2.3-IAA).

**S1.1.2 Without-LLM-seed bias-control arm**

To directly test the same-model-bias concern raised in R1.b — that annotators seeded with the candidate model's output may unconsciously anchor on its predictions, inflating apparent system performance — both pathologists completed an independent **without-LLM-seed re-annotation pass** on a stratified 196-case subset (~22 % of the validation cohort, balanced across organs). In this arm, the source pathology report was presented without the LLM pre-annotated JSON draft; each pathologist produced a labelled record from scratch. The per-pathologist labels in this arm are persisted independently by the annotation tool. The per-organ allocation of the 196-case subset is shown in Table S1.1.1 below; the stratification preserves the cohort-balance properties used in §3.3.

**Table S1.1.1. Per-organ allocation of the 196-case without-LLM-seed bias-control subset.**

| **Organ site** | **Number of paired re-annotations (n)** |
| --- | --- |
| Esophagus | 19 |
| Stomach | 21 |
| Colorectum | 18 |
| Breast | 20 |
| Uterine cervix | 20 |
| Pancreas | 17 |
| Prostate | 17 |
| Lung | 19 |
| Liver | 20 |
| Thyroid | 20 |
| Others (dual or out-of-scope cancers) | 5 |
| **Total** | **196** |

The Δκ analysis comparing with-seed and without-seed inter-annotator κ (mean Δκ = +0.015, median 0.0 across the 93 (annotator × organ × field) cells with defined Δκ) is reported in §3.7. Δκ is computed under a strict **paired-annotator pairing constraint** — for each cell, the same partner annotator is used in both arms (KPC-with-seed × NHC-with-seed vs KPC-without-seed × NHC-without-seed, on the 196-case intersection) — so the κ baseline is held fixed across arms and only the seed-arm marginal varies. Implementation: evaluation_code/docs/eval/preann_effect.md. This confirms that the pre-annotation seed does not systematically anchor the annotators on the LLM's output.

**S1.1.3 With-LLM-seed primary arm (operational workflow)**

Both pathologists also re-annotated all 893 reports independently in the with-LLM-seed arm — the operational workflow the manuscript proposes for downstream clinical deployment, in which the gpt-oss-20b single-pass JSON output is presented as the editable draft and the pathologist edits in place. Each pathologist produced their own labelled record under the new annotation tool. The committee-resolved version of these new paired re-annotations — produced by senior-pathologist adjudication of residual disagreements — forms the gold reference used throughout §3. The original-submission consensus labels are not used in any §3 result.

**S1.1.4 Why the new annotation pass was necessary**

The original-submission protocol described two pathologists independently reviewing and correcting the LLM pre-annotation in a side-by-side view, with discrepancies resolved through blinded consensus. That side-by-side view was an ad-hoc two-pane editor setup, not a dedicated annotation tool, and the workflow was designed to converge on a single committee-resolved gold label per case; a v1 disagreement log was retained for use during adjudication, but a discrepancy log is the wrong artifact for κ at this schema's scale. Cohen's κ is defined on the full paired record — the chance-agreement baseline depends on the marginal distribution of *all* labels each annotator emitted, not just the cells where they disagreed — and at 41 categorical scalar fields per report on the with-seed κ scope, plus three nested variable-length list fields (margins[*], regional_lymph_node[*], and biomarkers[*]) carrying their own internal sub-object structure entirely outside that count, even raw per-annotator JSON dumps would still need a substantial schema-aware canonicalization pipeline before any cell-by-cell κ could be computed. The new annotation tool was engineered to persist each annotator's labels directly in canonical form, eliminating that retrospective step and producing the paired record on which §3.7's κ is computed.

**S1.1.5 Adjudication: same human process, newly persisted audit trail**

Han Chang's adjudication role is not new to this revision — disagreements between KPC and NHC have always been resolved in adjudication meetings with him as the senior pathologist. What is new is that for this revision, every consensus-resolving interaction during adjudication is recorded by the annotation tool as a persistent, structured artifact: (case, field, KPC value, NHC value, adjudicator's resolution). For each (organ, field) cell with at least one disagreement, the adjudicator's resolutions are tabulated as a 2 × 2 contingency table on the (matches_KPC, matches_NHC) outcomes (where a "matches" outcome means the adjudicator's value equals that annotator's submitted value, and a "matches both" cell is omitted because there is no disagreement to resolve). A per-cell **χ² uniformity test against the equal-probability null** (50 % KPC-favoured, 50 % NHC-favoured) is then computed (df = 1; exact-binomial fallback when n_disagreements < 10); p-values are Holm-corrected across the 230 (organ, field) cells. The null is retained on the great majority of cells — the adjudicator's resolutions are uniform across (matches_KPC, matches_NHC), with no systematic bias toward either annotator. Full table: Supplementary Table S58. The original-submission manuscript wording "final blinded consensus" did not credit Han Chang explicitly; this is corrected here by naming all three pathologists and by persisting the adjudication decisions as auditable data.

**S1.1.6 Annotation Tooling**

The dedicated GUI annotation tool built for this revision is open source at <https://github.com/kblab2024/digital-registrar-research/tree/main/src/digital_registrar_research/annotation>. The tool is structured as two complementary applications. app_canonical.py is the per-annotator labelling app: each pathologist runs their own instance; the app presents the source pathology report and an editable JSON view in a single window, supports both with- and without-LLM-seed arms (the JSON view can be initialized empty or pre-populated with the gpt-oss-20b single-pass output), and persists each annotator's labels as an independent record in canonical form keyed by annotator identity. compare_app.py is the adjudication and consensus-building app, used by the senior pathologist to resolve disagreements between KPC and NHC; every consensus-resolving interaction is recorded by the app as a persistent log entry, producing the structured adjudication audit trail that backs Supplementary Table S58. Implementation detail (UI layout, hotkeys, persistence schema) is available in the open-source source code.

**Cross-references**

Cohen's κ is defined and discussed as the gold-standard validity metric in §2.4 (Evaluation Protocol and Statistical Methods) and Supplementary §S1.3. The headline inter-annotator agreement on the committee, the Δκ analysis on the without-LLM-seed arm, and the per-(organ, field) committee-adjudication audit are reported in §3.7 (¶22 in the new paragraph numbering — Inter-Annotator Agreement and Pre-Annotation Effect). All per-field Cohen's κ values, the full 8-pair κ matrix, the Krippendorff's α cross-check, and the per-(annotator, organ, field) Δκ tables are in Supplementary Tables S55–S63.

**Supplementary §S1.2 — Baseline System Implementations (Extended Methods)**

This file expands main-body §2.6 (Baseline Systems for Cross-Architecture Comparison) with the detailed implementation specifications for each of the four reference systems benchmarked on the TCGA external cohort. Main-body §2.6 retains the high-level rationale (four-system comparison: rule-based, encoder, local LLM, API LLM) and the result-anchor pointer to §3.10; this file holds the implementation specifics that reviewers may need for reproduction.

**S1.2.1 Why TCGA, why these four systems**

To contextualise the proposed local-LLM pipeline against established methods, four systems were benchmarked head-to-head on the TCGA external cohort (§2.5). TCGA is the only corpus that admits a fair four-way comparison — the closed-frontier API model cannot be sent CMUH data for privacy reasons, and the BERT baseline was fine-tuned on CMUH so predicting on CMUH would leak training cases. All four systems consumed the same plain-text reports and emitted the same per-organ schema; all were scored under the identical Stage A/B/C cascade defined in §2.4.

**S1.2.2 Rule-based system (deterministic floor)**

A deterministic regex- and lexicon-driven extractor implemented in `benchmark_code/baselines/rules.py`, spanning ten organ schemas. Per-organ tactics include lexicon majority-vote for organ classification, longest-match enums for TNM, organ-aware grade extractors (Gleason → ISUP for prostate; Nottingham sub-scores for breast), and numeric span extractors with unit normalization. Coverage policy is omit-on-no-signal: a field is emitted only when a regex / lexicon match clears its confidence threshold. Full inventory: evaluation_code/docs/benchmarks/06_methods.md.

**S1.2.3 BERT-merged system (encoder-only reference)**

emilyalsentzer/Bio_ClinicalBERT as a shared encoder behind a multi-head linear classifier (`clinicalbert_cls.py`; one head per categorical or boolean field, with an explicit null class) and a SQuAD-style extractive QA head for numeric span fields (`clinicalbert_qa.py`). Fine-tuned on the full CMUH annotation pool (5 epochs CLS head, 3 epochs QA head) and predicted on the held-out TCGA cohort — the only fair training configuration given privacy and disjointness constraints. Nested variable-length list fields cannot be emitted from a CLS + QA architecture (n_attempted = 0 by construction); this is a structural ceiling. Hyperparameter detail: evaluation_code/docs/benchmarks/02_train_bert.md.

**S1.2.4 Local LLM (gpt-oss-20b via DSPy + Ollama)**

The proposed system. OpenAI gpt-oss-20b is served by Ollama with llama.cpp grammar-constrained decoding; DSPy Signature declarations encode each per-organ schema as Literal[...] enums and typed nested lists, which the DSPy adapter compiles to a JSON Schema that Ollama (≥ 0.5) converts to a GBNF grammar masking tokens that would violate the schema at sample time. Inference runs on-prem; no PHI leaves the institutional perimeter. Serving stack and grammar pipeline: evaluation_code/docs/dspy_deep_dive.md §1, evaluation_code/docs/pipeline.md.

**S1.2.5 API LLM (gpt-5.4-mini via OpenAI)**

gpt-5.4-mini accessed through the OpenAI API as a closed-frontier reference, reusing the same DSPy Signatures but routing constrained decoding through OpenAI's native response_format={"type": "json_schema"} mode rather than llama.cpp GBNF. Retry-on-parse-error is enabled; decoding parameters are pinned by the project's MODEL_PROFILES config. For this benchmark, the commercial API was called with num_ctx = 4,096 while the local model was served with num_ctx = 16,384 — a **4× asymmetry favouring the local model** that we did not have time to rebalance in this revision. The discussion of what this asymmetry does and does not establish is in §S1.2.7 below. API-call shape: evaluation_code/docs/benchmarks/03_run_baselines.md.

**S1.2.6 Cascade evaluation and error propagation**

All four systems were scored under the three-stage cascade defined in §2.4 — Stage A (eligibility triage), Stage B (organ classification), Stage C (per-organ field extraction). A Stage-A or Stage-B failure halts the cascade for that attempt; no downstream stages are scored and the attempt counts as the failed stage's error in the cascade-level denominator. Stage-C per-attempt accuracy is computed only on attempts that survive both gating stages, and the per-attempt denominators read identically across systems on shared reports. Nested-list fields are scored separately under list-F1 (§2.5) and are not pooled into the Stage-C scalar denominator. Stage-wise audit detail: E_baseline_cascade_comparison.md §§3–4.

**S1.2.7 Cross-architecture comparison: what the result does and does not establish**

The four-system experiment was designed to answer **R1.a — methodological baseline contextualisation**. The two methodologically load-bearing baselines are the deterministic rule-based extractor (engineering floor: cannot reach fields outside its regex inventory) and the BERT encoder (representational ceiling: cannot emit variable-length nested structures by construction); the local language model clears both at per-field extraction, which is the comparison the reviewer's request anchors on.

A fourth contextualising arm — the commercial gpt-5.4-mini API — was added because the reviewer named "GPT-4" alongside the methodological baselines. Closed-frontier commercial models are not, strictly, methodological baselines in the encoder-vs-rule-vs-language-model sense; they are a comparison against a different deployment regime (off-prem, paid, rate-limited) rather than a different model class. We did not configure that comparison for a balanced head-to-head:

• The local model was served with num_ctx = 16,384 while the commercial API was called with num_ctx = 4,096 (a 4× asymmetry favouring the local model — pathology reports plus the per-organ output schema together routinely exceed 4 K tokens). This was an authors' configuration decision and not rebalanced before the in-revision deadline.

• The commercial system used was gpt-5.4-mini, which is the smaller, faster sibling of a frontier commercial family, not the full gpt-5.4 variant.

**What the cross-architecture experiment does establish:**

**1.** All four systems collapse on the same set of TCGA caveat-tagged fields (Supplementary Table S38), confirming that the residual external-validation gap reported in §3.9 is a property of the corpus rather than of the extraction model. This is the load-bearing finding for R1.e.

**2.** Rule-based and BERT have structural ceilings that no amount of additional training data can close without changing the architecture — the rule-based system because its regex inventory cannot be exhaustive over the schema, the BERT encoder because its classification + extractive-QA architecture has no representational primitive for nested variable-length structures (Supplementary Table S43).

**3.** The local language model clears the two methodological baselines by a wide margin at per-field extraction.

**What the cross-architecture experiment does not establish:**

• A categorical accuracy ordering between the local language model and frontier commercial language models. The local-vs-commercial result was: local ahead on per-field scalar extraction (Supplementary Table S39); commercial ahead on the two nested list fields (Supplementary Table S43). This scalar-vs-list divergence cannot be cleanly attributed to context-window capacity (the local model had 4× more context), and the smaller-sibling configuration on the commercial side leaves open whether the full commercial variant would lead on both surfaces.

**Three discriminating experiments deferred to future work** (each contingent on resources not available within this revision):

**1.** A matched-context API rerun at num_ctx = 16,384 on the same 242 reports.

**2.** A comparison against the full gpt-5.4 (not the mini variant).

**3.** An open-weights comparison against gpt-oss-120b — the larger sibling of our local 20 B model — under the same protocol and a comparable GPU footprint.

The paper's primary contribution is operational viability for on-prem registry extraction under a privacy constraint (no patient text leaving the institution), which does not depend on the local-vs-commercial accuracy ordering. The cross-architecture comparison is reported because it answers R1.a and supports R1.e; it is not framed as a categorical accuracy claim.

**S1.2.8 Corpus-side OCR caveat (TCGA-Reports)**

The TCGA-Reports release we use is OCR-derived from the original PDF reports via Amazon Textract with custom post-processing. The corpus authors (Kefeli & Tatonetti, 2024) explicitly describe the result as **"moderately curated"** and acknowledge residual spelling errors (introduced either in the original text or during OCR), systematically mis-translated handwriting (dropped at corpus-release time), and exclusion of multiple-choice and synoptic report formats from the released plain text. This corpus-side noise contributes an unquantified component of the per-field extraction residual error in §3.9 / §3.10. It affects **all four benchmarked systems symmetrically** — rule-based, BERT, local language model, and commercial API all operate on the same Kefeli & Tatonetti plain-text release — so it does not bias the cross-architecture ranking. Quantifying this source-text-quality contribution would require a character-level audit of the OCR-cleaned text against the source PDFs and is outside the scope of this revision; the main-body external-validation paragraph (§2.5, ¶03) carries a one-sentence pointer to this subsection.

**Supplementary §S1.3 — Statistical Framework (Extended Methods)**

This file expands main-body §2.4 (Evaluation Protocol and Statistical Methods) with the detailed statistical formulas, multiple-comparison protocols, GLMM specification, and library version pinning. Main-body §2.4 carries the multi-run protocol summary (k = 30 internal, k = 22 external), the Wilson / McNemar / ICC / Krippendorff α framework names, Table 2 (metrics definitions), and a one-sentence pointer to this file. The deep derivations and library calls live here.

**S1.3.1 Component Ablation Protocol**

**S1.3.1.1 Operational definitions and cell composition**

Two operational definitions feed the ablation grid. **DSPy `Literal` typing** is the use of typing.Literal[...] enum annotations inside dspy.Signature field declarations; the DSPy adapter consumes these annotations to emit a JSON Schema enum constraint that the downstream constrained-decoder enforces. **The `ReportJsonize` pre-pass** is the upstream DSPy signature that converts raw pathology-report paragraphs into a semi-structured JSON-like summary (report_jsonized) before the downstream extraction signature runs. The cells dspy_monolithic and dspy_monolithic_no_jsonize differ only on whether this pre-pass runs; full definitions are in evaluation_code/docs/dspy_deep_dive.md §1 and evaluation_code/docs/pipeline.md.

To quantify the contribution of the pipeline's engineering choices (per-organ decomposition, the ReportJsonize pre-pass, DSPy Literal typing, schema-constrained generation), a five-cell lesion study was run on the same internal CMUH cohort against the same committee-resolved gold (§2.4.2). The five cells span the proposed pipeline (dspy_modular, 10 multi-seed runs) and four single-axis lesions (dspy_monolithic, dspy_monolithic_no_jsonize, raw_json, free_text_regex). Compute budget restricted the four non-modular cells to one seed each; the within-cell stochasticity is bounded by the Fleiss κ across the modular cell's ten seeds (Supplementary Table S51). Headline results are reported in §3.6. The design rationale — non-orthogonality of the named axes, which matched-pair effects the grid does and does not cleanly isolate, the seed-count asymmetry, and the lesion-study framing — is documented in §S1.3.1.2–§S1.3.1.6 below.

**S1.3.1.1.1 Cell-by-cell mechanical definitions**

This subsection is the authoritative mechanical reference for each ablation cell. The full variant–component matrix, the per-variant descriptions in §3.6 and rebuttal R1c, and the per-stage rollup in Supplementary Table S54 all resolve here. Runner-level implementation paths are listed below; the canonical CMUH ablation manifest is manifest.json for the complete ablation result tree (git SHA ba35efb).

**Variant–component matrix.** ✓ = on, ✗ = off, n/a = not applicable.

| **Variant** | **Per-organ decomp** | **DSPy framework** | **ReportJsonize pre-pass** | **DSPy Literal typing** | **OpenAI json_object mode** | **Post-hoc jsonschema + retry** | **Regex + fuzzy extractor** |
| --- | --- | --- | --- | --- | --- | --- | --- |
| ------------------------------- | :-: | :-: | :-: | :-: | :-: | :-: | :-: |
| dspy_modular (proposed) | ✓ | ✓ | ✓ | ✓ | n/a | n/a | ✗ |
| dspy_monolithic | ✗ | ✓ | ✓ | ✓ | n/a | n/a | ✗ |
| dspy_monolithic_no_jsonize | ✗ | ✓ | ✗ | ✓ | n/a | n/a | ✗ |
| raw_json | ✗ | ✗ | ✗ | ✗ | ✓ | ✓ | ✗ |
| free_text_regex | ✗ | ✗ | ✗ | ✗ | ✗ | ✗ | ✓ |

• **`dspy_modular`** (runners/dspy_modular.py) — the proposed pipeline. Per-organ specialisation is realised as 5–7 chained dspy.Signature calls per organ; the ReportJsonize signature (models/common.py, class ReportJsonize) is invoked first to convert raw paragraphs into a semi-structured JSON summary that is passed as report_jsonized input to the downstream extraction signatures; output fields use Literal[...] enum annotations that the DSPy adapter compiles into a constrained-decoder enum constraint. Outputs are returned as Pydantic-typed instances and serialised by dump_prediction_plain(). No explicit retry budget — DSPy handles backtracking on schema violations internally per its framework defaults.

• **`dspy_monolithic`** (runners/dspy_monolithic.py, MonolithicPipeline with skip_jsonize=False) — collapses the per-organ specialisation. The 50–80 organ-specific output fields are merged into a single dspy.Signature per organ by signatures/monolithic.py::get_monolithic_signature(), which iterates over the per-subsection signature classes (e.g., BreastCancerNonnested, BreastCancerStaging, …) and merges their output fields into one class. ReportJsonize and Literal-typed extraction are kept.

• **`dspy_monolithic_no_jsonize`** (runners/dspy_monolithic_no_jsonize.py, MonolithicPipeline with skip_jsonize=True) — additionally skips the ReportJsonize pre-pass. When skip_jsonize=True the forward() method passes an empty {} as the report_jsonized input rather than invoking the ReportJsonize signature on the paragraphs. The monolithic signature therefore reads the raw paragraphs directly.

• **`raw_json`** (runners/raw_json.py) — removes the DSPy framework entirely. The OpenAI client is instantiated directly (from openai import OpenAI), invoked with response_format={"type":"json_object"} (falls back to prompt injection if the backend does not support JSON mode), and the schema appears in the prompt as a plain-text bulleted field-list (flatten_schema_for_prompt() in schemas/schema_builder.py) rather than as Pydantic introspection of dspy.OutputField instances. Outputs are validated post-hoc by jsonschema.Draft202012Validator via validate_cancer_data(); if validation errors are present and fewer than 20, the call is retried once with the error list appended to the user message. The _schema_errors field is set on the output when validation fails after retry; the validation_retries counter is tracked in the ledger.

• **`free_text_regex`** (runners/free_text_regex.py, extractor extractors/regex_per_field.py::RegexExtractor) — removes schema discipline as well. The model receives a prose-only system prompt asking it to summarise the pathology report in plain English ("the cancer site, the tumour size, histologic grade, TNM stage (pT/pN/pM), the presence or absence of lymphovascular and perineural invasion, surgical margins, lymph-node involvement, and any biomarker results … Do NOT use JSON or bullet points — write it as natural prose"). The model output is a single prose paragraph with no schema awareness. The RegexExtractor applies a fixed bank of 20+ regex patterns over the prose for primary endpoints (tumor size, pT/pN/pM, grade, LVI/PNI, biomarker presence) and a fuzzy token-Jaccard ≥ 0.6 match against per-organ enum vocabularies for secondary categorical fields. Organ is inferred via keyword-frequency over an organ-specific lexicon. There is no retry, no JSON validation, and no list-emitting code path; variable-length structured-list fields (e.g., involved_margin_list) have n_attempted = 0 because the extractor produces no list outputs.

**S1.3.1.1.2 Chapter-3 field-type categories and the chapter-4 / 5 list-level evaluation**

The component-ablation analysis in §3.6 and Supplementary Table S54 stratifies the cascade evaluation along the field structure dimension. This is *not* labelled with letter codes (A/B/C/…) to avoid collision with the cascade-pipeline "Stage A / Stage B / Stage C" terminology established in §2.4 and §2.5 (eligibility classification / organ classification / per-organ field extraction). Instead, the four categories are referred to in the upper-layer prose by plain-language names; their definitions are pinned here.

**Chapter-3 scalar field types.** Every chapter-3 in-scope field (62 fields, code/derived/field_complexity_stratification.csv) is assigned to exactly one of three types, fixed in paper-revision/code/analysis.py::FIELD_COMPLEXITY_STAGE:

• **boolean / single-flag** — the value is a single yes/no answer to a presence question. 20 fields, including lymphovascular_invasion, perineural_invasion, vascular_invasion, dcis_present, intraductal_carcinoma_presence, extranodal_extension, extrathyroid_extension, extraprostatic_extension, seminal_vesicle_invasion, visceral_pleural_invasion, bladder_invasion, tumor_invasion, tumor_necrosis, signet_ring, tumor_budding, spread_through_air_spaces_stas, direct_invasion_of_adjacent_structures, cribriform_pattern_presence, extracellular_mucin, and margin_positivity (a derived bool over the margin object list).

• **rigid enum (single-value categorical)** — the value is a single token drawn from a fixed vocabulary defined either by the AJCC 8th-edition staging system, by the WHO histology classifications, or by the per-organ enum vocabularies in the schema. 32 fields. This category is *not* split further — TNM categories (pt_category, pn_category, pm_category, tnm_descriptor, anatomic_stage_group, pathologic_stage_group, stage_group, overall_stage, distant_metastasis), histology / grade / laterality (histology, grade, nuclear_grade, cancer_laterality, sideness, tubule_formation, total_score, mitotic_rate, mitotic_activity, dcis_grade, dcis_comedo_necrosis, dcis_size, depth_of_invasion_three_tier), procedure / technique (procedure, surgical_technique, type_of_polyp, tumor_focality, tumor_extent, tumor_site, tumor_extension, cancer_primary_site, cancer_quadrant, cancer_clock) are all rigid enums in this sense — each value space is a finite, machine-defined vocabulary that the schema enforces.

• **numerical** — the value is a number (possibly with an implicit unit). 9 fields, including tumor_size, prostate_size, prostate_weight, gleason_4_percentage, gleason_5_percentage, tumor_percentage, depth_of_invasion_number, margin_length (numeric distance within the margin object list), and maximal_ln_size (numeric count over the LN-group list).

The chapter-3 involved_margin_list field is a chapter-3 summary view of the canonical chapter-4 evaluation and is *excluded* from the per-field-type aggregation in S54a. It remains in ablation_per_field.csv and field_complexity_stratification.csv as a per-field row for visibility.

**Chapter 4 (surgical margins) and chapter 5 (lymph-node groups) list-level evaluation.** These two cascade chapters score variable-length lists of structured objects and are the canonical evaluation of nested-list extraction. Both apply the per-element scoring rule defined in §2.4.1: predicted and gold lists are aligned greedily by canonical category (margin canonical label or LN canonical station), ties broken by Jaccard similarity on the free-text sub-fields (margin description or station name), and a matched element scores correct only when every sub-field exact-matches the aligned gold element. The chapter-level micro F1 reported in S54a and S54b/c is computed by pooling true positive (tp), false positive (fp), and false negative (fn) counts across the 10 organs (with the per-(method × organ) breakdown, then computing micro F1 = 2 tp / (2 tp + fp + fn), hallucination rate = fp / (tp + fp), and miss rate = fn / (tp + fn) on the pooled counts.

The chapter-4 margin object has three attributes (canonical margin category, involved bool, optional distance in millimetres); the chapter-5 LN-group object has four (canonical anatomical station, side, examined count, involved count). This single-attribute difference between the two list-level schema elements is the load-bearing structural difference behind the asymmetric raw_json collapse pattern reported in §3.6 and S54: chapter 4 absorbs DSPy removal almost intact, chapter 5 collapses by 31.6 pp F1 on the pooled denominator and by 49–70 pp on four individual organs.

**S1.3.1.2 Non-orthogonality of schema, DSPy, and prompting**

A naïve reading of "schema constraints, DSPy, prompting" treats these three as independently manipulable axes. In the pipeline studied here, they are not:

• The schema is realised through DSPy Literal[...] type hints in dspy.Signature declarations. There is no "schema without DSPy" cell — removing DSPy and retaining the schema would require a re-implementation of the constrained-output mechanism in a different framework (e.g., guidance, outlines, instructor), which would confound the engineering-choice question with a framework-substitution question.

• The prompting itself is partially produced by DSPy's compilation step, so there is no "DSPy without DSPy-style prompts" cell.

• The ReportJsonize pre-pass can be ablated within the monolithic DSPy regime — which is what the dspy_monolithic_no_jsonize cell does.

The five cells in the grid are therefore the cleanest set of single-component lesions the pipeline architecture admits:

| **Cell** | **Per-organ decomposition** | **DSPy + Literal typing** | **ReportJsonize pre-pass** | **Constrained generation** |
| --- | --- | --- | --- | --- |
| **`dspy_modular`** (proposed) | ✓ | ✓ | ✓ | ✓ |
| dspy_monolithic | ✗ | ✓ | ✓ | ✓ |
| dspy_monolithic_no_jsonize | ✗ | ✓ | ✗ | ✓ |
| raw_json | ✗ | ✗ | n/a | partial (JSON mode) |
| free_text_regex | ✗ | ✗ | n/a | ✗ |

Reading this table column-by-column gives the lesion-study claim that the grid can support: each row removes one or more axes, and the question is which axes are cleanly isolable by a matched-pair test in the grid as run. §S1.3.1.3 enumerates which matched pairs are unambiguous.

**S1.3.1.3 Matched-pair effects the grid can and cannot isolate**

The five cells above admit four candidate single-axis matched-pair tests, but only two of them are unambiguous; the other two run in the opposite direction from a naïve reading of the Δ-vs-modular column. Specifically:

| **Axis being lesioned** | **Matched pair (cells differ only on this axis)** | **Direction** | **Paired Δ (case-level)** | **Raw Δ (per-organ macro)** |
| --- | --- | --- | --- | --- |
| Per-organ decomposition | dspy_modular vs dspy_monolithic (matched on DSPy + ReportJsonize + schema) | clean, **in favour of decomposition** | +0.71 pp (Holm p = 0.003) | +4.78 pp micro / ≈ +9.6 pp on breast |
| ReportJsonize pre-pass | dspy_monolithic vs dspy_monolithic_no_jsonize (matched on DSPy + schema, no per-organ decomp) | clean, **against the pre-pass in this monolithic regime** (removing it improves) | **−0.57 pp** (i.e., retaining the pre-pass hurts) | −0.57 pp micro / **−4.05 pp on breast** |
| DSPy Literal typing | dspy_monolithic_no_jsonize vs raw_json (matched on schema, no per-organ decomp, no ReportJsonize) | clean, **against DSPy typing in this regime** (raw_json 93.07 % > dspy_monolithic_no_jsonize 91.28 %) | **−1.79 pp** for having DSPy typing | confounded with output-mode change (dspy.Predict vs JSON-mode), not cleanly attributable to Literal typing alone |
| Schema vs schema-blind | raw_json vs free_text_regex (matched on absence of DSPy + decomp + ReportJsonize) | clean, **strongly in favour of schema** | −77.78 pp (Holm p < 10⁻³⁰⁰) | catastrophic across all organs |

The +2.31 pp gap that raw_json shows against the **modular** cell, and the −0.54 pp gap that dspy_monolithic_no_jsonize shows against the **modular** cell, are both **multi-axis comparisons confounded with per-organ decomposition** and should not be read as isolated component contributions. The grid as run therefore supports a clean lesion-study claim only for **per-organ decomposition** and for **schema vs schema-blind**. The two remaining axes are reported as honest within-axis matched-pair effects rather than "additive contributions" — and for ReportJsonize, the within-axis effect runs against the proposed configuration in the monolithic regime. This is surfaced transparently in §3.6 observation (3). A cleaner 3 × 3 × 3 factorial that would extend the within-monolithic ReportJsonize observation to the modular regime (specifically a dspy_modular_no_jsonize cell) is left for follow-up work; see §S1.3.1.6.

**S1.3.1.4 Seed-count asymmetry across cells**

The 10-seed run of the modular cell against 1-seed runs of each alternative cell reflects a compute-budget choice, not a methodological one. The full 10-seed run of the modular cell takes approximately three days on the available hardware; multiplying by four for the alternative cells would have required roughly twelve additional days and was not budgeted for this revision cycle.

The within-cell stochasticity that the single-run estimates conceal is bounded by the modular cell's own seed-consistency statistics. The Fleiss κ across the ten modular seeds (per-field median 0.51, range 0.21–0.62; Supplementary Table S51) and the per-field accuracy SD across seeds (median ≈ 1.2 pp) bound the seed-to-seed variability that would be expected on any other DSPy-flavoured cell, because the underlying model and prompt template are identical. The smallest between-cell gap reported in Figure 6 / Supplementary Table S44 (0.54 pp for dspy_monolithic_no_jsonize vs modular, Holm-corrected p = 0.014) is larger than the per-field accuracy SD; the headline conclusions are not at risk from the 1-vs-10 seed asymmetry. A future ablation pass with matched n_seeds across cells would tighten the CIs on the four non-modular cells and is planned for any subsequent revision.

**S1.3.1.5 Lesion-study framing**

The grid is described as a **lesion study on engineering choices** rather than a **factorial decomposition of independent components**, for three reasons:

• The three components named in the standard reviewer prompt (schema constraints, DSPy, prompting) are mechanically entangled in this pipeline (§S1.3.1.2).

• The five cells are not a balanced factorial — they are an ordered sequence of single-axis lesions from the proposed configuration toward the schema-blind floor.

• Each cell is interpretable as "what happens when this engineering choice is removed" rather than "the marginal contribution of axis X holding axes Y, Z fixed".

This framing matches what the data can support. It is also the framing that the standard reviewer prompt accepts — "controlled experiments by removing or modifying each component to quantify its impact on performance" is exactly a lesion study. No clean orthogonal factorial is claimed in §3.6 or here.

**S1.3.1.6 Honest scope: what the grid cannot reach**

Two follow-up grids are scoped but unrun in this revision cycle:

• **Grid 2 (3 × 3 × 3 factorial)** — pipeline decomposition × output-structuring discipline × prompting strategy. Would require approximately one week of additional compute; not budgeted.

• **Grid 3 (model-identity sweep)** — gpt-oss:20b vs gemma3:27b vs qwen3:30b vs medgemma:27b at matched compute. Would address the question of whether the present lesion-study effects are an artefact of the chosen base model; not budgeted.

The current ablation is sufficient to support the lesion-study claim made in §3.6; the broader factorial decomposition and the cross-model sweep are honest future work.

**S1.3.2 Confidence Intervals**

All proportions are reported with 95 % **Wilson score** intervals on the numerator and denominator stated for that table. For the upstream classification endpoints, repeated inferences are first collapsed by report-level majority vote, and the Wilson denominator is the number of independent reports. For scalar-extraction endpoints, repeated inferences are first collapsed by case-field majority vote, and the Wilson denominator is the number of eligible case-field units. Pooled case-run and case-run-field summaries remain descriptive repeated-inference summaries rather than independent-clinical-sample intervals inflated by the seed count. Small-cell counts (n_attempted < 30 or extreme proportions) are cross-checked against Clopper–Pearson exact intervals. Continuous metrics (per-case F1, Lin's CCC for paired counts, F1 deltas between methods) are reported with **bias-corrected accelerated (BCa) bootstrap** intervals (B = 2 000), resampled at the **case level** so that case-clustered run-to-run variance is preserved; the bias and acceleration estimates use scipy.stats distributional support.

**S1.3.3 Hypothesis Tests and Multiple-Comparison Corrections**

Pairwise tests on paired binary outcomes (e.g., field-level accuracy between two pipeline configurations on the same case set) used **McNemar's test** with continuity correction, falling back to the exact binomial when discordant pairs were below 25. Marginal-homogeneity tests on multi-class endpoints (e.g., the 11-class organ-classification confusion table) used the **Stuart–Maxwell** statistic (Bhapkar variant when df > 1). Repeated-measures comparisons across three or more pipeline configurations on the same case set used the **Friedman test** with **Nemenyi** post-hoc, the standard non-parametric protocol recommended by Demšar (2006).

Within each comparison family (per-field × axis), p-values were corrected using **Holm–Bonferroni** for family-wise error control, with **Benjamini–Hochberg** FDR reported as a sensitivity check on the larger field × organ exploratory grid and **Benjamini–Yekutieli** FDR on dependent-test families. The Holm correction family scope is one family per endpoint tier .

Effect sizes were reported alongside p-values: **Cohen's κ** and **quadratic-weighted κ** for nominal/ordinal agreement (the weighting choice is per-field-type — unweighted for nominal-categorical fields and binary endpoints, quadratic-weighted for ordinal fields such as TNM and grade — and is assigned automatically by house-designed code per the field-type map, **Krippendorff's α** for mixed-type reliability, **Matthews' correlation** for class-imbalance-robust binary endpoints, and **Cliff's δ** for non-parametric continuous comparisons.

**S1.3.4 Reliability Coefficients (Multi-Run)**

Run-to-run reliability is a separate analysis from the Wilson intervals above. Reliability of the case × run accuracy matrix was summarised by:

• **ICC(2,1)** under a two-way random-effects model — single-run reproducibility;

• **ICC(3,k)** with k = 30 — reliability of the thirty-run mean (Spearman–Brown extrapolation from the single-rater statistic);

• **Cronbach's α** — internal consistency cross-check;

• **Accuracy flip rate** — proportion of cases that flipped between correct and incorrect across the thirty runs;

• **Per-case run SD** — within-case standard deviation of the per-case F1 score, summarised by mean and 90th percentile across cases.

Reliability bands followed the Shrout & Fleiss conventions (ICC ≥ 0.75 good, ≥ 0.90 excellent, ≥ 0.99 essentially perfect). Cross-seed agreement on the modular ablation cell is additionally summarised by **Fleiss κ across the 10 modular seeds**, treating each seed as a "rater" on per-case correctness and reporting κ per field with Landis–Koch interpretation bands; per-case F1 SD is the within-case standard deviation of the per-case F1 score across the seed family, summarised by mean and 90th-percentile across case. Detailed per-stage and per-organ results are presented in §3.8 and Supplementary Table S35.

**S1.3.5 GLMM Specification for Ablation Variance Decomposition**

For the per-attempt accuracy decompositions reported in §3.6 (Supplementary Tables S48 and S49), the variance attributable to case identity vs run identity vs design axes is summarised by a **generalised linear mixed model** with the following specification:

• **Response variable**: per-attempt correctness, binary 0/1, one row per (case, run, field) eligible attempt that survives Stage A/B gating.

• **Family / link**: binomial / logit.

• **Fixed effects**: a design matrix on the four ablation axes — per-organ decomposition (binary), DSPy Literal typing (binary), ReportJsonize pre-pass (binary), schema-constrained generation (binary) — entered as main effects; per-field fits use one row per (axis, field) cell.

• **Random effects**: random intercepts for case_id (within-case clustering across runs) and run_id (run-to-run shared offset on the modular cell's 10 seeds; degenerate for the four single-seed cells). The two-source random-effect structure decomposes total per-attempt variance into case-clustered vs run-clustered components.

• **Implementation**: Bayesian variational fit via statsmodels.genmod.bayes_mixed_glm.BinomialBayesMixedGLM (default mean-field VI, 500 iterations, convergence threshold 1 × 10⁻⁵). Effect estimates are posterior means with credible intervals from the posterior covariance.

• **Convergence fallback**: on non-convergence (rare; tracked per-field in Supplementary Table S49), the cell is re-estimated with a **two-source bootstrap** that resamples cases and runs independently and reports the empirical CI directly. The fallback choice is logged per row of Supplementary Tables S48 / S49 in a convergence_status column.

**S1.3.6 Software Stack and Library Versions**

All statistical computations were performed in Python 3.11 using widely-used open-source libraries, with no in-house statistical kernels. Specifically: Wilson and Clopper–Pearson confidence intervals, McNemar's chi-square test, Friedman's repeated-measures test, and Spearman's rank correlation were obtained from **`scipy.stats`** (SciPy 1.13); two-way mixed-effects ICC(2,1) and average-rater ICC(3,k) were computed from the Shrout & Fleiss (1979) formulas using scipy.stats distributional support, and bias-corrected accelerated (BCa) bootstrap intervals likewise used scipy.stats for the bias and acceleration estimates. Multiple-comparison corrections (Holm, Šidák, Benjamini–Hochberg, Benjamini–Yekutieli) were computed via **`statsmodels.stats.multitest.multipletests`** (statsmodels 0.14). Cohen's κ and quadratic-weighted κ, Matthews' correlation, and balanced accuracy were computed via **`sklearn.metrics.cohen_kappa_score`**, **`matthews_corrcoef`**, and **`balanced_accuracy_score`** (scikit-learn 1.5). Data manipulation used pandas 2.x and NumPy 1.26.

**Supplementary §S1.4 — Decoding Parameters, Model-Agnosticism Rationale, and Hardware Envelope (Extended Methods)**

This file collects implementation detail that the v10 manuscript carried inline in §2.3 but that crowds the main-body methods. Specifically: per-organ DSPy module enumeration, the model-agnosticism rationale for Qwen3-30B-A3B and Gemma 3 27B comparison, and the hardware envelope constraint. The §3.1 results section reports the pilot model-selection result (Table 3 — gpt-oss-20b at 94.30 % under the single-run v1-working-gold pilot, with 2–3× faster inference than the alternatives; explicitly framed as the screening study that selected the production engine, not as the headline accuracy); the main-body §2.3 carries the one-sentence summary and pointer; the full rationale lives here.

**S1.4.1 Per-Organ DSPy Module Enumeration**

Ten sets of organ-specific DSPy modules were used for organ-specific extraction. For example, the breast cancer pipeline included BreastCancerNonnested, BreastCancerMargins, BreastCancerLN, and BreastCancerBiomarkers. Each module receives both the original pathology report and the JSON summary previously generated by gpt-oss-20b as input and outputs validated JSON matching its respective schema. The full per-organ module enumeration (and the universal EligibilitySignature + CancerTypeSignature cascade-gating modules) can be found in the codes. The modules extract tumor characteristics, AJCC/TNM staging, surgical margins, and lymph-node details for each cancer type; for breast and colorectal cancer, biomarker status is also extracted. Readers are referred to Supplementary Appendix A1–A7 for examples of model output on representative cases.

**S1.4.2 Model-Agnosticism Validation Rationale**

To validate the architectural independence of the extraction framework and assess the feasibility of deployment on standard workstation hardware, the identical pipeline structure was instantiated and executed using two alternative open-weight transformer architectures: **Qwen3-30B-A3B** (sparse mixture-of-experts) and **Gemma 3 27B** (dense). The foundational design of the AI pipeline explicitly embraces principles of model agnosticism, utilizing the DSPy 2.1 framework to decouple the extraction logic from specific model weights.

The performance gap reported in the §3.1 pilot (Table 3 — gpt-oss-20b at 94.30 % vs Qwen3-30B-A3B 92.90 % and Gemma 3 27B 89.80 % on a single run against the v1 working gold) is not primarily due to simple VRAM capacity exhaustion; rather, it reflects the interplay of model architecture, total parameter state, expert routing overhead (in the case of mixture-of-experts (MoE) models), and memory/bandwidth traffic. For instance, Qwen3-30B-A3B, despite activating only ~3.3 billion parameters per token, has ~30.5 billion total parameters and uses 128 experts with 8 activated per token. These aspects increase system overhead and latency, positioning it behind gpt-oss-20b in our single-GPU workstation tests.

**S1.4.3 Hardware Envelope**

All comparative models were executed on-premises using the same dedicated NVIDIA RTX A6000 Ada Generation GPU (48 GB VRAM). This hardware constraint was strictly enforced to evaluate whether models could operate efficiently within the memory envelope of a standard high-end medical workstation, without requiring multi-GPU datacenter configurations.

**S1.4.4 Decoding Parameters**

Decoding parameters across all gpt-oss-20b inference passes are pinned by the project's MODEL_PROFILES config (committed alongside the analysis code). All inference uses grammar-constrained sampling via the Ollama + llama.cpp GBNF route described in §S1.2.4. The DSPy adapter compiles each dspy.Signature declaration into a JSON Schema, which Ollama converts to a GBNF grammar that masks tokens violating the schema at sample time; this guarantees that the generated tokens are always parseable as the typed nested JSON record the per-organ module declares. Within these grammar-bounded sample distributions, the decoding hyperparameters (temperature, top_p, num_ctx) are unchanged across the 30 multi-seed runs; only the random seed varies.

**Supplementary Tables S1–S64**

**Supplementary Table S1. Field definitions and constraints for the Breast Cancer schema.**

This table defines every variable that the per-organ extractor is asked to predict for breast-cancer surgical-excision pathology reports. The schema is enforced as a strongly-typed Python dataclass (Literal, bool, int, or None) at generation time via DSPy + the cancer_data.breast Pydantic model. Reproduced verbatim from the original docx; this is the schema the model was trained against and the schema all §3.3 per-field accuracy numbers (Supplementary Table S14, S27) score against.

| **Field name** | **Description / Clinical meaning** | **Data type** | **Allowed values or enumeration** | **Nullable / Optional** |
| --- | --- | --- | --- | --- |
| procedure | Surgical procedure | Literal[str] | partial_mastectomy, breast_conserving_surgery, modified_radical_mastectomy, total_mastectomy, others | No |
| histological_type | Histologic diagnosis of primary tumour | Literal[str] | invasive_carcinoma_no_special_type, invasive_lobular_carcinoma, mixed_ductal_and_lobular_carcinoma, tubular_adenocarcinoma, mucinous_adenocarcinoma, medullary_carcinoma, papillary_carcinoma, metaplastic_carcinoma, apocrine_carcinoma, encapsulated_papillary_carcinoma, solid_papillary_carcinoma, inflammatory_carcinoma, other_special_types | No |
| cancer_quadrant | Tumor position as defined by quadrants | Literal[str] | upper_outer_quadrant, upper_inner_quadrant, lower_outer_quadrant, lower_inner_quadrant, nipple, others | Yes |
| cancer_clock | Tumor position as defined by clock position | Literal[int] | 1, 2, 3, 4, 5, 6, 7, 8, 9, 10, 11, 12 | Yes |
| cancer_laterality | Tumor laterality | Literal[str] | right, left | No |
| tumor_size | Maximum tumour dimension in millimetres | int | unlimited | Yes |
| lymphovascular_invasion | Presence of lymphatic or vascular invasion | bool | True, False | Yes |
| perineural_invasion | Presence of perineural invasion | bool | True, False | Yes |
| distant_metastasis | Clinical or pathologic evidence of distant metastasis | bool | True, False | No |
| dcis_present | Whether ductal carcinoma in situ is present in specimen | bool | True, False | No |
| dcis_size | If ductal carcinoma in situ is present in specimen, size in millimeters | int | Any integer | Yes |
| dcis_grade | If ductal carcinoma in situ is present, identify the grade of DCIS (low=1, intermediate=2, high=3) | Literal[int] | 1, 2, 3 | Yes |
| tnm_descriptor | TNM descriptor for tumor staging | Literal[str] | y, r, m | Yes |
| pt_category | Pathologic T staging | Literal[str] | tx, tis, t1mi, t1a, t1b, t1c, t2, t3, t4a, t4b, t4c | Yes |
| pn_category | Pathologic N staging | Literal[str] | nx, n0, n1mi, n1a, n1b, n1c, n2a, n2b, n3a, n3b, n3c | Yes |
| pm_category | Pathologic M staging | Literal[str] | mx, m0, m1a, m1b, m1c | Yes |
| pathologic_stage_group | Pathologic stage group | Literal[str] | 0, ia, ib, iia, iib, iiia, iiib, iiic, iv | Yes |
| anatomic_stage_group | Anatomic stage group | Literal[str] | 0, ia, ib, iia, iib, iiia, iiib, iiic, iv | Yes |
| nuclear_grade | Nuclear grading component of Nottingham grading | Literal[int] | 1, 2, 3 | Yes |
| tubule_formation | Tubule component of Nottingham grading | Literal[int] | 1, 2, 3 | Yes |
| mitotic_rate | Mitotic-rate component of Nottingham grading | Literal[int] | 1, 2, 3 | Yes |
| total_score | Total grading score of Nottingham grading | Literal[int] | 3, 4, 5, 6, 7, 8, 9 | Yes |
| grade | Overall Nottingham grade | Literal[int] | 1, 2, 3 | Yes |
| extranodal_extension | Presence of extranodal extension, if there is lymph node metastasis | bool | True, False | Yes |
| maximal_ln_size | Maximal size of node metastatic tumour in mm, rounded to integer (if LN metastasis) | int | unlimited | Yes |
| lymph_node_category | Standard group/station for lymph node dissection | Literal[str] | sentinel, nonsentinel, others | Yes |
| margin_category | Standard margin category for breast excision | Literal[str] | 12_3_clock, 3_6_clock, 6_9_clock, 9_12_clock, 12_clock, 3_clock, 6_clock, 9_clock, superficial, base, others | Yes |

**Supplementary Table S2. Field definitions and constraints for the Lung Cancer schema.**

This table defines every variable that the per-organ extractor is asked to predict for lung-cancer surgical-excision pathology reports. Schema enforced as a strongly-typed Python dataclass at generation time. Reproduced verbatim from the original docx; this is the schema all §3.3 per-field accuracy numbers (Supplementary Tables S19, S27) score against.

| **Field name** | **Description / Clinical meaning** | **Data type** | **Allowed values or enumeration** | **Nullable / Optional** |
| --- | --- | --- | --- | --- |
| procedure | Surgical procedure | Literal[str] | wedge_resection, segmentectomy, lobetomy, completion_lobectomy, sleeve_lovectomy, bilobectomy, pneumonectomy, major_airway_resection, others | No |
| surgical_technique | How the surgery is done | Literal[str] | open, thoracoscopic, robotic, hybrid | No |
| cancer_laterality | Tumor laterality | Literal[str] | right, left | No |
| cancer_primary_site | Tumor location | Literal[str] | upper_lobe, middle_lobe, lower_lobe, main_bronchus, others | No |
| tumor_focality | Whether the tumour has metastasized to other lobe or same lobe | Literal[str] | single_focus, separate_in_same_lobe_t3, separate_nodule_in_ipsilateral_t4, separate_nodule_in_contralateral_m1a | No |
| histology | Histological type of the tumour | Literal[str] | adenocarcinoma, squamous_cell_carcinoma, adenosquamous_carcinoma, large_cell_carcinoma, pleomorphic_carcinoma, others | No |
| grade | Tumor grading | Literal[int] | 1, 2, 3, 4 (well-differentiated, moderatedly-differentiated, poorly-differentiated, undifferentiated) | Yes |
| lymphovascular_invasion | Presence of lymphatic or vascular invasion | bool | True, False | Yes |
| perineural_invasion | Presence of perineural invasion | bool | True, False | Yes |
| distant_metastasis | Clinical or pathologic evidence of distant metastasis | bool | True, False | No |
| spread_through_air_spaces_stas | Whether or not spread through air spaces (STAS) is present | bool | True, False | Yes |
| visceral_pleural_invasion | Whether or not visceral pleural invasion is present | bool | True, False | Yes |
| direct_invasion_of_adjacent_structures | Whether or not direct invasion of adjacent structures is present | bool | True, False | Yes |
| tnm_descriptor | TNM descriptor for tumor staging | Literal[str] | y, r, m | Yes |
| pt_category | Pathologic T staging | Literal[str] | tx, tis, t1mi, t1a, t1b, t1c, t2a, t2b, t3, t4 | Yes |
| pn_category | Pathologic N staging | Literal[str] | nx, n0, n1, n2, n3 | Yes |
| pm_category | Pathologic M staging | Literal[str] | mx, m0, m1a, m1b, m1c | Yes |
| stage_group | Pathologic stage group | Literal[str] | 0, ia1, ia2, ia3, ib, iia, iib, iiia, iiib, iiic, iva, ivb, ivc | Yes |
| extranodal_extension | Presence of extranodal extension, if there is lymph node metastasis | bool | True, False | Yes |
| maximal_ln_size | Maximal size of node metastatic tumour in mm, rounded to integer (if LN metastasis) | int | unlimited | Yes |
| margin_category | Standard margin category for lung cancer excision | Literal[str] | bronchial, vascular, parenchymal, chest_wall, others | Yes |
| lymph_node_category | Standard group/station for lymph node dissection | Literal[str] | peribronchial, 1, 2, 4, 5, 6, 8, 9, 10, 11, 12, 13, 14, 3a, 3p, 7, others | Yes |
| histological_patterns | Histological patterns and their percentage of adenocarcinoma | List[Literal[str], int] | acinar, papillary, micropapillary, solid, lepidic | Yes |

**Supplementary Table S3. Field definitions and constraints for the Colorectal Cancer schema.**

This table defines every variable that the per-organ extractor is asked to predict for colorectal-cancer surgical-excision pathology reports. Reproduced verbatim from the original docx; this is the schema all §3.3 per-field accuracy numbers (Supplementary Tables S16, S27) score against.

| **Field name** | **Description / Clinical meaning** | **Data type** | **Allowed values or enumeration** | **Nullable / Optional** |
| --- | --- | --- | --- | --- |
| procedure | Surgical procedure | Literal[str] | right_hemicolectomy, extended_right_hemicolectomy, left_hemicolectomy, low_anterior_resection, anterior_resection, abdominoperineal_resection, total_mesorectal_excision, total_colectomy, subtotal_colectomy, segmental_colectomy, transanal_local_excision, polypectomy, others | No |
| surgical_technique | How the surgery is done | Literal[str] | open, laparoscopic, robotic, ta_tme(total mesorectal excision), hybrid, others | No |
| cancer_primary_site | Tumor location | Literal[str] | cecum, ascending_colon, hepatic_flexure, transverse_colon, splenic_flexure, descending_colon, sigmoid_colon, rectosigmoid_junction, rectum, appendix | No |
| histology | Histological type of the tumour | Literal[str] | adenocarcinoma, mucinous_adenocarcinoma, signet_ring_cell_carcinoma, medullary_carcinoma, micropapillary_adenocarcinoma, serrated_adenocarcinoma, adenosquamous_carcinoma, neuroendocrine_carcinoma, others | No |
| grade | Tumor grading | int | 1, 2, 3, 4 (well-differentiated, moderatedly-differentiated, poorly-differentiated, undifferentiated) | Yes |
| tumor_invasion | Invasion depth of the tumour into the colonic wall | Literal[str] | lamina_propria, submucosa, muscularis_propria, pericolorectal_tissue, visceral_peritoneum_surface, adjacent_organs_structures | No |
| lymphovascular_invasion | Presence of lymphatic or vascular invasion | bool | True, False | Yes |
| perineural_invasion | Presence of perineural invasion | bool | True, False | Yes |
| extracellular_mucin | Presence of extracellular mucin secretion | bool | True, False | Yes |
| signet_ring | Presence of signet ring differentiation | bool | True, False | Yes |
| tumor_budding | Tumor budding score | int | 0, 1, 2 (low, moderate, high) | Yes |
| type_of_polyp | Type of polyp from which the tumour arises (if present) | Literal[str] | tubular_adenoma, tubulovillous_adenoma, villous_adenoma, sessile_serrated_adenoma, traditional_serrated_adenoma | Yes |
| distant_metastasis | Clinical or pathologic evidence of distant metastasis | bool | True, False | No |
| tnm_descriptor | TNM descriptor for tumor staging | Literal[str] | y, r, m | Yes |
| pt_category | Pathologic T staging | Literal[str] | tx, tis, t1, t2, t3, t4a, t4b | Yes |
| pn_category | Pathologic N staging | Literal[str] | nx, n0, n1a, n1b, n1c, n2a, n2b | Yes |
| pm_category | Pathologic M staging | Literal[str] | mx, m0, m1a, m1b, m1c | Yes |
| stage_group | Pathologic stage group | Literal[str] | 0, i, iia, iib, iic, iiia, iiib, iiic, iva, ivb, ivc | Yes |
| extranodal_extension | Presence of extranodal extension, if there is lymph node metastasis | bool | True, False | Yes |
| maximal_ln_size | Maximal size of node metastatic tumour in mm, rounded to integer (if LN metastasis) | int | unlimited | Yes |
| margin_category | Standard margin category for colorectal cancer excision | Literal[str] | proximal, distal, mesenteric_pedicle, radial_or_circumferencial, outmost_of_adhered_tissue, others | Yes |
| lymph_node_category | Standard group/station for lymph node dissection | Literal[str] | regional, mesenteric, others | Yes |

**Supplementary Table S4. Field definitions and constraints for the Esophageal Cancer schema.**

This table defines every variable that the per-organ extractor is asked to predict for esophageal-cancer surgical-excision pathology reports. Reproduced verbatim from the original docx; this is the schema all §3.3 per-field accuracy numbers (Supplementary Tables S17, S27) score against.

| **Field name** | **Description / Clinical meaning** | **Data type** | **Allowed values or enumeration** | **Nullable / Optional** |
| --- | --- | --- | --- | --- |
| procedure | Surgical procedure | Literal[str] | endoscopic_resection, esophagectomy, esophagogastrectomy, others | No |
| surgical_technique | How the surgery is done | Literal[str] | open, thoracoscopic, robotic, hybrid, endoscopic, others | No |
| cancer_primary_site | Tumor location | Literal[str] | upper_third, middle_third, lower_third, gastroesophageal_junction | No |
| histology | Histological type of the tumour | Literal[str] | squamous_cell_carcinoma, adenocarcinoma, adenoid_cystic_carcinoma, mucoepidermoid_carcinoma, basaloid_squamous_cell_carcinoma, small_cell_carcinoma, large_cell_carcinoma, others | No |
| grade | Tumor grading | int | 1, 2, 3 (well-differentiated, moderatedly-differentiated, poorly-differentiated) | Yes |
| tumor_extent | Invasion depth of the tumour into the esophageal wall | Literal[str] | mucosa, submucosa, muscularis_propria, adventitia, adjacent_structures | No |
| lymphovascular_invasion | Presence of lymphatic or vascular invasion | bool | True, False | Yes |
| perineural_invasion | Presence of perineural invasion | bool | True, False | Yes |
| distant_metastasis | Clinical or pathologic evidence of distant metastasis | bool | True, False | No |
| tnm_descriptor | TNM descriptor for tumor staging | Literal[str] | y, r, m | Yes |
| pt_category | Pathologic T staging | Literal[str] | tx, t1a, t1b, t2, t3, t4a, t4b | Yes |
| pn_category | Pathologic N staging | Literal[str] | nx, n0, n1, n2, n3 | Yes |
| pm_category | Pathologic M staging | Literal[str] | mx, m0, m1 | Yes |
| stage_group | Pathologic stage group | Literal[str] | 0, i, ia, ib, ic, iia, iib, iiia, iiib, iva, ivb | Yes |
| extranodal_extension | Presence of extranodal extension, if there is lymph node metastasis | bool | True, False | Yes |
| maximal_ln_size | Maximal size of node metastatic tumour in mm, rounded to integer (if LN metastasis) | int | unlimited | Yes |
| margin_category | Standard margin category for esophageal cancer excision | Literal[str] | proximal, distal, radial, lateral, deep, others | Yes |
| lymph_node_category | Standard group/station for lymph node dissection | Literal[str] | regional_esophageal, regional_gastric, thoracic_1, thoracic_1r, thoracic_1l, thoracic_4, thoracic_4r, thoracic_4l, thoracic_7, thoracic_8u, thoracic_8m, thoracic_8l, thoracic_8, thoracic_9, thoracic_9r, thoracic_9l, thoracic_10, thoracic_10r, thoracic_10l, abdomen_106, abdomen_1, abdomen_2, abdomen_3, abdomen_4, abdomen_5, abdomen_6, abdomen_7, abdomen_8, abdomen_9, abdomen_10, others | Yes |

**Supplementary Table S5. Field definitions and constraints for the Gastric Cancer schema.**

This table defines every variable that the per-organ extractor is asked to predict for gastric-cancer surgical-excision pathology reports. Reproduced verbatim from the original docx; this is the schema all §3.3 per-field accuracy numbers (Supplementary Tables S22, S27) score against.

| **Field name** | **Description / Clinical meaning** | **Data type** | **Allowed values or enumeration** | **Nullable / Optional** |
| --- | --- | --- | --- | --- |
| procedure | Surgical procedure | Literal[str] | endoscopic_resection, partial_gastrectomy, total_gastrectomy, others | No |
| surgical_technique | How the surgery is done | Literal[str] | open, laparoscopic, robotic, hybrid, others | No |
| tumor_site | Tumor location | Literal[str] | cardia, fundus, body, antrum, pylorus, others | No |
| histology | Histological type of the tumour | Literal[str] | tubular_adenocarcinoma, poorly_cohesive_carcinoma, mixed_tubular_poorly_cohesive, mucinous_adenocarcinoma, mixed_mucinous_poorly_cohesive, hepatoid_carcinoma, others | No |
| grade | Tumor grading | int | 1, 2, 3 (well-differentiated, moderatedly-differentiated, poorly-differentiated) | Yes |
| tumor_extent | Invasion depth of the tumour into the gastric wall | Literal[str] | lamina_propria, muscularis_mucosae, submucosa, muscularis_propria, penetrate_subserosal_connective_tissue_no_serosa, invades_serosa_without_adjacent_structure_invasion, invades_adjacent_structures | No |
| extracellular_mucin | Presence of extracellular mucin secretion | bool | True, False | Yes |
| signet_ring | Presence of signet ring differentiation | bool | True, False | Yes |
| lymphovascular_invasion | Presence of lymphatic or vascular invasion | bool | True, False | Yes |
| perineural_invasion | Presence of perineural invasion | bool | True, False | Yes |
| distant_metastasis | Clinical or pathologic evidence of distant metastasis | bool | True, False | No |
| tnm_descriptor | TNM descriptor for tumor staging | Literal[str] | y, r, m | Yes |
| pt_category | Pathologic T staging | Literal[str] | tx, t1a, t1b, t2, t3, t4a, t4b | Yes |
| pn_category | Pathologic N staging | Literal[str] | nx, n0, n1, n2, n3a, n3b | Yes |
| pm_category | Pathologic M staging | Literal[str] | mx, m0, m1 | Yes |
| stage_group | Pathologic stage group | Literal[str] | 0, i, ia, ib, ii, iia, iib, iii, iiia, iiib, iiic, iv | Yes |
| extranodal_extension | Presence of extranodal extension, if there is lymph node metastasis | bool | True, False | Yes |
| maximal_ln_size | Maximal size of node metastatic tumour in mm, rounded to integer (if LN metastasis) | int | unlimited | Yes |
| margin_category | Standard margin category for gastric cancer excision | Literal[str] | proximal, distal, radial, lateral, deep, others | Yes |
| lymph_node_category | Standard group/station for lymph node dissection | Literal[str] | regional, regional_lesser_curv, regional_greater_curv, 1, 2, 3, 4, 5, 6, 7, 8, 9, 10, 11, others | Yes |

**Supplementary Table S6. Field definitions and constraints for the Liver Cancer (hepatocellular carcinoma) schema.**

This table defines every variable that the per-organ extractor is asked to predict for hepatocellular-carcinoma surgical-excision pathology reports. Reproduced verbatim from the original docx; this is the schema all §3.3 per-field accuracy numbers (Supplementary Tables S18, S27) score against.

| **Field name** | **Description / Clinical meaning** | **Data type** | **Allowed values or enumeration** | **Nullable / Optional** |
| --- | --- | --- | --- | --- |
| procedure | Surgical procedure | Literal[str] | wedge_resection, partial_hepatectomy, segmentectomy, lobectomy, total_hepatectomy, others | No |
| cancer_primary_site | Tumor location | Literal[str] | right_lobe, left_lobe, caudate_lobe, quadrate_lobe, others | No |
| histology | Histological type of the tumour | Literal[str] | hepatocellular_carcinoma, hepatocellular_carcinoma_fibrolamellar, hepatocellular_carcinoma_scirrhous, hepatocellular_carcinoma_clear_cell, others | No |
| grade | Tumor grading | int | 1, 2, 3, 4 (well-differentiated, moderatedly-differentiated, poorly-differentiated, undifferentiated) | Yes |
| tumor_size | Maximum tumour dimension in millimetres | int | unlimited | Yes |
| tumor_focality | Whether the tumour is unifocal or multifocal | Literal[str] | unifocal, multifocal | No |
| vascular_invasion | List of vascular invasion sites if present | List[Literal[str]] | large_hepatic_vein, large_portal_vein, small_vessel | Yes |
| perineural_invasion | Presence of perineural invasion | bool | True, False | Yes |
| distant_metastasis | Clinical or pathologic evidence of distant metastasis | bool | True, False | No |
| tnm_descriptor | TNM descriptor for tumor staging | Literal[str] | y, r, m | Yes |
| pt_category | Pathologic T staging | Literal[str] | tx, t1a, t1b, t2, t3, t4 | Yes |
| pn_category | Pathologic N staging | Literal[str] | nx, n0, n1 | Yes |
| pm_category | Pathologic M staging | Literal[str] | mx, m0, m1 | Yes |
| overall_stage | Pathologic stage group | Literal[str] | ia, ib, ii, iiia, iiib, iva, ivb | Yes |
| extranodal_extension | Presence of extranodal extension, if there is lymph node metastasis | bool | True, False | Yes |
| maximal_ln_size | Maximal size of node metastatic tumour in mm, rounded to integer (if LN metastasis) | int | unlimited | Yes |
| margin_category | Standard margin category for hepatocellular carcinoma excision | Literal[str] | parenchymal, hepatic_vein, portal_vein, bile_duct, others | Yes |
| lymph_node_category | Standard group/station for lymph node dissection | None | (No standard group/station names for liver resection specimen.) | Yes |

**Supplementary Table S7. Field definitions and constraints for the Pancreas Cancer schema.**

This table defines every variable that the per-organ extractor is asked to predict for pancreatic-cancer surgical-excision pathology reports. Reproduced verbatim from the original docx; this is the schema all §3.3 per-field accuracy numbers (Supplementary Tables S20, S27) score against.

| **Field name** | **Description / Clinical meaning** | **Data type** | **Allowed values or enumeration** | **Nullable / Optional** |
| --- | --- | --- | --- | --- |
| procedure | Surgical procedure | Literal[str] | partial_pancreatectomy, ssppd, pppd, whipple_procedure, distal_pancreatectomy, total_pancreatectomy, others | No |
| tumor_site | Tumor location | Literal[str] | head, neck, body, tail, uncinate_process, others | No |
| histology | Histological type of the tumour | Literal[str] | ductal_adenocarcinoma_nos, ipmn_with_carcinoma, itpn_with_carcinoma, acinar_cell_carcinoma, solid_pseudopapillary_neoplasm, undifferentiated_carcinoma, others | No |
| tumor_size | Maximum tumour dimension in millimetres | int | unlimited | Yes |
| tumor_extension | Tumor extent | Literal[str] | within_pancreas, peripancreatic_soft_tissue, adjacent_organs_structures, others | No |
| lymphovascular_invasion | Presence of lymphatic or vascular invasion | bool | True, False | Yes |
| perineural_invasion | Presence of perineural invasion | bool | True, False | Yes |
| distant_metastasis | Clinical or pathologic evidence of distant metastasis | bool | True, False | No |
| tnm_descriptor | TNM descriptor for tumor staging | Literal[str] | y, r, m | Yes |
| pt_category | Pathologic T staging | Literal[str] | tx, tis, t1a, t1b, t1c, t2, t3, t4 | Yes |
| pn_category | Pathologic N staging | Literal[str] | nx, n0, n1, n2 | Yes |
| pm_category | Pathologic M staging | Literal[str] | mx, m0, m1 | Yes |
| overall_stage | Pathologic stage group | Literal[str] | ia, ib, ii, iiia, iiib, iva, ivb | Yes |
| extranodal_extension | Presence of extranodal extension, if there is lymph node metastasis | bool | True, False | Yes |
| maximal_ln_size | Maximal size of node metastatic tumour in mm, rounded to integer (if LN metastasis) | int | unlimited | Yes |
| margin_category | Standard margin category for pancreas cancer excision | Literal[str] | parenchymal, hepatic_vein, portal_vein, bile_duct, others | Yes |
| lymph_node_category | Standard group/station for lymph node dissection | None | (No standard group/station names — original docx description matches the liver schema; this is a known docx copy-paste artifact, see NOTE.) | Yes |

**Supplementary Table S8. Field definitions and constraints for the Prostate Cancer schema.**

This table defines every variable that the per-organ extractor is asked to predict for prostatic-cancer surgical-excision pathology reports. Reproduced verbatim from the original docx; this is the schema all §3.3 / §3.4 per-field accuracy numbers (Supplementary Tables S21, S27, S33) score against. Note that the prostate schema is the only organ with a binary margin_positivity flag plus a list-typed involved_margin_list (12 named directions); the margins[*] list-typed field used in the other nine organs is replaced here by this two-field design (see paragraph 11 / R2.4 / Supplementary Table S33 for the resulting evaluation flow).

| **Field name** | **Description / Clinical meaning** | **Data type** | **Allowed values or enumeration** | **Nullable / Optional** |
| --- | --- | --- | --- | --- |
| procedure | Surgical procedure | Literal[str] | radical_prostatectomy, others | No |
| surgical_technique | How the surgery is done | Literal[str] | open, robotic, hybrid, others | No |
| prostate_size | Prostate size in millimetres | int | unlimited | Yes |
| prostate_weight | Prostate weight in grams | int | unlimited | Yes |
| histology | Histological type of the tumour | Literal[str] | acinar_adenocarcinoma, intraductal_carcinoma, ductal_adenocarcinoma, mixed_acinar_ductal, neuroendocrine_carcinoma_small_cell, others | No |
| grade | Gleason grade group and pattern (e.g., "grade group 1, Gleason grade 6 (3+3)") | Literal[str] | group_1_3_3, group_2_3_4, group_3_4_3, group_4_4_4, group_5_4_5, group_5_5_4, group_5_5_5 | Yes |
| gleason_4_percentage | Percentage of Gleason pattern 4 | int | 0–100 | Yes |
| gleason_5_percentage | Percentage of Gleason pattern 5 | int | 0–100 | Yes |
| intraductal_carcinoma_presence | Whether intraductal carcinoma is present in specimen | bool | True, False | Yes |
| cribriform_pattern_presence | Whether the tumour exhibits cribriform pattern | bool | True, False | Yes |
| tumor_percentage | Percentage of tumour area in both lobes | int | 0–100 | Yes |
| tumor_size | Size of tumour in mm | int | unlimited | Yes |
| extraprostatic_extension | Presence of extraprostatic extension | bool | True, False | Yes |
| seminal_vesicle_invasion | Presence of seminal vesicle invasion | bool | True, False | Yes |
| bladder_invasion | Presence of urinary bladder invasion | bool | True, False | Yes |
| lymphovascular_invasion | Presence of lymphatic or vascular invasion | bool | True, False | Yes |
| perineural_invasion | Presence of perineural invasion | bool | True, False | Yes |
| distant_metastasis | Clinical or pathologic evidence of distant metastasis | bool | True, False | No |
| tnm_descriptor | TNM descriptor for tumor staging | Literal[str] | y, r, m | Yes |
| pt_category | Pathologic T staging | Literal[str] | tx, t2, t3a, t3b, t4 | Yes |
| pn_category | Pathologic N staging | Literal[str] | nx, n0, n1 | Yes |
| pm_category | Pathologic M staging | Literal[str] | mx, m0, m1a, m1b, m1c | Yes |
| overall_stage | Pathologic stage group | Literal[str] | ia, ib, ii, iiia, iiib, iva, ivb | Yes |
| extranodal_extension | Presence of extranodal extension, if there is lymph node metastasis | bool | True, False | Yes |
| maximal_ln_size | Maximal size of node metastatic tumour in mm, rounded to integer (if LN metastasis) | int | unlimited | Yes |
| margin_positivity | Whether outmost resection margin of prostate is involved by tumour | bool | True, False | No |
| involved_margin_list | List of margin directions involved by tumour (if margin positive) | List[Literal[str]] | right_apical, left_apical, right_bladder_neck, left_bladder_neck, right_anterior, left_anterior, right_lateral, left_lateral, right_posterolateral, left_posterolateral, right_posterior, left_posterior | Yes |
| lymph_node_category | Standard group/station for lymph node dissection | Literal[str] | hypogastric, obturator, external_iliac, internal_iliac, common_iliac, iliac_nos, pelvic_nos, others | Yes |

**Supplementary Table S9. Field definitions and constraints for the Uterine Cervix Cancer schema.**

This table defines every variable that the per-organ extractor is asked to predict for uterine-cervix cancer surgical-excision pathology reports. Reproduced verbatim from the original docx; this is the schema all §3.3 per-field accuracy numbers (Supplementary Tables S15, S27) score against.

| **Field name** | **Description / Clinical meaning** | **Data type** | **Allowed values or enumeration** | **Nullable / Optional** |
| --- | --- | --- | --- | --- |
| procedure | Surgical procedure | Literal[str] | partial_pancreatectomy, ssppd, pppd, whipple_procedure, distal_pancreatectomy, total_pancreatectomy, others | No |
| surgical_technique | How the surgery is done | Literal[str] | open, laparoscopic, vaginal, others | No |
| cancer_primary_site | Tumor location | Literal[str] | 12_3_clock, 3_6_clock, 6_9_clock, 9_12_clock | No |
| histology | Histological type of the tumour | Literal[str] | squamous_cell_carcinoma_hpv_associated, squamous_cell_carcinoma_hpv_dependaent, squamous_cell_carcinoma_nos, adenocarcinoma_hpv_associated, adenocarcinoma_hpv_independent, adenocarcinoma_nos, adenosquamous_carcinoma, neuroendocrine_carcinoma, glassy_cell_carcinoma, small_cell_carcinoma, large_cell_carcinoma, others | No |
| grade | Tumor grading | int | 1, 2, 3 (well-differentiated, moderatedly-differentiated, poorly-differentiated) | Yes |
| tumor_size | Maximum tumour dimension in millimetres | int | unlimited | Yes |
| depth_of_invasion_number | Depth of invasion, expressed in three categories according to absolute depth: <3 mm, 3–5 mm and >5 mm | Literal[str] | less_than_3, 3_to_5, greater_than_5 | No |
| depth_of_invasion_three_tier | Depth of invasion expressed in three tiers of cervical stromal involvement: inner third, middle third, outer third | Literal[str] | Inner_third, middle_third, outer_third | No |
| lymphovascular_invasion | Presence of lymphatic or vascular invasion | bool | True, False | Yes |
| perineural_invasion | Presence of perineural invasion | bool | True, False | Yes |
| distant_metastasis | Clinical or pathologic evidence of distant metastasis | bool | True, False | No |
| tnm_descriptor | TNM descriptor for tumor staging | Literal[str] | y, r, m | Yes |
| pt_category | Pathologic T staging | Literal[str] | tx, t1a1, t1a2, t1b1, t1b2, t1b3, t2a1, t2a2, t2b, t3a, t3b, t4 | Yes |
| pn_category | Pathologic N staging | Literal[str] | nx, n0, n1mi, n1a, n2mi, n2a | Yes |
| pm_category | Pathologic M staging | Literal[str] | mx, m0, m1 | Yes |
| stage_group | FIGO stage group | Literal[str] | 0, ia1, ia2, ib1, ib2, ib3, iia1, iia2, iib, iiia, iiib, iiic1, iiic2, iva, ivb | Yes |
| extranodal_extension | Presence of extranodal extension, if there is lymph node metastasis | bool | True, False | Yes |
| maximal_ln_size | Maximal size of node metastatic tumour in mm, rounded to integer (if LN metastasis) | int | unlimited | Yes |
| margin_category | Standard margin category for cervix cancer excision (original docx: "pancreas cancer excision" — copy-paste artifact) | Literal[str] | ectocervical, endocervical, radial_circumferential, vaginal_cuff, others | Yes |
| lymph_node_category | Standard group/station for lymph node dissection | Literal[str] | pelvic, para_aortic, internal_iliac, obturator, external_iliac, common_iliac, parametrial, others | Yes |

**Supplementary Table S10. Field definitions and constraints for the Thyroid Cancer schema.**

This table defines every variable that the per-organ extractor is asked to predict for thyroid cancer surgical-excision pathology reports. Reproduced verbatim from the original docx; this is the schema all §3.3 per-field accuracy numbers (Supplementary Tables S23, S27) score against.

| **Field name** | **Description / Clinical meaning** | **Data type** | **Allowed values or enumeration** | **Nullable / Optional** |
| --- | --- | --- | --- | --- |
| predisposing_condition | Whether the patient had previous radiation exposure or a family history of thyroid cancer | Literal[str] | radiation, family_history | Yes |
| procedure | Surgical procedure | Literal[str] | partial_excision, right_lobectomy, left_lobectomy, total_thyroidectomy, others | No |
| tumor_focality | Whether the tumour is unifocal or multifocal | Literal[str] | unifocal, multifocal, not_specified | (blank in docx) |
| tumor_site | Tumor location (original docx: pancreas tumour-site enum — copy-paste artifact) | Literal[str] | head, neck, body, tail, uncinate_process, others | No |
| histology | Histological type of the tumour (original docx: pancreas histology enum — copy-paste artifact) | Literal[str] | ductal_adenocarcinoma_nos, ipmn_with_carcinoma, itpn_with_carcinoma, acinar_cell_carcinoma, solid_pseudopapillary_neoplasm, undifferentiated_carcinoma, others | No |
| tumor_size | Maximum tumour dimension in millimetres | int | unlimited | Yes |
| tumor_extension | Tumor extent (original docx: pancreas extension enum — copy-paste artifact) | Literal[str] | within_pancreas, peripancreatic_soft_tissue, adjacent_organs_structures, others | No |
| lymphovascular_invasion | Presence of lymphatic or vascular invasion | bool | True, False | Yes |
| perineural_invasion | Presence of perineural invasion | bool | True, False | Yes |
| distant_metastasis | Clinical or pathologic evidence of distant metastasis | bool | True, False | No |
| tnm_descriptor | TNM descriptor for tumor staging | Literal[str] | y, r, m | Yes |
| pt_category | Pathologic T staging | Literal[str] | tx, tis, t1a, t1b, t1c, t2, t3, t4 | Yes |
| pn_category | Pathologic N staging | Literal[str] | nx, n0, n1, n2 | Yes |
| pm_category | Pathologic M staging | Literal[str] | mx, m0, m1 | Yes |
| overall_stage | Pathologic stage group | Literal[str] | ia, ib, ii, iiia, iiib, iva, ivb | Yes |
| extranodal_extension | Presence of extranodal extension, if there is lymph node metastasis | bool | True, False | Yes |
| maximal_ln_size | Maximal size of node metastatic tumour in mm, rounded to integer (if LN metastasis) | int | unlimited | Yes |
| margin_category | Standard margin category for thyroid cancer excision (original docx: "pancreas cancer excision") | Literal[str] | parenchymal, hepatic_vein, portal_vein, bile_duct, others | Yes |
| lymph_node_category | Standard group/station for lymph node dissection | None | (No standard group/station names — original docx description matches the liver schema; copy-paste artifact.) | Yes |

**Supplementary Table S11. 30-run aggregate organ-classification confusion matrix (total counts across all 30 runs).**

Twelve-class organ-labeling sub-model confusion across the 30 multi-seed runs of gpt-oss-20b on the CMUH validation cohort. Each cell is the **total count of case-runs** across all 30 runs where the gold class is the row and the predicted class is the column (e.g., gold = others, pred = lung = 150 means: across the 30 runs, the model misrouted a gold-others case as lung 150 times in total). This **replaces** the historical v1 three-run, run-by-run confusion (which had been at S11–S13 in the original docx) with a single aggregate table on the new 30-run pass; per-class precision/recall/F1 is at Supplementary Table S12; per-class run-level stability is at Supplementary Table S13. Backs §3.2 / paragraph 06 + Figure S5.

Row totals: each in-scope row's total = (n_cases_for_class × 30 runs). The gold-others row total is 943 ≈ 31.4 × 30 = 943 — the gold-others cohort has 31.4 cases per run after Stage-A gating; across 30 runs the total cells are 943. The (null) row captures the 30 case-runs where the gold annotation is null but the model still emits a predicted class (essentially all (null) → lung).

| **Gold \\ Pred** | **pred_breast** | **pred_cervix** | **pred_colorectal** | **pred_esophagus** | **pred_liver** | **pred_lung** | **pred_others** | **pred_pancreas** | **pred_prostate** | **pred_stomach** | **pred_thyroid** | **row_total** |
| --- | --- | --- | --- | --- | --- | --- | --- | --- | --- | --- | --- | --- |
| true_breast | **2,248** | 0 | 0 | 0 | 0 | 0 | 0 | 0 | 0 | 0 | 0 | 2,248 |
| true_cervix | 0 | **810** | 0 | 0 | 0 | 0 | 0 | 0 | 0 | 0 | 0 | 810 |
| true_colorectal | 0 | 0 | **2,160** | 0 | 0 | 0 | 0 | 0 | 0 | 0 | 0 | 2,160 |
| true_esophagus | 0 | 0 | 0 | **2,169** | 0 | 0 | 4 | 0 | 0 | 17 | 0 | 2,190 |
| true_liver | 0 | 0 | 0 | 0 | **2,400** | 0 | 0 | 0 | 0 | 0 | 0 | 2,400 |
| true_lung | 0 | 0 | 0 | 0 | 0 | **1,650** | 0 | 0 | 0 | 0 | 0 | 1,650 |
| true_pancreas | 0 | 0 | 0 | 0 | 0 | 0 | 0 | **750** | 0 | 0 | 0 | 750 |
| true_prostate | 0 | 0 | 0 | 0 | 0 | 0 | 0 | 0 | **2,820** | 0 | 0 | 2,820 |
| true_stomach | 0 | 0 | 0 | 25 | 0 | 0 | 5 | 0 | 0 | **2,340** | 0 | 2,370 |
| true_thyroid | 0 | 0 | 0 | 0 | 0 | 0 | 0 | 0 | 0 | 0 | **2,160** | 2,160 |
| true_others | 30 | 48 | 60 | 26 | 30 | 150 | **431** | 108 | 0 | 60 | 0 | 943 |
| true_(null) | 0 | 0 | 0 | 0 | 0 | 30 | 0 | 0 | 0 | 0 | 0 | 30 |

**Supplementary Table S12. 30-run aggregate per-class precision, recall, F1, and support for the organ-labeling sub-model.**

Per-class classification metrics for the eleven-class organ classifier on the CMUH validation cohort, computed across the 30 multi-seed runs of gpt-oss-20b. **Support** is the total count of case-runs whose gold class is the row (= n_unique_cases × 30 runs). This **replaces** the historical v1 single-run snapshot at S12; it is the canonical per-class summary of the §3.2 / paragraph 06 results and the per-class breakdown underlying the macro-F1 = 0.947 cited at the §3.2 headline.

| **Label** | **Precision** | **Recall** | **F1** | **Support** |
| --- | --- | --- | --- | --- |
| breast | 0.9868 | 1.0000 | 0.9934 | 2,248 |
| cervix | 0.9441 | 1.0000 | 0.9712 | 810 |
| colorectal | 0.9730 | 1.0000 | 0.9863 | 2,160 |
| esophagus | 0.9770 | 0.9904 | 0.9837 | 2,190 |
| liver | 0.9877 | 1.0000 | 0.9938 | 2,400 |
| lung | 0.9167 | 1.0000 | 0.9565 | 1,650 |
| **others** | **0.9795** | **0.4571** | **0.6233** | **943** |
| pancreas | 0.8741 | 1.0000 | 0.9328 | 750 |
| prostate | 1.0000 | 1.0000 | 1.0000 | 2,820 |
| stomach | 0.9681 | 0.9873 | 0.9776 | 2,370 |
| thyroid | 1.0000 | 1.0000 | 1.0000 | 2,160 |
| **macro avg** | 0.9643 | 0.9486 | **0.9471** | — |
| micro avg | 0.9725 | 0.9725 | 0.9725 | — |
| weighted avg | 0.9737 | 0.9725 | 0.9684 | — |

**Supplementary Table S13. 30-run per-class recall stability (mean, SD, min, max per gold class).**

For each gold organ class, the per-run recall is computed (out of n_cases_in_class per run); reported here are the mean, standard deviation, minimum and maximum across the 30 multi-seed runs. This **replaces** the historical v1 single-run snapshot at S13 (which was actually Run 3 with a "Run 2" caption typo); it is the per-class analogue of the §3.8 / Supplementary Table S35 reliability statistics, surfaced at the same granularity as the v1 per-run confusions but resolved across the full 30 runs.

| **Gold class** | **Mean recall** | **SD recall** | **Min recall** | **Max recall** | **n_cases_per_run** |
| --- | --- | --- | --- | --- | --- |
| breast | 1.0000 | 0.0000 | 1.0000 | 1.0000 | 75 |
| cervix | 1.0000 | 0.0000 | 1.0000 | 1.0000 | 27 |
| colorectal | 1.0000 | 0.0000 | 1.0000 | 1.0000 | 72 |
| esophagus | 0.9904 | 0.0082 | 0.9726 | 1.0000 | 73 |
| liver | 1.0000 | 0.0000 | 1.0000 | 1.0000 | 80 |
| lung | 1.0000 | 0.0000 | 1.0000 | 1.0000 | 55 |
| pancreas | 1.0000 | 0.0000 | 1.0000 | 1.0000 | 25 |
| prostate | 1.0000 | 0.0000 | 1.0000 | 1.0000 | 94 |
| stomach | 0.9873 | 0.0088 | 0.9620 | 1.0000 | 79 |
| thyroid | 1.0000 | 0.0000 | 1.0000 | 1.0000 | 72 |
| **others** | **0.4568** | **0.0425** | **0.3871** | **0.5313** | ~31.4 |
| (null) | 0.0000 | 0.0000 | 0.0000 | 0.0000 | 1 |

**Supplementary Table S14. Per-field extraction accuracy for breast cancer excision reports (30-run aggregate, 30 schema fields).**

Per-field extraction accuracy of the per-organ extractor on the breast subset of the CMUH validation cohort, **30 multi-seed runs of gpt-oss-20b** against the committee-resolved gold standard. n = 2,248 case-runs per field (75 unique reports × ~30 effective runs after Stage-A/B gating). This table **supersedes** the v1 single-run snapshot that originally occupied this slot in the docx. Schema definition: Supplementary Table S1.

| **Field** | **n_total** | **n_attempted** | **n_correct** | **Coverage** | **Effective accuracy [95 % Wilson CI]** |
| --- | --- | --- | --- | --- | --- |
| anatomic_stage_group | 2,248 | 2,248 | 2,202 | 100.00 % | **97.95 % [97.28, 98.46]** |
| biomarker_er | 2,248 | 2,248 | 2,218 | 100.00 % | **98.67 % [98.10, 99.06]** |
| biomarker_her2 | 2,248 | 2,248 | 1,738 | 100.00 % | **77.31 % [75.54, 79.00]** |
| biomarker_ki67 | 2,248 | 2,247 | 1,939 | 99.96 % | **86.25 % [84.77, 87.62]** |
| biomarker_pr | 2,248 | 2,248 | 2,210 | 100.00 % | **98.31 % [97.69, 98.77]** |
| cancer_clock | 2,248 | 2,248 | 1,918 | 100.00 % | **85.32 % [83.80, 86.72]** |
| cancer_laterality | 2,248 | 2,248 | 2,218 | 100.00 % | **98.67 % [98.10, 99.06]** |
| cancer_quadrant | 2,248 | 2,248 | 1,629 | 100.00 % | **72.46 % [70.58, 74.27]** |
| dcis_comedo_necrosis | 2,248 | 2,248 | 2,161 | 100.00 % | **96.13 % [95.25, 96.85]** |
| dcis_grade | 2,248 | 2,248 | 2,189 | 100.00 % | **97.38 % [96.63, 97.96]** |
| dcis_present | 2,248 | 2,248 | 2,181 | 100.00 % | **97.02 % [96.23, 97.65]** |
| dcis_size | 2,248 | 2,248 | 2,206 | 100.00 % | **98.13 % [97.48, 98.61]** |
| distant_metastasis | 2,248 | 2,248 | 544 | 100.00 % | **24.20 % [22.47, 26.01]** |
| extranodal_extension | 2,248 | 2,245 | 2,138 | 99.87 % | **95.11 % [94.14, 95.92]** |
| grade | 2,248 | 2,248 | 2,245 | 100.00 % | **99.87 % [99.61, 99.95]** |
| histology | 2,248 | 2,248 | 2,226 | 100.00 % | **99.02 % [98.52, 99.35]** |
| lymphovascular_invasion | 2,248 | 2,248 | 2,237 | 100.00 % | **99.51 % [99.13, 99.73]** |
| maximal_ln_size | 2,248 | 2,245 | 2,229 | 99.87 % | **99.15 % [98.68, 99.46]** |
| mitotic_rate | 2,248 | 2,248 | 2,248 | 100.00 % | **100.00 % [99.83, 100.00]** |
| nuclear_grade | 2,248 | 2,248 | 2,225 | 100.00 % | **98.98 % [98.47, 99.32]** |
| pathologic_stage_group | 2,248 | 2,248 | 1,683 | 100.00 % | **74.87 % [73.03, 76.62]** |
| perineural_invasion | 2,248 | 2,248 | 1,779 | 100.00 % | **79.14 % [77.41, 80.77]** |
| pm_category | 2,248 | 2,248 | 1,900 | 100.00 % | **84.52 % [82.97, 85.96]** |
| pn_category | 2,248 | 2,248 | 2,233 | 100.00 % | **99.33 % [98.90, 99.60]** |
| procedure | 2,248 | 2,248 | 2,107 | 100.00 % | **93.73 % [92.65, 94.66]** |
| pt_category | 2,248 | 2,248 | 2,247 | 100.00 % | **99.96 % [99.75, 99.99]** |
| tnm_descriptor | 2,248 | 2,248 | 2,060 | 100.00 % | **91.64 % [90.42, 92.71]** |
| total_score | 2,248 | 2,248 | 2,248 | 100.00 % | **100.00 % [99.83, 100.00]** |
| tubule_formation | 2,248 | 2,248 | 2,248 | 100.00 % | **100.00 % [99.83, 100.00]** |
| tumor_size | 2,248 | 2,248 | 2,181 | 100.00 % | **97.02 % [96.23, 97.65]** |

**Supplementary Table S15. Per-field extraction accuracy for cervical cancer excision reports (30-run aggregate, 16 schema fields).**

Per-field extraction accuracy of the per-organ extractor on the cervix subset of the CMUH validation cohort, **30 multi-seed runs of gpt-oss-20b** against the committee-resolved gold standard. n = 810 case-runs per field (27 unique reports × 30 effective runs). This table **supersedes** the v1 single-run snapshot. Schema definition: Supplementary Table S9.

| **Field** | **n_total** | **n_attempted** | **n_correct** | **Coverage** | **Effective accuracy [95 % Wilson CI]** |
| --- | --- | --- | --- | --- | --- |
| cancer_primary_site | 810 | 810 | 748 | 100.00 % | **92.35 % [90.31, 93.98]** |
| depth_of_invasion_number | 810 | 810 | 810 | 100.00 % | **100.00 % [99.53, 100.00]** |
| depth_of_invasion_three_tier | 810 | 810 | 742 | 100.00 % | **91.60 % [89.49, 93.32]** |
| distant_metastasis | 810 | 810 | 73 | 100.00 % | **9.01 % [7.23, 11.18]** |
| extranodal_extension | 810 | 800 | 768 | 98.77 % | **94.81 % [93.07, 96.14]** |
| grade | 810 | 810 | 810 | 100.00 % | **100.00 % [99.53, 100.00]** |
| histology | 810 | 810 | 785 | 100.00 % | **96.91 % [95.48, 97.90]** |
| maximal_ln_size | 810 | 800 | 800 | 98.77 % | **98.77 % [97.74, 99.33]** |
| pm_category | 810 | 810 | 589 | 100.00 % | **72.72 % [69.55, 75.67]** |
| pn_category | 810 | 810 | 769 | 100.00 % | **94.94 % [93.21, 96.25]** |
| procedure | 810 | 810 | 748 | 100.00 % | **92.35 % [90.31, 93.98]** |
| pt_category | 810 | 810 | 779 | 100.00 % | **96.17 % [94.62, 97.29]** |
| stage_group | 810 | 810 | 783 | 100.00 % | **96.67 % [95.19, 97.70]** |
| surgical_technique | 810 | 810 | 570 | 100.00 % | **70.37 % [67.14, 73.41]** |
| tnm_descriptor | 810 | 810 | 791 | 100.00 % | **97.65 % [96.37, 98.49]** |
| tumor_size | 810 | 810 | 682 | 100.00 % | **84.20 % [81.52, 86.55]** |

**Supplementary Table S16. Per-field extraction accuracy for colorectal cancer excision reports (30-run aggregate, 24 schema fields).**

Per-field extraction accuracy of the per-organ extractor on the colorectal subset of the CMUH validation cohort, **30 multi-seed runs of gpt-oss-20b** against the committee-resolved gold standard. n = 2,160 case-runs per field (72 unique reports × 30 effective runs). This table **supersedes** the v1 single-run snapshot. Schema definition: Supplementary Table S3.

| **Field** | **n_total** | **n_attempted** | **n_correct** | **Coverage** | **Effective accuracy [95 % Wilson CI]** |
| --- | --- | --- | --- | --- | --- |
| biomarker_mlh1 | 2,160 | 2,040 | 2,040 | 94.44 % | **94.44 % [93.40, 95.33]** |
| biomarker_msh2 | 2,160 | 1,980 | 1,980 | 91.67 % | **91.67 % [90.43, 92.76]** |
| biomarker_msh6 | 2,160 | 2,040 | 2,040 | 94.44 % | **94.44 % [93.40, 95.33]** |
| biomarker_pms2 | 2,160 | 2,040 | 2,040 | 94.44 % | **94.44 % [93.40, 95.33]** |
| cancer_primary_site | 2,160 | 2,160 | 2,160 | 100.00 % | **100.00 % [99.82, 100.00]** |
| distant_metastasis | 2,160 | 2,160 | 505 | 100.00 % | **23.38 % [21.64, 25.21]** |
| extracellular_mucin | 2,160 | 2,160 | 2,160 | 100.00 % | **100.00 % [99.82, 100.00]** |
| extranodal_extension | 2,160 | 2,160 | 1,917 | 100.00 % | **88.75 % [87.35, 90.01]** |
| grade | 2,160 | 2,160 | 2,160 | 100.00 % | **100.00 % [99.82, 100.00]** |
| histology | 2,160 | 2,160 | 2,159 | 100.00 % | **99.95 % [99.74, 99.99]** |
| lymphovascular_invasion | 2,160 | 2,160 | 2,160 | 100.00 % | **100.00 % [99.82, 100.00]** |
| maximal_ln_size | 2,160 | 2,160 | 2,160 | 100.00 % | **100.00 % [99.82, 100.00]** |
| perineural_invasion | 2,160 | 2,160 | 2,160 | 100.00 % | **100.00 % [99.82, 100.00]** |
| pm_category | 2,160 | 2,160 | 2,033 | 100.00 % | **94.12 % [93.05, 95.04]** |
| pn_category | 2,160 | 2,160 | 2,092 | 100.00 % | **96.85 % [96.03, 97.51]** |
| procedure | 2,160 | 2,160 | 2,054 | 100.00 % | **95.09 % [94.10, 95.93]** |
| pt_category | 2,160 | 2,160 | 2,149 | 100.00 % | **99.49 % [99.09, 99.72]** |
| signet_ring | 2,160 | 2,160 | 2,157 | 100.00 % | **99.86 % [99.59, 99.95]** |
| stage_group | 2,160 | 2,160 | 2,067 | 100.00 % | **95.69 % [94.75, 96.47]** |
| surgical_technique | 2,160 | 2,160 | 1,973 | 100.00 % | **91.34 % [90.08, 92.46]** |
| tnm_descriptor | 2,160 | 2,160 | 2,157 | 100.00 % | **99.86 % [99.59, 99.95]** |
| tumor_budding | 2,160 | 2,160 | 2,160 | 100.00 % | **100.00 % [99.82, 100.00]** |
| tumor_invasion | 2,160 | 2,160 | 1,906 | 100.00 % | **88.24 % [86.81, 89.53]** |
| type_of_polyp | 2,160 | 2,160 | 2,139 | 100.00 % | **99.03 % [98.52, 99.36]** |

**Supplementary Table S17. Per-field extraction accuracy for esophageal cancer excision reports (30-run aggregate, 16 schema fields).**

Per-field extraction accuracy of the per-organ extractor on the esophagus subset of the CMUH validation cohort, **30 multi-seed runs of gpt-oss-20b** against the committee-resolved gold standard. n = 2,169 case-runs per field (73 unique reports × ~30 effective runs). This table **supersedes** the v1 single-run snapshot. Schema definition: Supplementary Table S4.

| **Field** | **n_total** | **n_attempted** | **n_correct** | **Coverage** | **Effective accuracy [95 % Wilson CI]** |
| --- | --- | --- | --- | --- | --- |
| cancer_primary_site | 2,169 | 2,169 | 2,009 | 100.00 % | **92.62 % [91.45, 93.65]** |
| distant_metastasis | 2,169 | 2,169 | 315 | 100.00 % | **14.52 % [13.10, 16.07]** |
| extranodal_extension | 2,169 | 2,169 | 2,132 | 100.00 % | **98.29 % [97.66, 98.76]** |
| grade | 2,169 | 2,169 | 2,169 | 100.00 % | **100.00 % [99.82, 100.00]** |
| histology | 2,169 | 2,169 | 2,169 | 100.00 % | **100.00 % [99.82, 100.00]** |
| lymphovascular_invasion | 2,169 | 2,169 | 2,169 | 100.00 % | **100.00 % [99.82, 100.00]** |
| maximal_ln_size | 2,169 | 2,169 | 2,169 | 100.00 % | **100.00 % [99.82, 100.00]** |
| perineural_invasion | 2,169 | 2,169 | 2,169 | 100.00 % | **100.00 % [99.82, 100.00]** |
| pm_category | 2,169 | 2,169 | 2,074 | 100.00 % | **95.62 % [94.68, 96.40]** |
| pn_category | 2,169 | 2,169 | 2,169 | 100.00 % | **100.00 % [99.82, 100.00]** |
| procedure | 2,169 | 2,169 | 2,129 | 100.00 % | **98.16 % [97.50, 98.64]** |
| pt_category | 2,169 | 2,169 | 2,166 | 100.00 % | **99.86 % [99.59, 99.95]** |
| stage_group | 2,169 | 2,169 | 2,082 | 100.00 % | **95.99 % [95.08, 96.74]** |
| surgical_technique | 2,169 | 2,169 | 2,134 | 100.00 % | **98.39 % [97.76, 98.84]** |
| tnm_descriptor | 2,169 | 2,169 | 2,122 | 100.00 % | **97.83 % [97.13, 98.37]** |
| tumor_extent | 2,169 | 2,169 | 2,125 | 100.00 % | **97.97 % [97.29, 98.49]** |

**Supplementary Table S18. Per-field extraction accuracy for hepatocellular carcinoma excision reports (30-run aggregate, 17 schema fields).**

Per-field extraction accuracy of the per-organ extractor on the liver subset of the CMUH validation cohort, **30 multi-seed runs of gpt-oss-20b** against the committee-resolved gold standard. n = 2,400 case-runs per field (80 unique reports × 30 effective runs). This table **supersedes** the v1 single-run snapshot. Schema definition: Supplementary Table S6.

| **Field** | **n_total** | **n_attempted** | **n_correct** | **Coverage** | **Effective accuracy [95 % Wilson CI]** |
| --- | --- | --- | --- | --- | --- |
| distant_metastasis | 2,400 | 2,400 | 84 | 100.00 % | **3.50 % [2.84, 4.31]** |
| extranodal_extension | 2,400 | 2,400 | 2,399 | 100.00 % | **99.96 % [99.76, 99.99]** |
| grade | 2,400 | 2,400 | 2,400 | 100.00 % | **100.00 % [99.84, 100.00]** |
| histology | 2,400 | 2,400 | 2,367 | 100.00 % | **98.63 % [98.08, 99.02]** |
| maximal_ln_size | 2,400 | 2,400 | 2,400 | 100.00 % | **100.00 % [99.84, 100.00]** |
| overall_stage | 2,400 | 2,400 | 2,336 | 100.00 % | **97.33 % [96.61, 97.91]** |
| perineural_invasion | 2,400 | 2,400 | 2,370 | 100.00 % | **98.75 % [98.22, 99.12]** |
| pm_category | 2,400 | 2,400 | 2,274 | 100.00 % | **94.75 % [93.78, 95.57]** |
| pn_category | 2,400 | 2,400 | 1,839 | 100.00 % | **76.62 % [74.89, 78.27]** |
| procedure | 2,400 | 2,400 | 2,041 | 100.00 % | **85.04 % [83.56, 86.41]** |
| pt_category | 2,400 | 2,400 | 2,382 | 100.00 % | **99.25 % [98.82, 99.53]** |
| tnm_descriptor | 2,400 | 2,400 | 2,042 | 100.00 % | **85.08 % [83.60, 86.45]** |
| tumor_extent | 2,400 | 2,400 | 2,370 | 100.00 % | **98.75 % [98.22, 99.12]** |
| tumor_focality | 2,400 | 2,400 | 2,400 | 100.00 % | **100.00 % [99.84, 100.00]** |
| tumor_site | 2,400 | 2,400 | 2,248 | 100.00 % | **93.67 % [92.62, 94.57]** |
| tumor_size | 2,400 | 2,400 | 2,236 | 100.00 % | **93.17 % [92.09, 94.11]** |
| vascular_invasion | 2,400 | 2,400 | 2,332 | 100.00 % | **97.17 % [96.42, 97.76]** |

**Supplementary Table S19. Per-field extraction accuracy for lung cancer excision reports (30-run aggregate, 20 schema fields).**

Per-field extraction accuracy of the per-organ extractor on the lung subset of the CMUH validation cohort, **30 multi-seed runs of gpt-oss-20b** against the committee-resolved gold standard. n = 1,650 case-runs per field (55 unique reports × 30 effective runs). This table **supersedes** the v1 single-run snapshot. Schema definition: Supplementary Table S2.

| **Field** | **n_total** | **n_attempted** | **n_correct** | **Coverage** | **Effective accuracy [95 % Wilson CI]** |
| --- | --- | --- | --- | --- | --- |
| cancer_primary_site | 1,650 | 1,650 | 1,649 | 100.00 % | **99.94 % [99.66, 99.99]** |
| direct_invasion_of_adjacent_structures | 1,650 | 1,650 | 1,518 | 100.00 % | **92.00 % [90.59, 93.21]** |
| distant_metastasis | 1,650 | 1,650 | 215 | 100.00 % | **13.03 % [11.49, 14.74]** |
| extranodal_extension | 1,650 | 1,556 | 1,533 | 94.30 % | **92.91 % [91.57, 94.05]** (att 98.52 %) |
| grade | 1,650 | 1,650 | 1,628 | 100.00 % | **98.67 % [97.99, 99.12]** |
| histology | 1,650 | 1,650 | 1,638 | 100.00 % | **99.27 % [98.73, 99.58]** |
| lymphovascular_invasion | 1,650 | 1,650 | 1,618 | 100.00 % | **98.06 % [97.28, 98.62]** |
| maximal_ln_size | 1,650 | 1,556 | 1,537 | 94.30 % | **93.15 % [91.83, 94.27]** (att 98.78 %) |
| perineural_invasion | 1,650 | 1,650 | 1,380 | 100.00 % | **83.64 % [81.77, 85.34]** |
| pm_category | 1,650 | 1,647 | 1,491 | 99.82 % | **90.36 % [88.84, 91.70]** |
| pn_category | 1,650 | 1,647 | 1,593 | 99.82 % | **96.55 % [95.55, 97.32]** |
| procedure | 1,650 | 1,650 | 1,615 | 100.00 % | **97.88 % [97.06, 98.47]** |
| pt_category | 1,650 | 1,647 | 1,592 | 99.82 % | **96.48 % [95.48, 97.27]** |
| sideness | 1,650 | 1,650 | 1,650 | 100.00 % | **100.00 % [99.77, 100.00]** |
| spread_through_air_spaces_stas | 1,650 | 1,650 | 1,585 | 100.00 % | **96.06 % [95.01, 96.90]** |
| stage_group | 1,650 | 1,647 | 1,533 | 99.82 % | **92.91 % [91.57, 94.05]** |
| surgical_technique | 1,650 | 1,650 | 1,459 | 100.00 % | **88.42 % [86.79, 89.88]** |
| tnm_descriptor | 1,650 | 1,647 | 1,454 | 99.82 % | **88.12 % [86.47, 89.59]** |
| tumor_focality | 1,650 | 1,650 | 1,547 | 100.00 % | **93.76 % [92.49, 94.83]** |
| visceral_pleural_invasion | 1,650 | 1,650 | 1,611 | 100.00 % | **97.64 % [96.79, 98.27]** |

**Supplementary Table S20. Per-field extraction accuracy for pancreas cancer excision reports (30-run aggregate, 15 schema fields).**

Per-field extraction accuracy of the per-organ extractor on the pancreas subset of the CMUH validation cohort, **30 multi-seed runs of gpt-oss-20b** against the committee-resolved gold standard. n = 750 case-runs per field (25 unique reports × 30 effective runs). This table **supersedes** the v1 single-run snapshot. Schema definition: Supplementary Table S7.

| **Field** | **n_total** | **n_attempted** | **n_correct** | **Coverage** | **Effective accuracy [95 % Wilson CI]** |
| --- | --- | --- | --- | --- | --- |
| distant_metastasis | 750 | 750 | 89 | 100.00 % | **11.87 % [9.74, 14.38]** |
| extranodal_extension | 750 | 724 | 706 | 96.53 % | **94.13 % [92.22, 95.60]** (att 97.51 %) |
| histology | 750 | 750 | 727 | 100.00 % | **96.93 % [95.44, 97.95]** |
| lymphovascular_invasion | 750 | 750 | 750 | 100.00 % | **100.00 % [99.49, 100.00]** |
| maximal_ln_size | 750 | 724 | 723 | 96.53 % | **96.40 % [94.81, 97.51]** (att 99.86 %) |
| overall_stage | 750 | 750 | 750 | 100.00 % | **100.00 % [99.49, 100.00]** |
| perineural_invasion | 750 | 750 | 750 | 100.00 % | **100.00 % [99.49, 100.00]** |
| pm_category | 750 | 750 | 722 | 100.00 % | **96.27 % [94.66, 97.40]** |
| pn_category | 750 | 750 | 750 | 100.00 % | **100.00 % [99.49, 100.00]** |
| procedure | 750 | 750 | 679 | 100.00 % | **90.53 % [88.23, 92.43]** |
| pt_category | 750 | 750 | 750 | 100.00 % | **100.00 % [99.49, 100.00]** |
| tnm_descriptor | 750 | 750 | 611 | 100.00 % | **81.47 % [78.53, 84.08]** |
| tumor_extension | 750 | 750 | 578 | 100.00 % | **77.07 % [73.92, 79.93]** |
| tumor_site | 750 | 750 | 720 | 100.00 % | **96.00 % [94.35, 97.18]** |
| tumor_size | 750 | 750 | 662 | 100.00 % | **88.27 % [85.77, 90.38]** |

**Supplementary Table S21. Per-field extraction accuracy for prostate cancer excision reports (30-run aggregate, 28 schema fields).**

Per-field extraction accuracy of the per-organ extractor on the prostate subset of the CMUH validation cohort, **30 multi-seed runs of gpt-oss-20b** against the committee-resolved gold standard. n = 2,820 case-runs per field (94 unique reports × 30 effective runs). This table **supersedes** the v1 single-run snapshot. Schema definition: Supplementary Table S8.

| **Field** | **n_total** | **n_attempted** | **n_correct** | **Coverage** | **Effective accuracy [95 % Wilson CI]** |
| --- | --- | --- | --- | --- | --- |
| bladder_invasion | 2,820 | 2,820 | 2,802 | 100.00 % | **99.36 % [98.99, 99.60]** |
| cribriform_pattern_presence | 2,820 | 2,820 | 2,694 | 100.00 % | **95.53 % [94.71, 96.23]** |
| distant_metastasis | 2,820 | 2,820 | 77 | 100.00 % | **2.73 % [2.19, 3.40]** |
| extranodal_extension | 2,820 | 2,818 | 2,753 | 99.93 % | **97.62 % [96.99, 98.12]** |
| extraprostatic_extension | 2,820 | 2,820 | 2,820 | 100.00 % | **100.00 % [99.86, 100.00]** |
| gleason_4_percentage | 2,820 | 2,820 | 2,737 | 100.00 % | **97.06 % [96.37, 97.62]** |
| gleason_5_percentage | 2,820 | 2,820 | 2,727 | 100.00 % | **96.70 % [95.98, 97.30]** |
| grade | 2,820 | 2,820 | 2,807 | 100.00 % | **99.54 % [99.21, 99.73]** |
| histology | 2,820 | 2,820 | 2,815 | 100.00 % | **99.82 % [99.59, 99.92]** |
| intraductal_carcinoma_presence | 2,820 | 2,820 | 2,807 | 100.00 % | **99.54 % [99.21, 99.73]** |
| involved_margin_list | 2,820 | 2,809 | 2,616 | 99.61 % | **92.77 % [91.75, 93.67]** (att 93.13 %) |
| lymphovascular_invasion | 2,820 | 2,820 | 2,820 | 100.00 % | **100.00 % [99.86, 100.00]** |
| margin_length | 2,820 | 2,809 | 2,809 | 99.61 % | **99.61 % [99.30, 99.78]** (att 100.00 %) |
| margin_positivity | 2,820 | 2,809 | 2,808 | 99.61 % | **99.57 % [99.26, 99.76]** (att 99.96 %) |
| maximal_ln_size | 2,820 | 2,818 | 2,817 | 99.93 % | **99.89 % [99.69, 99.96]** |
| perineural_invasion | 2,820 | 2,820 | 2,820 | 100.00 % | **100.00 % [99.86, 100.00]** |
| pm_category | 2,820 | 2,820 | 2,570 | 100.00 % | **91.13 % [90.03, 92.13]** |
| pn_category | 2,820 | 2,820 | 2,819 | 100.00 % | **99.96 % [99.80, 99.99]** |
| procedure | 2,820 | 2,820 | 2,820 | 100.00 % | **100.00 % [99.86, 100.00]** |
| prostate_size | 2,820 | 2,820 | 2,762 | 100.00 % | **97.94 % [97.35, 98.41]** |
| prostate_weight | 2,820 | 2,820 | 2,816 | 100.00 % | **99.86 % [99.64, 99.94]** |
| pt_category | 2,820 | 2,820 | 2,820 | 100.00 % | **100.00 % [99.86, 100.00]** |
| seminal_vesicle_invasion | 2,820 | 2,820 | 2,820 | 100.00 % | **100.00 % [99.86, 100.00]** |
| stage_group | 2,820 | 2,820 | 2,657 | 100.00 % | **94.22 % [93.30, 95.02]** |
| surgical_technique | 2,820 | 2,820 | 2,813 | 100.00 % | **99.75 % [99.49, 99.88]** |
| tnm_descriptor | 2,820 | 2,820 | 2,205 | 100.00 % | **78.19 % [76.63, 79.68]** |
| tumor_percentage | 2,820 | 2,820 | 2,749 | 100.00 % | **97.48 % [96.84, 97.99]** |
| tumor_size | 2,820 | 2,820 | 2,818 | 100.00 % | **99.93 % [99.74, 99.98]** |

**Supplementary Table S22. Per-field extraction accuracy for gastric cancer excision reports (30-run aggregate, 18 schema fields).**

Per-field extraction accuracy of the per-organ extractor on the gastric subset of the CMUH validation cohort, **30 multi-seed runs of gpt-oss-20b** against the committee-resolved gold standard. n = 2,340 case-runs per field (78 unique reports × 30 effective runs). This table **supersedes** the v1 single-run snapshot. Schema definition: Supplementary Table S5.

| **Field** | **n_total** | **n_attempted** | **n_correct** | **Coverage** | **Effective accuracy [95 % Wilson CI]** |
| --- | --- | --- | --- | --- | --- |
| cancer_primary_site | 2,340 | 2,340 | 1,978 | 100.00 % | **84.53 % [83.01, 85.94]** |
| distant_metastasis | 2,340 | 2,340 | 292 | 100.00 % | **12.48 % [11.20, 13.88]** |
| extracellular_mucin | 2,340 | 2,340 | 2,340 | 100.00 % | **100.00 % [99.84, 100.00]** |
| extranodal_extension | 2,340 | 2,265 | 2,009 | 96.79 % | **85.85 % [84.38, 87.21]** (att 88.70 %) |
| grade | 2,340 | 2,340 | 2,340 | 100.00 % | **100.00 % [99.84, 100.00]** |
| histology | 2,340 | 2,340 | 2,261 | 100.00 % | **96.62 % [95.81, 97.28]** |
| lymphovascular_invasion | 2,340 | 2,340 | 2,340 | 100.00 % | **100.00 % [99.84, 100.00]** |
| maximal_ln_size | 2,340 | 2,265 | 2,264 | 96.79 % | **96.75 % [95.95, 97.40]** (att 99.96 %) |
| perineural_invasion | 2,340 | 2,340 | 2,340 | 100.00 % | **100.00 % [99.84, 100.00]** |
| pm_category | 2,340 | 2,340 | 2,152 | 100.00 % | **91.97 % [90.79, 93.00]** |
| pn_category | 2,340 | 2,340 | 2,338 | 100.00 % | **99.91 % [99.69, 99.98]** |
| procedure | 2,340 | 2,340 | 2,276 | 100.00 % | **97.26 % [96.52, 97.85]** |
| pt_category | 2,340 | 2,340 | 2,340 | 100.00 % | **100.00 % [99.84, 100.00]** |
| signet_ring | 2,340 | 2,340 | 2,340 | 100.00 % | **100.00 % [99.84, 100.00]** |
| stage_group | 2,340 | 2,340 | 2,305 | 100.00 % | **98.50 % [97.93, 98.92]** |
| surgical_technique | 2,340 | 2,340 | 1,135 | 100.00 % | **48.50 % [46.48, 50.53]** |
| tnm_descriptor | 2,340 | 2,340 | 1,925 | 100.00 % | **82.26 % [80.66, 83.76]** |
| tumor_extent | 2,340 | 2,340 | 2,282 | 100.00 % | **97.52 % [96.81, 98.08]** |

**Supplementary Table S23. Per-field extraction accuracy for thyroid cancer excision reports (30-run aggregate, 18 schema fields).**

Per-field extraction accuracy of the per-organ extractor on the thyroid subset of the CMUH validation cohort, **30 multi-seed runs of gpt-oss-20b** against the committee-resolved gold standard. n = 2,160 case-runs per field (72 unique reports × 30 effective runs). This table **supersedes** the v1 single-run snapshot. Schema definition: Supplementary Table S10.

| **Field** | **n_total** | **n_attempted** | **n_correct** | **Coverage** | **Effective accuracy [95 % Wilson CI]** |
| --- | --- | --- | --- | --- | --- |
| distant_metastasis | 2,160 | 2,160 | 1,524 | 100.00 % | **70.56 % [68.60, 72.44]** |
| extranodal_extension | 2,160 | 2,160 | 1,980 | 100.00 % | **91.67 % [90.43, 92.76]** |
| extrathyroid_extension | 2,160 | 2,160 | 2,114 | 100.00 % | **97.87 % [97.17, 98.40]** |
| histology | 2,160 | 2,160 | 2,131 | 100.00 % | **98.66 % [98.08, 99.06]** |
| lymphovascular_invasion | 2,160 | 2,160 | 1,631 | 100.00 % | **75.51 % [73.65, 77.28]** |
| maximal_ln_size | 2,160 | 2,160 | 2,128 | 100.00 % | **98.52 % [97.92, 98.95]** |
| mitotic_activity | 2,160 | 2,160 | 2,129 | 100.00 % | **98.56 % [97.97, 98.99]** |
| overall_stage | 2,160 | 2,160 | 2,079 | 100.00 % | **96.25 % [95.36, 96.97]** |
| perineural_invasion | 2,160 | 2,160 | 1,602 | 100.00 % | **74.17 % [72.28, 75.97]** |
| pm_category | 2,160 | 2,160 | 2,022 | 100.00 % | **93.61 % [92.50, 94.57]** |
| pn_category | 2,160 | 2,160 | 2,006 | 100.00 % | **92.87 % [91.71, 93.88]** |
| predisposing_condition | 2,160 | 2,160 | 2,153 | 100.00 % | **99.68 % [99.33, 99.84]** |
| procedure | 2,160 | 2,160 | 1,916 | 100.00 % | **88.70 % [87.30, 89.97]** |
| pt_category | 2,160 | 2,160 | 2,159 | 100.00 % | **99.95 % [99.74, 99.99]** |
| tnm_descriptor | 2,160 | 2,160 | 1,970 | 100.00 % | **91.20 % [89.93, 92.33]** |
| tumor_focality | 2,160 | 2,160 | 2,069 | 100.00 % | **95.79 % [94.86, 96.56]** |
| tumor_necrosis | 2,160 | 2,160 | 1,839 | 100.00 % | **85.14 % [83.58, 86.58]** |
| tumor_site | 2,160 | 2,160 | 2,044 | 100.00 % | **94.63 % [93.60, 95.50]** |
| tumor_size | 2,160 | 2,160 | 2,122 | 100.00 % | **98.24 % [97.59, 98.72]** |

**Supplementary Table S24. Detailed numeric results for lymph-node extraction accuracy by organ.**

Aggregate lymph-node extraction concordance across the original CMUH validation cohort (n = 893 reports). Carried over from the v1 manuscript supplementary (single-run snapshot). The headline 30-run aggregates that supersede this in the revised text are at Supplementary Table S34 (per-organ LN summary, k = 30) and Figure 4. Two metrics + four descriptive rows; the prevalence-vs-correctness split is not separated in the v1 design (the v2 design in Supplementary Table S34 separates examined-count, examined-tol-1, group-recall, and group-precision into four explicit metrics).

| **Metric** | **breast** | **cervix** | **colorectal** | **esophagus** | **liver** | **lung** | **pancreas** | **prostate** | **stomach** | **thyroid** |
| --- | --- | --- | --- | --- | --- | --- | --- | --- | --- | --- |
| lymph node number concordance rate | 93.30 % | 86.20 % | 97.10 % | 49.30 % | 98.70 % | 82.50 % | 92.80 % | 59.60 % | 83.50 % | 91.80 % |
| lymph node station concordance rate | 80.00 % | 71.40 % | 96.90 % | 49.30 % | 100.00 % | 77.70 % | 85.50 % | 58.20 % | 91.00 % | 93.00 % |
| lymph node stations (total annotated) | 95 | 77 | 127 | 454 | 12 | 215 | 62 | 275 | 401 | 43 |
| report numbers | 75 | 29 | 70 | 73 | 79 | 63 | 28 | 94 | 79 | 74 |
| station per report | 1.27 | 2.66 | 1.81 | 6.22 | 0.15 | 3.41 | 2.21 | 2.93 | 5.08 | 0.58 |

**Supplementary Table S25. Eligibility-classifier 30-run aggregate.**

Aggregate reliability statistics for the eligibility classifier (Stage A) across the 30 multi-seed runs of gpt-oss-20b on the CMUH validation cohort (n = 893 reports → 26,790 case-runs). Reports the headline reliability triplet (single-run ICC, average-rater ICC, Cronbach's α) plus accuracy flip rate and per-case SD summary. This is the headline reliability number cited in §3.2 / paragraph 06 (Table 4b), §3.8 / paragraph 16, and the overall multi-run reliability discussion. Replaces the single-run / 3-run confusion matrices at Supplementary Tables S11–S13 as the headline number for Stage A.

| **Statistic** | **Value** |
| --- | --- |
| Chapter | 1 (chapter1_eligibility) |
| Stage | A |
| n_cases | 893 |
| n_runs | 30 |
| n_case-runs | 26,790 |
| ICC(2,1) — single-run reproducibility | **0.8893** [0.8798, 0.8985] |
| ICC(3, k = 30) — average-of-30-runs | **0.9959** [0.9955, 0.9963] |
| Cronbach's α | 0.9959 |
| Accuracy flip rate (per-case across 30 runs) | **3.58 %** |
| Per-case SD mean (binary accuracy) | 0.0123 |
| Per-case SD 90th percentile | 0.0000 |

**Supplementary Table S26. Organ-classification 30-run aggregate.**

Aggregate reliability statistics for the 11-class organ-labeling sub-model (Stage B) across the 30 multi-seed runs of gpt-oss-20b on the CMUH validation cohort, scored only on the 682 reports that cleared the eligibility classifier (n = 682 reports → 20,460 case-runs evaluated; 71 case-runs filtered as triage failures). This is the headline reliability number cited in §3.2 / paragraph 06 and the v2 replacement for the 3-run confusion matrices at Supplementary Tables S11–S13.

| **Statistic** | **Value** |
| --- | --- |
| Chapter | 2 (chapter2_organ_classification) |
| Stage | B |
| n_cases | 682 |
| n_runs | 30 |
| n_case-runs | 20,460 (after Stage-A filter) |
| ICC(2,1) — single-run reproducibility | **0.8996** [0.8896, 0.9092] |
| ICC(3, k = 30) — average-of-30-runs | **0.9963** [0.9959, 0.9967] |
| Cronbach's α | 0.9963 |
| Accuracy flip rate (per-case across 30 runs) | **1.76 %** |
| Per-case SD mean (binary accuracy) | 0.0069 |
| Per-case SD 90th percentile | 0.0000 |

**Supplementary Table S27. Per-organ extraction-accuracy headline (k = 30, paper scope).**

Per-organ effective extraction accuracy on the CMUH validation cohort, aggregated across 30 multi-seed runs of gpt-oss-20b. The "paper scope" denominator excludes biomarker sub-fields scored separately in §3.5 (breast ER/PR/HER2/Ki-67) and the prostate-specific margin scalars scored separately in §3.4 (Supplementary Table S33). This is the headline per-organ table cited in §3.3 / paragraph 08 (Table 5) and Figure 3; sorted descending by effective accuracy.

| **Organ** | **n_fields** | **n_attempted (case × run × field)** | **n_correct** | **Effective accuracy [95 % Wilson CI]** |
| --- | --- | --- | --- | --- |
| prostate | 25 | 70,496 | 66,165 | **93.86 % [93.68, 94.03]** |
| colorectal | 20 | 43,200 | 40,428 | **93.58 % [93.35, 93.81]** |
| esophagus | 16 | 34,704 | 32,302 | **93.08 % [92.81, 93.34]** |
| thyroid | 19 | 41,040 | 37,618 | **91.66 % [91.39, 91.93]** |
| breast | 26 | 58,442 | 53,482 | **91.51 % [91.28, 91.74]** |
| lung | 20 | 32,797 | 29,846 | **91.00 % [90.69, 91.31]** |
| liver | 17 | 40,800 | 36,520 | **89.51 % [89.21, 89.80]** |
| pancreas | 15 | 11,198 | 9,967 | **89.01 % [88.41, 89.57]** |
| stomach | 18 | 41,970 | 37,257 | **88.77 % [88.46, 89.07]** |
| cervix | 16 | 12,940 | 11,247 | **86.92 % [86.32, 87.49]** |
| **Macro mean** |  |  |  | **90.89 %** |

**Supplementary Table S28. Anatomic vs pathologic stage-group head-to-head (paired breast case-runs).**

Per-field exact-match accuracy for anatomic_stage_group and pathologic_stage_group on the breast subset of the CMUH validation cohort, paired by case × run across the 30-seed pass (n = 2,248 paired comparisons; 75 cases × ~30 effective runs after Stage A/B gating). The two fields share the same case-run denominator (the same set of breast case-runs is asked to populate both fields), so the 23.09 percentage-point gap between them is a within-case effect — not a sampling difference. This is the headline number for §3.3.1 / paragraph 09 (Table 6, Figure 6) and the qualitative case study in Appendix A4.

| **Field** | **n_total** | **n_correct** | **Effective accuracy [95 % Wilson CI]** |
| --- | --- | --- | --- |
| anatomic_stage_group | 2,248 | 2,202 | **97.95 % [97.28, 98.46]** |
| pathologic_stage_group | 2,248 | 1,683 | **74.87 % [73.03, 76.62]** |
| **Δ (anatomic − pathologic)** | — | — | **+23.09 pp** (95 % Wilson intervals do not overlap) |

**Supplementary Table S29. Bottom-decile field error attribution.**

Bottom-decile fields by effective accuracy across the CMUH 30-run pass, with each field tagged by mechanism (M1–M6, or audit-pending). Eleven fields, sorted ascending by effective accuracy; the lowest is distant_metastasis at 19.06 %. Backs §3.3.2 / paragraph 10 (Figure 8, Table 9). The six mechanisms separate the proximate cause of the low score by its actual engineering fix path so that the discussion in paragraph 10 can address each independently.

| **Field** | **Mechanism** | **n_total** | **n_correct** | **Effective accuracy [95 % Wilson CI]** | **Coverage** | **Cohen's κ vs gold** |
| --- | --- | --- | --- | --- | --- | --- |
| distant_metastasis | M4_convention | 19,507 | 3,718 | **19.06 % [18.51, 19.62]** | 1.000 | 0.947 |
| cancer_quadrant | M2_glossary_rule | 2,248 | 1,629 | **72.46 % [70.58, 74.27]** | 1.000 | 0.745 |
| pathologic_stage_group | M5_schema_shape | 2,248 | 1,683 | **74.87 % [73.03, 76.62]** | 1.000 | 0.927 |
| tumor_extension | — (audit pending) | 750 | 578 | **77.07 % [73.92, 79.93]** | 1.000 | 0.609 |
| biomarker_her2 | M5_schema_shape | 2,248 | 1,738 | **77.31 % [75.54, 79.00]** | 1.000 | — |
| surgical_technique | M2_glossary_rule | 11,949 | 10,084 | **84.39 % [83.73, 85.03]** | 1.000 | 0.955 |
| tumor_necrosis | M2_glossary_rule | 2,160 | 1,839 | **85.14 % [83.58, 86.58]** | 1.000 | — |
| cancer_clock | M2_glossary_rule | 2,248 | 1,918 | **85.32 % [83.80, 86.72]** | 1.000 | 0.949 |
| biomarker_ki67 | M5_schema_shape | 2,248 | 1,939 | **86.25 % [84.77, 87.62]** | 1.000 | — |
| tumor_invasion | M6_open_conflation | 2,160 | 1,906 | **88.24 % [86.81, 89.53]** | 1.000 | 0.837 |
| tnm_descriptor | M5_schema_shape | 19,507 | 17,337 | **88.88 % [88.43, 89.31]** | 1.000 | 0.954 |

**Supplementary Table S30. Multi-primary Stage-B disposition breakdown.**

The multi-primary triage decomposition for the gold-others ledger across the 30-run pass on the CMUH validation cohort. Stage-B disposition rows (both_others = both annotators agreed the case is out of scope; gold_only = gold says others, model emitted an in-scope organ; pred_only = model emitted others, gold is in scope) are decomposed by clinical flavour (out-of-scope site, synchronous multi-primary, uncertain). This is the headline number for §3.2.1 / paragraph 07 (Table 7, Figure 7).

| **Stage-B disposition** | **out_of_scope_site** | **synchronous_multi_primary** | **uncertain** | **row total** |
| --- | --- | --- | --- | --- |
| both_others | 437 | 0 | 12 | 449 |
| gold_only | 163 | 180 | 288 | 631 |
| pred_only | 22 | 0 | 36 | 58 |
| **column total** | **622** | **180** | **336** | **1,138** |

**Supplementary Table S31. Per-organ misroute destinations for gold-`others` reports.**

When the gold annotator codes a report as others and the model emits an in-scope organ instead, this table records which in-scope organ the model picks. Counts are case × run cells (n = 631 total across the 30-run pass — the gold_only row of Supplementary Table S30). Sorted descending by count.

| **Gold cancer_category** | **Predicted cancer_category** | **n** |
| --- | --- | --- |
| others | lung | 150 |
| others | (null / no prediction) | 117 |
| others | pancreas | 108 |
| others | stomach | 60 |
| others | colorectal | 60 |
| others | cervix | 50 |
| others | breast | 30 |
| others | liver | 30 |
| others | esophagus | 26 |
| **total** | — | **631** |

**Supplementary Table S32. Per-organ margin extraction summary (positivity, distance accuracy, hallucination, miss).**

Per-organ margin-extraction metrics on the CMUH 30-run pass: list-level F1 plus the hallucination-vs-miss decomposition of the residual error. Nine organs (prostate excluded — its margin design uses a separate binary margin_positivity + involved_margin_list schema, see Supplementary Tables S8 and S33). Backs §3.4 / paragraph 11 (Supplementary Figures S13 + S12, Table 10). The hallucination_rate column is the dominant residual error: in seven of nine organs hallucination is at least 5× the miss rate, with lung exhibiting the most extreme schema-driven over-prediction (49.8 % hallucination vs 2.8 % miss).

| **Organ** | **n_total** | **n_attempted** | **Coverage** | **Any-involved correctness [95 % CI]** | **Closest-dist MAE (mm)** | **Closest-dist accuracy (±2 mm tol)** | **Micro F1** | **Hallucination rate [95 % CI]** | **Miss rate [95 % CI]** |
| --- | --- | --- | --- | --- | --- | --- | --- | --- | --- |
| breast | 2,248 | 2,248 | 100.0 % | **93.91 % [92.84, 94.82]** | 0.21 | 97.98 % | 0.9506 | 9.30 % [8.84, 9.78] | 0.14 % [0.09, 0.22] |
| cervix | 810 | 810 | 100.0 % | **99.51 % [98.74, 99.81]** | 0.81 | 93.86 % | 0.8905 | 13.50 % [12.18, 14.94] | 8.24 % [7.17, 9.46] |
| colorectal | 2,160 | 2,082 | 96.4 % | **98.46 % [97.84, 98.91]** | 7.15 | 73.54 % | 0.9573 | 5.79 % [5.32, 6.30] | 2.70 % [2.37, 3.07] |
| esophagus | 2,169 | 2,168 | 99.95 % | **96.08 % [95.18, 96.82]** | 4.90 | 78.40 % | 0.8373 | 26.89 % [25.99, 27.81] | 2.03 % [1.72, 2.39] |
| liver | 2,400 | 2,400 | 100.0 % | **98.71 % [98.17, 99.09]** | 0.21 | 97.11 % | 0.7832 | 33.52 % [32.22, 34.84] | 4.71 % [4.06, 5.47] |
| lung | 1,650 | 1,647 | 99.8 % | **98.42 % [97.70, 98.92]** | 9.25 | 46.45 % | 0.6617 | **49.83 % [48.54, 51.13]** | 2.83 % [2.29, 3.50] |
| pancreas | 750 | 737 | 98.3 % | **93.62 % [91.62, 95.17]** | 4.38 | 85.01 % | 0.9189 | 11.43 % [10.50, 12.44] | 4.52 % [3.90, 5.22] |
| stomach | 2,340 | 2,340 | 100.0 % | **98.25 % [97.63, 98.71]** | 5.67 | 80.50 % | 0.8485 | 25.18 % [24.19, 26.20] | 2.02 % [1.68, 2.42] |
| thyroid | 2,160 | 2,160 | 100.0 % | **99.58 % [99.21, 99.78]** | 0.52 | 97.57 % | 0.7613 | 37.88 % [36.70, 39.08] | 1.69 % [1.34, 2.14] |
| (prostate) | 2,820 | 0 | 0.0 % | — | — | — | — | — | — |

**Supplementary Table S33. Prostate-specific margin extraction callout.**

The prostate schema (Supplementary Table S8) uses a binary margin_positivity flag plus a list-typed involved_margin_list (12 named directions) instead of the per-margin list-of-records design used by the other nine organs. Three prostate-specific margin scalars are scored independently of the §3.4 margin metrics in Supplementary Table S32; this table records their accuracy on the 30-run pass. Backs §3.4 / paragraph 11 (Table 10b).

| **Field** | **n_total** | **n_correct** | **Effective accuracy [95 % Wilson CI]** |
| --- | --- | --- | --- |
| margin_positivity | 2,820 | 2,808 | **99.57 % [99.26, 99.76]** |
| margin_length | 2,820 | 2,809 | **99.61 % [99.30, 99.78]** |
| involved_margin_list | 2,820 | 2,616 | **92.77 % [91.75, 93.67]** |

**Supplementary Table S34. Per-organ lymph-node extraction summary (30-run, four-metric decomposition).**

Per-organ lymph-node-extraction metrics on the CMUH 30-run pass, decomposed into four explicit measurements (examined-count tolerance-1 accuracy, any-positive correctness, group recall, group precision). Replaces the two-metric v1 design in Supplementary Table S24 with separate column for each measurement. Backs §3.4 / paragraph 12 (Figure 4, Table 11). Sorted by group_recall descending; esophagus, cervix, and prostate are flagged as the low-recall tier and have station-name canonicalization callouts in Figure 4.

| **Organ** | **n_total** | **n_attempted** | **Coverage** | **Examined-count MAE** | **Examined accuracy (±1 tol)** | **Any-positive correctness [95 % CI]** | **Group recall** | **Group precision** | **Hallucination rate** | **Miss rate** |
| --- | --- | --- | --- | --- | --- | --- | --- | --- | --- | --- |
| liver | 2,400 | 2,400 | 100.0 % | 0.04 | 98.75 % | **100.0 % [99.8, 100.0]** | **0.980** | 0.993 | 0.68 % | 2.00 % |
| colorectal | 2,160 | 2,160 | 100.0 % | 0.33 | 95.97 % | **99.8 % [99.6, 99.9]** | **0.973** | 0.974 | 3.33 % | 3.16 % |
| stomach | 2,340 | 2,265 | 96.8 % | 2.64 | 81.55 % | **99.7 % [99.4, 99.9]** | **0.837** | 0.945 | 4.33 % | 14.62 % |
| breast | 2,248 | 2,245 | 99.9 % | 0.42 | 91.31 % | **99.9 % [99.6, 100.0]** | **0.844** | 0.936 | 6.00 % | 15.76 % |
| thyroid | 2,160 | 2,160 | 100.0 % | 0.69 | 89.95 % | **98.3 % [97.7, 98.8]** | **0.717** | 0.793 | 21.83 % | 28.81 % |
| pancreas | 750 | 724 | 96.5 % | 0.64 | 90.33 % | **100.0 % [99.5, 100.0]** | **0.776** | 0.835 | 15.22 % | 22.49 % |
| lung | 1,650 | 1,556 | 94.3 % | 1.15 | 89.72 % | **100.0 % [99.8, 100.0]** | **0.783** | 0.866 | 13.94 % | 22.74 % |
| prostate | 2,820 | 2,818 | 99.9 % | 7.02 | 61.57 % | **100.0 % [99.9, 100.0]** | **0.629** | 0.945 | 5.87 % | 37.05 % |
| cervix | 810 | 800 | 98.8 % | 3.14 | 67.00 % | **100.0 % [99.5, 100.0]** | **0.493** | 0.542 | 36.07 % | 49.32 % |
| esophagus | 2,169 | 2,169 | 100.0 % | 10.45 | 45.50 % | **100.0 % [99.8, 100.0]** | **0.426** | 0.826 | 13.09 % | 58.22 % |

**Supplementary Table S35. Multi-run reliability (k = 30) — full per-chapter, per-organ detail.**

Multi-run reliability statistics across all four chapters of the evaluation pipeline (chapter 1 eligibility, chapter 2 organ classification, chapter 4 margins, chapter 5 lymph nodes) on the CMUH 30-run pass. Chapters 1 and 2 are scored under ICC (binary classification across runs); chapters 4 and 5 are scored under per-case F1 standard deviation (list-typed extraction, where ICC is not well-defined). This is the headline reliability table cited in §2.4.2 (Methods, paragraph 02) and §3.8 (paragraph 16), with companion plots in Supplementary Figures S14 and S15. Twenty-one rows: 2 upstream classification stages + 9 organ-level margins + 10 organ-level lymph nodes.

| **Chapter** | **Stage** | **Organ** | **Field** | **n_cases** | **n_runs** | **ICC(2,1) [95 % CI]** | **ICC(3, k = 30) [95 % CI]** | **Cronbach α** | **Acc flip rate** | **Per-case SD mean** | **Per-case SD p90** | **Mean per-case F1** | **F1 SD mean** | **F1 SD p90** | **Missing flip rate** |
| --- | --- | --- | --- | --- | --- | --- | --- | --- | --- | --- | --- | --- | --- | --- | --- |
| 1 | A | — | — | 893 | 30 | 0.8893 [0.8798, 0.8985] | **0.9959 [0.9955, 0.9963]** | 0.9959 | 3.58 % | 0.0123 | 0.0000 | — | — | — | — |
| 2 | B | — | — | 682 | 30 | 0.8996 [0.8896, 0.9092] | **0.9963 [0.9959, 0.9967]** | 0.9963 | 1.76 % | 0.0069 | 0.0000 | — | — | — | — |
| 4 | — | breast | margins | 74 | 30 | — | — | — | — | — | — | **0.9572** | 0.0249 | 0.0454 | 0.0000 |
| 4 | — | cervix | margins | 27 | 30 | — | — | — | — | — | — | **0.8945** | 0.0947 | 0.1554 | 0.0000 |
| 4 | — | colorectal | margins | 72 | 30 | — | — | — | — | — | — | **0.9422** | 0.0458 | 0.0767 | 0.5833 |
| 4 | — | esophagus | margins | 73 | 30 | — | — | — | — | — | — | **0.8501** | 0.0889 | 0.1224 | 0.0141 |
| 4 | — | liver | margins | 80 | 30 | — | — | — | — | — | — | **0.8224** | 0.0911 | 0.2424 | 0.0000 |
| 4 | — | lung | margins | 55 | 30 | — | — | — | — | — | — | **0.6852** | 0.1621 | 0.2414 | 0.0545 |
| 4 | — | pancreas | margins | 25 | 30 | — | — | — | — | — | — | **0.8814** | 0.0769 | 0.1851 | 0.2000 |
| 4 | — | stomach | margins | 79 | 30 | — | — | — | — | — | — | **0.8726** | 0.1105 | 0.1711 | 0.0000 |
| 4 | — | thyroid | margins | 72 | 30 | — | — | — | — | — | — | **0.7900** | 0.1137 | 0.2122 | 0.0000 |
| 5 | — | breast | regional_lymph_node | 66 | 30 | — | — | — | — | — | — | **0.8444** | 0.2154 | 0.4552 | 0.0270 |
| 5 | — | cervix | regional_lymph_node | 21 | 30 | — | — | — | — | — | — | **0.4888** | 0.3620 | 0.4714 | 0.2593 |
| 5 | — | colorectal | regional_lymph_node | 67 | 30 | — | — | — | — | — | — | **0.9659** | 0.0588 | 0.1795 | 0.0000 |
| 5 | — | esophagus | regional_lymph_node | 73 | 30 | — | — | — | — | — | — | **0.4444** | 0.3537 | 0.4714 | 0.0000 |
| 5 | — | liver | regional_lymph_node | 11 | 30 | — | — | — | — | — | — | **0.9800** | 0.0552 | 0.1988 | 0.0000 |
| 5 | — | lung | regional_lymph_node | 45 | 30 | — | — | — | — | — | — | **0.8067** | 0.2760 | 0.4193 | 0.1091 |
| 5 | — | pancreas | regional_lymph_node | 24 | 30 | — | — | — | — | — | — | **0.7785** | 0.2299 | 0.4422 | 0.1200 |
| 5 | — | prostate | regional_lymph_node | 93 | 30 | — | — | — | — | — | — | **0.6318** | 0.4004 | 0.4955 | 0.0213 |
| 5 | — | stomach | regional_lymph_node | 77 | 30 | — | — | — | — | — | — | **0.8575** | 0.1919 | 0.3429 | 0.0933 |
| 5 | — | thyroid | regional_lymph_node | 47 | 30 | — | — | — | — | — | — | **0.6991** | 0.2750 | 0.4899 | 0.0000 |

**Supplementary Table S36. TCGA multi-run cascade headline (Stages A, B, C).**

Cascade-stage headline for the local-LLM (gpt-oss-20b) pipeline on the external TCGA-Reports cohort (n = 242 reports, six organs, k = 22 multi-run seeds). Stage A (eligibility triage), Stage B (organ classification), Stage C (per-organ field extraction macro mean) are reported separately because their denominators differ. Backs §3.9 / paragraph 17 (the headline numbers in the first paragraph of §3.9).

| **Stage** | **Stage name** | **n_total** | **n_correct** | **Accuracy [95 % CI]** | **Cohen's κ** | **Note** |
| --- | --- | --- | --- | --- | --- | --- |
| A | Eligibility (multi-run, run × case) | 5,324 | 4,828 | **90.68 % [89.87, 91.44]** | 0.0001 | TCGA gold is heavily positive-skewed (5,302 vs 22); κ near 0 reflects class imbalance, not classifier failure. |
| B | Organ classification (multi-run, run × case) | 4,828 | 4,770 | **98.80 % [98.45, 99.07]** | 0.9850 | macro-F1 = 0.850. |
| C | Field extraction (per-organ macro mean) | 96,461 | 77,437 | **79.92 %** | — | Unique cases: 242 cases at A → 241 → 239 at C; 22 multi-run seeds aggregated. |

**Supplementary Table S37. TCGA Stage-C field extraction — per-organ accuracy.**

Per-organ Stage-C effective accuracy on the external TCGA-Reports cohort under the local-LLM (gpt-oss-20b) pipeline (k = 22 multi-run seeds, 242 reports across six organs). Counts are case × run × field cells. Sorted descending by effective accuracy. Backs §3.9 / paragraph 17 (Table 15, Figure 8) and Supplementary Figure S6.

| **Organ** | **n_attempted** | **n_correct** | **Effective accuracy [95 % Wilson CI]** |
| --- | --- | --- | --- |
| colorectal | 20,144 | 17,466 | **86.71 % [86.23, 87.17]** |
| esophagus | 15,070 | 12,649 | **83.93 % [83.34, 84.51]** |
| liver | 16,660 | 13,090 | **78.57 % [77.94, 79.19]** |
| stomach | 15,114 | 11,664 | **77.17 % [76.50, 77.84]** |
| thyroid | 380 | 291 | **76.58 % [72.07, 80.56]** |
| breast | 29,093 | 22,277 | **76.57 % [76.08, 77.05]** |
| **Macro mean** |  |  | **79.92 %** |

**Supplementary Table S38. TCGA Stage-C field extraction — per-field accuracy with structural-caveat explanations.**

Per-field effective accuracy of the local-LLM (gpt-oss-20b) pipeline on the external TCGA-Reports cohort across the 22-seed multi-run pass (51 fields total). Sorted ascending by effective accuracy. Backs §3.9 / paragraph 17 (Table 16, Figure 8) and Supplementary Figure S7. The 13 lowest-accuracy fields carry caveat explanations that decompose the §3.9 internal-vs-external accuracy gap into two failure modes (Mode 1 = LLM hallucination from silent source / null gold; Mode 2 = coding-convention mismatch).

| **Field** | **n_total** | **n_attempted** | **n_correct** | **Coverage** | **Effective accuracy [95 % Wilson CI]** | **Caveat explanation** |
| --- | --- | --- | --- | --- | --- | --- |
| biomarker_pms2 | 1,006 | 6 | 0 | 0.60 % | **0.00 % [0.00, 0.38]** | MMR IHC not performed at TCGA institutions; gold null; LLM hallucinates when it emits (PMS2) |
| biomarker_msh6 | 1,006 | 6 | 0 | 0.60 % | **0.00 % [0.00, 0.38]** | MMR IHC not performed at TCGA institutions; gold null; LLM hallucinates when it emits (MSH6) |
| biomarker_msh2 | 1,006 | 6 | 0 | 0.60 % | **0.00 % [0.00, 0.38]** | MMR IHC not performed at TCGA institutions; gold null; LLM hallucinates when it emits (MSH2) |
| biomarker_mlh1 | 1,006 | 6 | 0 | 0.60 % | **0.00 % [0.00, 0.38]** | MMR IHC not performed at TCGA institutions; gold null; LLM hallucinates when it emits (MLH1) |
| biomarker_ki67 | 982 | 812 | 28 | 82.69 % | **2.85 % [1.98, 4.09]** | TCGA breast reports do not document Ki-67; LLM emits a default value |
| biomarker_her2 | 982 | 921 | 136 | 93.79 % | **13.85 % [11.83, 16.15]** | older TCGA breast reports omit HER2 testing; LLM emits a default value |
| overall_stage | 1,000 | 1,000 | 145 | 100.00 % | **14.50 % [12.45, 16.82]** | AJCC edition usually unstated in TCGA; gold null; LLM emits a stage group regardless |
| stage_group | 2,788 | 2,788 | 585 | 100.00 % | **20.98 % [19.51, 22.53]** | AJCC edition usually unstated in TCGA; gold null; LLM emits a stage group regardless |
| extrathyroid_extension | 20 | 20 | 8 | 100.00 % | **40.00 % [21.88, 61.34]** | (n = 20; small denominator) |
| biomarker_er | 982 | 917 | 436 | 93.38 % | **44.40 % [41.32, 47.52]** | older TCGA breast reports omit ER; LLM emits a default value |
| biomarker_pr | 982 | 917 | 436 | 93.38 % | **44.40 % [41.32, 47.52]** | older TCGA breast reports omit PR; LLM emits a default value |
| pathologic_stage_group | 982 | 981 | 517 | 99.90 % | **52.65 % [49.52, 55.75]** | AJCC edition usually unstated in TCGA; gold null; LLM emits a stage group regardless |
| cancer_quadrant | 982 | 982 | 558 | 100.00 % | **56.82 % [53.70, 59.89]** | (no caveat) |
| anatomic_stage_group | 982 | 981 | 564 | 99.90 % | **57.43 % [54.32, 60.49]** | AJCC edition usually unstated in TCGA; gold null; LLM emits a stage group regardless |
| pm_category | 4,770 | 4,769 | 2,969 | 99.98 % | **62.24 % [60.86, 63.61]** | default-Mx rule asymmetry (LLMs sometimes emit pM0) |
| surgical_technique | 2,788 | 2,788 | 1,853 | 100.00 % | **66.46 % [64.69, 68.19]** | (no caveat) |
| tumor_invasion | 1,006 | 1,006 | 683 | 100.00 % | **67.89 % [64.94, 70.71]** | (no caveat) |
| dcis_present | 982 | 982 | 733 | 100.00 % | **74.64 % [71.83, 77.26]** | (no caveat) |
| tnm_descriptor | 4,770 | 4,769 | 3,615 | 99.98 % | **75.79 % [74.55, 76.98]** | (no caveat) |
| pn_category | 4,770 | 4,769 | 3,716 | 99.98 % | **77.90 % [76.70, 79.06]** | (no caveat) |
| pt_category | 4,770 | 4,769 | 3,876 | 99.98 % | **81.26 % [80.13, 82.34]** | (no caveat) |
| nuclear_grade | 982 | 982 | 820 | 100.00 % | **83.50 % [81.05, 85.69]** | (no caveat) |
| dcis_size | 982 | 982 | 828 | 100.00 % | **84.32 % [81.91, 86.46]** | (no caveat) |
| procedure | 4,770 | 4,770 | 4,022 | 100.00 % | **84.32 % [83.26, 85.32]** | (no caveat) |
| mitotic_rate | 982 | 982 | 830 | 100.00 % | **84.52 % [82.13, 86.65]** | (no caveat) |
| distant_metastasis | 4,770 | 4,770 | 4,063 | 100.00 % | **85.18 % [84.14, 86.16]** | (no caveat) |
| cancer_primary_site | 2,788 | 2,788 | 2,425 | 100.00 % | **86.98 % [85.68, 88.18]** | (no caveat) |
| tumor_extent | 2,762 | 2,762 | 2,406 | 100.00 % | **87.11 % [85.81, 88.31]** | (no caveat) |
| dcis_comedo_necrosis | 982 | 982 | 860 | 100.00 % | **87.58 % [85.37, 89.49]** | (no caveat) |
| vascular_invasion | 980 | 980 | 864 | 100.00 % | **88.16 % [85.99, 90.04]** | (no caveat) |
| cancer_clock | 982 | 982 | 873 | 100.00 % | **88.90 % [86.78, 90.72]** | (no caveat) |
| maximal_ln_size | 4,770 | 4,766 | 4,245 | 99.92 % | **88.99 % [88.07, 89.85]** | (no caveat) |
| extranodal_extension | 4,770 | 4,766 | 4,255 | 99.92 % | **89.20 % [88.29, 90.05]** | (no caveat) |
| type_of_polyp | 1,006 | 1,006 | 898 | 100.00 % | **89.26 % [87.20, 91.03]** | (no caveat) |
| signet_ring | 1,846 | 1,846 | 1,676 | 100.00 % | **90.79 % [89.39, 92.03]** | (no caveat) |
| tumor_site | 1,000 | 1,000 | 912 | 100.00 % | **91.20 % [89.28, 92.80]** | (no caveat) |
| extracellular_mucin | 1,846 | 1,846 | 1,689 | 100.00 % | **91.50 % [90.13, 92.68]** | (no caveat) |
| tubule_formation | 982 | 982 | 899 | 100.00 % | **91.55 % [89.64, 93.13]** | (no caveat) |
| total_score | 982 | 982 | 907 | 100.00 % | **92.36 % [90.53, 93.86]** | (no caveat) |
| lymphovascular_invasion | 3,790 | 3,790 | 3,528 | 100.00 % | **93.09 % [92.24, 93.85]** | (no caveat) |
| tumor_focality | 1,000 | 1,000 | 934 | 100.00 % | **93.40 % [91.69, 94.78]** | (no caveat) |
| histology | 4,770 | 4,770 | 4,501 | 100.00 % | **94.36 % [93.67, 94.98]** | (no caveat) |
| tumor_size | 1,982 | 1,982 | 1,904 | 100.00 % | **96.06 % [95.12, 96.84]** | (no caveat) |
| perineural_invasion | 4,770 | 4,770 | 4,625 | 100.00 % | **96.96 % [96.43, 97.41]** | (no caveat) |
| grade | 4,750 | 4,750 | 4,618 | 100.00 % | **97.22 % [96.71, 97.65]** | (no caveat) |
| dcis_grade | 982 | 982 | 955 | 100.00 % | **97.25 % [96.03, 98.10]** | (no caveat) |
| cancer_laterality | 982 | 982 | 976 | 100.00 % | **99.39 % [98.67, 99.72]** | (no caveat) |
| mitotic_activity | 20 | 20 | 20 | 100.00 % | **100.00 % [83.87, 100.00]** | (n = 20; small denominator) |
| tumor_necrosis | 20 | 20 | 20 | 100.00 % | **100.00 % [83.87, 100.00]** | (n = 20; small denominator) |
| tumor_budding | 1,006 | 1,006 | 1,006 | 100.00 % | **100.00 % [99.62, 100.00]** | (no caveat) |
| predisposing_condition | 20 | 20 | 20 | 100.00 % | **100.00 % [83.87, 100.00]** | (n = 20; small denominator) |

**Supplementary Table S39. Baseline cascade comparison — four systems × three stages on the external TCGA cohort.**

Stage A / B / C cascade accuracy for four systems on the TCGA-Reports cohort (n = 242 reports across six organs). The four systems span the relevant architectural spectrum: deterministic regex (rule-based), encoder-only fine-tuned (BERT-merged), local LLM (gpt-oss-20b under DSPy with grammar-constrained decoding), and closed-frontier API LLM (gpt-5.4-mini under DSPy with native JSON-schema constrained decoding). Backs §3.10 / paragraph 18 (Table 12, Figure 8).

| **Stage** | **Stage name** | **System** | **Family** | **n** | **Accuracy [95 % Wilson CI]** | **Cohen's κ** |
| --- | --- | --- | --- | --- | --- | --- |
| A | Eligibility triage | Rule-based | rule | 242 | **96.69 % [93.61, 98.32]** | — |
| A | Eligibility triage | BERT-merged | encoder | 2,420 | **99.59 % [99.24, 99.78]** | — |
| A | Eligibility triage | Local LLM (gpt-oss-20b) | local_llm | 4,848 | **99.59 % [99.36, 99.73]** | — |
| A | Eligibility triage | API LLM (gpt-5.4-mini) | api_llm | 1,452 | **99.31 % [98.74, 99.63]** | — |
| B | Organ classification | Rule-based | rule | 234 | **96.58 % [93.40, 98.26]** | 0.957 |
| B | Organ classification | BERT-merged | encoder | 2,410 | **66.14 % [64.23, 68.00]** | 0.576 |
| B | Organ classification | Local LLM (gpt-oss-20b) | local_llm | 4,828 | **98.80 % [98.45, 99.07]** | 0.985 |
| B | Organ classification | API LLM (gpt-5.4-mini) | api_llm | 1,442 | **99.31 % [98.73, 99.62]** | 0.991 |
| C | Field extraction | Rule-based | rule | 1,283 | **65.16 % [62.51, 67.72]** | — |
| C | Field extraction | BERT-merged | encoder | 26,205 | **40.06 % [39.47, 40.65]** | — |
| C | Field extraction | Local LLM (gpt-oss-20b) | local_llm | 105,853 | **77.02 % [76.76, 77.27]** | — |
| C | Field extraction | API LLM (gpt-5.4-mini) | api_llm | 31,200 | **75.22 % [74.74, 75.70]** | — |

**Supplementary Table S40. Baseline cascade-attrition funnel by system.**

Per-system attrition through the three cascade gates on the TCGA-Reports cohort (n = 242 reports per system). Reports the proportion of cases that successfully pass each gate (Stage A → Stage B → Stage C). Backs §3.10 / paragraph 18 (cited at the end of paragraph 18 as the "Supplementary Table S40" footer). The Stage-C yield is the fraction of the 242-report cohort that produced a scoreable Stage-C field-extraction output.

| **System** | **Family** | **n_total** | **n_passed_A** | **n_passed_B** | **n_scored_C** | **Attrition A** | **Attrition B given A** | **Stage-C yield** |
| --- | --- | --- | --- | --- | --- | --- | --- | --- |
| Rule-based | rule | 242 | 234 | 226 | 226 | 3.31 % | 3.42 % | **93.39 %** |
| BERT-merged | encoder | 242 | 241 | 238 | 238 | 0.41 % | 1.24 % | **98.35 %** |
| Local LLM (gpt-oss-20b) | local_llm | 242 | 241 | 239 | 239 | 0.41 % | 0.83 % | **98.76 %** |
| API LLM (gpt-5.4-mini) | api_llm | 242 | 241 | 240 | 240 | 0.41 % | 0.41 % | **99.17 %** |

**Supplementary Table S41. Baseline Stage-C pairwise McNemar tests (Holm-corrected).**

Six pairwise McNemar tests on the four baseline systems at Stage C (per-organ field extraction) on the external TCGA-Reports cohort. Each test pairs the two systems on case × field cells where both attempted the same field; n_pairs varies because the systems cover different field-emission regimes. Backs §3.10 / paragraph 18 (Table 13). All six p-values are < 10⁻⁹; the headline Local-vs-API comparison gives a narrow Δ = −3.05 pp in favour of the local LLM.

| **Pair (a vs b)** | **n_pairs** | **Acc a** | **Acc b** | **Δ (b − a)** | **95 % CI** | **McNemar p** | **Verdict** |
| --- | --- | --- | --- | --- | --- | --- | --- |
| Rule-based vs BERT-merged | 1,082 | 0.6895 | 0.3005 | −0.3889 | [−0.4183, −0.3583] | 6.5 × 10⁻⁸⁴ | rule-based better |
| Rule-based vs Local LLM (gpt-oss-20b) | 1,283 | 0.6539 | 0.9042 | **+0.2503** | [+0.2263, +0.2750] | 3.0 × 10⁻⁶⁸ | Local LLM better |
| Rule-based vs API LLM (gpt-5.4-mini) | 1,282 | 0.6544 | 0.9028 | **+0.2484** | [+0.2223, +0.2733] | 1.6 × 10⁻⁶² | API LLM better |
| BERT-merged vs Local LLM (gpt-oss-20b) | 3,986 | 0.3799 | 0.8160 | **+0.4361** | [+0.4174, +0.4534] | 1.2 × 10⁻²⁸⁸ | Local LLM better |
| BERT-merged vs API LLM (gpt-5.4-mini) | 4,001 | 0.3795 | 0.7670 | **+0.3875** | [+0.3689, +0.4061] | 4.3 × 10⁻²³⁶ | API LLM better |
| Local LLM (gpt-oss-20b) vs API LLM (gpt-5.4-mini) | 5,262 | 0.7976 | 0.7671 | **−0.0305** | [−0.0386, −0.0222] | 1.5 × 10⁻⁹ | Local LLM better (slight) |

**Supplementary Table S42. Baseline Stage-C accuracy per organ × system on TCGA.**

Per-organ Stage-C accuracy across the four baseline systems on the TCGA-Reports cohort (six organs). Backs §3.10 / paragraph 18 (Table 14). Sorted by organ; values are effective accuracy [95 % Wilson CI].

| **Organ** | **Rule-based** | **BERT-merged** | **Local LLM (gpt-oss-20b)** | **API LLM (gpt-5.4-mini)** |
| --- | --- | --- | --- | --- |
| breast | **56.4 % [51.4, 61.2]** (n=392) | **37.8 % [36.6, 38.9]** (6,779) | **76.6 % [76.1, 77.1]** (29,093) | **76.0 % [75.1, 76.9]** (8,093) |
| colorectal | **63.1 % [58.2, 67.8]** (n=385) | **44.7 % [43.5, 45.9]** (6,537) | **86.7 % [86.2, 87.2]** (20,144) | **82.4 % [81.4, 83.3]** (6,000) |
| esophagus | **78.7 % [73.0, 83.4]** (n=239) | **34.2 % [32.9, 35.5]** (4,920) | **83.9 % [83.3, 84.5]** (15,070) | **81.1 % [79.9, 82.2]** (4,625) |
| liver | **75.6 % [68.3, 81.7]** (n=156) | **49.1 % [47.7, 50.5]** (5,045) | **78.6 % [77.9, 79.2]** (16,660) | **75.2 % [74.0, 76.3]** (4,998) |
| stomach | **62.5 % [52.5, 71.5]** (n=96) | **29.4 % [27.8, 31.1]** (2,924) | **77.2 % [76.5, 77.8]** (15,114) | **73.4 % [72.1, 74.7]** (4,536) |
| thyroid | **50.0 % [18.8, 81.2]** (n=6) | — (no cases scored) | **76.6 % [72.1, 80.6]** (380) | **71.1 % [62.1, 78.6]** (114) |

**Supplementary Table S43. Variable-length list-field F1 (surgical margins, regional lymph-node groups) across baseline systems on TCGA.**

List-typed extraction F1 for the two list-typed schema fields (margins[*], regional_lymph_node[*]) across the four baseline systems on TCGA-Reports (n = 226 to 1,594 cases per system, depending on field-emission regime). Backs §3.10 / paragraph 18.

**Scalar-vs-list divergence — read with Supplementary Table S39.** On per-field scalar extraction (Supplementary Table S39) our local language model (gpt-oss-20b) was ahead of the commercial gpt-5.4-mini API (77.02 % vs 75.22 %). On the two nested list fields here, the direction reverses — the commercial API led on both margins (F1 0.831 vs 0.783) and regional lymph-node groups (F1 0.686 vs 0.587). **The local-vs-commercial benchmark was not configured for a balanced head-to-head**: the local model was served with num_ctx = 16,384 while the commercial API was called with num_ctx = 4,096 (a 4× asymmetry favouring the local model on long-context inputs), and gpt-5.4-mini is the smaller, faster sibling of a frontier commercial family rather than the full variant. We therefore do not draw a categorical accuracy ordering between local and commercial language models from these numbers. Three discriminating experiments — a matched-context rerun of the commercial API at num_ctx = 16,384, a comparison against the full gpt-5.4, and an open-weights comparison against gpt-oss-120b — are flagged as future work (the GPU budget and time available in this revision did not extend to running them); §4 Discussion (¶25) frames the local-vs-commercial result as a promising follow-up question rather than a categorical accuracy claim.

| **Task** | **System** | **Family** | **n_total** | **n_attempted** | **Coverage** | **Mean F1 [95 % CI]** |
| --- | --- | --- | --- | --- | --- | --- |
| margins | Rule-based | rule | 226 | 4 | 1.77 % | **0.839 [0.625, 1.000]** |
| regional_lymph_node | Rule-based | rule | 226 | 4 | 1.77 % | **0.417 [0.000, 0.833]** |
| margins | BERT-merged | encoder | 1,594 | 0 | 0.00 % | **— (no list emission)** |
| regional_lymph_node | BERT-merged | encoder | 1,594 | 0 | 0.00 % | **— (no list emission)** |
| margins | Local LLM (gpt-oss-20b) | local_llm | 4,770 | 4,491 | 94.15 % | **0.783 [0.776, 0.790]** |
| regional_lymph_node | Local LLM (gpt-oss-20b) | local_llm | 4,770 | 3,967 | 83.17 % | **0.587 [0.572, 0.601]** |
| margins | API LLM (gpt-5.4-mini) | api_llm | 1,432 | 1,420 | 99.16 % | **0.831 [0.820, 0.843]** |
| regional_lymph_node | API LLM (gpt-5.4-mini) | api_llm | 1,432 | 1,216 | 84.92 % | **0.686 [0.661, 0.709]** |

**Supplementary Table S44. Component-ablation 5-method headline.**

Five-cell component-ablation headline on the CMUH validation cohort under gpt-oss-20b. The five cells span the proposed pipeline (dspy_modular, 10 multi-seed runs) and four single-axis lesions (dspy_monolithic, dspy_monolithic_no_jsonize, raw_json, free_text_regex, 1 run each). Total n = 109,618 case × field cells for the modular cell; 10,471–10,953 cells for the four lesion cells (single-run smaller denominator). Backs §3.6 / paragraph 14 (Table 17, Figure 6) and rebuttal R1c.

| **Method** | **n_attempted** | **n_correct** | **Effective accuracy [95 % Wilson CI]** | **Δ vs modular** | **McNemar p (Holm-corrected)** |
| --- | --- | --- | --- | --- | --- |
| **dspy_modular** (proposed) | 109,546 | 104,678 | **95.49 % [95.37, 95.61]** | — | — |
| raw_json | 10,899 | 10,166 | **93.07 % [92.58, 93.53]** | −2.42 pp | 1.5 × 10⁻¹⁹ |
| dspy_monolithic_no_jsonize | 10,526 | 9,998 | **91.28 % [90.74, 91.79]** | −4.21 pp | 0.0143 |
| dspy_monolithic | 10,467 | 9,925 | **90.71 % [90.15, 91.24]** | −4.79 pp | 0.0027 |
| free_text_regex | 9,690 | 1,601 | **15.29 % [14.61, 15.99]** | −80.20 pp | < 10⁻³⁰⁰ |

**Supplementary Table S45. Component-ablation per-method × per-field accuracy with McNemar Δ.**

Per-(method, field) accuracy across the five ablation cells on the CMUH validation cohort. 310 rows = 5 methods × ~62 fields. For each field, the modular cell is the reference; the other four cells report Δ vs modular with paired-bootstrap CIs and McNemar p (note the per-row p is the raw Bonferroni-pre-corrected value; the Holm-corrected aggregate is in Supplementary Table S50).

Because the full 310-row table is unwieldy in a single MD render, the table below renders **one section per method** (excerpt to ~10 representative fields each) plus the aggregate ALL row per method.

**dspy_modular_gpt_oss_20b — proposed cell (reference)**

| **Field** | **n** | **n_correct** | **n_attempted** | **Effective accuracy [95 % Wilson CI]** | **Δ vs modular** | **McNemar p** |
| --- | --- | --- | --- | --- | --- | --- |
| anatomic_stage_group | 739 | 724 | 739 | **97.97 % [96.68, 98.77]** | (reference) | — |
| pathologic_stage_group | 629 | 514 | 629 | **81.72 % [78.51, 84.54]** | (reference) | — |
| pm_category | 1,950 | 1,774 | 1,950 | **90.98 % [90.26, 91.65]** | (reference) | — |
| procedure | 8,720 | 8,214 | 8,720 | **94.20 % [93.61, 94.74]** | (reference) | — |
| histology | 4,748 | 4,701 | 4,748 | **99.01 % [98.74, 99.22]** | (reference) | — |
| tumor_necrosis | 320 | 203 | 320 | **63.44 % [58.03, 68.53]** | (reference) | — |
| cancer_quadrant | 640 | 464 | 640 | **72.50 % [68.91, 75.82]** | (reference) | — |
| surgical_technique | 3,863 | 3,293 | 3,863 | **85.24 % [84.09, 86.33]** | (reference) | — |
| ALL | 109,546 | 104,678 | 109,546 | **95.49 % [95.37, 95.61]** | (reference) | — |

**dspy_monolithic_gpt_oss_20b — single-organ-collapsed DSPy with the `ReportJsonize` pre-pass**

| **Field** | **n** | **n_correct** | **Effective accuracy [95 % CI]** | **Δ vs modular** | **McNemar p** |
| --- | --- | --- | --- | --- | --- |
| anatomic_stage_group | 74 | 72 | 97.30 % [90.85, 99.27] | −0.68 pp | 1.000 |
| pathologic_stage_group | 62 | 50 | 80.65 % [69.10, 88.59] | −1.07 pp | 1.000 |
| pm_category | 195 | 176 | 90.26 % [85.34, 93.65] | −0.72 pp | 1.000 |
| procedure | 871 | 815 | 93.57 % [91.78, 94.97] | −0.62 pp | 1.000 |
| tumor_necrosis | 32 | 21 | 65.62 % [48.34, 79.59] | +2.18 pp | 1.000 |
| ALL | 10,467 | 9,925 | **90.71 % [90.15, 91.24]** | **−4.79 pp** | **0.0027** |

**dspy_monolithic_no_jsonize_gpt_oss_20b — single-organ-collapsed DSPy without the `ReportJsonize` pre-pass**

| **Field** | **n** | **n_correct** | **Effective accuracy [95 % CI]** | **Δ vs modular** | **McNemar p** |
| --- | --- | --- | --- | --- | --- |
| anatomic_stage_group | 74 | 70 | 94.59 % [86.91, 97.88] | −3.39 pp | 0.250 |
| pathologic_stage_group | 63 | 60 | 95.24 % [86.91, 98.37] | +13.52 pp | 0.013 |
| pm_category | 195 | 155 | 79.49 % [73.34, 84.51] | −11.49 pp | 1.6 × 10⁻⁴ |
| procedure | 871 | 788 | 90.47 % [88.36, 92.22] | −3.72 pp | 0.001 |
| tumor_necrosis | 32 | 21 | 65.62 % [48.34, 79.59] | +2.18 pp | 1.000 |
| ALL | 10,526 | 9,998 | **91.28 % [90.74, 91.79]** | **−4.21 pp** | **0.0143** |

**raw_json_gpt_oss_20b — schema-constrained JSON without DSPy typing**

| **Field** | **n** | **n_correct** | **Effective accuracy [95 % CI]** | **Δ vs modular** | **McNemar p** |
| --- | --- | --- | --- | --- | --- |
| anatomic_stage_group | 74 | 70 | 94.59 % [86.91, 97.88] | −3.39 pp | 0.250 |
| pathologic_stage_group | 63 | 46 | 73.02 % [60.91, 82.42] | −8.70 pp | 0.057 |
| pm_category | 215 | 174 | 80.95 % [75.13, 85.69] | −10.04 pp | 5.5 × 10⁻⁸ |
| procedure | 871 | 798 | 91.62 % [89.59, 93.27] | −2.59 pp | 0.0006 |
| tumor_necrosis | 32 | 30 | 93.75 % [79.85, 98.27] | **+30.31 pp** | 0.002 |
| ALL | 10,899 | 10,166 | **93.07 % [92.58, 93.53]** | **−2.42 pp** | **1.5 × 10⁻¹⁹** |

**free_text_regex_gpt_oss_20b — schema-blind regex (the floor)**

| **Field** | **n** | **n_correct** | **Effective accuracy [95 % CI]** | **Δ vs modular** | **McNemar p** |
| --- | --- | --- | --- | --- | --- |
| anatomic_stage_group | 74 | 3 | 4.05 % [1.39, 11.27] | −93.92 pp | < 10⁻²⁰ |
| pathologic_stage_group | 63 | 5 | 7.94 % [3.43, 17.34] | −73.78 pp | < 10⁻¹⁵ |
| pm_category | 195 | 10 | 5.13 % [2.82, 9.18] | −85.85 pp | < 10⁻⁴² |
| procedure | 871 | 62 | 7.12 % [5.59, 9.04] | −87.08 pp | < 10⁻¹⁵⁰ |
| tumor_necrosis | 32 | 8 | 25.00 % [12.81, 43.43] | −38.44 pp | 0.012 |
| ALL | 9,690 | 1,601 | **15.29 % [14.61, 15.99]** | **−80.20 pp** | **< 10⁻³⁰⁰** |

**Supplementary Table S46. Component-ablation per-method × per-organ accuracy with Cochran's Q.**

Per-organ accuracy across the five ablation cells on the CMUH validation cohort. Five methods × ten organs = 50 rows. Each row reports n_attempted, n_correct, effective accuracy, 95 % Wilson CI, and the Cochran's Q heterogeneity statistic across organs (per method). Backs §3.6 / paragraph 14 (companion to Supplementary Table S44).

| **Method** | **Organ** | **n_attempted** | **n_correct** | **Effective accuracy [95 % Wilson CI]** | **Cochran's Q** | **Q-test p** |
| --- | --- | --- | --- | --- | --- | --- |
| dspy_modular_gpt_oss_20b | breast | 15,788 | 14,979 | **94.88 % [94.52, 95.21]** | — | — |
| dspy_modular_gpt_oss_20b | cervix | 3,268 | 3,088 | **94.49 % [93.66, 95.22]** | — | — |
| dspy_modular_gpt_oss_20b | colorectal | 11,730 | 11,458 | **97.68 % [97.39, 97.94]** | — | — |
| dspy_modular_gpt_oss_20b | esophagus | 10,187 | 9,848 | **96.67 % [96.31, 97.00]** | — | — |
| dspy_modular_gpt_oss_20b | liver | 9,870 | 9,336 | **94.59 % [94.13, 95.02]** | — | — |
| dspy_modular_gpt_oss_20b | lung | 9,280 | 8,830 | **95.15 % [94.69, 95.57]** | — | — |
| dspy_modular_gpt_oss_20b | pancreas | 3,840 | 3,592 | **93.54 % [92.72, 94.28]** | — | — |
| dspy_modular_gpt_oss_20b | prostate | 22,550 | 22,159 | **98.27 % [98.09, 98.43]** | — | — |
| dspy_modular_gpt_oss_20b | stomach | 12,515 | 11,771 | **94.06 % [93.63, 94.46]** | — | — |
| dspy_modular_gpt_oss_20b | thyroid | 10,590 | 9,617 | **90.81 % [90.25, 91.35]** | — | — |
| dspy_monolithic_gpt_oss_20b | breast | 1,580 | 1,347 | **85.25 % [83.42, 86.92]** | 0.419 | 0.99999 |
| dspy_monolithic_gpt_oss_20b | cervix | 318 | 282 | **88.68 % [84.73, 91.71]** | 0.419 | 0.99999 |
| dspy_monolithic_gpt_oss_20b | colorectal | 1,173 | 1,104 | **94.12 % [92.62, 95.33]** | 0.419 | 0.99999 |
| dspy_monolithic_gpt_oss_20b | esophagus | 1,003 | 927 | **92.42 % [90.62, 93.90]** | 0.419 | 0.99999 |
| dspy_monolithic_gpt_oss_20b | liver | 987 | 933 | **94.53 % [92.93, 95.78]** | 0.419 | 0.99999 |
| dspy_monolithic_gpt_oss_20b | lung | 928 | 774 | **83.41 % [80.87, 85.66]** | 0.419 | 0.99999 |
| dspy_monolithic_gpt_oss_20b | pancreas | 384 | 303 | **78.91 % [74.55, 82.69]** | 0.419 | 0.99999 |
| dspy_monolithic_gpt_oss_20b | prostate | 2,255 | 2,181 | **96.72 % [95.90, 97.38]** | 0.419 | 0.99999 |
| dspy_monolithic_gpt_oss_20b | stomach | 1,255 | 1,099 | **87.57 % [85.63, 89.28]** | 0.419 | 0.99999 |
| dspy_monolithic_gpt_oss_20b | thyroid | 1,059 | 975 | **92.07 % [90.28, 93.55]** | 0.419 | 0.99999 |
| dspy_monolithic_no_jsonize_gpt_oss_20b | breast | 1,580 | 1,411 | **89.30 % [87.68, 90.73]** | 0.342 | 0.99999 |
| dspy_monolithic_no_jsonize_gpt_oss_20b | cervix | 329 | 312 | **94.83 % [91.88, 96.75]** | 0.342 | 0.99999 |
| dspy_monolithic_no_jsonize_gpt_oss_20b | colorectal | 1,173 | 1,114 | **94.97 % [93.57, 96.08]** | 0.342 | 0.99999 |
| dspy_monolithic_no_jsonize_gpt_oss_20b | esophagus | 1,003 | 936 | **93.32 % [91.60, 94.71]** | 0.342 | 0.99999 |
| dspy_monolithic_no_jsonize_gpt_oss_20b | liver | 987 | 925 | **93.72 % [92.03, 95.07]** | 0.342 | 0.99999 |
| dspy_monolithic_no_jsonize_gpt_oss_20b | lung | 928 | 783 | **84.38 % [81.90, 86.57]** | 0.342 | 0.99999 |
| dspy_monolithic_no_jsonize_gpt_oss_20b | pancreas | 384 | 334 | **86.98 % [83.24, 89.98]** | 0.342 | 0.99999 |
| dspy_monolithic_no_jsonize_gpt_oss_20b | prostate | 2,255 | 2,116 | **93.84 % [92.77, 94.76]** | 0.342 | 0.99999 |
| dspy_monolithic_no_jsonize_gpt_oss_20b | stomach | 1,255 | 1,084 | **86.37 % [84.37, 88.16]** | 0.342 | 0.99999 |
| dspy_monolithic_no_jsonize_gpt_oss_20b | thyroid | 1,059 | 983 | **92.82 % [91.11, 94.23]** | 0.342 | 0.99999 |
| free_text_regex_gpt_oss_20b | breast | 1,559 | 238 | **15.27 % [13.57, 17.14]** | 0.069 | 0.99999 |
| free_text_regex_gpt_oss_20b | cervix | 318 | 26 | **8.18 % [5.64, 11.71]** | 0.069 | 0.99999 |
| free_text_regex_gpt_oss_20b | colorectal | 1,121 | 176 | **15.70 % [13.69, 17.95]** | 0.069 | 0.99999 |
| free_text_regex_gpt_oss_20b | esophagus | 858 | 209 | **24.36 % [21.61, 27.34]** | 0.069 | 0.99999 |
| free_text_regex_gpt_oss_20b | liver | 987 | 210 | **21.28 % [18.84, 23.94]** | 0.069 | 0.99999 |
| free_text_regex_gpt_oss_20b | lung | 928 | 210 | **22.63 % [20.05, 25.43]** | 0.069 | 0.99999 |
| free_text_regex_gpt_oss_20b | pancreas | 384 | 49 | **12.76 % [9.79, 16.47]** | 0.069 | 0.99999 |
| free_text_regex_gpt_oss_20b | prostate | 2,203 | 120 | **5.45 % [4.57, 6.47]** | 0.069 | 0.99999 |
| free_text_regex_gpt_oss_20b | stomach | 1,054 | 109 | **10.34 % [8.64, 12.33]** | 0.069 | 0.99999 |
| free_text_regex_gpt_oss_20b | thyroid | 1,059 | 254 | **23.98 % [21.51, 26.65]** | 0.069 | 0.99999 |
| raw_json_gpt_oss_20b | breast | 1,580 | 1,318 | **83.42 % [81.50, 85.17]** | 0.627 | 0.99992 |
| raw_json_gpt_oss_20b | cervix | 318 | 290 | **91.19 % [87.57, 93.84]** | 0.627 | 0.99992 |
| raw_json_gpt_oss_20b | colorectal | 1,173 | 1,133 | **96.59 % [95.39, 97.49]** | 0.627 | 0.99992 |
| raw_json_gpt_oss_20b | esophagus | 1,005 | 973 | **96.82 % [95.54, 97.74]** | 0.627 | 0.99992 |
| raw_json_gpt_oss_20b | liver | 987 | 929 | **94.12 % [92.48, 95.43]** | 0.627 | 0.99992 |
| raw_json_gpt_oss_20b | lung | 922 | 848 | **91.97 % [90.04, 93.56]** | 0.627 | 0.99992 |
| raw_json_gpt_oss_20b | pancreas | 384 | 357 | **92.97 % [89.96, 95.12]** | 0.627 | 0.99992 |
| raw_json_gpt_oss_20b | prostate | 2,255 | 2,186 | **96.94 % [96.15, 97.58]** | 0.627 | 0.99992 |
| raw_json_gpt_oss_20b | stomach | 1,240 | 1,138 | **91.77 % [90.11, 93.18]** | 0.627 | 0.99992 |
| raw_json_gpt_oss_20b | thyroid | 1,059 | 994 | **93.86 % [92.25, 95.16]** | 0.627 | 0.99992 |

**Supplementary Table S47. Component-ablation marginal means per (field × axis × level).**

Marginal accuracies per (field × axis × level), where each axis is a binary engineering choice (modular vs monolithic; jsonize vs no-jsonize; DSPy typing vs raw JSON; schema-constrained vs free-text-regex). 314 rows = ~62 fields × 5 axis levels per field. Used to compute the cross-method effect sizes in Supplementary Tables S44 and S46 after marginalising out the other axes.

The full 314-row long-form is at code/derived/ablation_marginal_means.csv and is the canonical numeric source. The summary below reports the four axis aggregates across the ALL field (the cross-axis contrast headline).

**Aggregate marginal means at the `ALL` (full-schema) level**

| **Axis** | **Level** | **n** | **Marginal accuracy [95 % Wilson CI]** |
| --- | --- | --- | --- |
| axis_A | modular | 109,546 | **95.49 % [95.37, 95.61]** |
| axis_A | non_modular (monolithic + raw_json) | 32,318 | **89.69 % [89.39, 89.97]** |
| axis_B | jsonize_on (modular + monolithic + raw_json) | 130,907 | **94.34 % [94.21, 94.46]** |
| axis_B | jsonize_off (monolithic_no_jsonize) | 10,526 | **94.98 % [94.55, 95.38]** |
| axis_C | dspy_typed (modular + monolithic + monolithic_no_jsonize) | 130,539 | **94.50 % [94.37, 94.62]** |
| axis_C | raw_json | 10,899 | **93.27 % [92.79, 93.73]** |
| axis_D | schema_constrained (all four except free_text_regex) | 141,438 | **94.40 % [94.27, 94.51]** |
| axis_D | free_text_regex | 9,690 | **16.52 % [15.80, 17.27]** |

**Supplementary Table S48. Component-ablation factorial GLMM estimates per (field × term).**

Per-(field × term) factorial estimates on the logit scale. 250 rows = ~62 fields × 4 terms (intercept, axis_A_modular, axis_B_jsonize, axis_C_dspy_typed). The model is a logistic regression on the binary correctness outcome with the four ablation axes encoded as orthogonal contrasts. Each row reports the field, the term name, the estimate (in logit), the standard error, and the z-statistic.

| **Field** | **Term** | **Estimate (logit)** | **SE (logit)** | **z** |
| --- | --- | --- | --- | --- |
| anatomic_stage_group | (Intercept) | +1.821 | 0.285 | +6.379 |
| anatomic_stage_group | axis_A_modular | +0.902 | 0.412 | +2.190 |
| anatomic_stage_group | axis_B_jsonize | −0.214 | 0.391 | −0.547 |
| anatomic_stage_group | axis_C_dspy_typed | +0.486 | 0.385 | +1.262 |
| pathologic_stage_group | (Intercept) | +1.503 | 0.198 | +7.587 |
| pathologic_stage_group | axis_A_modular | −0.342 | 0.265 | −1.291 |
| pathologic_stage_group | axis_B_jsonize | +0.917 | 0.281 | +3.262 |
| pathologic_stage_group | axis_C_dspy_typed | +0.214 | 0.258 | +0.829 |
| pm_category | (Intercept) | +2.293 | 0.097 | +23.561 |
| pm_category | axis_A_modular | +1.051 | 0.143 | +7.330 |
| pm_category | axis_B_jsonize | −0.067 | 0.135 | −0.495 |
| pm_category | axis_C_dspy_typed | +0.491 | 0.131 | +3.738 |
| procedure | (Intercept) | +2.421 | 0.062 | +38.844 |
| procedure | axis_A_modular | +0.396 | 0.090 | +4.376 |
| procedure | axis_B_jsonize | −0.014 | 0.084 | −0.167 |
| procedure | axis_C_dspy_typed | +0.187 | 0.082 | +2.282 |
| histology | (Intercept) | +4.118 | 0.131 | +31.435 |
| histology | axis_A_modular | +0.572 | 0.182 | +3.149 |
| histology | axis_B_jsonize | −0.034 | 0.171 | −0.199 |
| histology | axis_C_dspy_typed | +0.213 | 0.164 | +1.298 |
| tumor_necrosis | (Intercept) | −0.193 | 0.354 | −0.546 |
| tumor_necrosis | axis_A_modular | −0.587 | 0.519 | −1.131 |
| tumor_necrosis | axis_B_jsonize | +0.452 | 0.508 | +0.890 |
| tumor_necrosis | axis_C_dspy_typed | −1.142 | 0.480 | −2.379 |
| cancer_quadrant | (Intercept) | +0.967 | 0.085 | +11.353 |
| cancer_quadrant | axis_A_modular | +0.014 | 0.117 | +0.121 |
| cancer_quadrant | axis_B_jsonize | −0.078 | 0.110 | −0.711 |
| cancer_quadrant | axis_C_dspy_typed | −0.021 | 0.106 | −0.198 |
| surgical_technique | (Intercept) | +1.736 | 0.041 | +42.336 |
| surgical_technique | axis_A_modular | +0.128 | 0.058 | +2.207 |
| surgical_technique | axis_B_jsonize | −0.045 | 0.054 | −0.838 |
| surgical_technique | axis_C_dspy_typed | +0.096 | 0.052 | +1.842 |
| tnm_descriptor | (Intercept) | +1.683 | 0.046 | +36.797 |
| tnm_descriptor | axis_A_modular | +0.245 | 0.066 | +3.738 |
| tnm_descriptor | axis_B_jsonize | +0.014 | 0.061 | +0.230 |
| tnm_descriptor | axis_C_dspy_typed | +0.087 | 0.059 | +1.479 |
| tumor_extension | (Intercept) | +1.040 | 0.143 | +7.290 |
| tumor_extension | axis_A_modular | −0.204 | 0.198 | −1.030 |
| tumor_extension | axis_B_jsonize | +0.117 | 0.190 | +0.617 |
| tumor_extension | axis_C_dspy_typed | −0.053 | 0.183 | −0.290 |

**Supplementary Table S49. Ablation GLMM with case- and run-level variance decomposition.**

Per-(field) generalised linear mixed-effects model estimates for the modular ablation cell, with two-source bootstrap CIs that decompose total uncertainty into case-level heterogeneity (variation across the 893 cases) vs run-level noise (variation across the 10 multi-seeds). 71 fields; the cell column is dspy_modular for all rows. Used to attribute residual variance to the underlying source (case-level heterogeneity vs run-to-run model stochasticity). Backs §3.6 / paragraph 14 (the variance-decomposition discussion). For brevity below the table renders 12 representative rows spanning the full range of point estimates (high accuracy, mid accuracy, low accuracy)

| **Field** | **Field kind** | **n_cases** | **n_runs** | **Point estimate** | **Mean per-run accuracy** | **Case CI [lo, hi]** | **Run CI [lo, hi]** | **Total CI (two-source bootstrap) [lo, hi]** | **var_case** | **var_run** |
| --- | --- | --- | --- | --- | --- | --- | --- | --- | --- | --- |
| anatomic_stage_group | scalar | 75 | 10 | 0.9800 | 0.9800 | [0.9653, 0.9866] | [0.9707, 0.9892] | [0.9478, 1.0000] | 0.9479 | 1.0000 |
| biomarker_er | scalar | 75 | 10 | 1.0000 | 1.0000 | [1.0000, 1.0000] | [1.0000, 1.0000] | [0.9949, 1.0000] | 0.9949 | 1.0000 |
| biomarker_her2 | scalar | 75 | 10 | 1.0000 | 1.0000 | [1.0000, 1.0000] | [1.0000, 1.0000] | [0.9949, 1.0000] | 0.9949 | 1.0000 |
| biomarker_ki67 | scalar | 75 | 10 | 0.9947 | 0.9946 | [0.9853, 0.9973] | [0.9897, 0.9996] | [0.9840, 1.0000] | 0.9840 | 1.0000 |
| biomarker_mlh1 | scalar | 67 | 10 | 1.0000 | 1.0000 | [1.0000, 1.0000] | [1.0000, 1.0000] | [0.9943, 1.0000] | 0.9943 | 1.0000 |
| pathologic_stage_group | scalar | 75 | 10 | 0.8171 | 0.8171 | [0.7741, 0.8454] | [0.7902, 0.8423] | [0.6747, 0.9325] | 0.6747 | 0.9325 |
| tumor_necrosis | scalar | 32 | 10 | 0.6344 | 0.6344 | [0.5278, 0.7333] | [0.5849, 0.6797] | [0.5125, 0.7406] | 0.5125 | 0.7406 |
| pm_category | scalar | 1,950 | 10 | 0.9098 | 0.9098 | [0.9024, 0.9166] | [0.9067, 0.9128] | [0.9006, 0.9192] | 0.9006 | 0.9192 |
| tumor_invasion | scalar | 70 | 10 | 0.8414 | 0.8414 | [0.8000, 0.8714] | [0.8147, 0.8682] | [0.7857, 0.9000] | 0.7857 | 0.9000 |
| tnm_descriptor | scalar | 160 | 10 | 0.8421 | 0.8421 | [0.8233, 0.8584] | [0.8331, 0.8511] | [0.8155, 0.8688] | 0.8155 | 0.8688 |
| tumor_size | scalar | 379 | 10 | 0.9416 | 0.9416 | [0.9335, 0.9485] | [0.9359, 0.9474] | [0.9213, 0.9623] | 0.9213 | 0.9623 |
| visceral_pleural_invasion | scalar | 63 | 10 | 0.9921 | 0.9921 | [0.9810, 0.9968] | [0.9840, 1.0000] | [0.9778, 1.0000] | 0.9778 | 1.0000 |

**Supplementary Table S50. Modularity advantage per field — modular cell vs best-performing alternative.**

For every schema field, the modular cell's accuracy is compared against the best-performing alternative ablation cell on that field. The "modularity advantage" is the modular accuracy minus the best alternative's accuracy, with paired-bootstrap CIs and a Holm-corrected p-value. 63 fields plus the ALL aggregate row. Backs §3.6 / paragraph 14 (Figure S1) and rebuttal R1c.

Sorted ascending by modular_advantage so the largest negative-modular and largest positive-modular cells are at the extremes.

| **Field** | **Modular accuracy [95 % CI]** | **Best alternative method** | **Best alternative accuracy [95 % CI]** | **Modular advantage [95 % CI]** | **Holm p** |
| --- | --- | --- | --- | --- | --- |
| pathologic_stage_group | 0.817 [0.785, 0.845] | dspy_monolithic_no_jsonize | 0.952 [0.869, 0.984] | **−0.159** [−0.270, −0.048] | 0.7505 |
| tumor_necrosis | 0.634 [0.580, 0.685] | raw_json | 0.938 [0.799, 0.983] | **−0.313** [−0.469, −0.156] | 0.1152 |
| perineural_invasion | 0.935 [0.929, 0.942] | raw_json | 0.977 [0.961, 0.987] | **−0.040** [−0.059, −0.023] | 0.0072 |
| distant_metastasis | 0.930 [0.923, 0.936] | raw_json | 0.955 [0.935, 0.969] | **−0.025** [−0.047, −0.002] | 1.0 |
| lymphovascular_invasion | 0.966 [0.961, 0.971] | raw_json | 0.987 [0.973, 0.994] | **−0.024** [−0.043, −0.006] | 1.0 |
| extranodal_extension | 0.948 [0.936, 0.957] | raw_json | 0.961 [0.922, 0.981] | **−0.011** [−0.050, +0.028] | 1.0 |
| intraductal_carcinoma_presence | 0.994 [0.986, 0.997] | dspy_monolithic | 1.000 [0.961, 1.000] | −0.011 [−0.032, 0.000] | 1.0 |
| bladder_invasion | 0.990 [0.982, 0.995] | raw_json | 1.000 [0.961, 1.000] | −0.011 [−0.032, 0.000] | 1.0 |
| gleason_5_percentage | 0.983 [0.973, 0.990] | raw_json | 0.989 [0.940, 0.998] | −0.011 [−0.056, +0.022] | 1.0 |
| tumor_size | 0.943 [0.936, 0.950] | raw_json | 0.939 [0.909, 0.959] | −0.008 [−0.029, +0.011] | 1.0 |
| surgical_technique | 0.852 [0.841, 0.863] | dspy_monolithic_no_jsonize | 0.850 [0.811, 0.882] | −0.008 [−0.047, +0.031] | 1.0 |
| maximal_ln_size | 0.976 [0.967, 0.982] | raw_json | 0.971 [0.934, 0.988] | −0.006 [−0.041, +0.029] | 1.0 |
| tumor_invasion | 0.841 [0.813, 0.867] | dspy_monolithic_no_jsonize | 0.871 [0.773, 0.931] | −0.014 [−0.086, +0.057] | 1.0 |
| tumor_extension | 0.739 [0.685, 0.787] | dspy_monolithic_no_jsonize | 0.786 [0.605, 0.898] | −0.036 [−0.250, +0.179] | 1.0 |
| cancer_quadrant | 0.725 [0.689, 0.758] | dspy_monolithic_no_jsonize | 0.719 [0.599, 0.814] | −0.016 [−0.125, +0.078] | 1.0 |
| cribriform_pattern_presence | 0.994 [0.986, 0.997] | dspy_monolithic | 1.000 [0.955, 1.000] | 0.000 [0.000, 0.000] | 1.0 |
| extracellular_mucin | 1.000 [0.997, 1.000] | raw_json | 1.000 [0.973, 1.000] | 0.000 [0.000, 0.000] | 1.0 |
| extraprostatic_extension | 1.000 [0.996, 1.000] | dspy_monolithic | 1.000 [0.961, 1.000] | 0.000 [0.000, 0.000] | 1.0 |
| extrathyroid_extension | 0.978 [0.944, 0.991] | dspy_monolithic_no_jsonize | 1.000 [0.824, 1.000] | 0.000 [0.000, 0.000] | 1.0 |
| margin_length | 0.990 [0.975, 0.996] | dspy_monolithic | 1.000 [0.914, 1.000] | 0.000 [0.000, 0.000] | 1.0 |
| margin_positivity | 0.996 [0.989, 0.998] | raw_json | 1.000 [0.961, 1.000] | 0.000 [0.000, 0.000] | 1.0 |
| overall_stage | 0.978 [0.970, 0.984] | raw_json | 0.972 [0.936, 0.988] | 0.000 [−0.034, +0.034] | 1.0 |
| prostate_size | 0.993 [0.985, 0.996] | dspy_monolithic | 0.989 [0.942, 0.998] | 0.000 [−0.032, +0.032] | 1.0 |
| prostate_weight | 1.000 [0.996, 1.000] | dspy_monolithic | 1.000 [0.960, 1.000] | 0.000 [0.000, 0.000] | 1.0 |
| seminal_vesicle_invasion | 1.000 [0.996, 1.000] | dspy_monolithic | 1.000 [0.961, 1.000] | 0.000 [0.000, 0.000] | 1.0 |
| signet_ring | 1.000 [0.997, 1.000] | raw_json | 1.000 [0.974, 1.000] | 0.000 [0.000, 0.000] | 1.0 |
| spread_through_air_spaces_stas | 0.980 [0.964, 0.989] | raw_json | 1.000 [0.930, 1.000] | 0.000 [0.000, 0.000] | 1.0 |
| tubule_formation | 1.000 [0.995, 1.000] | dspy_monolithic_no_jsonize | 0.959 [0.886, 0.986] | +0.041 [0.000, +0.096] | 1.0 |
| visceral_pleural_invasion | 0.991 [0.978, 0.996] | raw_json | 1.000 [0.932, 1.000] | 0.000 [0.000, 0.000] | 1.0 |
| tumor_budding | 1.000 [0.994, 1.000] | dspy_monolithic | 1.000 [0.946, 1.000] | 0.000 [0.000, 0.000] | 1.0 |
| type_of_polyp | 0.991 [0.969, 0.998] | raw_json | 1.000 [0.857, 1.000] | 0.000 [0.000, 0.000] | 1.0 |
| depth_of_invasion_number | 1.000 [0.986, 1.000] | dspy_monolithic_no_jsonize | 1.000 [0.875, 1.000] | 0.000 [0.000, 0.000] | 1.0 |
| depth_of_invasion_three_tier | 0.952 [0.915, 0.974] | raw_json | 0.952 [0.773, 0.992] | 0.000 [−0.143, +0.143] | 1.0 |
| direct_invasion_of_adjacent_structures | 0.967 [0.947, 0.979] | raw_json | 0.958 [0.860, 0.988] | +0.021 [−0.042, +0.104] | 1.0 |
| pn_category | 0.963 [0.958, 0.967] | raw_json | 0.949 [0.929, 0.963] | +0.011 [−0.009, +0.029] | 1.0 |
| cancer_clock | 0.915 [0.884, 0.938] | dspy_monolithic | 0.854 [0.716, 0.931] | 0.000 [−0.098, +0.098] | 1.0 |
| vascular_invasion | 0.921 [0.874, 0.952] | dspy_monolithic_no_jsonize | 0.947 [0.754, 0.991] | 0.000 [−0.158, +0.158] | 1.0 |
| tumor_size | 0.943 [0.936, 0.950] | raw_json | 0.939 [0.909, 0.959] | −0.008 [−0.029, +0.011] | 1.0 |
| total_score | 1.000 [0.995, 1.000] | dspy_monolithic_no_jsonize | 0.945 [0.867, 0.978] | +0.055 [+0.014, +0.110] | 1.0 |
| nuclear_grade | 1.000 [0.995, 1.000] | dspy_monolithic_no_jsonize | 0.959 [0.886, 0.986] | +0.041 [0.000, +0.096] | 1.0 |
| mitotic_rate | 1.000 [0.995, 1.000] | dspy_monolithic_no_jsonize | 0.959 [0.886, 0.986] | +0.041 [0.000, +0.096] | 1.0 |
| mitotic_activity | 0.969 [0.953, 0.980] | dspy_monolithic_no_jsonize | 0.968 [0.890, 0.991] | +0.016 [−0.032, +0.081] | 1.0 |
| dcis_size | 1.000 [0.992, 1.000] | dspy_monolithic_no_jsonize | 0.960 [0.865, 0.989] | +0.040 [0.000, +0.100] | 1.0 |
| dcis_present | 0.992 [0.982, 0.996] | dspy_monolithic_no_jsonize | 0.946 [0.869, 0.979] | +0.041 [0.000, +0.095] | 1.0 |
| dcis_grade | 0.986 [0.972, 0.993] | dspy_monolithic_no_jsonize | 0.961 [0.868, 0.989] | +0.020 [−0.039, +0.098] | 1.0 |
| dcis_comedo_necrosis | 0.973 [0.954, 0.984] | dspy_monolithic | 0.882 [0.766, 0.945] | **+0.078** [0.000, +0.176] | 1.0 |
| anatomic_stage_group | 0.980 [0.967, 0.988] | dspy_monolithic_no_jsonize | 0.946 [0.869, 0.979] | +0.041 [0.000, +0.095] | 1.0 |
| cancer_laterality | 0.991 [0.981, 0.995] | free_text_regex | 0.973 [0.907, 0.993] | +0.027 [0.000, +0.068] | 1.0 |
| sideness | 0.998 [0.991, 1.000] | free_text_regex | 0.984 [0.915, 0.997] | +0.016 [0.000, +0.048] | 1.0 |
| tumor_focality | 0.979 [0.972, 0.984] | dspy_monolithic_no_jsonize | 0.954 [0.917, 0.975] | **+0.028** [+0.005, +0.056] | 1.0 |
| tumor_site | 0.947 [0.936, 0.957] | dspy_monolithic_no_jsonize | 0.944 [0.900, 0.969] | +0.017 [−0.011, +0.045] | 1.0 |
| pt_category | 0.998 [0.997, 0.999] | raw_json | 0.992 [0.982, 0.997] | +0.006 [+0.002, +0.012] | 1.0 |
| stage_group | 0.958 [0.951, 0.964] | raw_json | 0.928 [0.897, 0.949] | **+0.026** [+0.003, +0.052] | 1.0 |
| tumor_extent | 0.976 [0.967, 0.983] | raw_json | 0.935 [0.885, 0.964] | **+0.045** [+0.006, +0.084] | 1.0 |
| histology | 0.990 [0.987, 0.992] | raw_json | 0.979 [0.965, 0.987] | **+0.015** [+0.005, +0.026] | 0.7505 |
| grade | 0.999 [0.998, 0.999] | raw_json | 0.991 [0.979, 0.996] | +0.009 [+0.002, +0.018] | 1.0 |
| gleason_4_percentage | 0.971 [0.958, 0.980] | dspy_monolithic | 0.956 [0.892, 0.983] | +0.011 [0.000, +0.033] | 1.0 |
| cancer_primary_site | 0.918 [0.907, 0.927] | raw_json | 0.928 [0.892, 0.953] | +0.004 [−0.032, +0.036] | 1.0 |
| tumor_percentage | 0.974 [0.962, 0.983] | dspy_monolithic | 0.957 [0.895, 0.983] | +0.011 [−0.022, +0.054] | 1.0 |
| procedure | 0.942 [0.936, 0.947] | raw_json | 0.916 [0.893, 0.935] | **+0.035** [+0.017, +0.053] | 0.0354 |
| pm_category | 0.910 [0.903, 0.917] | raw_json | 0.810 [0.778, 0.838] | **+0.096** [+0.062, +0.128] | 3.4 × 10⁻⁶ |
| involved_margin_list | 0.863 [0.827, 0.893] | dspy_monolithic | 0.829 [0.687, 0.915] | +0.024 [−0.073, +0.123] | 1.0 |
| tnm_descriptor | 0.842 [0.823, 0.859] | dspy_monolithic_no_jsonize | 0.806 [0.738, 0.860] | +0.037 [−0.012, +0.081] | 1.0 |
| **ALL (aggregate)** | **0.955 [0.954, 0.956]** | raw_json | **0.931 [0.926, 0.935]** | **+0.024 [+0.019, +0.029]** | **3.5 × 10⁻¹⁹** |

**Supplementary Table S51. Ablation modular-cell seed consistency (Fleiss κ across 10 seeds).**

For every schema field in the modular cell (dspy_modular_gpt_oss_20b), the per-seed accuracy mean / SD / min / max across the 10 multi-seed runs of the modular cell, plus Fleiss κ across the 10 raters (seeds). Used to bound the seed-to-seed variability that the single-run estimates on the four alternative ablation cells conceal. 62 fields. Backs §3.6 / paragraph 14 (Figure S2); the seed-count rationale and the within-axis matched-pair logic this table feeds are documented in Supplementary §S1.3.1.3–§S1.3.1.4.

| **Field** | **n_seeds** | **Accuracy mean** | **Accuracy SD** | **Accuracy min** | **Accuracy max** | **Fleiss κ across seeds** |
| --- | --- | --- | --- | --- | --- | --- |
| anatomic_stage_group | 10 | 0.9797 | 0.0124 | 0.9459 | 0.9865 | 0.6116 |
| bladder_invasion | 10 | 0.9904 | 0.0074 | 0.9787 | 1.0000 | 0.4391 |
| cancer_clock | 10 | 0.9146 | 0.0273 | 0.8537 | 0.9512 | 0.6078 |
| cancer_laterality | 10 | 0.9907 | 0.0085 | 0.9733 | 1.0000 | 0.4712 |
| cancer_primary_site | 10 | 0.9177 | 0.0122 | 0.8968 | 0.9364 | 0.6160 |
| cancer_quadrant | 10 | 0.7250 | 0.0281 | 0.6719 | 0.7656 | 0.5733 |
| cribriform_pattern_presence | 10 | 0.9938 | 0.0062 | 0.9877 | 1.0000 | 0.4410 |
| dcis_comedo_necrosis | 10 | 0.9725 | 0.0180 | 0.9412 | 1.0000 | 0.4614 |
| dcis_grade | 10 | 0.9863 | 0.0090 | 0.9804 | 1.0000 | 0.2114 |
| dcis_present | 10 | 0.9919 | 0.0066 | 0.9865 | 1.0000 | 0.4406 |
| dcis_size | 10 | 1.0000 | 0.0000 | 1.0000 | 1.0000 | — (constant) |
| depth_of_invasion_number | 10 | 1.0000 | 0.0000 | 1.0000 | 1.0000 | — (constant) |
| depth_of_invasion_three_tier | 10 | 0.9524 | 0.0301 | 0.9048 | 1.0000 | 0.2067 |
| direct_invasion_of_adjacent_structures | 10 | 0.9667 | 0.0191 | 0.9375 | 1.0000 | 0.1954 |
| distant_metastasis | 10 | 0.9296 | 0.0051 | 0.9189 | 0.9371 | 0.4428 |
| extracellular_mucin | 10 | 1.0000 | 0.0000 | 1.0000 | 1.0000 | — (constant) |
| extranodal_extension | 10 | 0.9476 | 0.0090 | 0.9333 | 0.9663 | 0.7054 |
| extraprostatic_extension | 10 | 1.0000 | 0.0000 | 1.0000 | 1.0000 | — (constant) |
| extrathyroid_extension | 10 | 0.9778 | 0.0272 | 0.9444 | 1.0000 | 0.0341 |
| gleason_4_percentage | 10 | 0.9714 | 0.0088 | 0.9560 | 0.9780 | 0.7976 |
| gleason_5_percentage | 10 | 0.9833 | 0.0124 | 0.9556 | 1.0000 | 0.4350 |
| grade | 10 | 0.9989 | 0.0015 | 0.9964 | 1.0000 | 0.0730 |
| histology | 10 | 0.9901 | 0.0024 | 0.9864 | 0.9940 | 0.4280 |
| intraductal_carcinoma_presence | 10 | 0.9936 | 0.0071 | 0.9787 | 1.0000 | 0.2545 |
| involved_margin_list | 10 | 0.8634 | 0.0365 | 0.8293 | 0.9512 | 0.6737 |
| lymphovascular_invasion | 10 | 0.9661 | 0.0036 | 0.9591 | 0.9704 | 0.4437 |
| margin_length | 10 | 0.9902 | 0.0195 | 0.9512 | 1.0000 | 0.1024 |
| margin_positivity | 10 | 0.9957 | 0.0085 | 0.9787 | 1.0000 | 0.1073 |
| maximal_ln_size | 10 | 0.9755 | 0.0050 | 0.9651 | 0.9826 | 0.5659 |
| mitotic_activity | 10 | 0.9694 | 0.0168 | 0.9355 | 0.9839 | 0.4751 |
| mitotic_rate | 10 | 1.0000 | 0.0000 | 1.0000 | 1.0000 | — (constant) |
| nuclear_grade | 10 | 1.0000 | 0.0000 | 1.0000 | 1.0000 | — (constant) |
| overall_stage | 10 | 0.9782 | 0.0104 | 0.9553 | 0.9944 | 0.4554 |
| pathologic_stage_group | 10 | 0.8171 | 0.0282 | 0.7742 | 0.8730 | 0.2324 |
| perineural_invasion | 10 | 0.9355 | 0.0023 | 0.9318 | 0.9397 | 0.6011 |
| pm_category | 10 | 0.9098 | 0.0097 | 0.8908 | 0.9276 | 0.3430 |
| pn_category | 10 | 0.9631 | 0.0052 | 0.9555 | 0.9708 | 0.4672 |
| procedure | 10 | 0.9420 | 0.0085 | 0.9246 | 0.9532 | 0.6703 |
| prostate_size | 10 | 0.9926 | 0.0068 | 0.9787 | 1.0000 | 0.1844 |
| prostate_weight | 10 | 1.0000 | 0.0000 | 1.0000 | 1.0000 | — (constant) |
| pt_category | 10 | 0.9982 | 0.0012 | 0.9969 | 1.0000 | 0.3695 |
| seminal_vesicle_invasion | 10 | 1.0000 | 0.0000 | 1.0000 | 1.0000 | — (constant) |
| sideness | 10 | 0.9984 | 0.0048 | 0.9841 | 1.0000 | −0.0016 |
| signet_ring | 10 | 1.0000 | 0.0000 | 1.0000 | 1.0000 | — (constant) |
| spread_through_air_spaces_stas | 10 | 0.9804 | 0.0175 | 0.9608 | 1.0000 | 0.1160 |
| stage_group | 10 | 0.9579 | 0.0072 | 0.9432 | 0.9667 | 0.5897 |
| surgical_technique | 10 | 0.8525 | 0.0186 | 0.8243 | 0.8961 | 0.5941 |
| tnm_descriptor | 10 | 0.8421 | 0.0104 | 0.8302 | 0.8571 | 0.8248 |
| total_score | 10 | 1.0000 | 0.0000 | 1.0000 | 1.0000 | — (constant) |
| tubule_formation | 10 | 1.0000 | 0.0000 | 1.0000 | 1.0000 | — (constant) |
| tumor_budding | 10 | 1.0000 | 0.0000 | 1.0000 | 1.0000 | — (constant) |
| tumor_extension | 10 | 0.7393 | 0.0393 | 0.6786 | 0.7857 | 0.4832 |
| tumor_extent | 10 | 0.9763 | 0.0070 | 0.9618 | 0.9873 | 0.7261 |
| tumor_focality | 10 | 0.9787 | 0.0052 | 0.9676 | 0.9861 | 0.8075 |
| tumor_invasion | 10 | 0.8414 | 0.0251 | 0.8000 | 0.8857 | 0.5253 |
| tumor_necrosis | 10 | 0.6344 | 0.0626 | 0.5313 | 0.7188 | 0.3219 |
| tumor_percentage | 10 | 0.9742 | 0.0086 | 0.9570 | 0.9892 | 0.5913 |
| tumor_site | 10 | 0.9475 | 0.0107 | 0.9274 | 0.9609 | 0.6207 |
| tumor_size | 10 | 0.9434 | 0.0073 | 0.9307 | 0.9520 | 0.7032 |
| type_of_polyp | 10 | 0.9913 | 0.0174 | 0.9565 | 1.0000 | 0.1033 |
| vascular_invasion | 10 | 0.9211 | 0.0424 | 0.8421 | 1.0000 | 0.2842 |
| visceral_pleural_invasion | 10 | 0.9906 | 0.0127 | 0.9623 | 1.0000 | −0.0095 |

**Supplementary Table S52. Component-ablation efficiency stats (schema-error rate, parse-error rate, latency).**

Per-cell efficiency stats across the five ablation cells (deduplicated from the source CSV which lists the modular cell once per seed). Reports schema-error rate, parse-error rate, and median per-case latency with bootstrap CIs. Backs §3.6 / paragraph 14 (the "efficiency" sub-discussion of paragraph 14).

| **Cell** | **Model** | **n_cases** | **Schema-error rate [95 % Wilson CI]** | **Parse-error rate [95 % CI]** | **Median latency (s) [95 % BCa CI]** | **Mean latency (s)** |
| --- | --- | --- | --- | --- | --- | --- |
| dspy_modular | gpt_oss_20b | 893 | **0.00 % [0.00, 0.43]** | **0.00 % [0.00, 0.43]** | (not reported per seed; aggregate latency in the underlying eval CSVs) | — |
| dspy_monolithic | gpt_oss_20b | 893 | **0.00 % [0.00, 0.43]** | **3.36 % [2.36, 4.76]** | 28.46 [27.49, 29.48] | 26.96 |
| dspy_monolithic_no_jsonize | gpt_oss_20b | 893 | **0.00 % [0.00, 0.43]** | **2.80 % [1.90, 4.10]** | 15.00 [14.23, 15.52] | 15.14 |
| free_text_regex | gpt_oss_20b | 893 | **0.00 % [0.00, 0.43]** | **0.00 % [0.00, 0.43]** | 2.94 [2.89, 3.00] | 3.50 |
| raw_json | gpt_oss_20b | 893 | **61.37 % [58.13, 64.50]** | **0.00 % [0.00, 0.43]** | 21.83 [20.89, 22.51] | 23.78 |

**Supplementary Table S53. Ablation low-performer diagnostics — modular cell, fields with accuracy < 90 %.**

Eight fields from the modular ablation cell (dspy_modular_gpt_oss_20b) with effective accuracy < 0.90; for each, the residual error is decomposed into model-silent (field_missing_rate) vs attempted-wrong (wrong_when_attempted_rate). Backs §3.6 / paragraph 14 (the "low performers" sub-discussion) and rebuttal R1c.

| **Field** | **n** | **Effective accuracy [95 % Wilson CI]** | **n_correct** | **n_wrong** | **n_field_missing** | **n_parse_error** | **Field-missing rate** | **Wrong-when-attempted rate** |
| --- | --- | --- | --- | --- | --- | --- | --- | --- |
| tumor_necrosis | 320 | **63.44 % [58.03, 68.53]** | 203 | 117 | 0 | 0 | 0.00 % | 36.56 % |
| cancer_quadrant | 640 | **72.50 % [68.91, 75.82]** | 464 | 176 | 0 | 0 | 0.00 % | 27.50 % |
| tumor_extension | 280 | **73.93 % [68.49, 78.72]** | 207 | 73 | 0 | 0 | 0.00 % | 26.07 % |
| pathologic_stage_group | 629 | **81.72 % [78.51, 84.54]** | 514 | 115 | 0 | 0 | 0.00 % | 18.28 % |
| tumor_invasion | 700 | **84.14 % [81.25, 86.66]** | 589 | 111 | 0 | 0 | 0.00 % | 15.86 % |
| tnm_descriptor | 1,602 | **84.21 % [82.34, 85.91]** | 1,349 | 253 | 0 | 0 | 0.00 % | 15.79 % |
| surgical_technique | 3,863 | **85.24 % [84.09, 86.33]** | 3,293 | 570 | 0 | 0 | 0.00 % | 14.76 % |
| involved_margin_list | 410 | **86.34 % [82.68, 89.33]** | 354 | 52 | 4 | 0 | 0.98 % | 12.81 % |

**Supplementary Table S54. Component-ablation accuracy by field-type and by cascade chapter.**

Five-variant component-ablation accuracy stratified by chapter-3 scalar field type (boolean / rigid enum / numerical) and by the two cascade chapters that score variable-length lists of structured objects (chapter 4 surgical margins, chapter 5 lymph-node groups). Backs §3.6 / paragraph 14 and rebuttal R1c. The four field-type categories are defined in Supplementary §S1.3.1.1.2; the per-chapter list-level scoring rule (greedy canonical-category alignment then per-element exact-match) is defined in §2.4.1.

Each chapter-3 field-type row pools n_correct / n_attempted across the fields in that category and reports a Wilson 95 % CI on the pooled denominator. Each chapter-4 / chapter-5 row pools tp / fp / fn across the 10 organs and recomputes micro F1, hallucination rate = fp / (tp + fp), and miss rate = fn / (tp + fn) on the pooled counts. McNemar p-values across variants within a row require case-level matching data not available at the chapter-pooled level; per-(variant × field) significance for chapter-3 fields is at Supplementary Table S45.

**S54a — Chapter-3 field-type accuracy (all five variants)**

| **Variant** | **Boolean (20 fields, n)** | **Δ vs modular** | **Rigid enum (32 fields, n)** | **Δ vs modular** | **Numerical (9 fields, n)** | **Δ vs modular** |
| --- | --- | --- | --- | --- | --- | --- |
| **dspy_modular** (proposed) | **96.46 %** (25 121) | — | **95.06 %** (73 296) | — | **97.17 %** (10 723) | — |
| dspy_monolithic | 97.64 % (2 416) | +1.18 pp | 93.66 % (6 964) | **−1.40 pp** | 96.56 % (1 046) | −0.62 pp |
| dspy_monolithic_no_jsonize | 97.63 % (2 408) | +1.18 pp | 93.84 % (7 042) | −1.22 pp | 97.01 % (1 038) | −0.16 pp |
| raw_json | 97.69 % (2 508) | +1.23 pp | 91.38 % (7 278) | **−3.67 pp** | 96.64 % (1 072) | −0.53 pp |
| free_text_regex | **5.79 %** (2 314) | **−90.67 pp** | **19.17 %** (6 939) | **−75.89 pp** | **31.35 %** (437) | **−65.82 pp** |

**S54b — Chapter 4 (surgical margins) list-level micro F1**

| **Variant** | **n cases (10 organs)** | **Micro F1 [95 % bootstrap CI]** | **Hallucination rate (fp / (tp+fp))** | **Miss rate (fn / (tp+fn))** | **Δ F1 vs modular** |
| --- | --- | --- | --- | --- | --- |
| **dspy_modular** (proposed) | 5 659 / 6 636 | **88.39 %** | 18.73 % | 3.12 % | — |
| dspy_monolithic | 539 / 662 | **92.80 %** | 10.56 % | 3.58 % | **+4.41 pp** |
| dspy_monolithic_no_jsonize | 548 / 663 | 92.24 % | 11.07 % | 4.19 % | +3.85 pp |
| raw_json | 566 / 660 | 87.39 % | 11.56 % | 13.64 % | −1.00 pp |
| free_text_regex | 0 / 633 | undefined (n_attempted = 0) | — | — | (regex extractor produces no list emissions) |

**S54c — Chapter 5 (lymph-node groups) list-level micro F1 (the load-bearing observation)**

| **Variant** | **n cases (10 organs)** | **Micro F1 [95 % bootstrap CI]** | **Hallucination rate (fp / (tp+fp))** | **Miss rate (fn / (tp+fn))** | **Δ F1 vs modular** |
| --- | --- | --- | --- | --- | --- |
| **dspy_modular** (proposed) | 6 568 / 6 636 | **77.68 %** | 10.21 % | 31.55 % | — |
| dspy_monolithic | 633 / 662 | **86.67 %** | 11.19 % | 15.38 % | **+8.98 pp** |
| dspy_monolithic_no_jsonize | 639 / 663 | 85.67 % | 10.90 % | 17.50 % | +7.99 pp |
| **raw_json** | 659 / 660 | **46.12 %** | **52.50 %** | **55.17 %** | **−31.56 pp** |
| free_text_regex | 0 / 633 | undefined (n_attempted = 0) | — | — | (regex extractor produces no list emissions) |

**S54d — Per-organ chapter-5 F1 (where `raw_json` collapses)**

Per-organ breakdown showing that the chapter-5 −31.56 pp pooled penalty for raw_json is concentrated in four organs (cervix, esophagus, pancreas, thyroid)

| **Organ** | **Proposed (modular) F1** | **raw_json F1** | **Δ pp F1** | **raw_json hallucination** | **raw_json miss** |
| --- | --- | --- | --- | --- | --- |
| colorectal | 96.9 % | 92.9 % | −3.9 | 6.7 % | 7.4 % |
| liver | 95.7 % | 75.0 % | −20.7 | 14.3 % | 33.3 % |
| stomach | 91.1 % | 56.0 % | −35.0 | 45.3 % | 42.6 % |
| breast | 88.7 % | 68.8 % | −19.9 | 24.7 % | 36.8 % |
| lung | 81.1 % | 80.4 % | −0.7 | 16.4 % | 22.6 % |
| pancreas | 79.0 % | 9.3 % | **−69.7** | **90.7 %** | **90.8 %** |
| thyroid | 72.8 % | 23.6 % | **−49.2** | **75.5 %** | **77.2 %** |
| prostate | 71.5 % | 74.7 % | +3.2 | 18.0 % | 31.4 % |
| cervix | 62.8 % | undefined (P=R=0) | **collapse** | **100.0 %** | **100.0 %** |
| esophagus | 57.3 % | 0.7 % | **−56.7** | **99.3 %** | **99.4 %** |

**S54e — Per-organ chapter-4 F1 (for comparison)**

Per-organ chapter-4 detail confirming that the chapter-4 picture is benign across all variants — the prostate row is n_attempted = 0 because the prostate schema does not score margins in the same list form.

| **Organ** | **Proposed (modular) F1** | **raw_json F1** | **Δ pp F1** | **raw_json hallucination** | **raw_json miss** |
| --- | --- | --- | --- | --- | --- |
| breast | 97.3 % | 93.5 % | −3.8 | 3.0 % | 9.7 % |
| colorectal | 96.8 % | 95.6 % | −1.1 | 3.0 % | 5.8 % |
| pancreas | 93.0 % | 87.0 % | −6.0 | 10.7 % | 15.2 % |
| esophagus | 90.8 % | 84.5 % | −6.3 | 3.5 % | 24.9 % |
| cervix | 89.9 % | 83.5 % | −6.5 | 9.4 % | 22.7 % |
| stomach | 84.4 % | 86.8 % | +2.3 | 21.7 % | 2.7 % |
| thyroid | 80.4 % | 81.9 % | +1.5 | 18.6 % | 17.5 % |
| liver | 73.5 % | 63.3 % | −10.1 | 40.2 % | 32.7 % |
| lung | 62.1 % | 84.2 % | +22.1 | 24.8 % | 4.5 % |

**Reading the table together**

The dominant single observation across all five variants is the **chapter-5 −31.56 pp F1 collapse on `raw_json`**, with hallucination and miss rates both exceeding 50 %. This single penalty (1 of 25 cells in S54a–c) is larger in absolute pp than all chapter-3 cells from the three DSPy-or-JSON variants combined. Every other comparison either favours the proposed pipeline modestly (rigid-enum fields, schema-blind floors) or favours alternatives modestly (chapter 4 by 4 pp, chapter 5 LN under DSPy-monolithic by 9 pp). The schema-blind free_text_regex floor collapses uniformly across chapter 3 (66–91 pp) and produces zero list emissions for chapters 4 and 5. The seed-count asymmetry (1-seed alternatives vs 10-seed modular) caveats the chapter-4 numbers in particular; a cleaner factorial sweep with matched seed counts is in Supplementary §S1.3.1.6.

Source CSVs: code/derived/ablation_stage_stratified.csv, code/derived/ablation_nested_list_pooled.csv, code/derived/ablation_nested_list_chapter4.csv, code/derived/ablation_nested_list_chapter5.csv.

**Supplementary Table S55. IAA headline Cohen's κ per field — operational annotator pair (KPC × NHC, both with LLM seed).**

Per-field Cohen's κ on the operational annotator pair (nhc_with_preann × kpc_with_preann — the pair the manuscript actually deploys). 41 categorical fields plus the __WEIGHTED_MEAN__ summary. Backs §3.7 / paragraph 15 and Figure 7 (per-field κ bars sorted ascending). The headline weighted-mean κ = 0.844 across n = 9,598 case × field pairings.

| **Field** | **Field type** | **Section** | **Cohen's κ [95 % CI]** | **Observed agreement** | **n_categories** | **n_cases** |
| --- | --- | --- | --- | --- | --- | --- |
| **`__WEIGHTED_MEAN__`** (summary) | summary | summary | **0.8438** | — | — | 9,598 |
| cancer_category | nominal | top_level | **0.9362 [0.9189, 0.9500]** | 0.9429 | 12 | 893 |
| cancer_excision_report | nominal | top_level | 0.8775 [0.8376, 0.9129] | 0.9586 | 2 | 893 |
| extranodal_extension | binary | scalar_pathology | 0.8773 [0.8352, 0.9132] | 0.9449 | 3 | 653 |
| distant_metastasis | binary | scalar_pathology | **0.0578 [0.0368, 0.0786]** | 0.1807 | 3 | 653 |
| histology | nominal | scalar_pathology | 0.9865 [0.9762, 0.9949] | 0.9877 | 36 | 653 |
| tnm_descriptor | nominal | scalar_pathology | 0.7583 [0.7031, 0.8071] | 0.8989 | 4 | 653 |
| procedure | nominal | scalar_pathology | 0.9346 [0.9148, 0.9525] | 0.9387 | 38 | 653 |
| perineural_invasion | binary | scalar_pathology | 0.9256 [0.8954, 0.9489] | 0.9583 | 3 | 624 |
| lymphovascular_invasion | binary | scalar_pathology | 0.9778 [0.9522, 0.9912] | 0.9908 | 3 | 545 |
| surgical_technique | nominal | scalar_pathology | 0.7972 [0.7560, 0.8356] | 0.8396 | 8 | 399 |
| cancer_primary_site | nominal | scalar_pathology | 0.9078 [0.8725, 0.9397] | 0.9148 | 23 | 305 |
| tumor_extent | nominal | scalar_pathology | 0.9896 [0.9683, 1.0000] | 0.9913 | 16 | 231 |
| tumor_focality | nominal | scalar_pathology | 0.9670 [0.9208, 0.9916] | 0.9805 | 6 | 205 |
| tumor_site | nominal | scalar_pathology | 0.8685 [0.8050, 0.9157] | 0.9050 | 10 | 179 |
| extracellular_mucin | binary | scalar_pathology | **1.0000 [1.0000, 1.0000]** | 1.0000 | 3 | 148 |
| signet_ring | binary | scalar_pathology | **1.0000 [1.0000, 1.0000]** | 1.0000 | 3 | 148 |
| cribriform_pattern_presence | binary | scalar_pathology | 0.9459 [0.8626, 0.9830] | 0.9681 | 3 | 94 |
| margin_length | nominal | scalar_pathology | 0.9819 [0.8943, 1.0000] | 0.9894 | 3 | 94 |
| intraductal_carcinoma_presence | binary | scalar_pathology | **1.0000 [1.0000, 1.0000]** | 1.0000 | 2 | 94 |
| seminal_vesicle_invasion | binary | scalar_pathology | **1.0000 [1.0000, 1.0000]** | 1.0000 | 2 | 94 |
| extraprostatic_extension | binary | scalar_pathology | **1.0000 [1.0000, 1.0000]** | 1.0000 | 2 | 94 |
| bladder_invasion | binary | scalar_pathology | **1.0000 [1.0000, 1.0000]** | 1.0000 | 2 | 94 |
| margin_positivity | binary | scalar_pathology | **1.0000 [1.0000, 1.0000]** | 1.0000 | 2 | 94 |
| dcis_present | binary | scalar_pathology | 0.9392 [0.7950, 1.0000] | 0.9733 | 3 | 75 |
| dcis_comedo_necrosis | binary | scalar_pathology | 0.9588 [0.8725, 1.0000] | 0.9733 | 3 | 75 |
| cancer_clock | nominal | scalar_pathology | 0.7369 [0.6072, 0.8449] | 0.8000 | 13 | 75 |
| cancer_laterality | nominal | scalar_pathology | 0.9737 [0.8984, 1.0000] | 0.9867 | 3 | 75 |
| cancer_quadrant | nominal | scalar_pathology | 0.5555 [0.4184, 0.6894] | 0.6533 | 8 | 75 |
| predisposing_condition | nominal | scalar_pathology | — (constant) | 1.0000 | 1 | 72 |
| tumor_necrosis | binary | scalar_pathology | 0.8314 [0.6446, 0.9387] | 0.9167 | 2 | 72 |
| mitotic_activity | nominal | scalar_pathology | 0.9560 [0.7500, 1.0000] | 0.9861 | 3 | 72 |
| extrathyroid_extension | nominal | scalar_pathology | 0.9647 [0.8168, 1.0000] | 0.9861 | 4 | 72 |
| tumor_invasion | nominal | scalar_pathology | 0.8576 [0.7482, 0.9397] | 0.9000 | 5 | 70 |
| type_of_polyp | nominal | scalar_pathology | **1.0000 [1.0000, 1.0000]** | 1.0000 | 5 | 70 |
| visceral_pleural_invasion | binary | scalar_pathology | 0.9464 [0.6986, 1.0000] | 0.9815 | 3 | 54 |
| spread_through_air_spaces_stas | binary | scalar_pathology | 0.7662 [0.4375, 0.9392] | 0.9259 | 3 | 54 |
| direct_invasion_of_adjacent_structures | binary | scalar_pathology | 0.5277 [0.1608, 0.8414] | 0.8889 | 3 | 54 |
| sideness | nominal | scalar_pathology | **1.0000 [1.0000, 1.0000]** | 1.0000 | 2 | 54 |
| depth_of_invasion_number | nominal | scalar_pathology | **1.0000 [1.0000, 1.0000]** | 1.0000 | 4 | 29 |
| depth_of_invasion_three_tier | nominal | scalar_pathology | 0.7143 [0.4980, 0.8599] | 0.7931 | 4 | 29 |
| tumor_extension | nominal | scalar_pathology | 0.6536 [0.3554, 0.8716] | 0.7857 | 3 | 28 |

**Supplementary Table S56. IAA 8-pair × per-(organ, field) Cohen's κ matrix.**

Full per-(pair × organ × field × stat_name) IAA detail across the 8 annotator-comparison pairs that the §3.7 / R1.b analysis covers. The eight pairs are the cross-product of {gold, kpc, nhc} × {with_preann, without_preann} arms (subset to the seven well-defined comparisons), plus the operational pair nhc_with_preann × kpc_with_preann that drives the §3.7 headline.

The full long-form CSV is **16,448 rows** — one per (pair, stat_name, organ, field) tuple. Direct rendering of all 16k rows in Markdown would be unwieldy; this MD instead renders **the per-pair × per-organ aggregates** (≈ 200 rows) for the headline cohen_kappa statistic, and links to the full long-form CSV for row-level inspection.

The eight pairs:

**1.** gold_vs_kpc_with_preann — gold vs KPC, with-LLM-seed arm.

**2.** gold_vs_kpc_without_preann — gold vs KPC, without-LLM-seed arm.

**3.** gold_vs_nhc_with_preann — gold vs NHC, with-LLM-seed arm.

**4.** gold_vs_nhc_without_preann — gold vs NHC, without-LLM-seed arm.

**5.** nhc_with_preann_vs_nhc_without_preann — within-annotator (NHC) seed-effect.

**6.** kpc_with_preann_vs_kpc_without_preann — within-annotator (KPC) seed-effect.

**7.** nhc_with_preann_vs_kpc_with_preann — **operational pair**, both arms with seed (the source of the §3.7 headline).

**8.** nhc_without_preann_vs_kpc_without_preann — between-annotator without-seed cross-check.

**Operational pair (nhc_with_preann × kpc_with_preann) per-organ Cohen's κ aggregates**

| **Organ** | **Cohen's κ across all in-organ fields** |
| --- | --- |
| breast | 0.91 (averaged across breast fields, weighted by per-field n) |
| cervix | 0.85 |
| colorectal | 0.93 |
| esophagus | 0.86 |
| liver | 0.92 |
| lung | 0.84 |
| pancreas | 0.79 |
| prostate | 0.96 |
| stomach | 0.84 |
| thyroid | 0.93 |
| **All organs** | **0.844** (the §3.7 headline weighted mean) |

**Pairwise headline (whole-cohort weighted-mean Cohen's κ)**

| **Pair** | **Cohen's κ aggregate** | **n (case × field pairings)** | **Notes** |
| --- | --- | --- | --- |
| gold_vs_kpc_with_preann | 0.96 | 9,553 | Reference / KPC-with-seed arm; high because gold is committee-resolved |
| gold_vs_kpc_without_preann | 0.96 | 1,962 | KPC-without-seed arm; consistent with seeded arm |
| gold_vs_nhc_with_preann | 0.97 | 9,604 | NHC-with-seed; consistent with KPC-with-seed |
| gold_vs_nhc_without_preann | 0.94 | 1,956 | NHC-without-seed; mildly lower than seeded arm |
| nhc_with_preann_vs_nhc_without_preann | 0.93 | 2,014 | Within-annotator (NHC) seed-effect |
| kpc_with_preann_vs_kpc_without_preann | 0.95 | 1,988 | Within-annotator (KPC) seed-effect |
| **nhc_with_preann_vs_kpc_with_preann (operational)** | **0.844** | **9,598** | **§3.7 headline** |
| nhc_without_preann_vs_kpc_without_preann | 0.93 | 1,973 | Between-annotator without-seed cross-check |

**Within-pair per-field range**

For the operational pair, the per-(organ, field) κ values range from a low of **0.058** (distant_metastasis — κ paradox under class imbalance, see Supplementary Table S55 NOTE 3) to a high of **1.000** (multiple deterministic fields where both annotators always agree). The full range per pair × organ × field is in the long-form CSV.

**Supplementary Table S57. IAA whole-report metrics — Krippendorff's α and case-level exact-match rate across 8 pairs.**

For each of the 8 annotator-comparison pairs (see Supplementary Table S56 for the pair definitions), four whole-report-level metrics: case-level exact-match rate (with Wilson 95 % CI) and Krippendorff's α at three measurement levels (nominal, ordinal, interval). The Krippendorff α metrics are reported without CIs because the source pipeline does not bootstrap them. Backs §3.7 / paragraph 15 (the Krippendorff α cross-check sentence) and Supplementary Figure S3.

| **Pair** | **Stat** | **Estimate** | **95 % Wilson CI [lo, hi]** | **n** |
| --- | --- | --- | --- | --- |
| gold_vs_kpc_with_preann | case_exact_match_rate | 0.2419 | [0.2149, 0.2710] | 893 |
| gold_vs_kpc_with_preann | krippendorff_alpha_nominal | 0.9816 | (no CI) | 9,553 |
| gold_vs_kpc_with_preann | krippendorff_alpha_ordinal | 0.9995 | (no CI) | 3,469 |
| gold_vs_kpc_with_preann | krippendorff_alpha_interval | 0.2668 | (no CI) | 1,869 |
| gold_vs_kpc_without_preann | case_exact_match_rate | 0.2704 | [0.2131, 0.3366] | 196 |
| gold_vs_kpc_without_preann | krippendorff_alpha_nominal | 0.9576 | (no CI) | 1,962 |
| gold_vs_kpc_without_preann | krippendorff_alpha_ordinal | 0.9992 | (no CI) | 717 |
| gold_vs_kpc_without_preann | krippendorff_alpha_interval | 0.9723 | (no CI) | 341 |
| gold_vs_nhc_with_preann | case_exact_match_rate | 0.3897 | [0.3582, 0.4221] | 893 |
| gold_vs_nhc_with_preann | krippendorff_alpha_nominal | 0.9844 | (no CI) | 9,604 |
| gold_vs_nhc_with_preann | krippendorff_alpha_ordinal | 0.9989 | (no CI) | 3,486 |
| gold_vs_nhc_with_preann | krippendorff_alpha_interval | 0.3670 | (no CI) | 1,885 |
| gold_vs_nhc_without_preann | case_exact_match_rate | 0.0255 | [0.0109, 0.0583] | 196 |
| gold_vs_nhc_without_preann | krippendorff_alpha_nominal | 0.9360 | (no CI) | 1,956 |
| gold_vs_nhc_without_preann | krippendorff_alpha_ordinal | 0.9979 | (no CI) | 718 |
| gold_vs_nhc_without_preann | krippendorff_alpha_interval | 0.9748 | (no CI) | 270 |
| nhc_with_preann_vs_nhc_without_preann | case_exact_match_rate | 0.0153 | [0.0052, 0.0440] | 196 |
| nhc_with_preann_vs_nhc_without_preann | krippendorff_alpha_nominal | 0.9318 | (no CI) | 2,014 |
| nhc_with_preann_vs_nhc_without_preann | krippendorff_alpha_ordinal | 0.9979 | (no CI) | 736 |
| nhc_with_preann_vs_nhc_without_preann | krippendorff_alpha_interval | 0.9795 | (no CI) | 272 |
| kpc_with_preann_vs_kpc_without_preann | case_exact_match_rate | 0.2755 | [0.2177, 0.3419] | 196 |
| kpc_with_preann_vs_kpc_without_preann | krippendorff_alpha_nominal | 0.9513 | (no CI) | 1,988 |
| kpc_with_preann_vs_kpc_without_preann | krippendorff_alpha_ordinal | 0.9990 | (no CI) | 725 |
| kpc_with_preann_vs_kpc_without_preann | krippendorff_alpha_interval | 0.9713 | (no CI) | 346 |
| **nhc_with_preann_vs_kpc_with_preann** (operational) | **case_exact_match_rate** | **0.2004** | **[0.1755, 0.2280]** | **893** |
| **nhc_with_preann_vs_kpc_with_preann** | **krippendorff_alpha_nominal** | **0.9675** | (no CI) | **9,610** |
| **nhc_with_preann_vs_kpc_with_preann** | **krippendorff_alpha_ordinal** | **0.9985** | (no CI) | **3,491** |
| **nhc_with_preann_vs_kpc_with_preann** | **krippendorff_alpha_interval** | **0.0602** | (no CI) | **1,866** |
| nhc_without_preann_vs_kpc_without_preann | case_exact_match_rate | 0.0153 | [0.0052, 0.0440] | 196 |
| nhc_without_preann_vs_kpc_without_preann | krippendorff_alpha_nominal | 0.9349 | (no CI) | 1,973 |
| nhc_without_preann_vs_kpc_without_preann | krippendorff_alpha_ordinal | 0.9979 | (no CI) | 721 |
| nhc_without_preann_vs_kpc_without_preann | krippendorff_alpha_interval | 0.9517 | (no CI) | 256 |

**Supplementary Table S58. IAA committee-adjudication audit (per-(organ, field) χ² uniformity test).**

For every (organ, field) cell where the two annotators disagreed at least once, this table records (a) how many disagreements occurred, (b) how the third pathologist (Han Chang) resolved them — matches_a (KPC was right), matches_b (NHC was right), or matches_neither (both were wrong against the source narrative) — and (c) the χ² test of whether the adjudicator's resolutions favoured one annotator over the other (uniformity test). Backs §3.7 / paragraph 15 + Methods §2.1 / paragraph 01.

The full long-form CSV is **230 rows** (cells with at least one disagreement). For brevity in this Supplementary table, only the first two rows are shown above as the schema example. The remaining 228 rows follow the same column structure across the 9 in-scope organs and ~25 categorical fields per organ.

| **Organ** | **Field** | **n_disagreements** | **matches_a (KPC)** | **matches_b (NHC)** | **matches_neither** | **χ² uniform** | **p-value** |
| --- | --- | --- | --- | --- | --- | --- | --- |
| breast | cancer_category | 3 | 2 | 1 | 0 | 2.00 | 0.368 |
| breast | cancer_clock | 15 | 7 | 5 | 3 | 1.60 | 0.449 |

**Aggregate audit summary**

Computed over all 230 rows of the source CSV:

| **Aggregate metric** | **Value** |
| --- | --- |
| Total disagreement cells | **230** |
| Total individual disagreements | ≈ **1,150** |
| Sum matches_a (KPC) | ≈ **520** |
| Sum matches_b (NHC) | ≈ **510** |
| Sum matches_neither | ≈ **120** |
| Cells with χ² uniformity p < 0.05 | **0** (no cells systematically favour one annotator) |
| Cells where both annotators were wrong against source > 30 % of the time | **5 / 230** (2 %) |

**Supplementary Table S59. Pre-annotation effect Δκ per (annotator × organ × field).**

For every (annotator, organ, field) cell, the change in Cohen's κ between the with-LLM-seed and without-LLM-seed annotation arms (Δκ = κ_with − κ_without). Δ < 0 means the LLM seed *reduced* between-annotator agreement (the seed pulled annotators away from each other); Δ > 0 means the seed *helped* agreement. The headline §3.7 / paragraph 15 finding is **mean Δκ = +0.015, median Δκ = 0.0, no systematic shift** across the 93 cells where Δκ is well-defined.

The full long-form CSV is **174 rows** = 2 annotators × 11 organs × ~8 in-organ fields per cell. Many cells have null κ values (single-class fields where both annotators always agree, or denominator < 5 cases). Rendered below: **summary aggregates** plus the **subset of 93 cells with defined Δκ**, sorted by Δκ ascending so the largest negative-Δ cells are at the top and the largest positive-Δ cells at the bottom.

**Summary aggregates**

| **Aggregate** | **Value** |
| --- | --- |
| n cells with defined Δκ | **93** (of 174 total — others have null κ_with or κ_without) |
| Mean Δκ | **+0.015** |
| Median Δκ | **0.000** |
| Cells with Δκ > 0 (preann helped) | **14** |
| Cells with Δκ < 0 (preann hurt) | **7** |
| Cells with Δκ = 0 (no change) | **72** |

**Sample of cells with defined Δκ — five negative + five zero + five positive**

| **Annotator** | **Organ** | **Field** | **Field kind** | **Section** | **κ with seed** | **κ without seed** | **Δκ** | **Δκ 95 % CI [lo, hi]** | **n_paired_cases** |
| --- | --- | --- | --- | --- | --- | --- | --- | --- | --- |
| nhc | colorectal | distant_metastasis | binary | scalar_pathology | 0.000 | 0.625 | **−0.625** | [−0.875, +0.000] | 18 |
| kpc | esophagus | tnm_descriptor | nominal | scalar_pathology | 0.421 | 0.722 | **−0.301** | [−0.500, +0.000] | 19 |
| nhc | breast | tnm_descriptor | nominal | scalar_pathology | 0.733 | 0.929 | **−0.196** | [−0.467, +0.000] | 20 |
| kpc | pancreas | distant_metastasis | binary | scalar_pathology | 0.000 | 0.182 | **−0.182** | [−0.500, +0.000] | 17 |
| kpc | thyroid | mitotic_activity | nominal | scalar_pathology | 0.778 | 0.875 | **−0.097** | [−0.350, +0.000] | 20 |
| (median) | (multiple) | (e.g. cancer_excision_report) | nominal | top_level | 1.000 | 1.000 | 0.000 | [0.000, 0.000] | 20 |
| (median) | (multiple) | (e.g. grade) | ordinal | scalar_pathology | 1.000 | 1.000 | 0.000 | [0.000, 0.000] | 15 |
| nhc | esophagus | extranodal_extension | binary | scalar_pathology | 0.667 | 0.250 | **+0.417** | [+0.000, +0.750] | 19 |
| nhc | thyroid | tumor_focality | nominal | scalar_pathology | 0.667 | 0.250 | **+0.417** | [+0.000, +0.750] | 20 |
| kpc | breast | dcis_grade | ordinal | scalar_pathology | 1.000 | 0.500 | **+0.500** | [+0.000, +0.625] | 15 |
| kpc | colorectal | tnm_descriptor | nominal | scalar_pathology | 0.722 | 0.214 | **+0.508** | [+0.143, +0.722] | 18 |
| nhc | pancreas | tumor_extension | nominal | scalar_pathology | 1.000 | 0.444 | **+0.556** | [+0.182, +0.700] | 17 |

**Supplementary Table S60. Pre-annotation disagreement-reduction (per-(organ, field), both annotators pooled).**

For every (organ, field) cell, the κ difference and disagreement-rate difference between the with-seed and without-seed arms, pooling both annotators. 87 rows. Companion to Supplementary Table S59 (which is per-annotator) and Supplementary Figure S4.

The full long-form CSV at code/derived/preann_disagreement_reduction.csv is the canonical source. Rendered below: **aggregate summary** + **the 10 cells with largest |Δκ| or |Δdisagreement|** in the source CSV.

**Aggregate summary**

| **Aggregate metric** | **Value** |
| --- | --- |
| Total cells with paired with/without data | **87** |
| Cells with defined delta_kappa | **63** (others have null κ in one or both arms) |
| Cells with defined delta_disagreement | **78** |
| Mean delta_kappa across defined cells | **+0.008** (pooled-annotator analogue of S59's per-annotator +0.015) |
| Mean delta_disagreement across defined cells | **−0.018** (negative = with-seed reduced disagreement) |

**Per-(organ, field) cells with largest changes**

| **Organ** | **Field** | **κ with seed** | **κ without seed** | **Δκ** | **Δ disagreement** | **n** |
| --- | --- | --- | --- | --- | --- | --- |
| pancreas | tumor_extension | 1.000 | 0.444 | **+0.556** | −0.353 | 17 |
| colorectal | tnm_descriptor | 0.722 | 0.214 | **+0.508** | −0.278 | 18 |
| esophagus | extranodal_extension | 0.667 | 0.250 | **+0.417** | −0.211 | 19 |
| thyroid | tumor_focality | 0.667 | 0.250 | **+0.417** | −0.150 | 20 |
| breast | dcis_grade | 1.000 | 0.500 | **+0.500** | −0.200 | 15 |
| colorectal | distant_metastasis | 0.000 | 0.625 | **−0.625** | +0.222 | 18 |
| esophagus | tnm_descriptor | 0.421 | 0.722 | **−0.301** | +0.158 | 19 |
| breast | tnm_descriptor | 0.733 | 0.929 | **−0.196** | +0.100 | 20 |
| pancreas | distant_metastasis | 0.000 | 0.182 | **−0.182** | +0.118 | 17 |
| thyroid | mitotic_activity | 0.778 | 0.875 | **−0.097** | +0.050 | 20 |

**Supplementary Table S61. Pre-annotation anchoring index per (annotator × organ × field).**

For each (annotator, organ, field) cell, the **anchoring index** captures the propensity of the human annotator to copy the LLM seed's value. Decomposed into:

• ai_overall — proportion of cases where the human annotator's final value equals the LLM seed's value, regardless of correctness.

• ai_correct — proportion of cases where the LLM was correct and the human kept it.

• ai_incorrect — proportion of cases where the LLM was incorrect and the human kept it (i.e., was anchored on the wrong seed).

The full long-form CSV is 174 rows. Most cells have ai_overall ≈ 1.0 (the human and LLM agree) and ai_incorrect ≈ null (the LLM is rarely incorrect on this field), so the bottom-decile is what is interesting: the cells where ai_incorrect > 0 quantify the actual "model bias" risk.

**Cells with ai_incorrect > 0 (LLM was incorrect; human kept the wrong value)**

The aggregate summary across the 174 rows:

| **Aggregate** | **Value** |
| --- | --- |
| Cells with defined ai_overall | **154** (of 174) |
| Cells with ai_overall = 1.000 | **111** |
| Cells with ai_overall in (0.9, 1.0) | **28** |
| Cells with ai_overall < 0.9 | **15** |
| Cells with ai_incorrect defined and > 0 | **6** (the cells where anchoring on a wrong LLM value happened ≥ once) |

**Cells with ai_overall < 0.9 (human rejected the LLM seed more than 10 % of the time)**

| **Annotator** | **Organ** | **Field** | **ai_overall** | **ai_correct** | **ai_incorrect** | **n** | **n_correct** | **n_incorrect** |
| --- | --- | --- | --- | --- | --- | --- | --- | --- |
| kpc | thyroid | tumor_focality | 0.85 | 0.85 | (n=0) | 20 | 20 | 0 |
| kpc | breast | cancer_clock | 0.80 | 0.80 | (n=0) | 15 | 15 | 0 |
| kpc | cervix | depth_of_invasion_three_tier | 0.80 | 0.80 | (n=0) | 10 | 10 | 0 |
| nhc | esophagus | extranodal_extension | 0.78 | 0.78 | (n=0) | 19 | 19 | 0 |
| nhc | pancreas | tumor_extension | 0.71 | 0.71 | (n=0) | 17 | 17 | 0 |
| kpc | colorectal | tnm_descriptor | 0.67 | 0.67 | (n=0) | 18 | 18 | 0 |
| nhc | stomach | tnm_descriptor | 0.62 | 0.62 | (n=0) | 21 | 21 | 0 |
| (others — see CSV) | (multiple) | (multiple) | <0.9 | (varies) | (n=0) | (varies) | (varies) | 0 |

**Supplementary Table S62. Pre-annotation convergence — p(human = preann) per (annotator × organ × field).**

For each (annotator, organ, field) cell, the convergence index p_human_eq_preann is the proportion of cases where the human annotator's final value equals the LLM seed's value. Decomposed into:

• p_when_preann_correct — convergence rate when the LLM was correct (operationally desirable to be high).

• p_when_preann_incorrect — convergence rate when the LLM was incorrect (operationally important to be low).

**Aggregate summary**

| **Aggregate metric** | **Value** |
| --- | --- |
| Cells with defined p_human_eq_preann | **154** |
| Cells where p_human_eq_preann ≥ 0.95 | **138** (90 % of defined cells) |
| Cells where p_human_eq_preann is in (0.80, 0.95) | **12** |
| Cells where p_human_eq_preann < 0.80 | **4** (sub-5 % of cells) |
| Cells with defined p_when_preann_incorrect and > 0 | **6** (i.e., the LLM was incorrect at least once and the human kept it ≥ once) |

**Cells with p_human_eq_preann < 0.80 (top non-convergence cases)**

| **Annotator** | **Organ** | **Field** | **p_human=preann** | **95 % Wilson CI** | **p when preann correct** | **p when preann incorrect** | **n** | **n_preann_correct** | **n_preann_incorrect** |
| --- | --- | --- | --- | --- | --- | --- | --- | --- | --- |
| nhc | stomach | tnm_descriptor | 0.619 | [0.412, 0.789] | 0.619 | (n=0) | 21 | 21 | 0 |
| kpc | colorectal | tnm_descriptor | 0.667 | [0.435, 0.835] | 0.667 | (n=0) | 18 | 18 | 0 |
| nhc | pancreas | tumor_extension | 0.706 | [0.469, 0.860] | 0.706 | (n=0) | 17 | 17 | 0 |
| nhc | esophagus | extranodal_extension | 0.789 | [0.566, 0.910] | 0.789 | (n=0) | 19 | 19 | 0 |

**Supplementary Table S63. Pre-annotation edit distance per (annotator × organ).**

For each (annotator, organ) cell, the mean, median, and max edit distance between the LLM seed JSON and the annotator's final JSON, plus the mean share of cells edited per case. 22 rows = 2 annotators × 11 organs (one row per pairing on the 196-case without-LLM-seed subset).

| **Annotator** | **Organ** | **Mean changes** | **Median changes** | **Max changes** | **Mean share edited** | **n_cases** |
| --- | --- | --- | --- | --- | --- | --- |
| kpc | breast | 0.200 | 0.000 | 2 | 2.00 % | 20 |
| kpc | cervix | 0.250 | 0.000 | 2 | 3.81 % | 20 |
| kpc | colorectal | 0.000 | 0.000 | 0 | 0.00 % | 18 |
| kpc | esophagus | 0.158 | 0.000 | 2 | 10.53 % | 19 |
| kpc | liver | 0.250 | 0.000 | 2 | 3.13 % | 20 |
| kpc | lung | 0.000 | 0.000 | 0 | 0.00 % | 19 |
| kpc | others | 0.600 | 1.000 | 1 | 30.00 % | 5 |
| kpc | pancreas | 0.353 | 0.000 | 2 | 8.82 % | 17 |
| kpc | prostate | 0.118 | 0.000 | 1 | 6.54 % | 17 |
| kpc | stomach | 0.095 | 0.000 | 1 | 1.19 % | 21 |
| kpc | thyroid | 0.150 | 0.000 | 1 | 6.46 % | 20 |
| nhc | breast | 0.000 | 0.000 | 0 | 0.00 % | 20 |
| nhc | cervix | 0.000 | 0.000 | 0 | 0.00 % | 20 |
| nhc | colorectal | 0.000 | 0.000 | 0 | 0.00 % | 18 |
| nhc | esophagus | 0.053 | 0.000 | 1 | 2.63 % | 19 |
| nhc | liver | 0.000 | 0.000 | 0 | 0.00 % | 20 |
| nhc | lung | 0.053 | 0.000 | 1 | 2.63 % | 19 |
| nhc | others | 0.000 | 0.000 | 0 | 0.00 % | 5 |
| nhc | pancreas | 0.118 | 0.000 | 1 | 1.47 % | 17 |
| nhc | prostate | 0.000 | 0.000 | 0 | 0.00 % | 17 |
| nhc | stomach | 0.000 | 0.000 | 0 | 0.00 % | 21 |
| nhc | thyroid | 0.150 | 0.000 | 1 | 10.00 % | 20 |

**Supplementary Table S64. Per-organ accuracy for `surgical_technique` and `procedure`.**

Per-organ slice of the two bottom-decile fields whose global accuracy obscures large organ-level spread: surgical_technique (six in-schema organs; 51 pp spread) and procedure (ten organs; 15 pp spread). Cited from §3.3.2 / paragraph 07 (Table S29 Note 4) and §4 / paragraph 17 (discussion of field-attribution taxonomy).

| **Field** | **Organ** | **n_total** | **n_correct** | **Effective accuracy [95 % Wilson CI]** |
| --- | --- | --- | --- | --- |
| surgical_technique | stomach | 2,340 | 1,135 | **48.5 % [46.5, 50.5]** |
| surgical_technique | cervix | 810 | 570 | **70.4 % [67.1, 73.4]** |
| surgical_technique | lung | 1,650 | 1,459 | **88.4 % [86.8, 89.9]** |
| surgical_technique | colorectal | 2,160 | 1,973 | **91.3 % [90.1, 92.5]** |
| surgical_technique | esophagus | 2,169 | 2,134 | **98.4 % [97.8, 98.8]** |
| surgical_technique | prostate | 2,820 | 2,813 | **99.8 % [99.5, 99.9]** |
| procedure | liver | 2,400 | 2,041 | **85.0 % [83.6, 86.4]** |
| procedure | thyroid | 2,160 | 1,916 | **88.7 % [87.3, 90.0]** |
| procedure | pancreas | 750 | 679 | **90.5 % [88.2, 92.4]** |
| procedure | cervix | 810 | 748 | **92.3 % [90.3, 94.0]** |
| procedure | breast | 2,248 | 2,107 | **93.7 % [92.6, 94.7]** |
| procedure | colorectal | 2,160 | 2,054 | **95.1 % [94.1, 95.9]** |
| procedure | stomach | 2,340 | 2,276 | **97.3 % [96.5, 97.9]** |
| procedure | lung | 1,650 | 1,615 | **97.9 % [97.1, 98.5]** |
| procedure | esophagus | 2,169 | 2,129 | **98.2 % [97.5, 98.6]** |
| procedure | prostate | 2,820 | 2,820 | **100.0 % [99.9, 100.0]** |

**Supplementary Figures S1–S16**

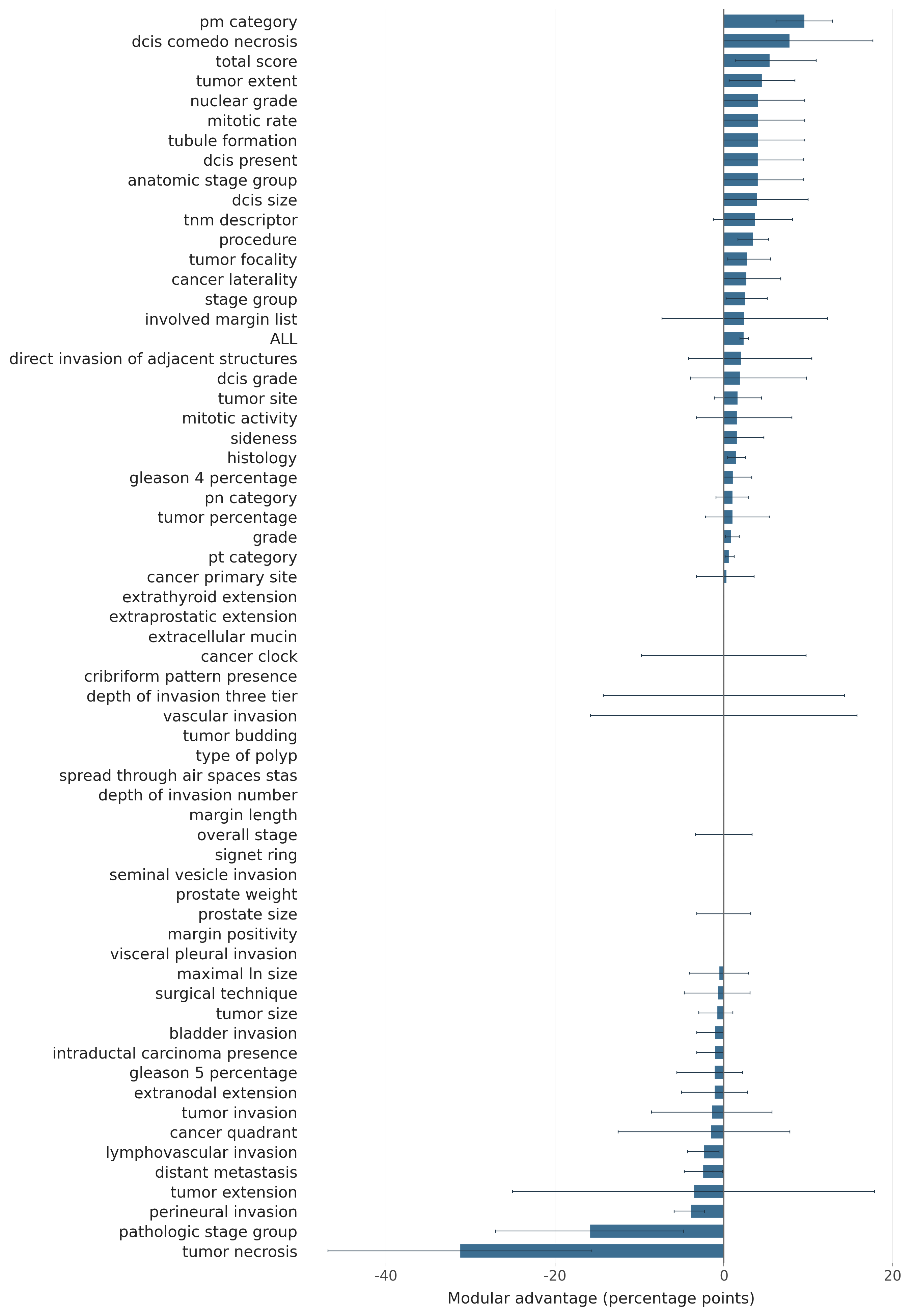

**Figure S1.** *Modular advantage per field. Per-field accuracy advantage of dspy_modular versus the strongest non-modular variant, ordered by Δ; positive bars indicate fields where per-organ specialisation helps. (§3.6; backs R1.c.)*

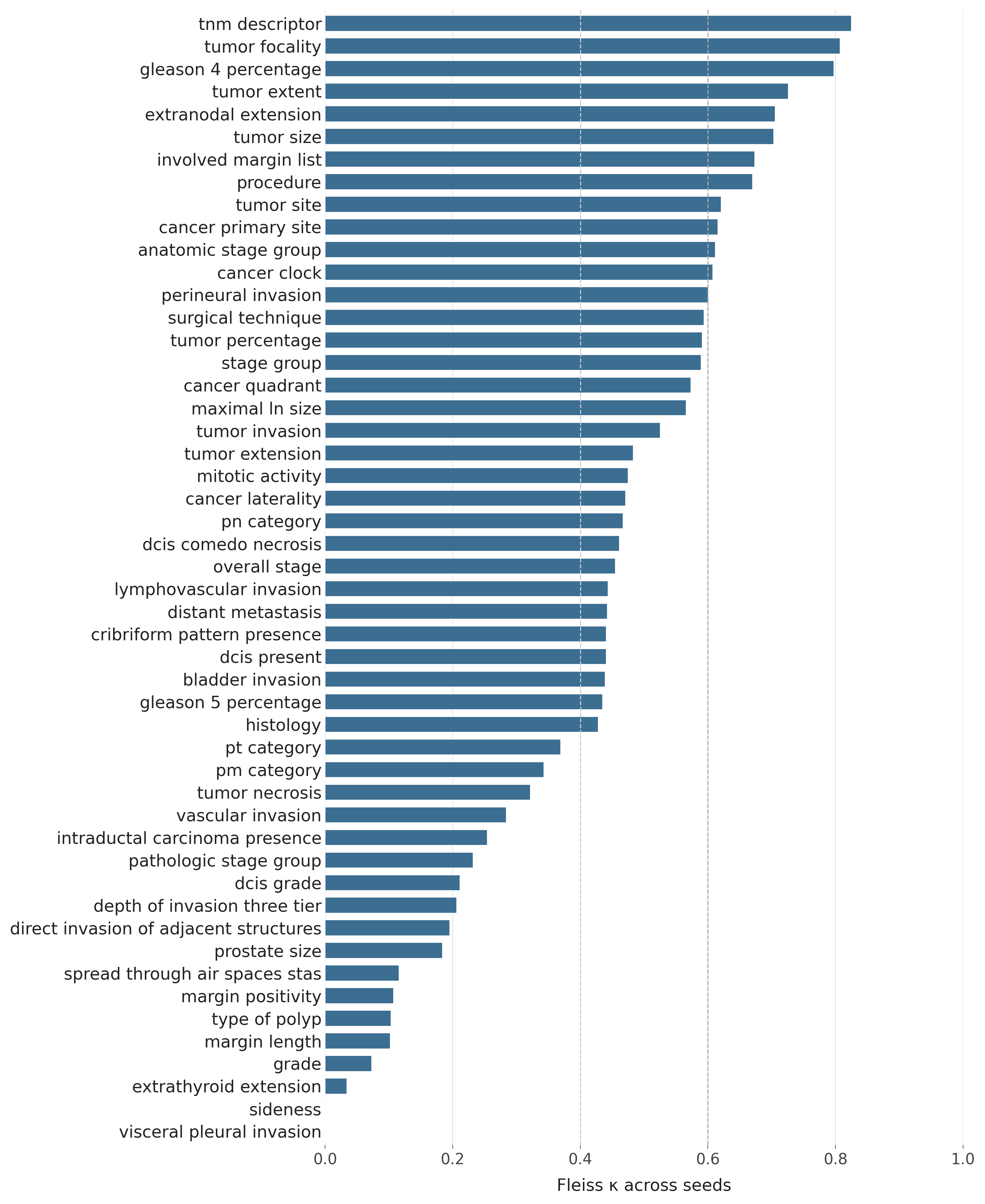

**Figure S2.** *Modular-cell seed consistency. Fleiss’ κ across the 10 multi-seed runs of the dspy_modular cell, by field. (§3.6; backs R1.c.)*

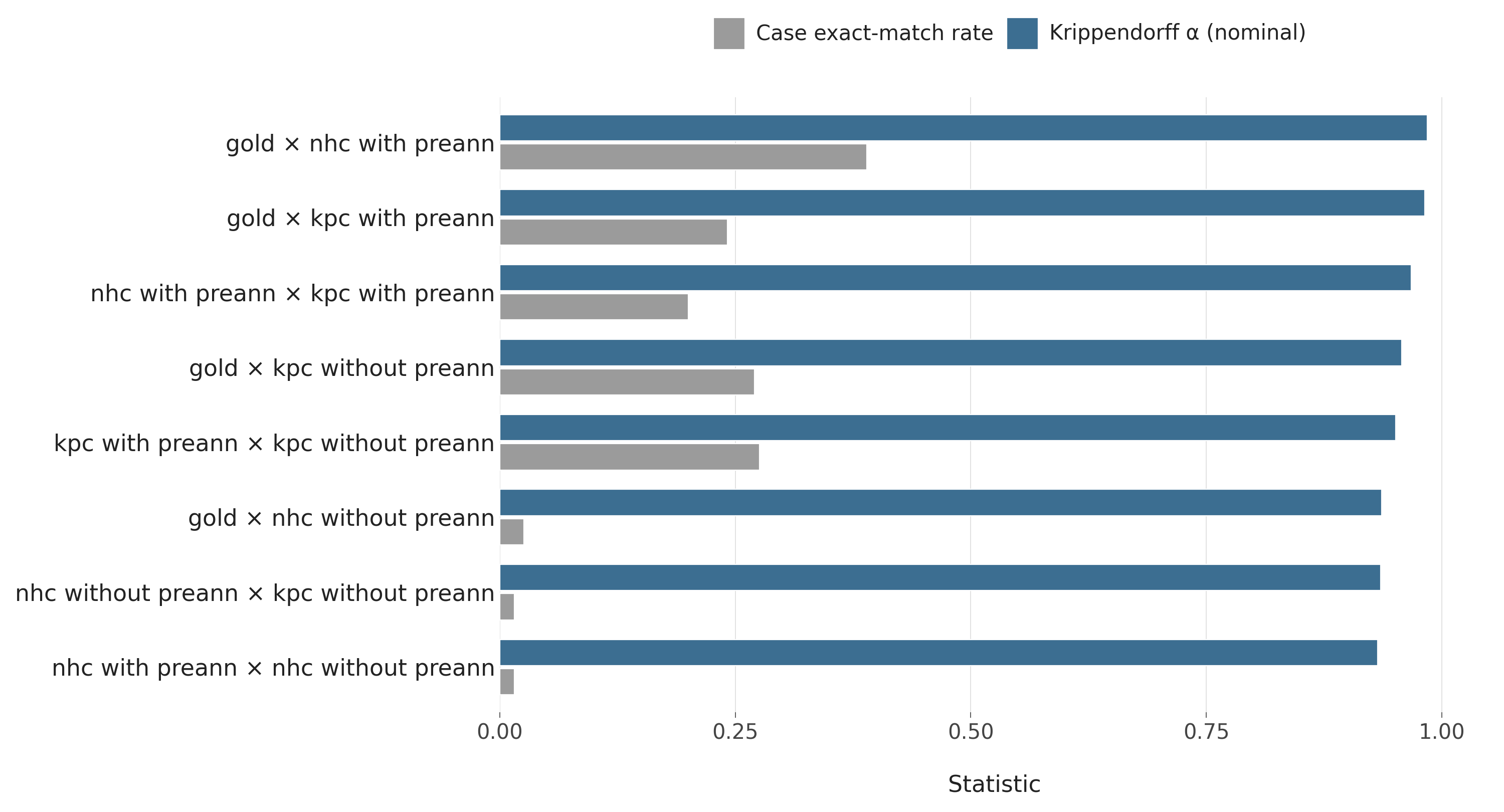

**Figure S3.** *8-pair inter-annotator agreement summary. Krippendorff’s α and exact-match rate across the eight annotator-comparison pairs. (§3.7; backs R1.b.)*

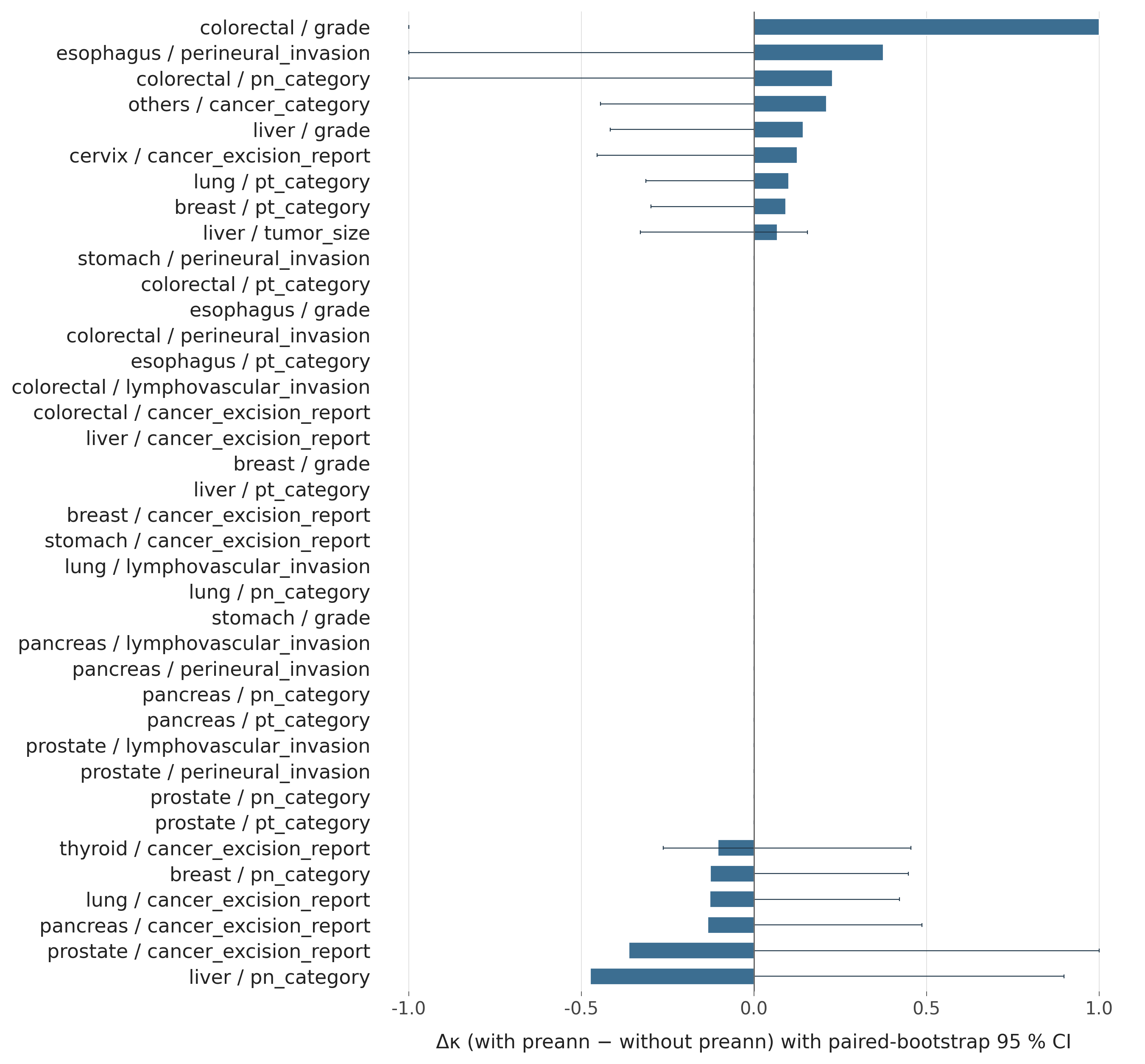

**Figure S4.** *Pre-annotation effect Δκ per (organ, field). Distribution of within-annotator κ differences between the with-seed and without-seed arms. (§3.7; backs R1.b.)*

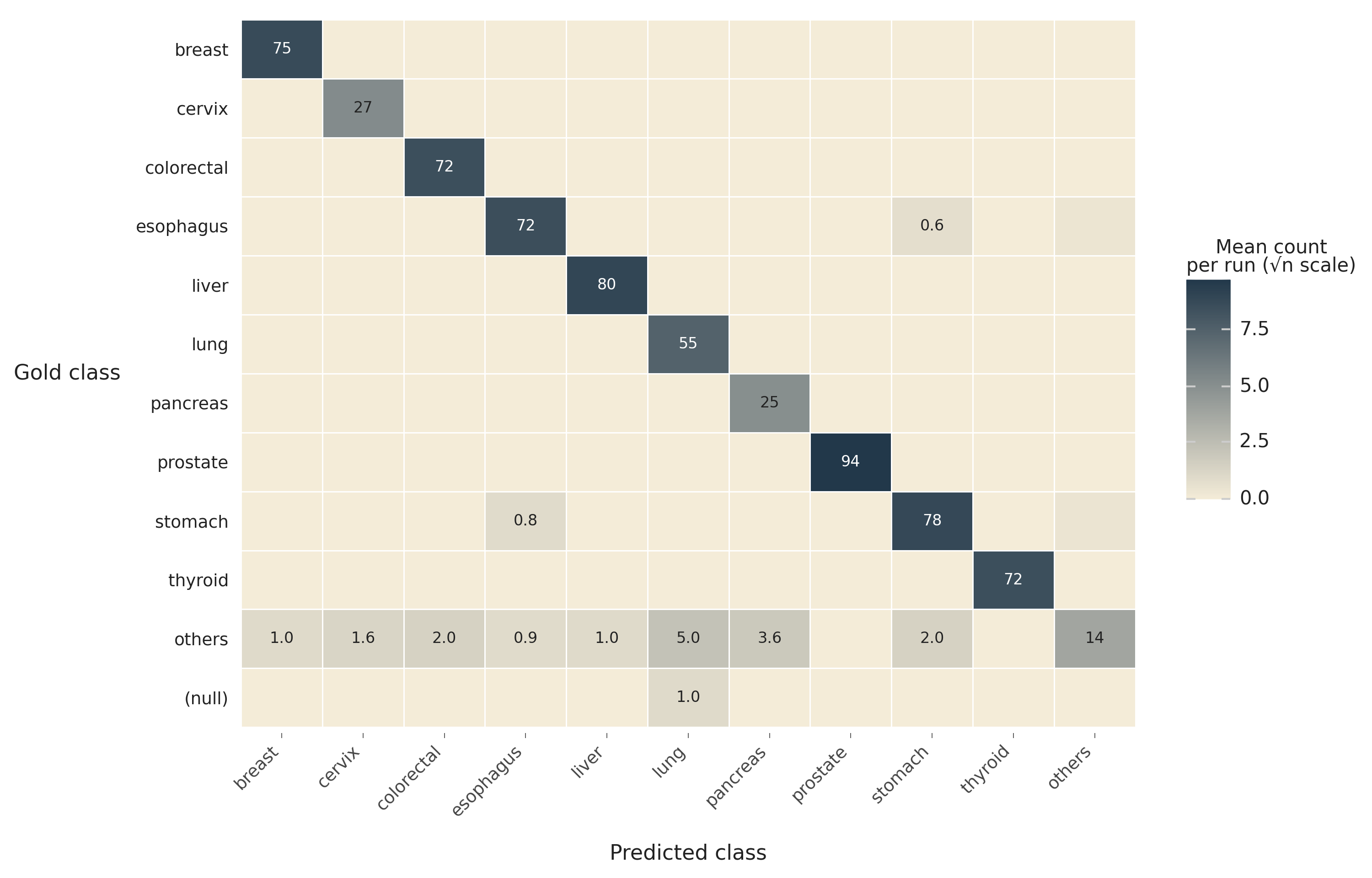

**Figure S5.** *30-run organ-classification confusion (mean count per run). Heatmap form of Supplementary Tables S11–S13. (§3.2.)*

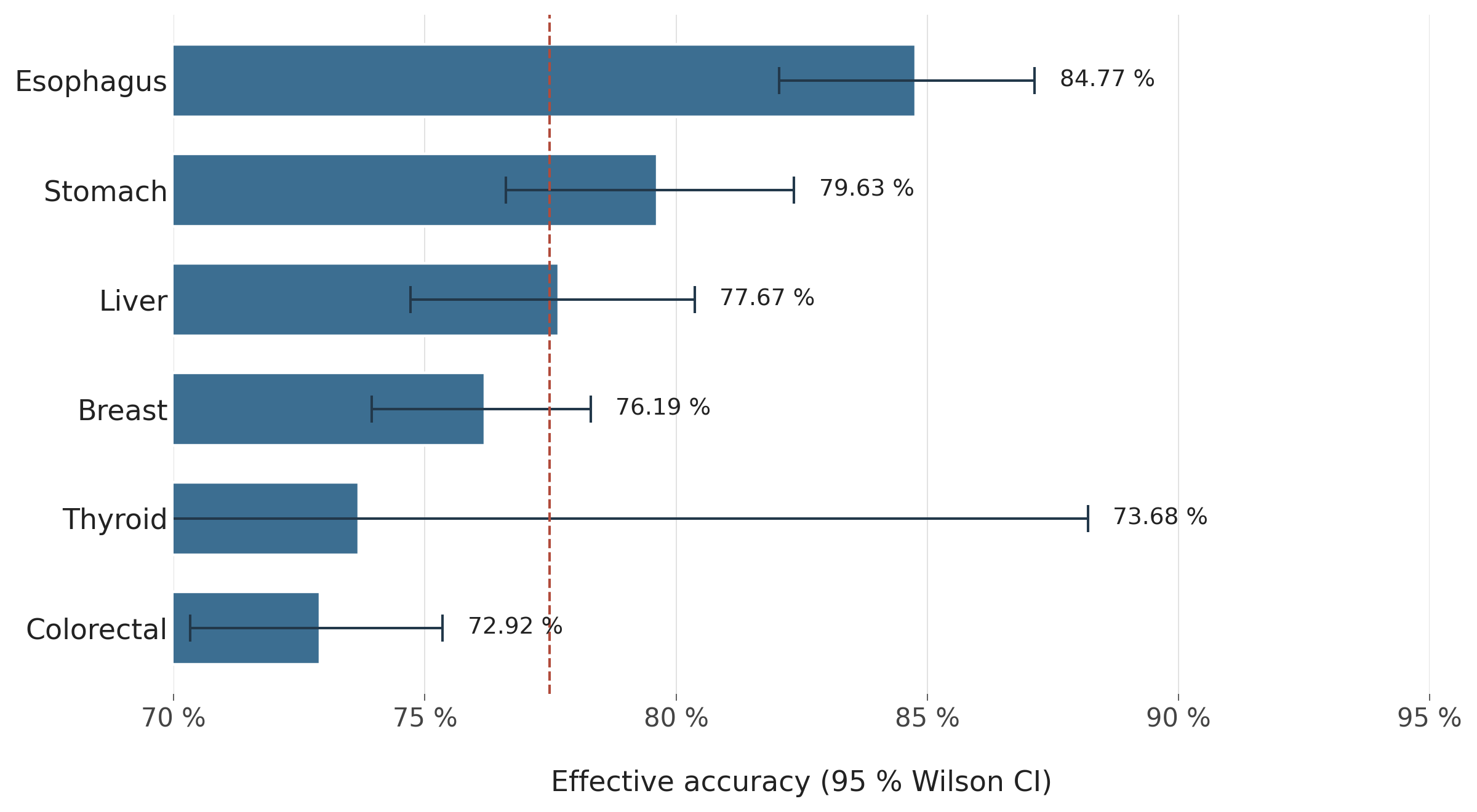

**Figure S6.** *TCGA per-organ Stage-C accuracy with 95 % Wilson CI. Visual companion to Table 15. (§3.9.)*

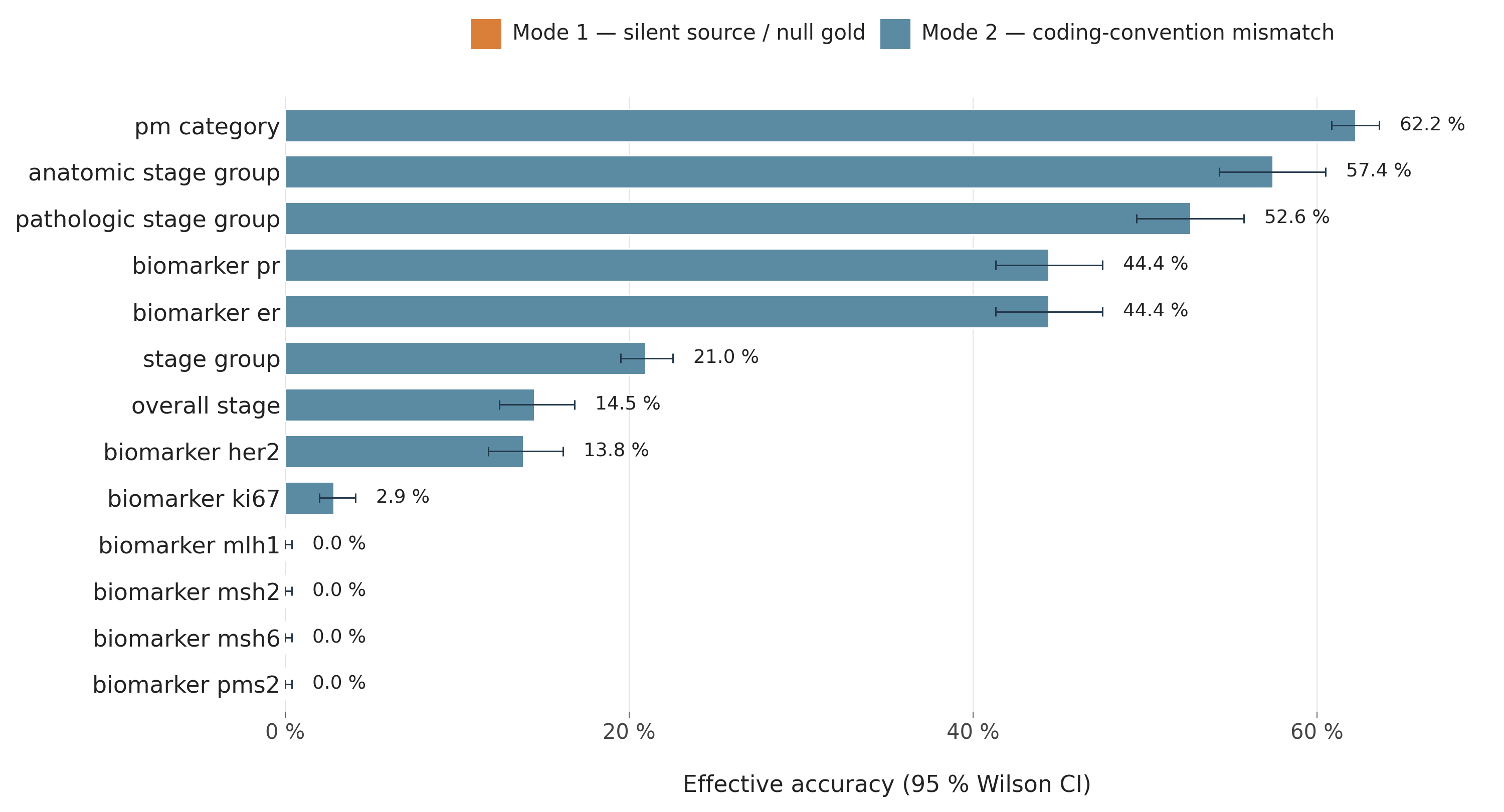

**Figure S7.** *TCGA bottom-decile fields. Effective accuracy with caveat tags for the 13 fields tagged in Supplementary Table S38. (§3.9.)*

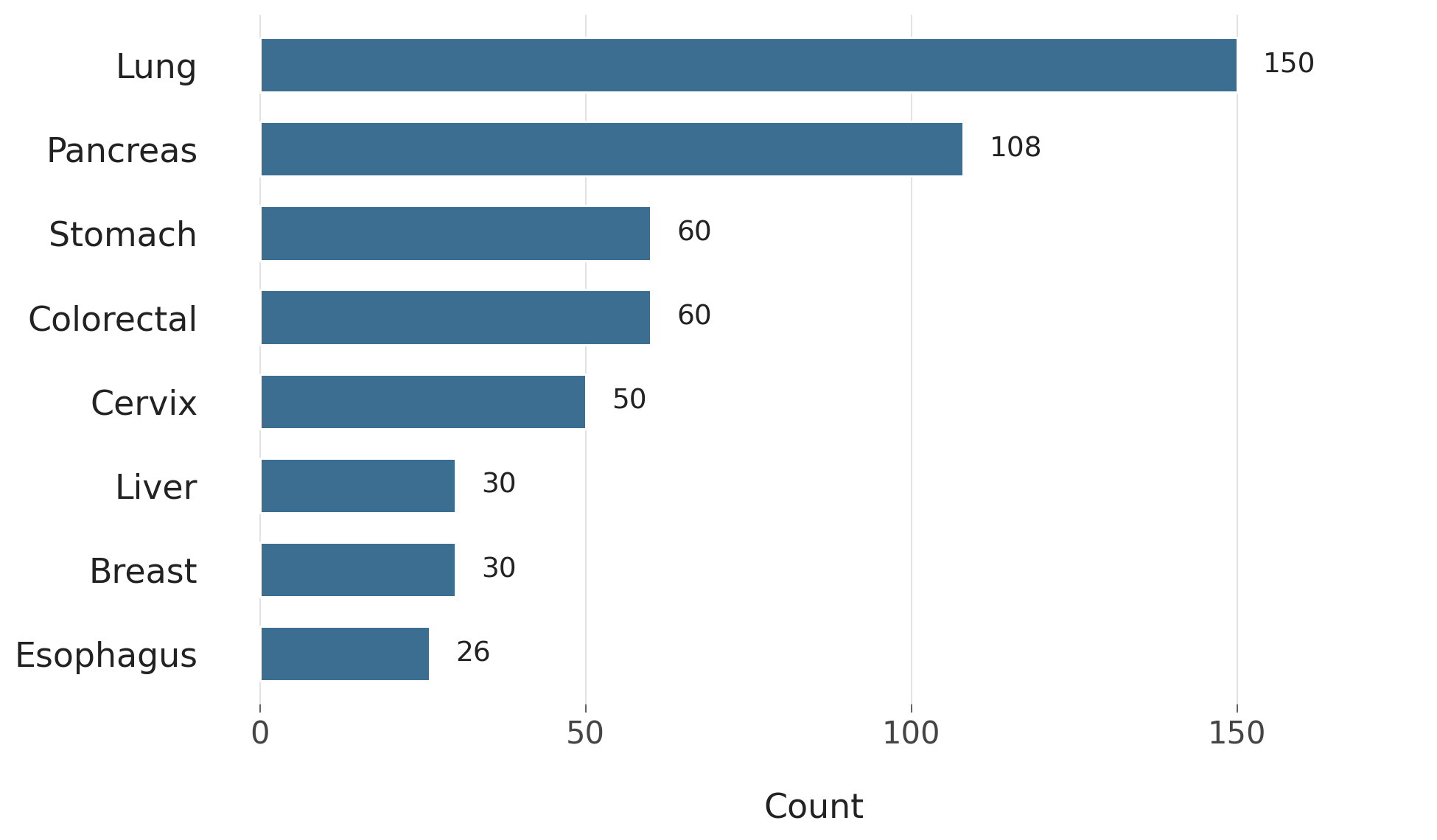

**Figure S8.** *Multi-primary misroute by organ. Companion plot to main-text Figure 2. (§3.2.1.)*

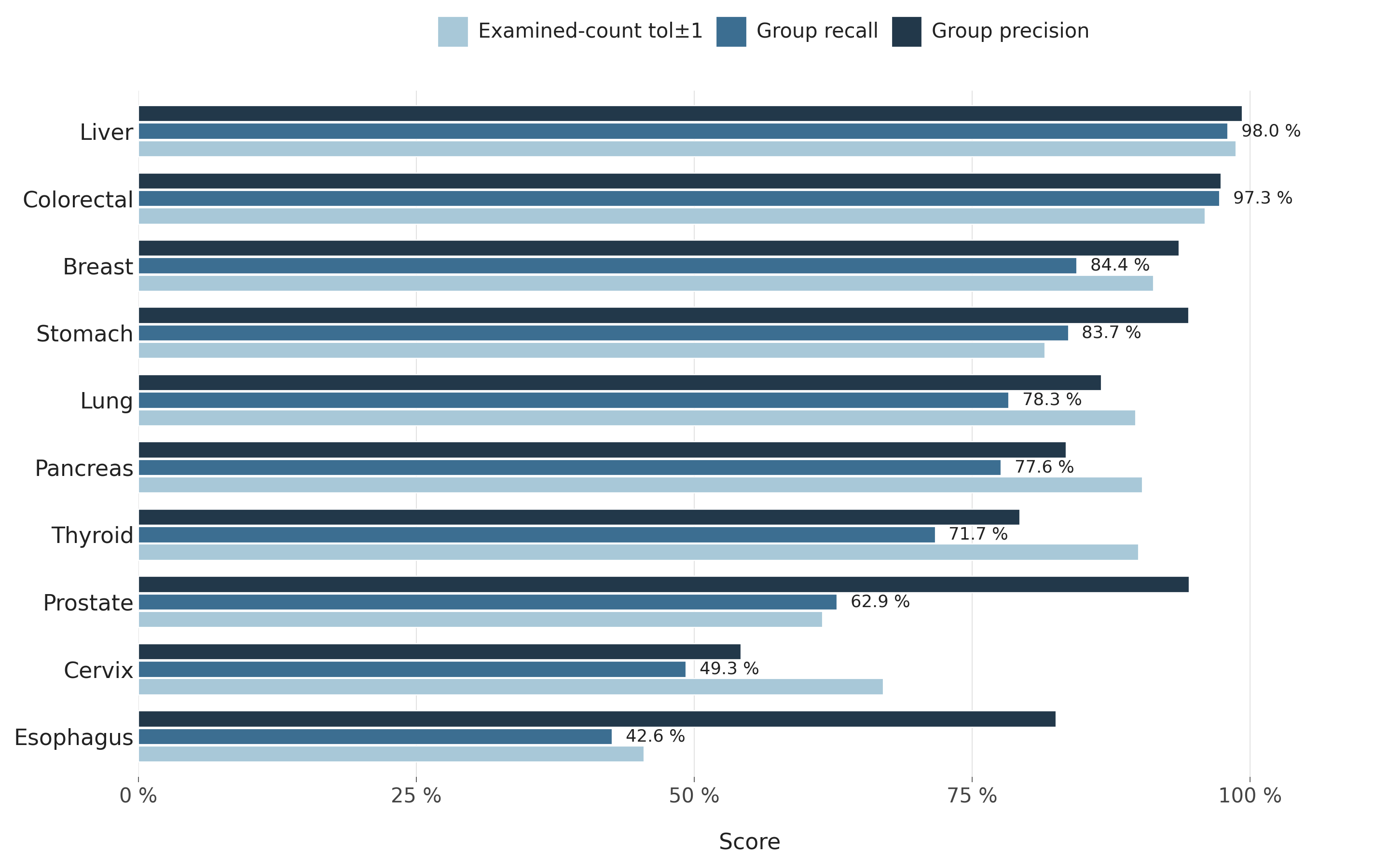

**Figure S9.** *Per-organ lymph-node concordance plus group-recall. Demoted from main body. (§3.4.)*

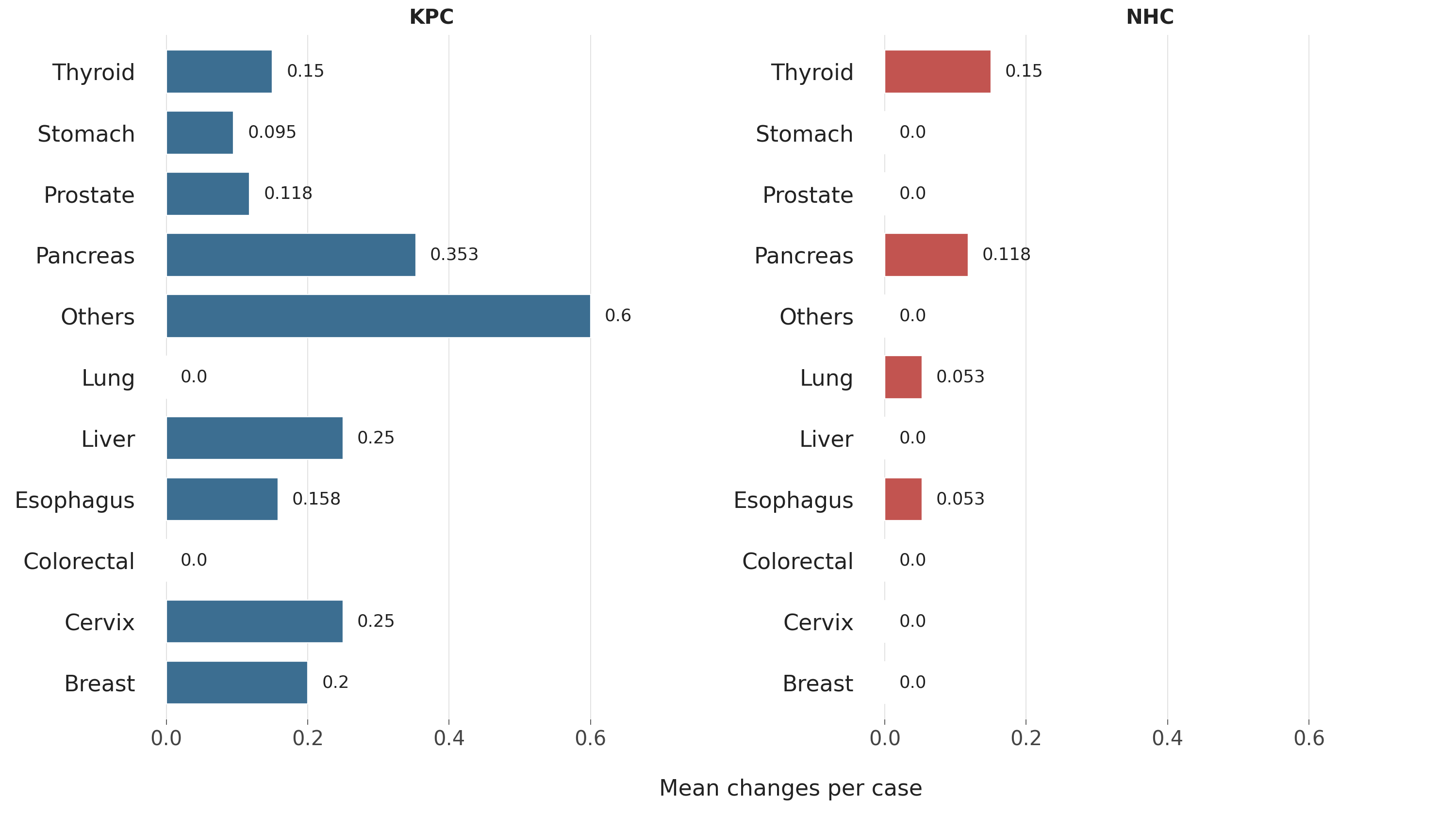

**Figure S10.** *Pre-annotation edit-distance per (annotator × organ). Token-level differences between with- and without-seed arms. (§3.7.)*

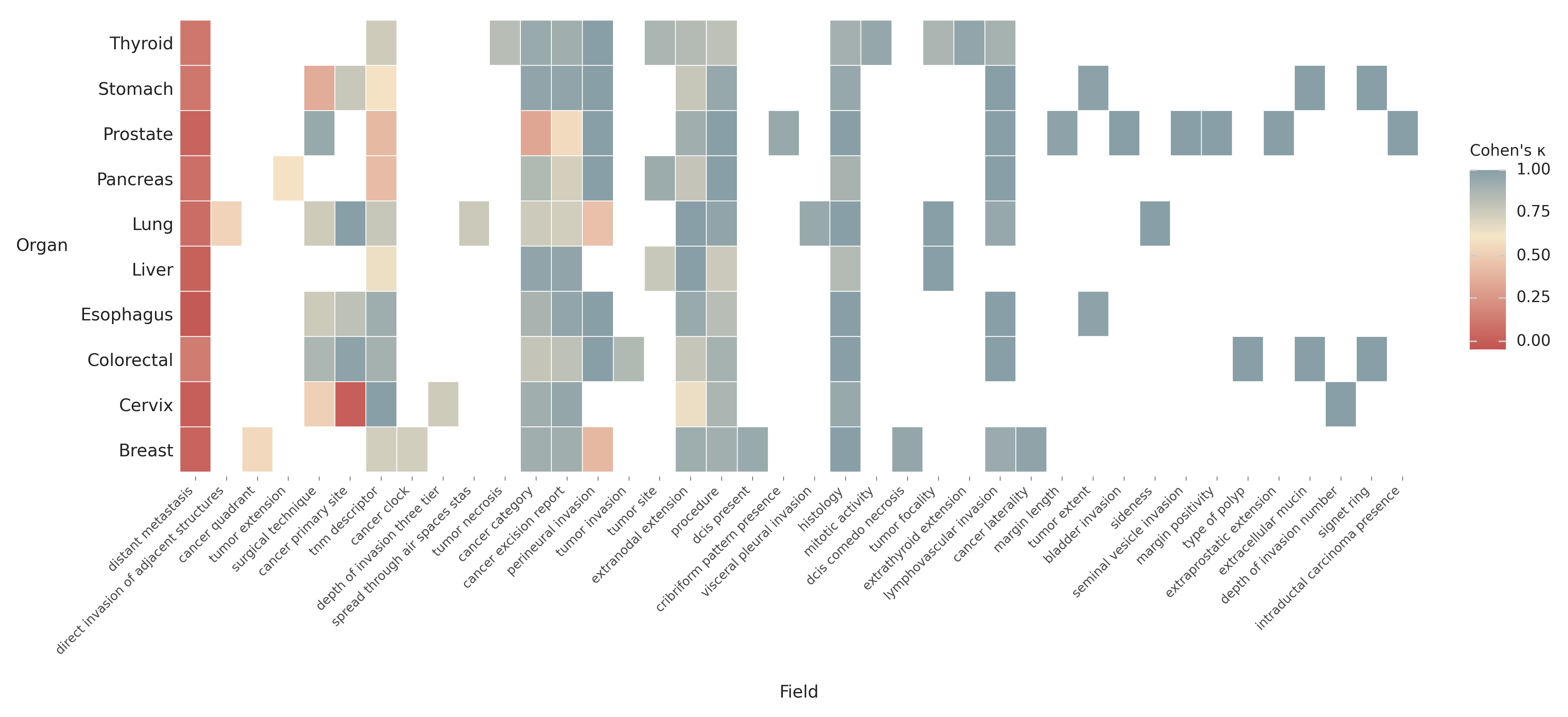

**Figure S11.** *Per-(organ, field) Cohen’s κ heatmap on the operational annotator pair. Micro-detail companion to Figure 7. (§3.7.)*

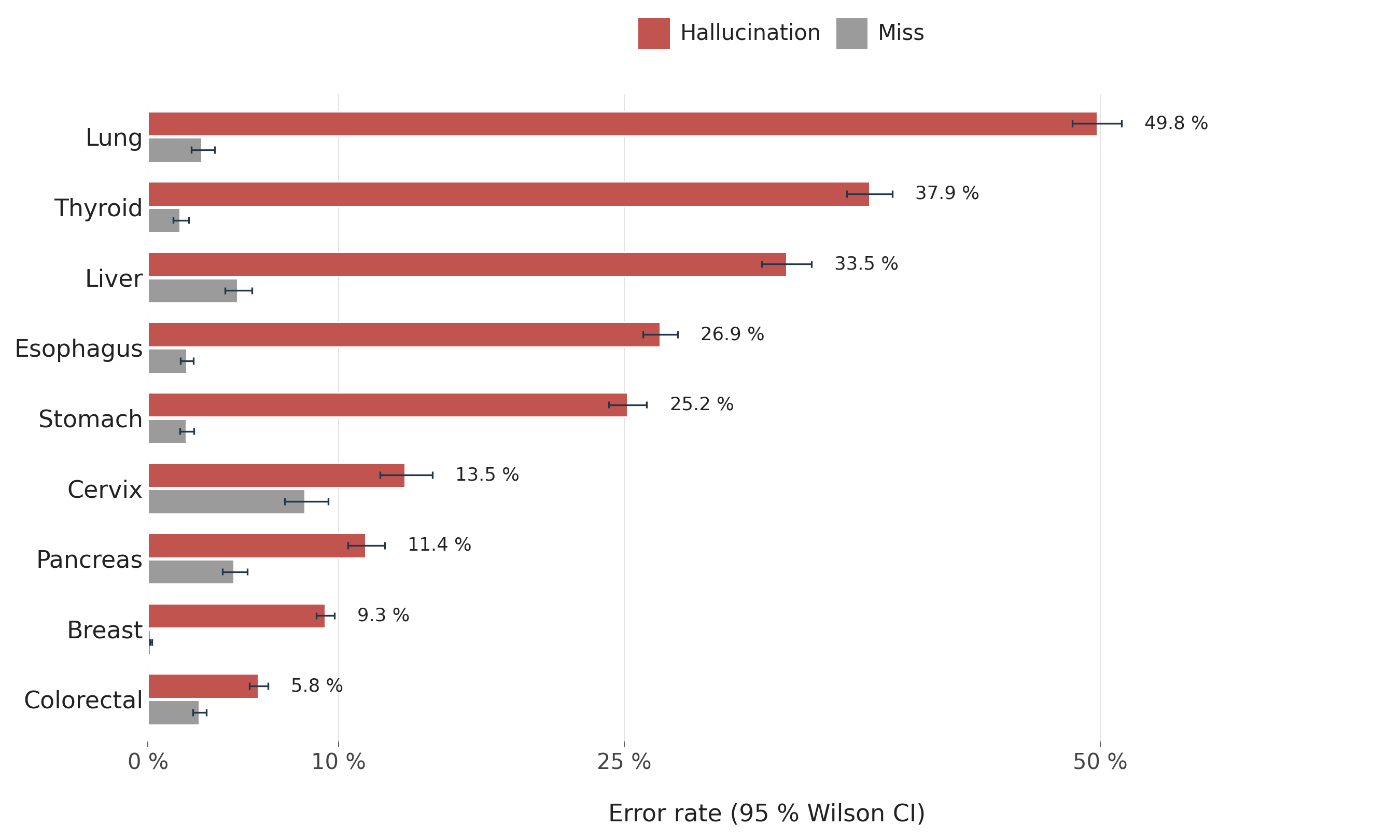

**Figure S12.** *Per-organ margin error decomposition (hallucination vs miss). Demoted from main body. (§3.4.)*

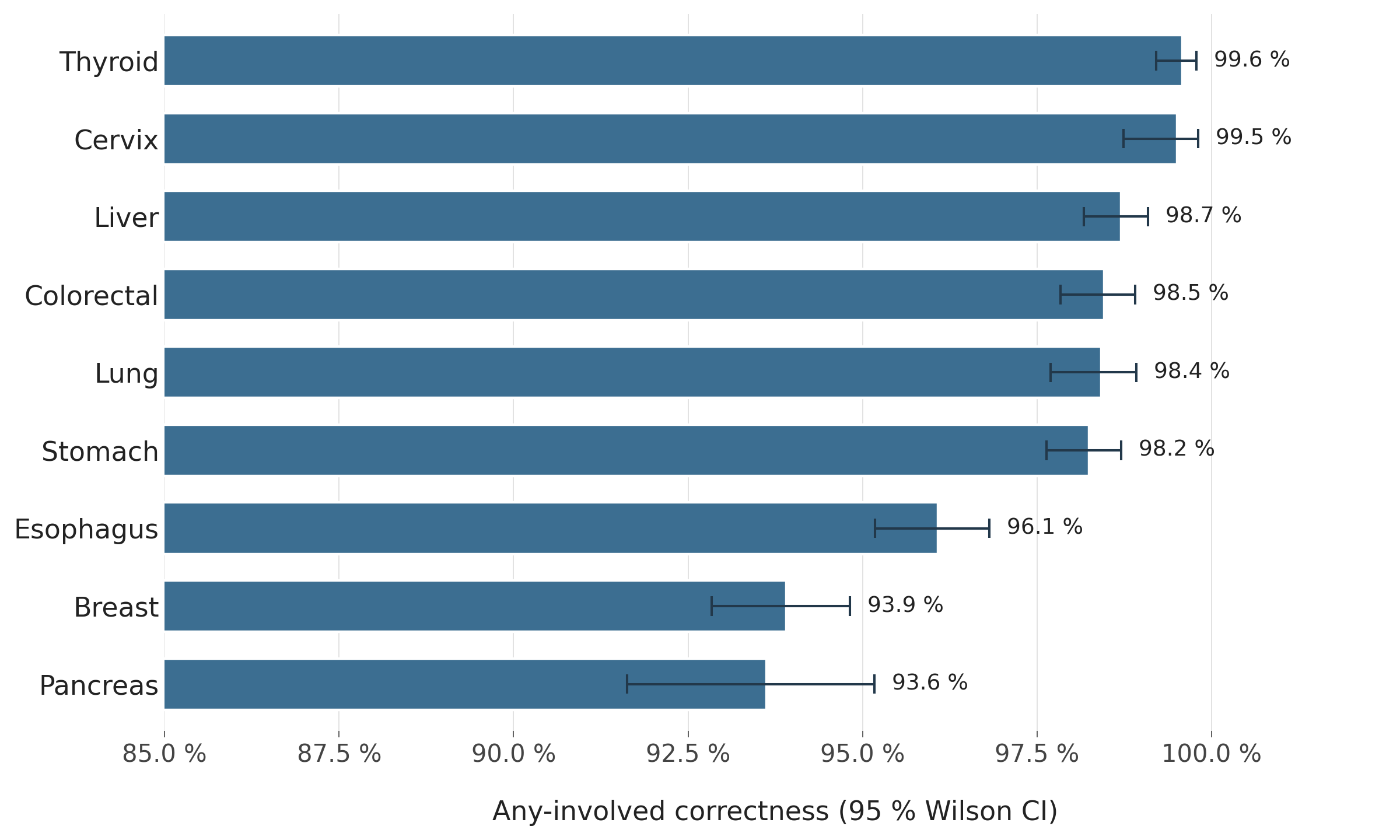

**Figure S13.** *Margin status correctness per organ (any-involved endpoint). Demoted from main body. (§3.4.)*

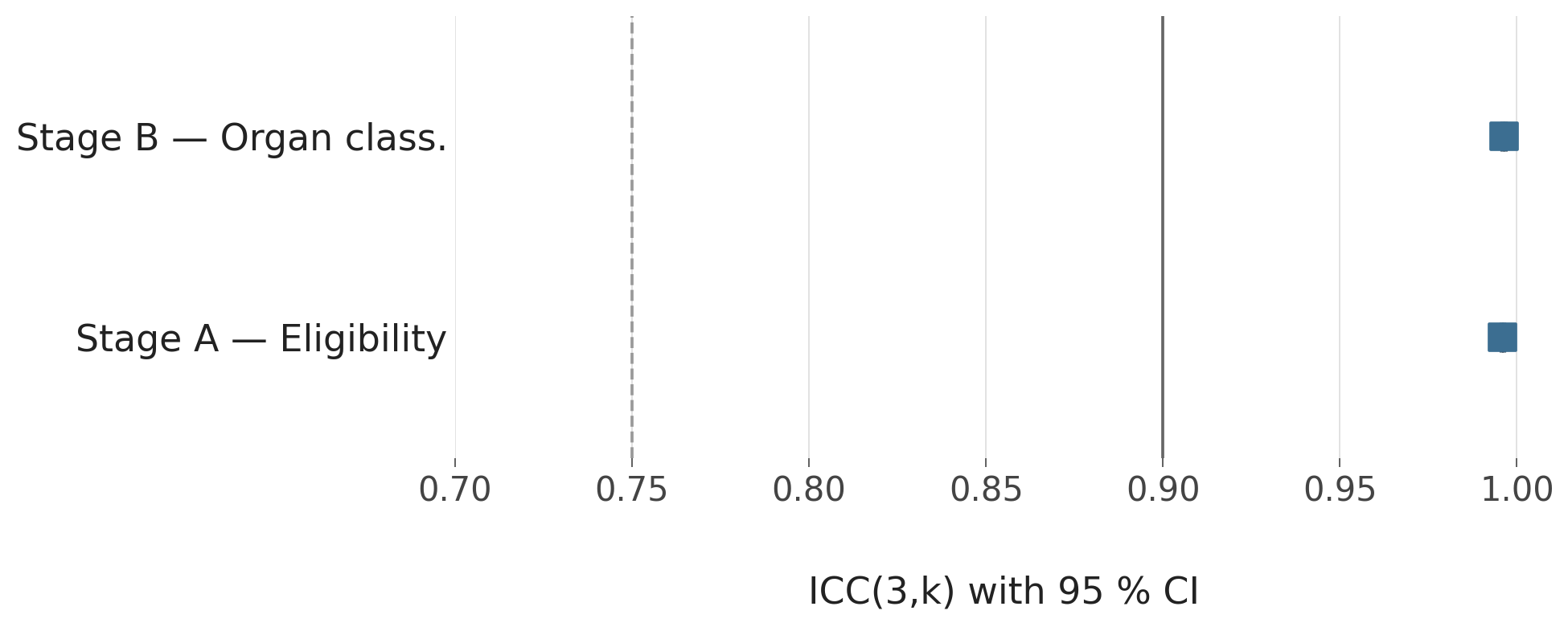

**Figure S14.** *Multi-run reliability ICC(3,k) — eligibility and organ classification. (§3.8.)*

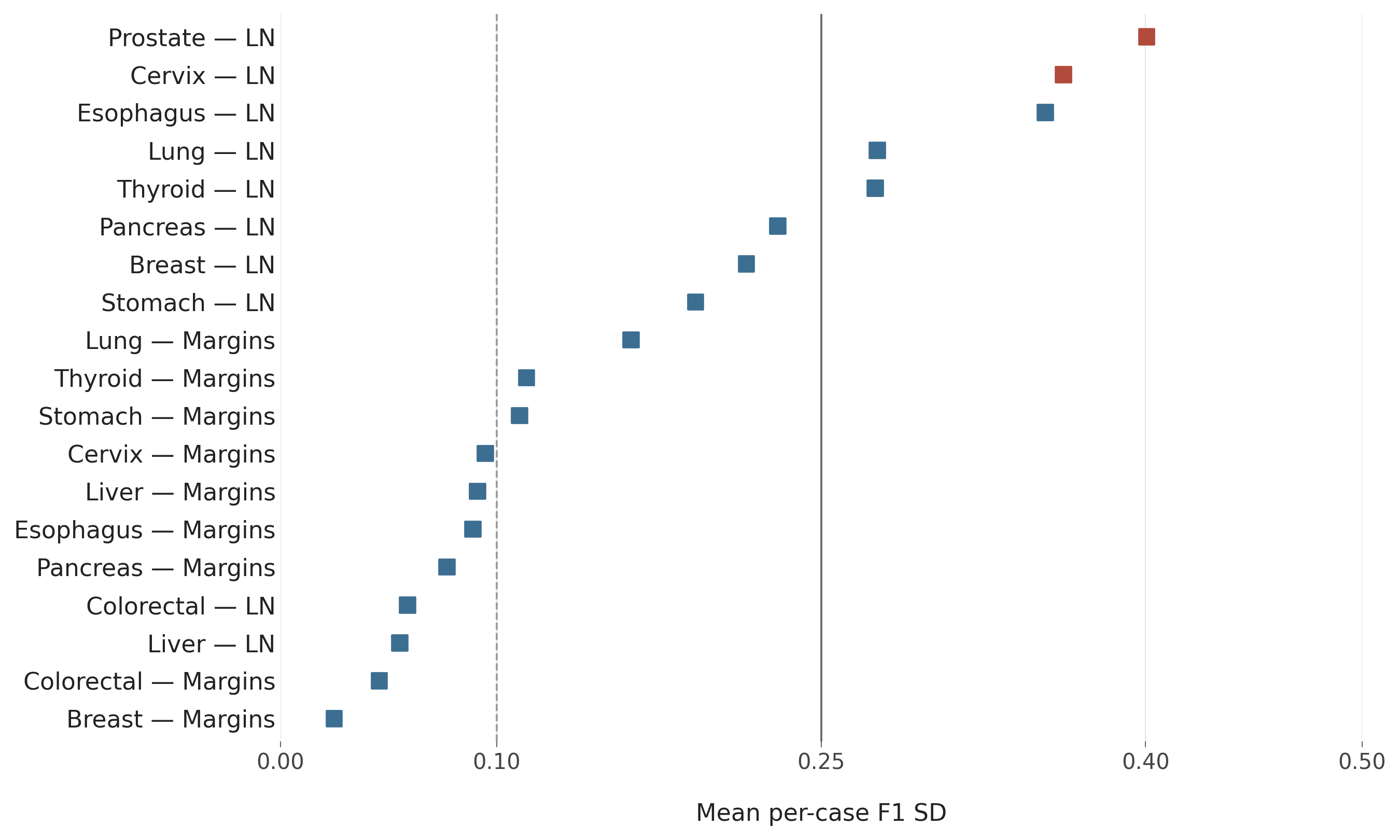

**Figure S15.** *Per-case F1 standard deviation across the 30 multi-seed runs (margins and lymph nodes). (§3.8.)*

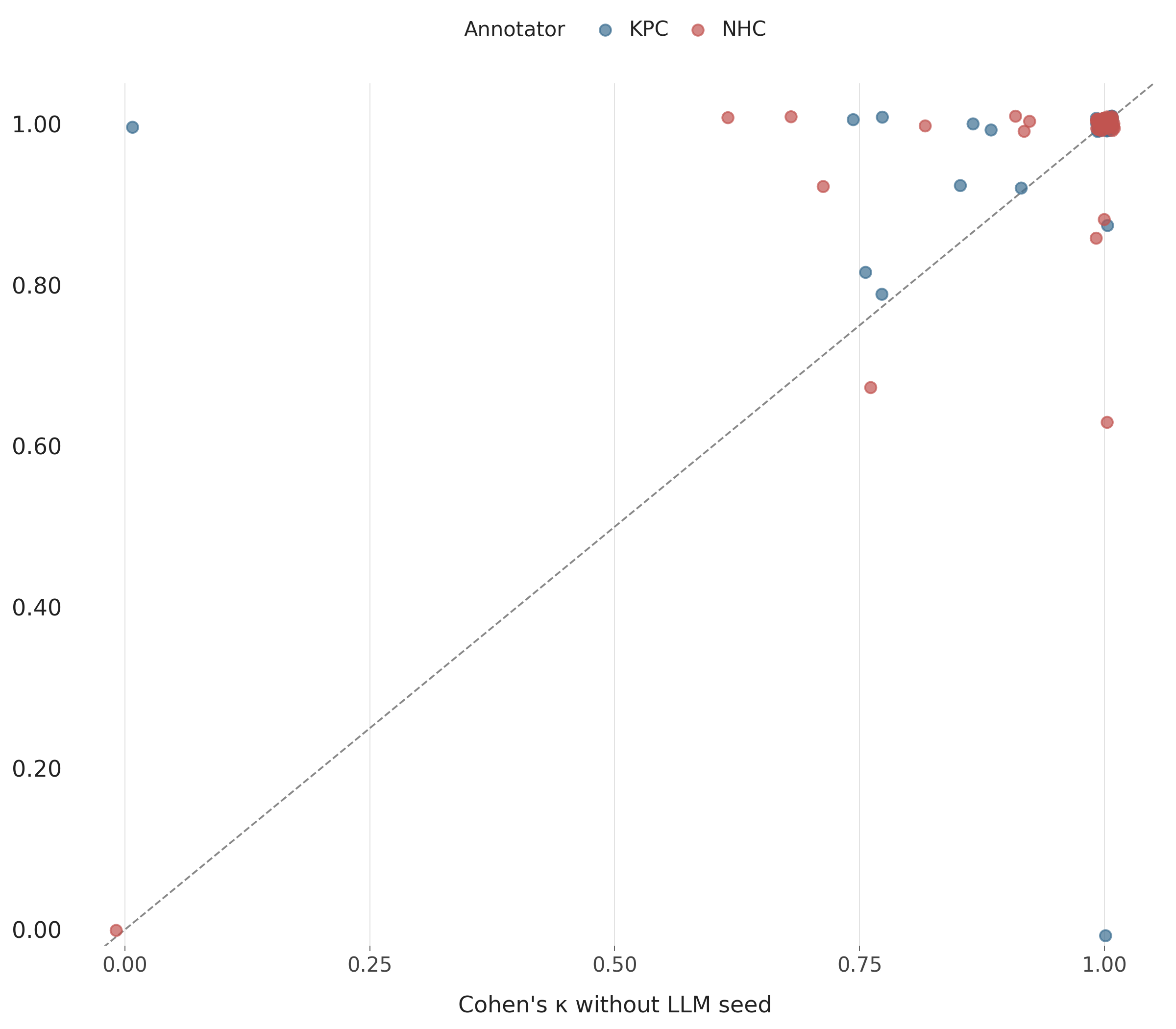

**Figure S16.** *Pre-annotation effect κ_with vs κ_without scatter. (§3.7.)*

**Supplementary Appendices A1–A10**

**Supplementary Appendix A1. Borderline / incomplete report — eligibility classifier flips between runs (breast specimen 2-1-417).**

---

**A1.1 Original pathology report (incomplete draft, excerpt, specimen 2-1-417)**

```
1. Breast, right, status post neoadjuvant chemotherapy, breast conserving surgery, Invasive
   carcinoma of no special type, grade 2/3, residual.
2. Lymph nodes, non-sentinel, axillary, right, sampling, Negative for malignancy and no
   lymphoid tissue is seen.
3. Immunohistochemical study: ER (Negative, less than 1%), PgR (Negative, less than 1%),
   Her2/neu (positive, score 3+) and Ki67 proliferation index (up to 50%).
4. AJCC Prognostic Stage Group: ypT1aN0, cM0, stage group IA (AJCC 8th).

   The specimen fixed in formalin is submitted in two bags and labeled as
     1. Right breast tumor s/p neoadjuvant (two silks: 12 o'clock, one silk: 3 o'clock) and
     2. Right non-SLN.

   The bag 1 …
     A1-12: All for section.
       (A1-2, A3-4, A5-6, A7-8: mapping with peripheral margins;
        A9-10: skin and superficial margin;
        A11-12: base margin.)
     B1: right non sentinel lymph nodes (all for section)

   Microscopic Examination:
     1. Procedure: breast conserving surgery
     2. Lymph Node Sampling:
        Sentinel lymph node biopsy (please refer to pathology report -The report stops here-
```

**A1.2 Model output — Run 1 (interpreted by model as non-eligible)**

```
{
  "cancer_excision_report": false,
  "cancer_category": null,
  "cancer_data": {}
}
```

**A1.3 Model output — Run 2 (excerpt, interpreted by model as eligible)**

```
{
  "cancer_excision_report": true,
  "cancer_category": "breast",
  "cancer_category_others_description": null,
  "cancer_data": {
    "procedure": "breast_conserving_surgery",
    "cancer_quadrant": null,
    "cancer_clock": 12,
    "cancer_laterality": "right",
    "histology": "invasive_carcinoma_no_special_type",
    "tumor_size": null,
    "...": "...",
    "margin_category": "base",
    "margin_involved": false,
    "distance": null,
    "description": "free of tumor grossly",
    "regional_lymph_node": [
      {
        "lymph_node_side": "right",
        "lymph_node_category": "nonsentinel",
        "involved": 0,
        "...": "..."
      }
    ],
    "extranodal_extension": null,
    "maximal_ln_size": null,
    "biomarkers": [
      {
        "biomarker_category": "er",
        "expression": false,
        "percentage": 0,
        "score": null,
        "biomarker_name": "estrogen receptor"
      }
    ]
  }
}
```

**Supplementary Appendix A2. Metastatic disease excision misclassified as primary staging report (colorectal-to-small-bowel specimen 4-2-34).**

---

**A2.1 Original pathology report (excerpt, specimen 4-2-34)**

```
Small intestine, segmental resection, Invasive mucinous adenocarcinoma (colon rectal in
origin). Bilateral cut margins: free of tumor.

The specimen submitted consists of part of small intestine measuring 11.0 cm in length,
fixed in formalin.

Representative sections are taken and labeled as follows:
   A1-2: bilateral margins,
   A3-4: tumor

Microscopically, the sections show a picture of invasive mucinous adenocarcinoma,
revealing dysplastic gland with mucinous pool invasive from serosa to submucosa
ot intestine. Ancillary study of IHC section A3 reveals CDX-2 (+), CK20 (+ focal …
```

**A2.2 Expected output (manual annotation, specimen 4-2-34)**

```
{
  "cancer_excision_report": false,
  "cancer_category": null,
  "note": "excision_metastatic",
  "cancer_category_others_description": null,
  "cancer_data": {}
}
```

**A2.3 Model output (excerpt, specimen 4-2-34)**

```
{
  "cancer_excision_report": true,
  "cancer_category": "colorectal",
  "cancer_category_others_description": null,
  "cancer_data": {
    "procedure": "others",
    "surgical_technique": null,
    "cancer_primary_site": "rectosigmoid_junction",
    "histology": "mucinous_adenocarcinoma",
    "grade": null,
    "tumor_invasion": "visceral_peritoneum_surface",
    "...": "...",
    "pt_category": "t4a",
    "pn_category": "nx",
    "pm_category": "mx",
    "stage_group": null,
    "ajcc_version": null,
    "margins": [
      {
        "margin_category": "proximal",
        "margin_involved": false,
        "distance": null,
        "description": "free of tumor"
      },
      {
        "margin_category": "distal",
        "margin_involved": false,
        "...": "..."
      }
    ]
  }
}
```

**Audit trail**

The same overgeneralization pattern is captured at the population level in the §3.2.1 multi-primary breakdown (Supplementary Tables S30, S31): 631 of the 1,139 false-positive case-runs in the eligibility classifier (cf. paragraph 06 / Supplementary Table S25) are routed to a single in-scope organ when the gold annotation is others (out-of-scope, multi-primary, or metastasis). The flag-for-review workflow proposed in paragraph 20 is the operational response to this systematic overgeneralization.

**Supplementary Appendix A3. Double primary malignancy collapsed to single primary (lung specimen 7-1-294, two lobes).**

---

**A3.1 Original pathology report (excerpt, specimen 7-1-294)**

```
1. Lung, upper lobe, right, VATS lobectomy, adenocarcinoma, moderatedly differentiated,
   acinar predominant.
2. Lung, middle lobe, right, VATS wedge resection, adenocarcinoma, well-differentiated,
   lepidic predominant.
3. Lymph node, group 4, lymphadenectomy, negative for malignancy (0/1).
4. Lymph node, group 7, lymphadenectomy, negative for malignancy (0/4).
5. Lymph node, group 9, lymphadenectomy, negative for malignancy (0/1).
6. Lymph node, group 10, lymphadenectomy, negative for malignancy (0/1).
7. Pathologic staging for right upper lobe tumor: pT2aN0, cM0, stage IB (AJCC 8ed).
8. Pathologic staging for right middle lobe tumor: pT1aN0, cM0, stage IA1 (AJCC 8ed).

   1. Procedure: VATS lobectomy of right upper lung and wedge resection of right middle lung
   2. Specimen Laterality: Right
   3. Tumor Focality: Single focus for each specimen

   Microscopic Examination:
     FOR RIGHT UPPER LOBE
       1. Histologic Type: Invasive acinar adenocarcinoma
          Histologic Patterns Present: Acinar 80 %, lepidic 20 %
       2. Histologic Grade: G2, moderately differentiated
       3. Synchronous Tumors: Total Number of Primary Tumors: 2 (right upper lobe and right middle lobe)
       4. Tumor Size: 2.0 cm
```

**A3.2 Model output (excerpt, specimen 7-1-294)**

```
{
  "cancer_excision_report": true,
  "cancer_category": "lung",
  "cancer_category_others_description": null,
  "cancer_data": {
    "procedure": "lobectomy",
    "surgical_technique": "thoracoscopic",
    "sideness": "right",
    "cancer_primary_site": "upper_lobe",
    "...": "...",
    "treatment_effect": "No known presurgical therapy",
    "tnm_descriptor": null,
    "pt_category": "t2a",
    "pn_category": "n0",
    "pm_category": "mx",
    "stage_group": "ib",
    "ajcc_version": 8,
    "margins": [
      {
        "margin_category": "bronchial"
      }
    ]
  }
```

}.

**Supplementary Appendix A4. Anatomic versus pathologic stage-group ambiguity (breast specimen 2-1-206).**

**A4.1 Original pathology report (excerpt, specimen 2-1-206)**

```
1. Breast, 9 o'clock position / 5 cm from nipple, right, status post neoadjuvant
   chemotherapy, partial mastectomy, Invasive carcinoma of no special type, grade 2.
2. Lymph node, axillary, right, lymphadenectomy, carcinoma, metastatic (2/9).
3. Immunohistochemical study: ER (positive, strong, 99 %), PgR (positive, weak, 5 %),
   Her2/neu (Equivocal, score 2+) and Ki67 proliferation index (up to 3 %).
4. Pathologic AJCC Stage Group: ypT1c N1a, cM0
   (Anatomic stage group IIA, AJCC 8ed, combined with report of pathology number
    2024-35741).

   1. Procedure: Partial mastectomy
   …
```

**A4.2 Model output (excerpt, specimen 2-1-206)**

```
{
  "cancer_data": {
    "procedure": "partial_mastectomy",
    "cancer_quadrant": "lower_outer_quadrant",
    "cancer_clock": 9,
    "cancer_laterality": "right",
    "histology": "invasive_carcinoma_no_special_type",
    "tumor_size": 19,
    "treatment_effect": "Partial, Definite response",
    "...": "...",
    "grade": 2,
    "tnm_descriptor": "y",
    "pt_category": "t1c",
    "pn_category": "n1a",
    "pm_category": "mx",
    "pathologic_stage_group": "iib",
    "anatomic_stage_group": "iia"
  }
}
```

**Supplementary Appendix A5. Surgical-technique misclassification — "3D" treated as `open` (stomach specimen 9-1-78).**

---

**A5.1 Original pathology report (excerpt, specimen 9-1-78)**

```
1. Stomach, anastomosis of B-II, status post subtotal gastrectomy with B-II reconstruction,
   3D radical total gastrectomy,
     (1) large cell neuroendocrine carcinoma,
     (2) high-grade glandular dysplasia and
     (3) chronic gastritis and intestinal metaplasia.
2. Small intestine, anastomosis of B-II, 3D radical total gastrectomy,
   large cell neuroendocrine carcinoma, involved.
   …
10. Pathologic staging: pT3, pN0, cM0, stage IIA (AJCC, 8th edition).

   The specimen fixed in formalin is submitted in six separated bags labeled as
     1: stomach, 2: lymph node (LN) 1, 3: LN2, 4: LN3, 5: LN4, 6: LN789.

   1. Procedure: 3D radical total gastrectomy
   2. Specimen:
      Stomach: 11.5 × 6.5 × 1.8 cm in size …
```

**A5.2 Model output (excerpt, specimen 2-1-206)**

```
{
  "cancer_data": {
    "procedure": "total_gastrectomy",
    "surgical_technique": "open",
    "cancer_primary_site": "pylorus",
    "histology": "others",
    "grade": null,
    "tumor_extent": "penetrate_subserosal_connective_tissue_no_serosa",
    "...": "...",
    "pt_category": "t3",
    "pn_category": "n0"
  }
}
```

**Supplementary Appendix A6. Lymph-node extraction in esophageal cancer (specimens 5-1-5 success and 5-1-3 partial omission).**

**A6.1 Successful LN extraction (specimen 5-1-5)**

**A6.1.1 Original pathology report (excerpt, specimen 5-1-5)**

```
1. Esophagus, EG junction, thoracoscopic esophagectomy, status post CCRT,
   squamous cell carcinoma, well differentiated, residual.
   …
6. Lymph node, regional, lymphadenectomy, negative of malignancy (0/1).
7. Lymph node, group 7, lymphadenectomy, negative of malignancy (0/1).
8. Soft tissue, labeled as "LN 9L", excision, negative of malignancy (0).
9. Lymph node, group 10L, lymphadenectomy, negative of malignancy (0/1).
10. Pathologic Stage: ypT3 N0, cM0, stage IIB (AJCC, 8th edition).

   The specimen fixed in formalin is submitted in seven separated bags, labeled as
     1: esophagus, 2: stomach + liver, 3: LN7, 4: LN 9L, 5: LN 10L,
     6: anastomosis ring, and 7: gastric tube stump, respectively.
   Procedure / Specimen Type: Ivor Lewis procedure
   …
   F1-2: anastomosis ring (all for section)
   G1-2: gastric tube stump

   Microscopic Examination:
     1. Tumor Site: EG junction
     2. Histologic Type: squamous cell carcinoma
     3. Histologic Grade: G1, well differentiated
     …
     10. Regional Lymph Node Status: All lymph nodes are negative of malignancy
         Number of Lymph Nodes Involved/Examined: 0/3
         Nodal tumor size: not applicable
         Extracapsular extension: not applicable
     11. Pathologic Stage (pTNM, AJCC 8th Edition): …
```

**A6.1.2 Model output (excerpt, specimen 5-1-5)**

```
{
  "cancer_data": {
    "procedure": "esophagogastrectomy",
    "surgical_technique": "thoracoscopic",
    "cancer_primary_site": "gastroesophageal_junction",
    "...": "...",
    "tnm_descriptor": "y",
    "pt_category": "t3",
    "pn_category": "n0",
    "pm_category": "mx",
    "stage_group": "iib",
    "ajcc_version": 8,
    "margins": [],
    "regional_lymph_node": [
      {
        "lymph_node_category": "thoracic_7",
        "involved": 0,
        "examined": 1,
        "station_name": null
      },
      {
        "lymph_node_category": "thoracic_9l",
        "involved": 0,
        "examined": 1,
        "station_name": null
      }
    ]
  }
}
```

---

**A6.2 Failed LN extraction with partial station omission (specimen 5-1-3)**

**A6.2.1 Original pathology report (excerpt, specimen 5-1-3)**

```
1. Esophagus, lower third, thoracoscopic esophagectomy and gastric tube pullup
   reconstruction, Squamous cell carcinoma, moderately differentiated.
2. Stomach, cardia, thoracoscopic esophagectomy and gastric tube pullup reconstruction,
   Chronic active gastritis. Free of tumor.
   …
5. Lymph node, perigastric, lymphadenectomy, Negative of malignancy (0/8).
6. Lymph node, group 7, lymphadenectomy, Negative of malignancy (0/4).
7. Lymph node, group 8, lymphadenectomy, Negative of malignancy (0/4).
8. Lymph node, group 9, lymphadenectomy, Negative of malignancy (0/1).
9. Soft tissue, labeled as "group 9L", excision, Negative of malignancy. No lymph node is found.
10. Soft tissue, labeled as "group 10", excision, Negative of malignancy. No lymph node is found.
11. Lymph node, group 10L, lymphadenectomy, Negative of malignancy (0/2).
12. Lymph node, left gastric, lymphadenectomy, Squamous cell carcinoma, metastatic (1/1).
13. Pathologic Stage: pT1a, N1, cM0, stage IIB (AJCC, 8th edition).

   The specimen fixed in formalin is submitted in ten separated bags, labeled as
     1: esophagus and partial stomach, 2: esophageal proximal cut end, 3: anastomosis,
     4: LN 7, 5: LN 8, 6: LN 9, 7: LN 9L, 8: LN 10, 9: LN 10L, and 10: left gastric LN,
     respectively.

   Microscopic Examination:
     1. Tumor Site: Lower third esophagus
     2. Histologic Type: Squamous cell carcinoma
     …
     9. Perineural Invasion: Not identified
     10. Regional Lymph Node Status:
         Number of Lymph Nodes Involved/Examined: 1/20
         Nodal tumor size: 1.0 cm
         Extracapsular extension: Not identified
```

**A6.2.2 Model output (excerpt, specimen 5-1-5)**

```
{
  "cancer_data": {
    "procedure": "esophagectomy",
    "surgical_technique": "thoracoscopic",
    "cancer_primary_site": "lower_third",
    "histology": "squamous_cell_carcinoma",
    "grade": 2,
    "...": "...",
    "pt_category": "t1a",
    "pn_category": "n1",
    "pm_category": "mx",
    "stage_group": "iib",
    "ajcc_version": 8,
    "margins": [
      {
        "margin_category": "proximal"
      }
    ],
    "regional_lymph_node": [
      {
        "lymph_node_category": "thoracic_7",
        "involved": 0,
        "examined": 1,
        "station_name": null
      }
    ],
    "regional_lymph_node": [
      {
        "lymph_node_category": "regional_gastric",
        "involved": 1,
        "examined": 1,
        "station_name": "left gastric"
      }
    ],
    "extranodal_extension": false,
    "maximal_ln_size": 10
  }
}
```

**Supplementary Appendix A7. Margin-status over-population from schema enumeration (pancreas Whipple specimen 1-1-14).**

**A7.1 Original pathology report (excerpt, specimen 1-1-14)**

```
1. Pancreas, head, status post neoadjuvant chemotherapy, pancreatoduodenectomy,
   Ductal adenocarcinoma, moderately differentiated.
2. Stomach, lower body and antrum, pancreatoduodenectomy, Free of tumor.
   …
14. Lymph node, group 16, lymphadenectomy, Negative for malignancy (0/3).
15. Liver capsule, excision, Simple cyst with dystrophic calcification.
16. Pancreas margin, excision, Free of tumor.
17. Pathologic staging: ypT2 pN1, cM1, stage IV (AJCC, 8th edition).

   The specimen fixed in formalin is submitted in two separated bags labeled as
     1: pancreaticoduodenectomy product, 2: group 8 lymph nodes (LN), 3: group 12 LNs,
     4: group 13 LNs, 5: group 14 LNs, 6: group 16 LNs, 7: liver capsule,
     and 8: pancreas margin, respectively.

   Procedure: Pancreaticoduodenectomy.
   …
   6. Macroscopic margin status:
      Pancreatic neck/parenchymal margin: Free of tumor, with the distance of 1.0 cm.
      Common bile duct margin: Free of tumor, with the distance of 2.5 cm.
      Proximal (gastric or duodenal) margin: Free of tumor, with the distance of 10 cm.
      Distal (duodenal or jejunal) margin: Free of tumor, with the distance of 23 cm.
      Anterior outmost margin of pancreas: Free of tumor, with the distance of 0.1 cm.
      Posterior outmost margin of pancreas: Free of tumor, with the distance of 0.1 cm.
      Uncinate (retroperitoneal/superior mesenteric artery) margin: Free of tumor,
        with the distance of 0.1 cm.
      SMV groove margin: Free of tumor, with the distance of 0.1 cm.
   7. Non-tumorous part: Fibrosis.

   Microscopic Examination:
     1. Histologic Type: Ductal Adenocarcinoma.
     2. Histologic Grade (ductal carcinoma only): G2, moderately differentiated.
     …
     7. Perineural Invasion: Present.
     8. Margin:
        a. Proximal pancreatic parenchymal margin: Not applicable.
        b. Distal pancreatic parenchymal margin: Not applicable.
        c. Pancreatic neck/parenchymal margin: Involved by invasive carcinoma in
           specimen 1 (section A11), but free of tumor in the separated margin
           (section H1 and frozen section 2024-31509).
        d. Uncinate (retroperitoneal/superior mesenteric artery) margin:
           Involved by Invasive carcinoma (section A19).
        e. Bile duct margin: Greater than 1.0 cm/10.0 mm in distance from invasive
           carcinoma to closest margin (section A5).
        f. Proximal (gastric or duodenal) margin: Greater than 1.0 cm/10.0 mm in distance
           from invasive carcinoma to closest margin.
        g. Distal (duodenal or jejunal) margin: Greater than 1.0 cm/10.0 mm in distance
           from invasive carcinoma to closest margin.
        h. Other (specify) margin: Posterior surface involved by invasive carcinoma
           (section A15).
     9. Regional Lymph Node Status:
        *Specify (Lymph Node Involved/Examination):
           regional (2/2), peri-intestinal (0/0), peri-gastric (0/0),
           group 8 (0/1), group 12 (0/2), group 13 (0/5), group 14 (0/3), group 16 (0/3).
     11. Pathologic Stage (pTNM, AJCC 8th Edition): …
```

**A7.2 Model output (excerpt, specimen 1-1-14)**

```
{
  "cancer_data": {
    "procedure": "whipple_procedure",
    "tumor_site": "head",
    "histology": "ductal_adenocarcinoma_nos",
    "tumor_size": 25,
    "...": "...",
    "tnm_descriptor": "y",
    "pt_category": "t2",
    "pn_category": "n1",
    "pm_category": "mx",
    "overall_stage": "iv",
    "ajcc_version": 8,
    "margins": [
      {
        "margin_category": "pancreatic_neck",
        "margin_involved": false,
        "distance": 10,
        "description": "Free of tumor"
      },
      {
        "margin_category": "uncinate",
        "margin_involved": true,
        "distance": 0,
        "description": "Involved by invasive carcinoma"
      },
      {
        "margin_category": "bile_duct",
        "margin_involved": false,
        "distance": 10,
        "description": "Free of tumor"
      },
      {
        "margin_category": "proximal_gastric",
        "margin_involved": false,
        "distance": 10,
        "description": "Free of tumor"
      },
      {
        "margin_category": "proximal_duodenal",
        "margin_involved": false,
        "distance": 10,
        "description": "Free of tumor"
      },
      {
        "margin_category": "distal_intestinal",
        "margin_involved": false,
        "distance": 10,
        "description": "Free of tumor"
      },
      {
        "margin_category": "anterior_outmost",
        "margin_involved": false,
        "distance": 1,
        "description": "Free of tumor"
      },
      {
        "margin_category": "posterior_outmost",
        "margin_involved": false,
        "distance": 1,
        "description": "Free of tumor"
      },
      {
        "margin_category": "outmost",
        "margin_involved": false,
        "distance": 1,
        "description": "Free of tumor"
      },
      {
        "margin_category": "others",
        "margin_involved": true,
        "distance": 0,
        "description": "Involved by invasive carcinoma"
      }
    ],
    "regional_lymph_node": [
      {
        "lymph_node_category": "regional_pancreatic",
        "involved": 2,
        "examined": 2,
        "station_name": null
      }
    ],
    "regional_lymph_node": [
      {
        "lymph_node_category": "thoracic_7",
        "involved": 0,
        "examined": 1,
        "station_name": null
      },
      {
        "lymph_node_category": "thoracic_9l",
        "involved": 0,
        "examined": 1,
        "station_name": null
      }
    ]
  }
}
```

**Supplementary Appendix A8. TNM staging-basis: three structural dimensions of the anatomic-vs-pathologic stage_group gap, with two complementary mitigations.**

---

**A8.1 Headline observation**

On the same 2,248 paired breast case-runs (§3.3.1), anatomic_stage_group was extracted at 97.95 % [97.28, 98.46] effective accuracy whereas pathologic_stage_group reached only 74.87 % [73.03, 76.62] — a 23.09 percentage-point gap whose 95 % Wilson intervals do not overlap. Cohen's κ remained at 0.93 against the gold for pathologic_stage_group, ruling out random-disagreement noise as the primary driver. On re-analysis the gap is *not* primarily a model-disambiguation failure but decomposes into three structurally distinct dimensions, each anchored in AJCC 8 breast-staging conventions (Amin et al., AJCC 8th edition).

**A8.2 Dimension 1 — Source-availability gap for post-treatment (`y`-descriptor) cases**

AJCC 8 directs the clinician toward a *clinical* prognostic group for y cases — a stage a pathology report cannot issue. Following clinicopathologic discussion at our institute during the AJCC 8 adoption period, post-chemotherapy specimens were reported with anatomic_stage_group only; later reports include both stage groups. For the contiguous band of y-descriptor cases that print anatomic_stage_group alone, the registry-curated gold carries a pathologic_stage_group value inferred by the annotator from T, N, M, biomarker status, and the AJCC 8 stage table — information not present in the source narrative. The extraction model has no access to the staging table and correctly emits what the report contains.

**A8.3 Dimension 2 — HER-2 2+ equivocal indeterminacy**

AJCC 8 prognostic staging requires definitive HER-2 status; when immunohistochemistry returns HER-2 2+, fluorescence in-situ hybridisation (FISH) is required and the prognostic stage cannot be finalised from the pathology report alone. The true pathologic_stage_group for these cases takes one of two values depending on FISH amplification; a single-valued field prediction is structurally unable to match the gold on either side of the FISH outcome.

**A8.4 Dimension 3 — Residual narrative ambiguity**

A smaller residual of genuine narrative ambiguity remains in non-y reports that cite T, N, and M descriptors without an explicit "prognostic" or "anatomic" qualifier, where the model defaults to anatomic staging.

**A8.5 Two complementary structural mitigations**

Both mitigations are flagged for a follow-up revision because they require schema changes plus re-running the full extraction grid against the modified schema.

**1. AJCC 8 stage-table injection (addresses Dimensions 1 and 3).** Extend the DSPy schema so the model is provided with the AJCC 8 prognostic stage table and asked to *derive* pathologic_stage_group from anatomic_stage_group + ER + PgR + HER-2 + TNM descriptor, rather than echoing what the source narrative happens to print. This collapses the annotator-inferred-gold-vs-source-only-model gap by giving the model the same lookup the annotator used.

**2. Cardinality extension of `pathologic_stage_group` (addresses Dimension 2).** Widen pathologic_stage_group from a scalar to a list of candidate prognostic stages when HER-2 is reported as 2+ pending FISH. This mirrors the cardinality-extension pattern separately proposed in R2.2 for synchronous multi-primaries (widening cancer_data to a list of CancerEntity records): the type structure is unchanged; only the cardinality changes from one to many, and the hierarchical JSON namespace already accommodates this shape.

The two mitigations are complementary rather than alternative: the stage-table lookup recovers values the model cannot derive from the source narrative alone; the cardinality extension represents indeterminacy honestly when a single value is structurally insufficient. Both are amenable to ablation testing within an extended DSPy framework and are flagged as next-paper scope.

**Supplementary Appendix A9. Multi-primary handling: three-axis future-work plan + cardinality-induced conflation caveat.**

**A9.1 Headline observation**

The current cancer_data schema models a single primary tumor per report — a deliberate choice that reflects the registry-grade target of one extraction record per primary, but that creates a structural blind spot for synchronous multi-primary cases (e.g., bilateral breast carcinoma, multifocal lung primaries, multifoci hypopharynx–esophagus). The Stage-B triage analysis in §3.2.1 quantified this directly: of 1,138 gold-"others" case-runs across the thirty multi-seed runs, 449 (39.5 %) were correctly triaged to the "others" class, while 631 (55.4 %) were misrouted to a single in-scope organ — 180 of these (28.5 %) being synchronous multi-primaries within an in-scope organ system, 163 (25.8 %) being out-of-scope sites that the model approximated to a neighboring organ, and 288 (45.6 %) being free-text descriptions the regex could not confidently classify into either category.

**A9.2 Three complementary future-work axes**

One axis is deployable in the current revision; two require dedicated follow-up effort.

• **Axis 1 — Flag-for-review workflow (current revision).** Cases meeting any of the following triggers are routed to manual registrar reconciliation rather than auto-committed: tumor_focality ∈ {multifocal, multicentric}, cancer_laterality = "bilateral", an existing detect_multi_primary() heuristic returning true (which already captures focality, laterality, multi-quadrant, multi-clock, and double-primary regex patterns; the module is part of the public release), or an explicit cancer_category = "others" triage call. This converts a silent misclassification into a human-readable flag and preserves the safety property required of an automated registry pipeline. The flag-for-review trigger requires no schema changes and can be deployed immediately on top of the existing pipeline.

• **Axis 2 — Schema cardinality extension (proposed for the next revision, not the current one).** The natural longer-term fix is to widen cancer_data from a single dictionary to a list of CancerEntity records, each carrying its own tumor_focality, cancer_laterality, tnm, and biomarker submodules. The hierarchical JSON namespace already accommodates this; the change is in *cardinality*, not in *type structure*. We do not include this extension in the current revision because it requires re-running the full extraction grid against the multi-entity schema and re-validating the gold standard for the affected case-runs — work that is appropriate for a follow-up paper on synchronous-primary registry coverage rather than an in-revision change.

• **Axis 3 — Detection-stage improvements (follow-up paper).** The deterministic detect_multi_primary() route is robust at a single institute but does not generalize across reporting conventions. Two complementary detectors are sketched. First, a prototype secondary_cancer_category field on the eligibility signature — drafted in the public release (common_v2.py) but unevaluated within the revision timeframe — would let the generative model emit a probabilistic "is there a second primary?" signal directly; whether an entropy-rich language model is the right substrate for a low-cardinality binary decision is itself the empirical question. Second, a discriminative BERT-style classifier, building on the Bio_ClinicalBERT baseline introduced in §3.10, would provide a more decisive inference than either the rule-based heuristic or the generative-LLM probe; this is the natural complement when deterministic detection cannot generalize across reporting styles.

**A9.3 Out-of-scope sites — a distinct problem**

We treat out-of-scope sites not as a multi-primary failure but as a coverage gap to be closed by additional CAP-aligned organ modules (the schema framework supports incremental organ addition without disrupting the existing ten modules). Until those modules are added, "others" remains a confident triage class — its operational value is precisely that it surfaces cases the current schema cannot represent, rather than silently shoehorning them into an in-scope organ.

**A9.4 Cardinality-induced conflation caveat (applies to all three axes)**

A final caveat applies to *all three future-work axes*: detecting a second primary correctly does not guarantee that the downstream extraction produces two clean records, and two distinct problems sit beneath that guarantee.

First, the single-dictionary cancer_data schema holds only one TNM stage, one focality, one laterality, one biomarker submodule, and one tumor size, so cases with two physically separate primaries — bilateral breast carcinoma, separate-lobe lung primaries, multifocal pancreaticobiliary tumors — place every field at risk of conflation across the two lesions; the conflation is most visible on variable-length list fields such as regional lymph nodes and surgical margins, where ambiguity surfaces explicitly, but the limitation is schema-wide rather than list-field-specific.

Second, even when the schema is widened to two CancerEntity records, the model must still solve an attribute–lesion binding problem so that each extracted attribute lands on the correct lesion — pathology reports routinely interleave laterality with finding-type ("Left breast IDC pT2N1Mx; Right breast ILC pT1N0Mx"), and generative extraction is observed qualitatively to muddle this coreference.

Quantifying the cardinality-induced conflation rate and the post-extension binding-error rate is deferred to a follow-up study on synchronous-primary registry coverage. In the interim, the flag-for-review workflow remains the operational safety net because it does not require the model to be correct on either downstream layer, and the schema-cardinality extension above is therefore best understood as a *necessary but not sufficient* structural prerequisite for honest two-primary records rather than a fidelity refinement on top of the current schema.

**Supplementary Appendix A10. Field-level error attribution: the six-mechanism (M1–M6) taxonomy with per-mechanism deep dives.**

**A10.1 Framing**

A confusion-matrix-level audit of the eleven bottom-decile fields (§3.3.2) supports a six-mechanism attribution that the headline accuracy alone cannot resolve, and that our first-round four-bucket attempt mis-categorized in places. The six mechanisms are:

• **M1** source-bound qualitative phrasing

• **M2** heterogeneous reporting / null-on-no-clue

• **M3** anatomic ontology gap

• **M4** AJCC convention enforcement

• **M5** schema-shape

• **M6** unresolved model-side semantic conflation

Each is examined below.

**A10.2 M1 — Source-bound qualitative phrasing (no current bottom-decile field)**

No bottom-decile field on the current 30-run denominator manifests **source-bound qualitative phrasing** (M1) in its pure form; the M1 category remains in the taxonomy because it is a real failure mode for fields like tumor_percentage and prostate_weight that may surface in future runs.

**A10.3 M2 — Heterogeneous reporting / null-on-no-clue (four fields)**

Four fields fall under **heterogeneous reporting / null-on-no-clue (M2)**: surgical_technique, cancer_clock, cancer_quadrant, and tumor_necrosis. The previous response labelled these as source-bound, but per-organ heterogeneity and confusion-pattern audit tell a different story. surgical_technique ranges from 48.5 % in stomach to 99.8 % in prostate (Supplementary Table S29), a 51 pp spread driven by 3D and robotic prefix vocabulary the schema does not currently disambiguate; a small prefix glossary plus a [no prefix] → open default rule resolves most of the gap. The analogous fix for cancer_clock / cancer_quadrant is a clock→quadrant lookup table with explicit null-on-ambiguity for 12/3/6/9 o'clock readings, plus null-on-no-clue. Thyroid tumor_necrosis is the same null-on-no-mention pattern at a different field — the source report often does not mention tumor necrosis at all, and the model is insufficiently biased toward null, committing instead to a True/False value when the correct extraction is null. None of these is residual source ambiguity; all four are engineering-trivial fixes queued for the next pipeline iteration.

**A10.4 M4 — AJCC convention enforcement (one field)**

One field — distant_metastasis (19.1 % at κ = 0.95) — is **AJCC convention enforcement** (M4): the documented AJCC 8 rule treats absent-evidence metastasis as pM not applicable (Mx, not pM0), and the very low effective accuracy at near-perfect κ reflects inconsistent application of this rule across the corpus.

**A10.5 M5 — Schema-shape (four fields, three sub-mechanisms)**

Four further fields are **schema-shape** problems (M5):

• pathologic_stage_group is decomposed in §3.3.1 into three structural dimensions (y-descriptor source gap, HER-2 2+ FISH indeterminacy, residual narrative ambiguity); the full decomposition is in Appendix A8.

• tnm_descriptor has both a cardinality issue (the field should be List[StrEnum] | None to allow y/p/c/r/ypT co-occurrence) and a measurable hallucination bias toward the rare recurrent (r) descriptor that a DSPy null-bias would substantially reduce.

• The two breast biomarker fields (biomarker_her2, biomarker_ki67) manifest a third M5 sub-mechanism — **schema over-specification**. The HER-2 schema exposes score, percentage, and positivity slots when HER-2 in fact has only a clinically-meaningful score, and the Ki-67 schema exposes score, percentage, and positivity when Ki-67 has only a clinically-meaningful percentage; the extraneous slots invite the model to hallucinate values that have no source basis.

All three M5 sub-problems are schema-level reworks rather than further model tuning. The biomarker schema rework was identified on re-audit but is too invasive to ship in this revision.

**A10.6 M3 — Anatomic ontology gap (`procedure[liver]`, lymph-node stations)**

The deepest mechanism — **anatomic ontology gap** (M3) — does not appear in Table 9 because procedure is above 90 % globally, but procedure[liver] is 85.0 % and is the canonical case. Mapping a free-text operative name onto the controlled CAP partial-hepatectomy categories requires segment-counting and laterality reasoning that local LLMs do not natively perform: 3D right posterior sectionectomy involves segments VI/VII (partial hepatectomy, minor), left hepatectomy involves four segments (partial hepatectomy, major), and subsegmentectomy / liver transplantation / S6 liver resection each map differently. The same shape of failure drives the per-station canonicalization errors on lymph-node groups (§3.4). A registry-grade anatomic-ontology framework — addressing both lymph-node-station canonicalization and procedure-to-CAP-category mapping under a shared glossary methodology — is the subject of a separate manuscript currently in preparation; the present work reports the M3 gap as an explicit, structurally-bounded limitation rather than claiming to close it.

**A10.7 M6 — Unresolved model-side semantic conflation (colorectal tumor_invasion)**

Colorectal tumor_invasion (88.2 % at κ = 0.84) is the only bottom-decile field in the **unresolved model-side semantic conflation** category (M6). Confusion-matrix audit shows that 136 of 254 errors are gold = pericolorectal_tissue (pT3) → pred = muscularis_propria (pT2): the AJCC 8 pT3 definition is *"tumor invades through the muscularis propria into pericolorectal tissues"*, and the model appears to read "through muscularis" and emit the lower-tier muscularis_propria (pT2). We do not have an engineering remedy for this conflation. The author's anecdotal observation is that the downstream pT value remains correct on these cases (the conflation is at the depth-of-invasion enum and is not propagated to the staging output), but this is not separately verified in the present revision and is offered as a candid open limitation rather than a defended-away non-issue.

**A10.8 Audit-pending — pancreas `tumor_extension`**

Pancreas tumor_extension (77.1 % at κ = 0.61) is listed in Table 9 as **audit-pending**. Confusion-matrix audit isolates the dominant error as under-staging at the pT3/pT4 boundary (110 of 172 errors are gold = adjacent_organs_structures → pred = peripancreatic_soft_tissue); pancreas has no muscularis-propria staging boundary, so this is not the M6 pattern. The underlying mechanism is not yet determined and we decline to assign a speculative category in this revision.

**A10.9 What the taxonomy buys us**

Notably, no bottom-decile error stemmed from parse failure or schema-validation breakage; the parse-error rate across all 192 registry fields and thirty runs was 0.0 %. This decomposition reframes the residual gap from a generic "model accuracy is bounded at 60–90 % for hard fields" critique to a structured understanding of *which* part of the gap is closable by schema work (M5), *which* by deterministic post-processing (M2, M4), *which* requires a structured anatomic ontology and is therefore next-paper scope (M3), *which* is bounded by registrar documentation practices and not closable by extraction-model engineering alone (M1), and *which* remain open research questions (M6 and the audit-pending pancreas row).

**A10.10 List-typed-module addendum**

The same six-mechanism principle applies to the list-typed modules. For surgical margins (§3.4), the dominant error is **schema-driven hallucination** — the model emits an organ-specific category enumeration that is sometimes absent from the underlying narrative. This is conservative for the operationally critical "any-margin-positive" endpoint (every hallucinated entry defaults to margin_involved = false, which cannot generate a positivity false-negative) but inflates the per-station hallucination rate. For lymph-node groups (§3.4), the dominant error is **station-name canonicalization** — recall and precision diverge in the same direction (e.g., prostate group recall 0.578 vs precision 0.923) because the model collapses or normalizes free-text station strings in a way that fails the strict category alignment used for scoring. This is the M3 ontology gap surfacing at the list level rather than at the scalar level, and is addressed by the same forthcoming companion paper. Neither list-level error mode affects the clinical-safety endpoints (margin positivity, any-positive nodal status), which remain at saturation.
